# Age-, sex-, and ethnicity-stratified CMR strain reference ranges and centile-based categories from the Healthy Hearts Consortium

**DOI:** 10.64898/2026.10.08.26365067

**Authors:** Janek Salatzki, Dorina-Gabriela Condurache, Roman Roy, Liliana Szabo, Leo Westbury, Tesfamariam Akilu, Ahmed M Salih, Sucharitha Chadalavada, Robin Bülow, Giovanni Donato Aquaro, Thu-Thao Le, Dominika Suchá, Nay Aung, Nicholas C. Harvey, Tim Leiner, Calvin W.L. Chin, Andrea Barison, Marcus Dörr, Matthias G. Friedrich, Florian Andre, Steffen E. Petersen, Zahra Raisi-Estabragh

## Abstract

**Background:** This study presents reference ranges for cardiovascular magnetic resonance (CMR)-derived myocardial strain from the Healthy Hearts Consortium, while evaluating variations by demographic characteristics (age, sex, and ethnicity) and technical factors (scanner vendor, magnetic field strength). We derived reference ranges and centile-based categories for ventricular, atrial, and rapid strain parameters. The prognostic relevance of these categories was assessed by examining their associations with incident cardiovascular events.

**Methods:** The dataset included 9,088 CMR studies. Automated image analysis pipelines were established using suiteHEART (NeoSoft Inc.) and cvi42 (Circle Cardiovascular Imaging). A three-step quality control framework was applied including exclusion of incomplete data, visual assessment of image and segmentation quality, and statistical outlier removal. Ventricular strain and strain rate were measured using both software solutions. Additional cvi42-derived metrics included velocity and displacement, annular plane systolic excursion metrics, and rapid strain parameters. Age- and sex-specific reference ranges were generated using lambda–mu–sigma (LMS) modeling with penalized splines. Reference ranges were defined as values between the 2.5^th^ and 97.5^th^ percentiles. For lower-tail classification, values below the 2.5th percentile were considered mildly out-of-range and values below the 0.5th percentile at least moderately out-of-range. Differences across scanner vendors (Siemens, Philips, GE) and magnetic field strengths (1.5 T vs. 3 T) were evaluated using adjusted linear models. The prognostic relevance of selected cvi42-derived categories was evaluated in approximately 45,000 UK Biobank participants using Cox regression adjusted for age, sex, and body mass index.

**Results:** After quality control, 8,975 CMR examinations (cvi42) and 8,992 (suiteHEART) were available for analysis; these represent software-specific analytic cohorts. Most exclusions occurred during visual quality control. Women had higher absolute ventricular and atrial strain values than men across all age groups. Left ventricular (LV) longitudinal strain and strain rate were slightly lower in older ages, whereas circumferential and radial parameters remained largely stable. Left atrial strain was also lower in older age groups, while right atrial strain did not vary by age. Annular plane systolic excursion and rapid strain metrics showed similar sex-specific patterns, with modestly lower atrial and displacement values in older groups. Ethnicity-related differences were modest, heterogeneous, and substantially smaller in magnitude than age- and sex-related differences. Technical effects were parameter specific. While LV longitudinal and circumferential strains were largely unaffected, radial strain was higher at 3 T and showed vendor-related differences. LMS-derived reference ranges and centile thresholds were applied both across all ethnicities and within ethnicity-specific analyses, and are presented stratified by age and sex. In the prognostic analysis, centile categories were most consistently associated with incident heart failure. LV longitudinal, circumferential, and radial strain below the 0.5th centile were associated with significantly increased risk of incident heart failure (HRs: 9.9 to 12.4, 95% CIs 7.00–13.90 to 8.49–18.06).

**Conclusion:** This study provides age- and sex-specific reference ranges and centile-based categories for CMR-derived strain parameters in a large reference population. Age and sex represent the primary determinants of myocardial deformation, whereas scanner-related effects are parameter-specific. Values in the lower extremes of the healthy reference distribution, particularly for LV strain, are associated with increased risk of incident heart failure. These findings enable standardized, centile-based classification of myocardial strain, facilitating consistent interpretation and improving risk stratification in research and clinical practice.

## Introduction

Cardiovascular magnetic resonance (CMR) enables accurate, non-invasive assessment of cardiac structure and function and has become a key tool for evaluating myocardial deformation [1]. Feature tracking CMR (FT-CMR) has emerged as a practical and widely used approach for myocardial strain quantification, as it can be applied to routinely acquired cine images without the need for dedicated strain acquisition sequences and allows relatively efficient post-processing [2–4]. FT-CMR-derived strain provides important diagnostic and prognostic information across a range of cardiovascular diseases, including heart failure, ischemic heart disease, cardiomyopathy and myocarditis [5–9].

Myocardial strain is influenced by demographic factors. Strain values are lower in older age groups, and women typically exhibit higher (more negative) strain values than men [10,11]. In addition, ethnicity-related differences and variability across magnetic field strengths and software vendors have been reported [10,12–14].

Despite its growing clinical importance, robust population-based reference ranges for CMR-derived strain are scarce. Existing studies are often small, ethnically homogeneous, and restricted in age range, limiting generalizability [15–20]. The lack of standardized reference values hampers clinical implementation and may lead to misclassification and suboptimal risk stratification. In addition, FT-CMR strain values have been shown to vary across post-processing software packages, underscoring the need for software-specific reference ranges [21].

The Healthy Hearts Consortium (HHC) is a multicenter collaborative comprising over 9,000 CMR examinations from verified healthy adults across six institutions [22]. The dataset encompasses a broad adult age range and diverse ethnic backgrounds, providing a unique platform to derive robust and clinically applicable CMR reference ranges.

This study established automated analysis pipelines using two clinically used software solutions to derive age-, sex-, and ethnicity-specific reference ranges and centile categories for ventricular and atrial FT-CMR strain, strain rate, rapid strain, and annular plane systolic excursion using data from the HHC. The effects of demographic and technical factors were assessed, and the prognostic relevance of the derived centiles was evaluated.

The work provides a comprehensive framework for standardized strain interpretation and supports improved clinical application of CMR-derived deformation metrics.

## Materials and Methods

### Healthy Hearts Consortium

The HHC is an international collaboration including the largest global collection of CMR studies of healthy adults without cardiovascular disease, with 9,088 scans from six contributing institutions [22]. This dataset is notable for its size, broad age range, and ethnic diversity. The contributing sites comprise published CMR reference datasets from Utrecht (The Netherlands) [23], Pisa (Italy) [24], Heidelberg (Germany) [25], Singapore [26] and healthy subsets from SHIP (Study of Health in Pomerania) [27] and UK Biobank [28].

The dataset includes anonymized CMR series in DICOM format, demographic information, body size measurements, and selected technical parameters. The dataset is centrally collated at the William Harvey Research Institute, Queen Mary University of London, London, UK. Further details are available online: https://healthy-hearts.org.uk [22].

### Definition of healthy cohorts

The contributing cohorts applied study-specific criteria to define cardiovascular health. All cohorts excluded individuals with known cardiovascular disease, although the screening procedures differed between studies. Some cohorts included dedicated clinical assessments or additional investigations to exclude cardiovascular risk factors or subclinical disease, whereas the population-based SHIP and UK Biobank cohorts identified eligible healthy subsets using research assessments and linked health-record data. For UK Biobank, additional sampling methods were applied to better align the selected healthy subset with the demographic, socioeconomic, and cardiovascular risk-factor distribution of the general population using data from the Health Survey for England, while also increasing the representation of participants from diverse ethnic backgrounds. Full details of the cohort-specific eligibility criteria and sampling procedures are provided in our previous Healthy Hearts Consortium publication and the original source publications for each contributing cohort [22–29].

### Population-related and technical sources of variation

We considered age, sex, ethnicity, and technical factors as potential sources of variation in CMR-derived strain metrics. Technical factors included scanner vendor, magnetic field strength, and post-processing software, given the known variability in FT-CMR measurements across software platforms. Each HHC site provided details on CMR scanner vendor and magnetic field strength (1.5 T or 3.0 T).

The Utrecht, Heidelberg, and SHIP cohorts comprised participants from White/Caucasian ethnic backgrounds. The Pisa cohort included one participant of Black ethnicity, with the remainder classified as White/Caucasian. The Singapore cohort comprised participants of Chinese ethnicity. Ethnicity categories provided by UK Biobank were “White,” “Black or Black British,” “Asian or Asian British,” “Chinese,” “Mixed,” and “Other.” In UK Biobank, “Asian or Asian British” refers to Indian, Pakistani, Bangladeshi, or other Asian backgrounds, in accordance with UK government classifications. Consistent with our previous Healthy Hearts Consortium publication, we labeled this group “South Asian” to distinguish it from the separately defined Chinese ethnicity group [29].

### CMR post-processing and imaging analysis

To evaluate software-related variability, CMR strain metrics were analyzed using two post-processing software platforms: cvi42 (prototype version 5.11.2, Circle Cardiovascular Imaging Inc.) and suiteHEART (prototype version 5.0.0, NeoSoft Inc.). This enabled assessment of whether reference ranges and observed population-related patterns were consistent across software solutions. The CMR-derived deformation metrics included in the analysis and their respective post-processing software platforms are summarized in **Table 1**.

**Table 1.** CMR-derived deformation metrics included in the analysis, by metric group and post-processing software.

| Metric group | Specific metrics | Post-processing software |
| --- | --- | --- |
| **Ventricular global strain** | LV GLS, LV GCS, LV GRS, RV GLS, RV GCS, RV GRS | cvi42; suiteHEART |
| **Ventricular global strain rate** | LV/RV systolic and diastolic GLSR, GCSR, GRSR | cvi42 |
| **LV global displacement** | LV GLD, GCD, GRD | cvi42 |
| **LV global velocity** | LV GLV, GCV, GRV | cvi42 |
| **Atrial global strain** | LA GLS, RA GLS | suiteHEART |
| **Annular plane systolic excursion** | MAPSE lateral, MAPSE septal, MAPSE inferior, MAPSE anterior, TAPSE | cvi42 |
| **Ventricular long-axis and atrioventricular junction strain** | LV long-axis strain, LV AV-junction strain | cvi42 |
| **Atrial long-axis and atrioventricular junction strain** | LA long-axis strain, LA AV-junction strain, RA long-axis strain, RA AV-junction strain | cvi42 |

**Footnote:** Metrics were derived using cvi42 (prototype version 5.11.2, Circle Cardiovascular Imaging Inc.), and suiteHEART (prototype version 5.0.0, NeoSoft Inc.). Ventricular global strain metrics were derived using both cvi42 and suiteHEART. Ventricular global strain rate, LV global displacement, LV global velocity, annular plane systolic excursion, and ventricular and atrial long-axis and atrioventricular junction strain metrics were derived using cvi42. Atrial global strain metrics were derived using suiteHEART. LV = left ventricle; RV = right ventricle; LA = left atrium; RA = right atrium; GLS = global longitudinal strain; GCS = global circumferential strain; GRS = global radial strain; GLSR = global longitudinal strain rate; GCSR = global circumferential strain rate; GRSR = global radial strain rate; GLD = global longitudinal displacement; GCD = global circumferential displacement; GRD = global radial displacement; GLV = global longitudinal velocity; GCV = global circumferential velocity; GRV = global radial velocity; MAPSE = mitral annular plane systolic excursion; TAPSE = tricuspid annular plane systolic excursion; AV = atrioventricular.

CMR series were transferred to two offline workstations, where FT-CMR analysis was performed using batch processing pipelines of the two software solutions.

Endocardial and epicardial borders in standard short-axis and long-axis (2-, 3-, and 4-chamber) cine views were automatically delineated. Contours were generated across all cardiac phases from end-diastole to end-systole. Trabeculae, papillary muscles, pericardium, and epicardial fat were excluded from the left ventricular myocardium.

Global strain metrics, including global radial strain (GRS), global circumferential strain (GCS), and global longitudinal strain (GLS) for the left and right ventricles, were obtained both software solutions, cvi42 and suiteHEART. Measurements were performed in short-axis slices for radial and circumferential strain and in three standard long-axis views (2-, 3- and 4-chamber views) for longitudinal strain. Systolic (sys) and diastolic (dia) strain rates (GRSR, GCSR, GLSR) were calculated using cvi42, following the same orientations as the corresponding strain metrics. Additional metrics, such as global longitudinal, circumferential, and radial displacement (GLD, GCD and GRD) and systolic and diastolic velocity metrics (GLV, GCV and GRV), with systolic and diastolic velocity measurements reported separately, were all derived using cvi42. For atrial strain, suiteHEART was used to assess longitudinal atrial strain, with endocardial borders traced on 2- and 4-chamber long-axis cine images.

Mitral annular plane systolic excursion (MAPSE) and tricuspid annular plane systolic excursion (TAPSE) were quantified using cvi42. In the 4-chamber view, lateral and septal MAPSE were calculated as the absolute displacement of the respective mitral annular insertion points between end-diastole and end-systole, without use of an external thoracic reference point. In the 2-chamber view, inferior and anterior MAPSE were derived analogously from the corresponding annular insertion points. TAPSE was measured in the 4-chamber view as the displacement of the tricuspid annulus relative to the right ventricular free wall between end-diastole and end-systole.

Rapid longitudinal shortening was assessed as a geometric surrogate of longitudinal ventricular deformation and was derived using cvi42. This parameter quantifies the relative percentage change in distance between the left ventricular (LV) epicardial apex and the atrioventricular (AV) junction from end-diastole to end-systole, thereby reflecting global longitudinal shortening without directly measuring myocardial strain. Two established measurement approaches were applied. Long-axis shortening (LS) was defined as the distance from the LV epicardial apex to the midpoint of a line connecting the mitral valve leaflet insertion points at end-diastole and end-systole in both 2- and 4-chamber views. In contrast, atrioventricular junction shortening (AVJS) was calculated as the shortening of the distance between the LV epicardial apex and the individual mitral valve insertion points – septal and lateral in the 4-chamber view, and anterior and inferior in the 2-chamber view – measured at end-diastole and end-systole. Both metrics were automatically derived and averaged across the two long-axis views to enhance robustness and reduce variability specific to each view.

### Quality control

Comprehensive quality control (QC) was applied to ensure accuracy of the obtained metrics. This multi-step QC approach included visual expert assessment of image and segmentation/tracking quality, removal of non-sensical values, and statistical outlier removal. Only available measurements proceeded to downstream analysis. Two independent readers performed a visual evaluation of the cine CMR images (J.S, D-G.C), scoring image quality, planning and tracking quality for each cardiac chamber using a three-point grading system (grade 1=perfect, grade 2=satisfactory, grade 3=unacceptable). All visual QC ratings were subsequently reviewed by J.S., a Level III-certified CMR cardiologist with >5 years of experience in CMR. GLS values were excluded from analysis if any ventricular chamber received a grade 3 rating in the 2-, 3-, or 4-chamber view. Similarly, GCS and GRS values were excluded if any ventricular chamber in the short-axis views received a grade 3 rating. The same quality-control framework was applied to atrial strain measurements, with atrial strain values excluded when the corresponding long-axis views received a grade 3 rating for image, planning, or tracking quality.

Following the visual assessment, non-sensical values were removed, including GLS and GCS values >0 and GRS values <0, indicating software tracking errors. Finally, a statistical outlier removal was applied to remove very extreme values using the 3 x IQR rule (three times the interquartile range (IQR) below the first and above the third quartile method) [30]. The **Supplementary Figures 1-3** detail the number of cases removed during this process.

### Incident outcome associations in UK Biobank

The prognostic relevance of the derived reference values and centile-based categories was evaluated in a UK Biobank validation cohort with available cvi42-derived CMR strain measurements, covariates, and linked outcome data.

Incident cardiovascular events occurring for the first time after imaging were ascertained through linked hospital inpatient and death registry data; these included heart failure, myocardial infarction, stroke, and all-cause death. Participants with the relevant outcome on or before the imaging date were excluded from the corresponding incident outcome analysis. Follow-up extended until the first occurrence of the endpoint of interest, death, or administrative censoring on March 31, 2023, whichever occurred first.

Age- and sex-specific reference ranges and centile-based thresholds were taken from the reference tables derived in the present study. Participants were assigned to the corresponding age stratum and sex category, and each observed CMR value was mapped to the relevant reference interval and centile-based category, as described in the statistical analysis section. The in-range category was the reference group.

### Statistical analysis

Statistical analyses were performed using R (version 4.3.3; R Foundation for Statistical Computing, Vienna, Austria) within RStudio (Posit Software). Continuous variables are presented as mean ± standard deviation (SD) or median with interquartile range (IQR), as appropriate. Distributional assumptions were assessed visually using histograms and Q–Q plots. All strain values are reported as absolute values to facilitate clinical interpretation[31].

Age-specific reference ranges were estimated using generalized additive models for location, scale and shape (GAMLSS) using the *gamlss* package (version 5.4.22) in R. For each cardiac measurement, sex- and ethnicity-specific models were fitted using the Box–Cox Cole and Green distribution (BCCG), which is suitable for modeling strictly positive variables that may deviate from a normal distribution [32]. An initial fully flexible model allowed the location (μ), scale (σ), and skewness (ν) parameters to vary smoothly with age. When this resulted in model instability, defined a priori as non-convergence, unsafe centile predictions, or implausible extreme centile estimates not supported by the observed data (e.g. unrealistically wide tails), progressively more constrained models were fitted. These included models with constant skewness (ν) and, where required, constant scale (σ) and skewness (ν), to ensure stable and clinically interpretable reference ranges and centiles. Where model instability persisted, additional fallback models with simplified spline, linear, or intercept-only age specifications were used where necessary. Subgroups with fewer than 30 observations were not modeled.

Centiles were derived for each metric stratified by sex and, where applicable, ethnicity. Reference ranges were defined using the conventional 95% reference range (2.5th-97.5th percentiles) [33,34]. Separately, for lower-tail classification of reduced values, measurements below the 2.5th percentile were classified as mildly out-of-range and those below the 0.5th percentile as at least moderately out-of-range; values at or above the 2.5th percentile served as the reference category for these lower-tail analyses. The 0.5th percentile was selected as the threshold for more marked out-of-range position, as this represented the most extreme tail threshold that remained robust and data-supported, whereas more extreme centiles yielded unstable estimates due to limited number of observations at the distribution extremes.

For tabular presentation, age-specific centile estimates were summarized within predefined age categories (18–29, 30–39, 40–49, 50–59, 60–69, and ≥70 years) by calculating median centile values within each age band. Age categories with fewer than 10 observations were excluded from the final reference tables.

For subgroup comparisons across ethnicity, analyses were performed separately within age- and sex-stratified groups using the Wilcoxon rank-sum test, with White participants serving as the reference group. To account for multiple testing across variables, age groups, and sex strata, p-values were adjusted using the Bonferroni method. Results are presented as directional differences relative to the reference group. Comparisons were not performed where either subgroup contained fewer than five observations. To assess scanner-related effects, separate multivariable linear regression models were fitted for each CMR-derived deformation parameter. These included ventricular strain, atrial strain, rapid strain, MAPSE, and TAPSE metrics derived from cvi42 and suiteHEART, where applicable.

Models included scanner vendor or magnet strength as the primary independent variable and were adjusted for age and sex, with Siemens and 1.5 T specified as reference categories, respectively. Adjusted marginal means were estimated using the estimated marginal means approach, and pairwise comparisons were derived using treatment-versus-control contrasts. Results are reported as adjusted mean differences with corresponding 95% confidence intervals. Global group effects were assessed using analysis of variance.

Agreement between the two post-processing platforms was assessed by direct paired comparison of ventricular strain metrics measured in the same participants analyzed with both cvi42 and suiteHEART. Examinations were matched across software exports using the participant identifier and imaging visit. For each metric, the difference (suiteHEART - cvi42) was plotted against the mean of the two measurements using Bland–Altman analysis. The bias was defined as the mean of the paired differences, and the 95% limits of agreement as the bias ± 1.96·SD of the differences, assuming approximately normally distributed differences. The strength of the linear association was summarized using the Pearson correlation coefficient. This analysis was restricted to ventricular strain (LV and RV GLS, GCS, and GRS), the metrics available from both platforms.

For each endpoint and CMR metric, we first calculated crude event counts, competing deaths, cumulative risk, total follow-up time, and incidence rates per 1,000 person-years across centile-based categories. Cox proportional hazards regression was then used to evaluate the association between centile-based categories and incident outcomes. Hazard ratios (HRs) and 95% confidence intervals (CIs) were estimated using the in-range category as the reference.

Primary models were adjusted for age and sex at imaging. Model assumptions, including the proportional hazards assumption, were assessed according to established survival modeling procedures and in line with prior UK Biobank CMR strain outcome analyses [9]. Participants with missing data for the relevant exposure, covariates, or endpoint-specific follow-up were excluded from the corresponding complete-case analysis.

Event-free survival was visualized using Kaplan–Meier curves stratified by centile-based category. Adjusted associations between CMR categories and incident outcomes were summarized using horizontal forest plots displaying HRs and 95% CIs. This analysis was designed to test whether values below the derived reference range, including mildly reduced and at least moderately out-of-range categories, were associated with higher risk of subsequent cardiovascular events compared with values within the in-range category.

A two-sided p-value <0.05 was considered statistically significant. For ethnicity analyses, adjusted p-values were interpreted in the context of multiple testing.

## Results

### Quality control

A structured three-tiered QC framework was applied across six datasets: UK Biobank (n = 7,672), SHIP (n = 634), Heidelberg (n = 192 for cvi42-derived measures, including one duplicate case; n = 191 for suiteHEART-derived measures), Utrecht (n = 133), Pisa (n = 292), and Singapore (n = 180). The QC framework comprised (i) initial technical exclusions, defined as removal of cases with incomplete batch processing, unavailable or incomplete output data, or duplicate entries, (ii) visual expert assessment, and (iii) statistical outlier control (**Supplementary Figures 1–3**).

For cvi42-derived global strain, 9,103 CMR examinations entered the QC workflow. Initial technical exclusions were minimal (n = 50 overall). Final available values ranged from 7,995 for LV-GLS to 8,575 for RV-GCS, corresponding to final data availability of approximately 88% to 94%. Exclusions varied by metric and cohort but were predominantly driven by visual QC, particularly for LV-GLS and RV-GLS in UK Biobank (727 and 388 values removed, respectively). Lower absolute numbers of exclusions were observed in the smaller contributing cohorts. Statistical QC had only a marginal additional impact (generally ≤1%). Consequently, the majority of LV and RV strain measurements remained available for analysis (**Supplementary Figure 1**).

For suiteHEART-derived strain, 9,102 CMR examinations entered the QC workflow. Initial technical exclusions were similarly low (n = 60 overall). Final available values ranged from 7,052 for RA-LS to 8,859 for LV-GCS, corresponding to final data availability of approximately 78% to 97%. After QC, LV-GRS, LV-GCS, and RV-GRS had the highest data availability, whereas atrial strain parameters showed the greatest number of exclusions, particularly LA-LS and RA-LS in UK Biobank (1,257 and 1,396 values removed by visual QC, respectively). Smaller but variable exclusions were observed across the other cohorts. Statistical QC again contributed minimally (**Supplementary Figure 2**).

For cvi42-derived MAPSE, TAPSE, and LAS, initial exclusions were negligible (≤7 cases per cohort). Visual QC accounted for the majority of removals, particularly for atrial LAS and septal/lateral MAPSE in UK Biobank, whereas other cohorts showed lower absolute numbers of exclusions. Statistical QC resulted in minimal additional exclusions.

Overall, visual QC was the dominant exclusion step across all parameters and cohorts, while statistical QC had only a minor impact. Most measurements remained available for downstream analyses (**Supplementary Figure 3**).

### Population characteristics

The final cvi42 cohort comprised 8,975 CMR examinations, including 4,595 women (51.2%) and 4,380 men (48.8%). The mean age was 61 ± 13 years (range 18–83 years). A total of 81.6% (n=7,327) participants were from White ethnic backgrounds, with smaller cohorts of South Asian (5.6%), Black (3.7%), Chinese (3.7%), and other/Mixed (5.3%) groups. The sex distribution was near equal across most ethnic group, except for the South Asians (male dominant) and the Other/Mixed (female dominant) groups. Detailed distributions stratified by sex are provided in **Table 2**.

**Table 2.**
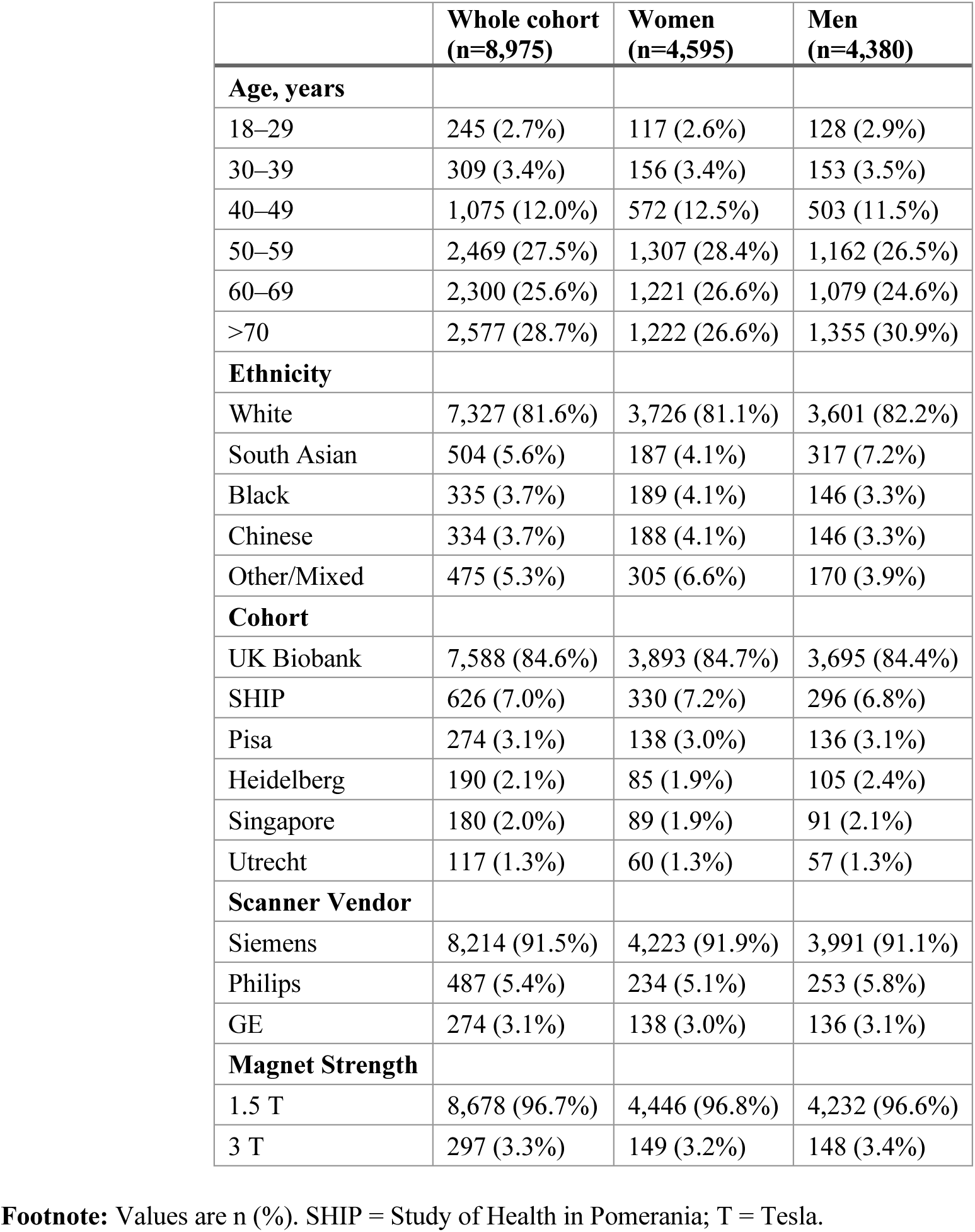
Baseline characteristics of cohorts after quality control using cvi42 (Circle Cardiovascular Imaging Inc.)

|  | **Whole cohort**<br>**(n=8,975)** | **Women**<br>**(n=4,595)** | **Men**<br>**(n=4,380)** |
| --- | --- | --- | --- |
| **Age, years** |  |  |  |
| 18–29 | 245 (2.7%) | 117 (2.6%) | 128 (2.9%) |
| 30–39 | 309 (3.4%) | 156 (3.4%) | 153 (3.5%) |
| 40–49 | 1,075 (12.0%) | 572 (12.5%) | 503 (11.5%) |
| 50–59 | 2,469 (27.5%) | 1,307 (28.4%) | 1,162 (26.5%) |
| 60–69 | 2,300 (25.6%) | 1,221 (26.6%) | 1,079 (24.6%) |
| >70 | 2,577 (28.7%) | 1,222 (26.6%) | 1,355 (30.9%) |
| **Ethnicity** |  |  |  |
| White | 7,327 (81.6%) | 3,726 (81.1%) | 3,601 (82.2%) |
| South Asian | 504 (5.6%) | 187 (4.1%) | 317 (7.2%) |
| Black | 335 (3.7%) | 189 (4.1%) | 146 (3.3%) |
| Chinese | 334 (3.7%) | 188 (4.1%) | 146 (3.3%) |
| Other/Mixed | 475 (5.3%) | 305 (6.6%) | 170 (3.9%) |
| **Cohort** |  |  |  |
| UK Biobank | 7,588 (84.6%) | 3,893 (84.7%) | 3,695 (84.4%) |
| SHIP | 626 (7.0%) | 330 (7.2%) | 296 (6.8%) |
| Pisa | 274 (3.1%) | 138 (3.0%) | 136 (3.1%) |
| Heidelberg | 190 (2.1%) | 85 (1.9%) | 105 (2.4%) |
| Singapore | 180 (2.0%) | 89 (1.9%) | 91 (2.1%) |
| Utrecht | 117 (1.3%) | 60 (1.3%) | 57 (1.3%) |
| **Scanner Vendor** |  |  |  |
| Siemens | 8,214 (91.5%) | 4,223 (91.9%) | 3,991 (91.1%) |
| Philips | 487 (5.4%) | 234 (5.1%) | 253 (5.8%) |
| GE | 274 (3.1%) | 138 (3.0%) | 136 (3.1%) |
| **Magnet Strength** |  |  |  |
| 1.5 T | 8,678 (96.7%) | 4,446 (96.8%) | 4,232 (96.6%) |
| 3 T | 297 (3.3%) | 149 (3.2%) | 148 (3.4%) |

**Footnote:** Values are n (%). SHIP = Study of Health in Pomerania; T = Tesla.

Following quality control using suiteHEART, 8,992 CMR examinations were available for analysis (4,597 women [51.1%] and 4,395 men [48.9%]). The age distribution was comparable (mean 61 ± 13 years; range 18–83 years). Detailed characteristics are presented in **Supplementary Table 1**.

For annular plane systolic excursion and rapid longitudinal shortening metrics derived using cvi42, the final cohort comprised 8,984 CMR examinations (4,598 women [51.2%] and 4,386 men [48.8%]). Detailed sex-stratified characteristics are provided in **Supplementary Table 2**.

### Age- and sex-specific patterns in strain metrics

Across all global strain parameters derived using cvi42, consistent sex-specific differences were observed (**Tables 3–4**; **Figures 1–2**). Age-related patterns were generally modest and parameter specific. LV GLS decreased slightly across age groups in women but was largely stable in men, while several circumferential and radial strain measures showed small increases with age.

**Figure 1.**
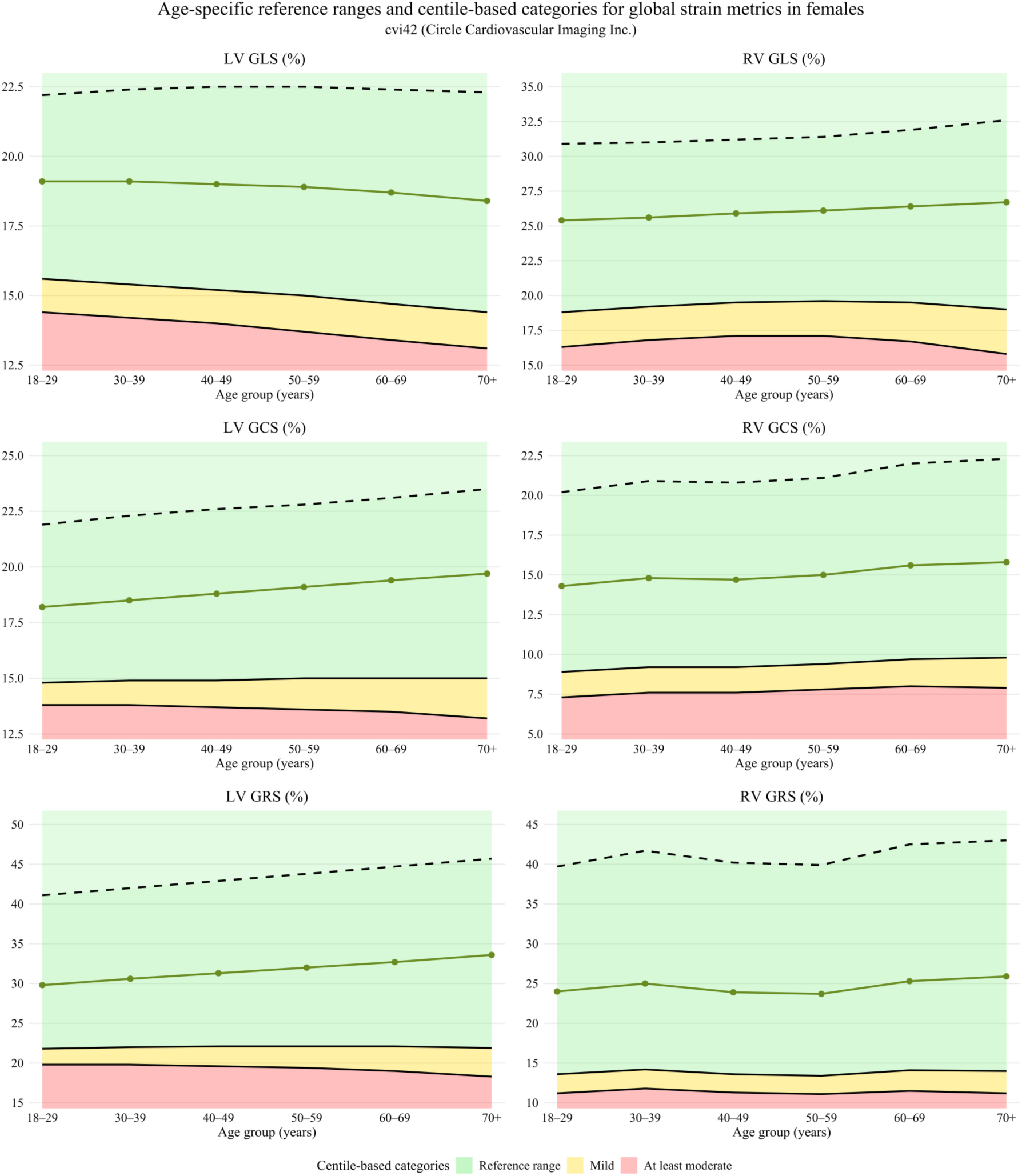
Age-specific reference ranges and centile-based categories for global ventricular strain metrics in females (*all ethnicities*), using cvi42 (Circle Cardiovascular Imaging Inc.) Age-specific reference ranges and centile-based categories for left ventricular (LV) and right ventricular (RV) global strain metrics derived from CMR feature tracking (cvi42, Circle Cardiovascular Imaging Inc.) in females. Solid lines represent median values across age groups, dashed lines indicate upper reference limits, and shaded areas depict centile-based categories: green (reference range, ≥P2.5), yellow (mild reduction, <P2.5), and red (at least moderate reduction, <P0.5). Age-related trends are shown for LV and RV global longitudinal (GLS), circumferential (GCS), and radial strain (GRS).

**Figure 2.**
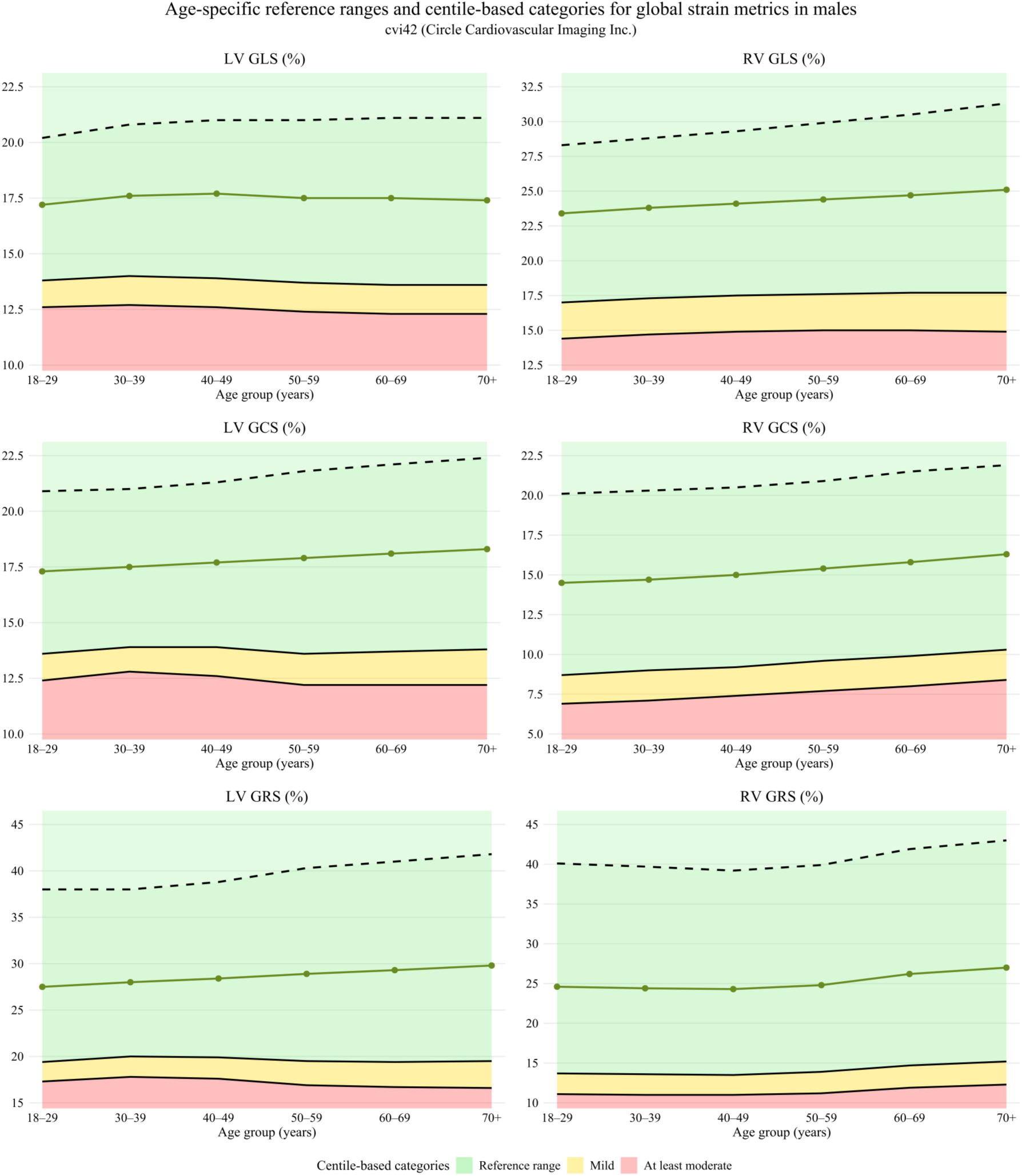
Age-specific reference ranges and centile-based categories for global ventricular strain metrics in males (*all ethnicities*), using cvi42 (Circle Cardiovascular Imaging Inc.) Age-specific reference ranges and centile-based categories for left ventricular (LV) and right ventricular (RV) global strain metrics derived from CMR feature tracking (cvi42, Circle Cardiovascular Imaging Inc.) in males. Solid lines represent median values across age groups, dashed lines indicate upper reference limits, and shaded areas depict centile-based categories: green (reference range, ≥P2.5), yellow (mild reduction, <P2.5), and red (at least moderate reduction, <P0.5). Age-related trends are shown for LV and RV global longitudinal (GLS), circumferential (GCS), and radial strain (GRS).

**Table 3.** Left and right ventricular global strain reference ranges and centile-based categories stratified by sex and age for all ethnicities using cvi42 (Circle Cardiovascular Imaging Inc.)

| Age group | Metrics (%) | Female |  |  |  | Male |  |  |  |
| --- | --- | --- | --- | --- | --- | --- | --- | --- | --- |
|  |  | Reference threshold | Mild | At least moderate | n | Reference threshold | Mild | At least moderate | n |
| 18–29 | LV GLS | ≥ 15.6 | 14.4–15.5 | < 14.4 | 76 | ≥ 13.8 | 12.6–13.7 | < 12.6 | 93 |
|  | LV GCS | ≥ 14.8 | 13.8–14.7 | < 13.8 | 105 | ≥ 13.6 | 12.4–13.5 | < 12.4 | 120 |
|  | LV GRS | ≥ 21.8 | 19.8–21.7 | < 19.8 | 105 | ≥ 19.4 | 17.3–19.3 | < 17.3 | 120 |
|  | RV GLS | ≥ 18.8 | 16.3–18.7 | < 16.3 | 89 | ≥ 17.0 | 14.4–16.9 | < 14.4 | 98 |
|  | RV GCS | ≥ 8.9 | 7.3–8.8 | < 7.3 | 100 | ≥ 8.7 | 6.9–8.6 | < 6.9 | 119 |
|  | RV GRS | ≥ 13.6 | 11.2–13.5 | < 11.2 | 100 | ≥ 13.7 | 11.1–13.6 | < 11.1 | 119 |
| 30–39 | LV GLS | ≥ 15.4 | 14.2–15.3 | < 14.2 | 125 | ≥ 14.0 | 12.7–13.9 | < 12.7 | 125 |
|  | LV GCS | ≥ 14.9 | 13.8–14.8 | < 13.8 | 140 | ≥ 13.9 | 12.8–13.8 | < 12.8 | 137 |
|  | LV GRS | ≥ 22.0 | 19.8–21.9 | < 19.8 | 140 | ≥ 20.0 | 17.8–19.9 | < 17.8 | 137 |
|  | RV GLS | ≥ 19.2 | 16.8–19.1 | < 16.8 | 126 | ≥ 17.3 | 14.7–17.2 | < 14.7 | 135 |
|  | RV GCS | ≥ 9.2 | 7.6–9.1 | < 7.6 | 141 | ≥ 9.0 | 7.1–8.9 | < 7.1 | 136 |
|  | RV GRS | ≥ 14.2 | 11.8–14.1 | < 11.8 | 141 | ≥ 13.6 | 11.0–13.5 | < 11.0 | 136 |
| 40–49 | LV GLS | ≥ 15.2 | 14.0–15.1 | < 14.0 | 530 | ≥ 13.9 | 12.6–13.8 | < 12.6 | 454 |
|  | LV GCS | ≥ 14.9 | 13.7–14.8 | < 13.7 | 521 | ≥ 13.9 | 12.6–13.8 | < 12.6 | 461 |
|  | LV GRS | ≥ 22.1 | 19.6–22.0 | < 19.6 | 521 | ≥ 19.9 | 17.6–19.8 | < 17.6 | 461 |
|  | RV GLS | ≥ 19.5 | 17.1–19.4 | < 17.1 | 540 | ≥ 17.5 | 14.9–17.4 | < 14.9 | 465 |
|  | RV GCS | ≥ 9.2 | 7.6–9.1 | < 7.6 | 546 | ≥ 9.2 | 7.4–9.1 | < 7.4 | 481 |
|  | RV GRS | ≥ 13.6 | 11.3–13.5 | < 11.3 | 546 | ≥ 13.5 | 11.0–13.4 | < 11.0 | 481 |
| 50–59 | LV GLS | ≥ 15.0 | 13.7–14.9 | < 13.7 | 1206 | ≥ 13.7 | 12.4–13.6 | < 12.4 | 1029 |
|  | LV GCS | ≥ 15.0 | 13.6–14.9 | < 13.6 | 1187 | ≥ 13.6 | 12.2–13.5 | < 12.2 | 1056 |
|  | LV GRS | ≥ 22.1 | 19.4–22.0 | < 19.4 | 1186 | ≥ 19.5 | 16.9–19.4 | < 16.9 | 1056 |
|  | RV GLS | ≥ 19.6 | 17.1–19.5 | < 17.1 | 1244 | ≥ 17.6 | 15.0–17.5 | < 15.0 | 1090 |
|  | RV GCS | ≥ 9.4 | 7.8–9.3 | < 7.8 | 1248 | ≥ 9.6 | 7.7–9.5 | < 7.7 | 1132 |
|  | RV GRS | ≥ 13.4 | 11.1–13.3 | < 11.1 | 1246 | ≥ 13.9 | 11.2–13.8 | < 11.2 | 1132 |
| 60–69 | LV GLS | ≥ 14.7 | 13.4–14.6 | < 13.4 | 1132 | ≥ 13.6 | 12.3–13.5 | < 12.3 | 953 |
|  | LV GCS | ≥ 15.0 | 13.5–14.9 | < 13.5 | 1128 | ≥ 13.7 | 12.2–13.6 | < 12.2 | 975 |
|  | LV GRS | ≥ 22.1 | 19.0–22.0 | < 19.0 | 1129 | ≥ 19.4 | 16.7–19.3 | < 16.7 | 975 |
|  | RV GLS | ≥ 19.5 | 16.7–19.4 | < 16.7 | 1153 | ≥ 17.7 | 15.0–17.6 | < 15.0 | 996 |
|  | RV GCS | ≥ 9.7 | 8.0–9.6 | < 8.0 | 1172 | ≥ 9.9 | 8.0–9.8 | < 8.0 | 1049 |
|  | RV GRS | ≥ 14.1 | 11.5–14.0 | < 11.5 | 1169 | ≥ 14.7 | 11.9–14.6 | < 11.9 | 1048 |
| 70+ | LV GLS | ≥ 14.4 | 13.1–14.3 | < 13.1 | 1105 | ≥ 13.6 | 12.3–13.5 | < 12.3 | 1167 |
|  | LV GCS | ≥ 15.0 | 13.2–14.9 | < 13.2 | 1130 | ≥ 13.8 | 12.2–13.7 | < 12.2 | 1215 |
|  | LV GRS | ≥ 21.9 | 18.3–21.8 | < 18.3 | 1130 | ≥ 19.5 | 16.6–19.4 | < 16.6 | 1216 |
|  | RV GLS | ≥ 19.0 | 15.8–18.9 | < 15.8 | 1158 | ≥ 17.7 | 14.9–17.6 | < 14.9 | 1238 |
|  | RV GCS | ≥ 9.8 | 7.9–9.7 | < 7.9 | 1153 | ≥ 10.3 | 8.4–10.2 | < 8.4 | 1298 |
|  | RV GRS | ≥ 14.0 | 11.2–13.9 | < 11.2 | 1152 | ≥ 15.2 | 12.3–15.1 | < 12.3 | 1298 |

**Footnote:** All strain measurements were derived using cvi42 software with standardized feature-tracking analysis. Global longitudinal strain (GLS), global circumferential strain (GCS), and global radial strain (GRS) are presented as absolute values (%). Reference ranges are defined as the interval between the 2.5th and 97.5th centiles derived from age- and sex-specific generalized additive models for location, scale and shape (GAMLSS) using the Box–Cox Cole–Green distribution. Centile-based categories beyond the reference range are derived from LMS-based thresholds (P0.5 and P2.5) and describe the position of a measurement within the reference distribution rather than disease severity or diagnostic abnormality. For clarity, only the lower reference threshold (2.5th percentile) and corresponding lower centile cut-offs are presented in this table. Reference values are based on all ethnicities. **Abbreviations:** LV = left ventricle; RV = right ventricle; GLS = global longitudinal strain; GCS = global circumferential strain; GRS = global radial strain.

**Table 4.**
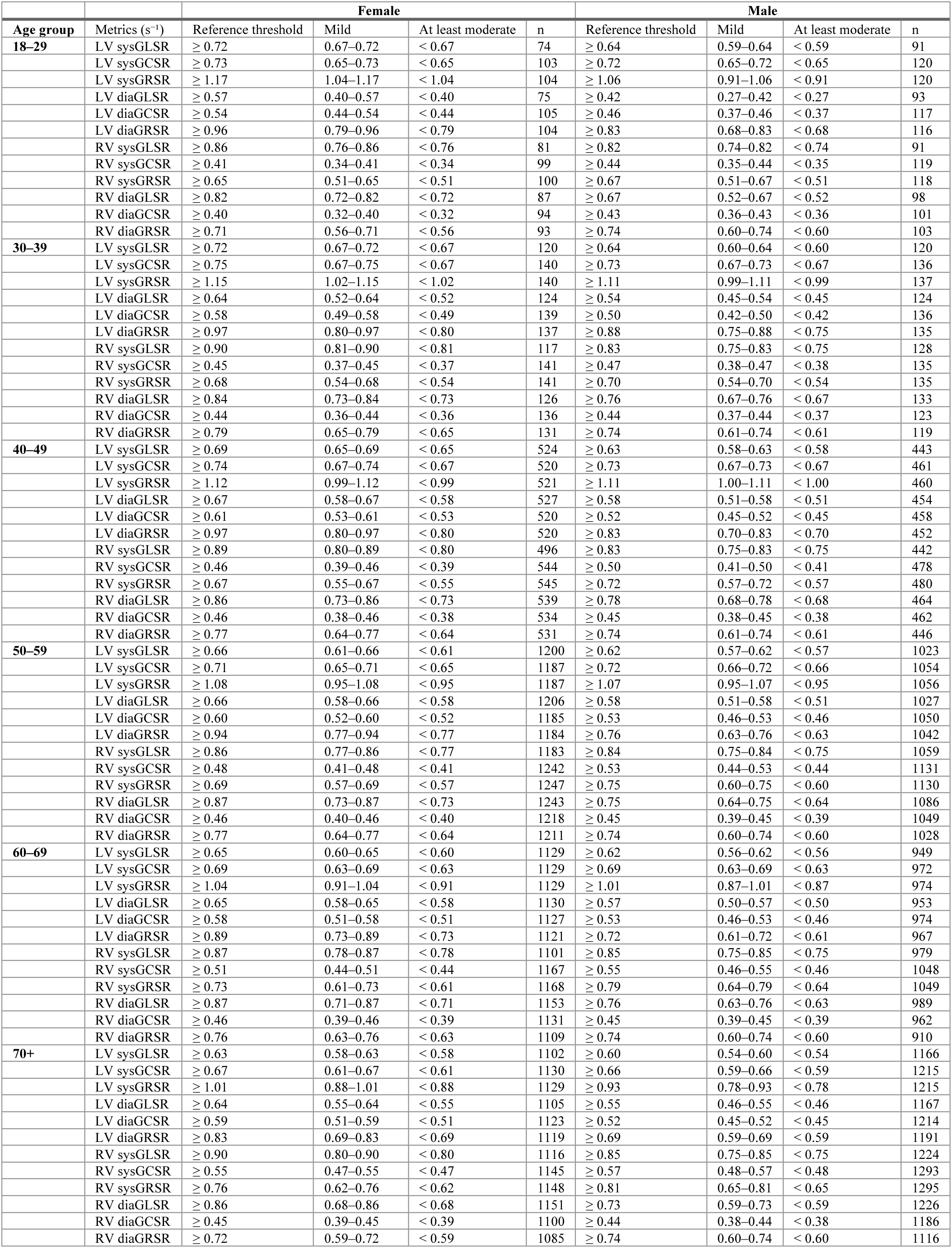
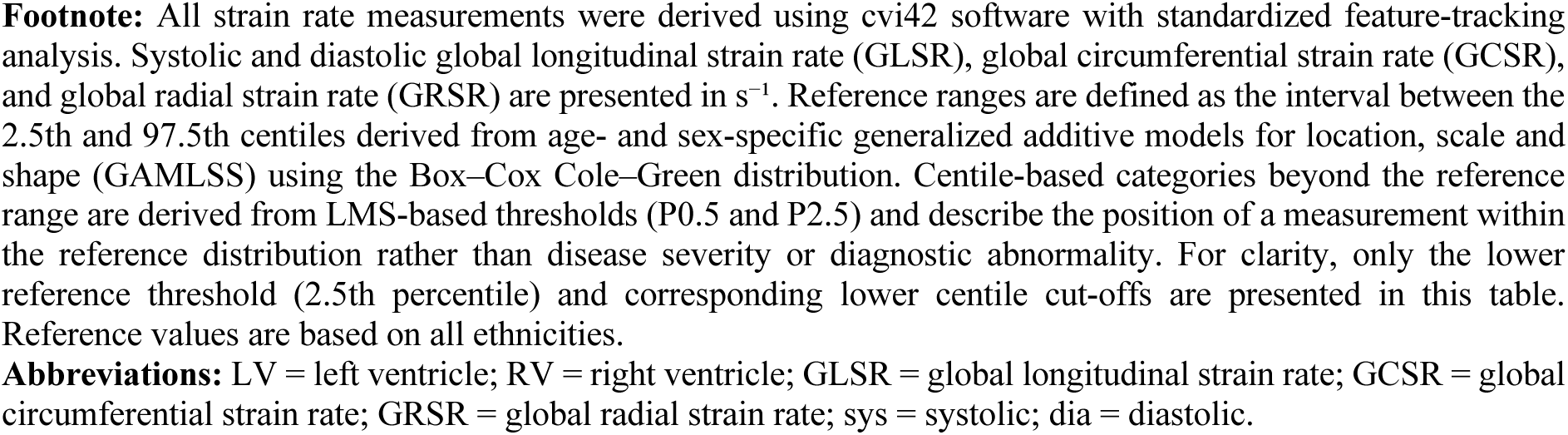
Left and right ventricular global strain rate reference ranges and centile-based categories stratified by sex and age for all ethnicities using cvi42 (Circle Cardiovascular Imaging Inc.)

Analyses using suiteHEART showed broadly consistent sex-specific differences, although age-related patterns were more heterogeneous than those observed with cvi42 (**Supplementary Figure 4–5; Table 5–6**). Women consistently exhibited higher absolute values across ventricular and atrial strain parameters, including LV GLS, RV GLS, LV and RV GRS, and LA and RA strain. In contrast to cvi42, suiteHEART-derived LV GLS was similar across age groups, while RV GLS and radial strain measures slightly higher in older age groups. LV GCS remained largely comparable across age groups. Atrial strain showed chamber-specific differences, with LA strain in older age groups, whereas RA strain was slightly higher in older age groups.

**Table 5.** Left and right ventricular global strain reference ranges and centile-based categories stratified by sex and age for all ethnicities using suiteHEART (NeoSoft)

| Age group | Metrics (%) | Female |  |  |  | Male |  |  |  |
| --- | --- | --- | --- | --- | --- | --- | --- | --- | --- |
|  |  | Reference threshold | Mild | At least moderate | n | Reference threshold | Mild | At least moderate | n |
| 18–29 | LV GLS | ≥ 16.2 | 15.1–16.1 | < 15.1 | 93 | ≥ 14.6 | 13.5–14.5 | < 13.5 | 111 |
|  | LV GCS | ≥ 13.7 | 12.3–13.6 | < 12.3 | 119 | ≥ 13.2 | 12.2–13.1 | < 12.2 | 123 |
|  | LV GRS | ≥ 69.4 | 63.5–69.3 | < 63.5 | 119 | ≥ 56.2 | 50.4–56.1 | < 50.4 | 123 |
|  | RV GLS | ≥ 17.8 | 15.3–17.7 | < 15.3 | 99 | ≥ 15.9 | 13.5–15.8 | < 13.5 | 112 |
|  | RV GCS | ≥ 6.9 | 5.4–6.8 | < 5.4 | 107 | ≥ 5.5 | 3.4–5.4 | < 3.4 | 116 |
|  | RV GRS | ≥ 97.8 | 83.4–97.7 | < 83.4 | 111 | ≥ 79.4 | 69.3–79.3 | < 69.3 | 120 |
| 30–39 | LV GLS | ≥ 16.3 | 14.7–16.2 | < 14.7 | 138 | ≥ 14.9 | 13.8–14.8 | < 13.8 | 133 |
|  | LV GCS | ≥ 13.8 | 12.5–13.7 | < 12.5 | 150 | ≥ 12.9 | 11.8–12.8 | < 11.8 | 148 |
|  | LV GRS | ≥ 65.4 | 59.3–65.3 | < 59.3 | 149 | ≥ 54.4 | 48.5–54.3 | < 48.5 | 148 |
|  | RV GLS | ≥ 18.5 | 15.8–18.4 | < 15.8 | 138 | ≥ 16.4 | 13.8–16.3 | < 13.8 | 133 |
|  | RV GCS | ≥ 6.8 | 5.2–6.7 | < 5.2 | 136 | ≥ 5.4 | 3.4–5.3 | < 3.4 | 142 |
|  | RV GRS | ≥ 87.4 | 72.5–87.3 | < 72.5 | 145 | ≥ 78.0 | 66.2–77.9 | < 66.2 | 145 |
| 40–49 | LV GLS | ≥ 17.5 | 16.2–17.4 | < 16.2 | 535 | ≥ 15.4 | 14.2–15.3 | < 14.2 | 459 |
|  | LV GCS | ≥ 11.1 | 9.1–11.0 | < 9.1 | 567 | ≥ 11.6 | 10.3–11.5 | < 10.3 | 495 |
|  | LV GRS | ≥ 67.3 | 60.4–67.2 | < 60.4 | 567 | ≥ 56.1 | 49.7–56.0 | < 49.7 | 495 |
|  | RV GLS | ≥ 19.0 | 16.1–18.9 | < 16.1 | 524 | ≥ 16.8 | 14.0–16.7 | < 14.0 | 442 |
|  | RV GCS | ≥ 6.2 | 4.5–6.1 | < 4.5 | 495 | ≥ 5.4 | 3.4–5.3 | < 3.4 | 441 |
|  | RV GRS | ≥ 97.2 | 77.8–97.1 | < 77.8 | 555 | ≥ 87.3 | 71.8–87.2 | < 71.8 | 481 |
| 50–59 | LV GLS | ≥ 18.0 | 16.8–17.9 | < 16.8 | 1164 | ≥ 15.7 | 14.3–15.6 | < 14.3 | 1023 |
|  | LV GCS | ≥ 9.1 | 6.7–9.0 | < 6.7 | 1286 | ≥ 10.4 | 8.8–10.3 | < 8.8 | 1145 |
|  | LV GRS | ≥ 70.3 | 62.4–70.2 | < 62.4 | 1286 | ≥ 58.2 | 51.1–58.1 | < 51.1 | 1144 |
|  | RV GLS | ≥ 19.2 | 16.1–19.1 | < 16.1 | 1179 | ≥ 17.1 | 14.1–17.0 | < 14.1 | 952 |
|  | RV GCS | ≥ 5.4 | 3.6–5.3 | < 3.6 | 1103 | ≥ 5.4 | 3.4–5.3 | < 3.4 | 995 |
|  | RV GRS | ≥ 141.1 | 108.6–141.0 | < 108.6 | 1262 | ≥ 113.4 | 91.1–113.3 | < 91.1 | 1116 |
| 60–69 | LV GLS | ≥ 17.6 | 16.2–17.5 | < 16.2 | 1054 | ≥ 15.6 | 14.2–15.5 | < 14.2 | 920 |
|  | LV GCS | ≥ 9.7 | 7.5–9.6 | < 7.5 | 1208 | ≥ 10.5 | 8.9–10.4 | < 8.9 | 1071 |
|  | LV GRS | ≥ 69.0 | 60.4–68.9 | < 60.4 | 1207 | ≥ 58.2 | 50.6–58.1 | < 50.6 | 1069 |
|  | RV GLS | ≥ 19.2 | 15.9–19.1 | < 15.9 | 1058 | ≥ 17.4 | 14.1–17.3 | < 14.1 | 840 |
|  | RV GCS | ≥ 4.9 | 3.0–4.8 | < 3.0 | 993 | ≥ 5.4 | 3.4–5.3 | < 3.4 | 938 |
|  | RV GRS | ≥ 143.7 | 110.8–143.6 | < 110.8 | 1151 | ≥ 117.8 | 94.9–117.7 | < 94.9 | 1045 |
| 70+ | LV GLS | ≥ 17.5 | 16.1–17.4 | < 16.1 | 1030 | ≥ 15.5 | 14.1–15.4 | < 14.1 | 1118 |
|  | LV GCS | ≥ 10.6 | 8.6–10.5 | < 8.6 | 1209 | ≥ 10.8 | 9.1–10.7 | < 9.1 | 1338 |
|  | LV GRS | ≥ 69.5 | 59.7–69.4 | < 59.7 | 1209 | ≥ 57.9 | 49.5–57.8 | < 49.5 | 1338 |
|  | RV GLS | ≥ 19.0 | 15.4–18.9 | < 15.4 | 1022 | ≥ 17.6 | 14.0–17.5 | < 14.0 | 977 |
|  | RV GCS | ≥ 4.9 | 2.9–4.8 | < 2.9 | 1005 | ≥ 5.4 | 3.4–5.3 | < 3.4 | 1180 |
|  | RV GRS | ≥ 155.3 | 123.4–155.2 | < 123.4 | 1152 | ≥ 130.0 | 107.6–129.9 | < 107.6 | 1297 |

**Footnote:** All strain measurements were derived using suiteHEART software with standardized feature-tracking analysis. Global longitudinal strain (GLS), global circumferential strain (GCS), and global radial strain (GRS) are presented as absolute values (%). Reference ranges are defined as the interval between the 2.5th and 97.5th centiles derived from age- and sex-specific generalized additive models for location, scale and shape (GAMLSS) using the Box–Cox Cole–Green distribution. Centile-based categories beyond the reference range are derived from LMS-based thresholds (P0.5 and P2.5) and describe the position of a measurement within the reference distribution rather than disease severity or diagnostic abnormality. For clarity, only the lower reference threshold (2.5th percentile) and corresponding lower centile cut-offs are presented in this table. Reference values are based on all ethnicities.
**Abbreviations:** LV = left ventricle; RV = right ventricle; GLS = global longitudinal strain; GCS = global circumferential strain; GRS = global radial strain.

**Table 6.**
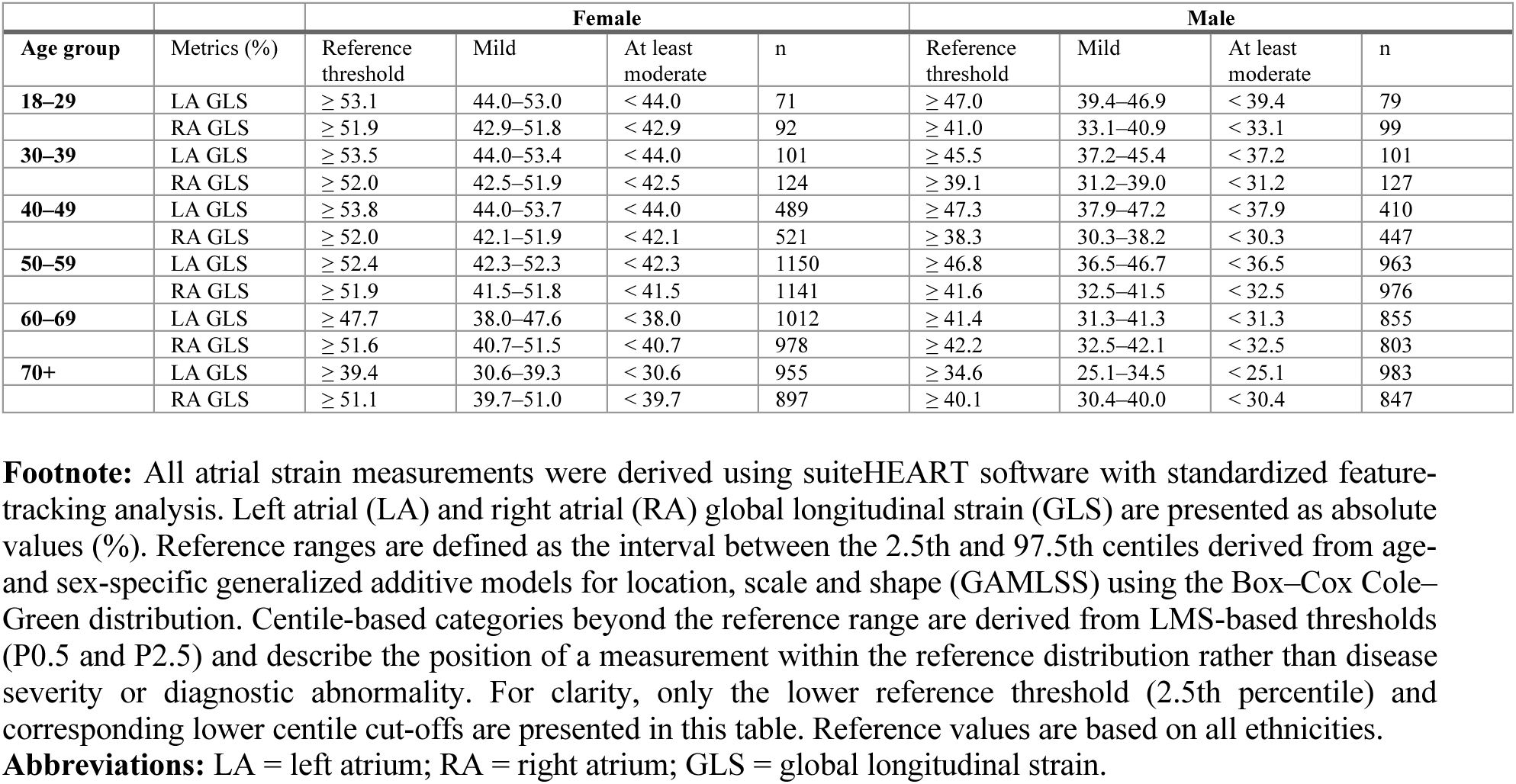
Left and right atrial global strain reference ranges and centile-based categories stratified by sex and age for all ethnicities using suiteHEART (NeoSoft)

| Age group | Metrics (%) | Female |  |  |  | Male |  |  |  |
| --- | --- | --- | --- | --- | --- | --- | --- | --- | --- |
|  |  | Reference threshold | Mild | At least moderate | n | Reference threshold | Mild | At least moderate | n |
| 18–29 | LA GLS | ≥ 53.1 | 44.0–53.0 | < 44.0 | 71 | ≥ 47.0 | 39.4–46.9 | < 39.4 | 79 |
|  | RA GLS | ≥ 51.9 | 42.9–51.8 | < 42.9 | 92 | ≥ 41.0 | 33.1–40.9 | < 33.1 | 99 |
| 30–39 | LA GLS | ≥ 53.5 | 44.0–53.4 | < 44.0 | 101 | ≥ 45.5 | 37.2–45.4 | < 37.2 | 101 |
|  | RA GLS | ≥ 52.0 | 42.5–51.9 | < 42.5 | 124 | ≥ 39.1 | 31.2–39.0 | < 31.2 | 127 |
| 40–49 | LA GLS | ≥ 53.8 | 44.0–53.7 | < 44.0 | 489 | ≥ 47.3 | 37.9–47.2 | < 37.9 | 410 |
|  | RA GLS | ≥ 52.0 | 42.1–51.9 | < 42.1 | 521 | ≥ 38.3 | 30.3–38.2 | < 30.3 | 447 |
| 50–59 | LA GLS | ≥ 52.4 | 42.3–52.3 | < 42.3 | 1150 | ≥ 46.8 | 36.5–46.7 | < 36.5 | 963 |
|  | RA GLS | ≥ 51.9 | 41.5–51.8 | < 41.5 | 1141 | ≥ 41.6 | 32.5–41.5 | < 32.5 | 976 |
| 60–69 | LA GLS | ≥ 47.7 | 38.0–47.6 | < 38.0 | 1012 | ≥ 41.4 | 31.3–41.3 | < 31.3 | 855 |
|  | RA GLS | ≥ 51.6 | 40.7–51.5 | < 40.7 | 978 | ≥ 42.2 | 32.5–42.1 | < 32.5 | 803 |
| 70+ | LA GLS | ≥ 39.4 | 30.6–39.3 | < 30.6 | 955 | ≥ 34.6 | 25.1–34.5 | < 25.1 | 983 |
|  | RA GLS | ≥ 51.1 | 39.7–51.0 | < 39.7 | 897 | ≥ 40.1 | 30.4–40.0 | < 30.4 | 847 |

**Footnote:** All atrial strain measurements were derived using suiteHEART software with standardized feature-tracking analysis. Left atrial (LA) and right atrial (RA) global longitudinal strain (GLS) are presented as absolute values (%). Reference ranges are defined as the interval between the 2.5th and 97.5th centiles derived from age- and sex-specific generalized additive models for location, scale and shape (GAMLSS) using the Box–Cox Cole–Green distribution. Centile-based categories beyond the reference range are derived from LMS-based thresholds (P0.5 and P2.5) and describe the position of a measurement within the reference distribution rather than disease severity or diagnostic abnormality. For clarity, only the lower reference threshold (2.5th percentile) and corresponding lower centile cut-offs are presented in this table. Reference values are based on all ethnicities.
**Abbreviations:** LA = left atrium; RA = right atrium; GLS = global longitudinal strain.

There were also sex differences in excursion-based metrics (**Supplementary Figure 6–7; Table 7; Supplementary Table 35**). Lateral MAPSE was slightly higher in women, whereas septal and inferior MAPSE tended to be higher in men. TAPSE values were broadly comparable between sexes, with marginally higher values in women. Both MAPSE and TAPSE slightly lower in older age groups, particularly in septal and anterior components, consistent lower longitudinal ventricular function in older participants.

**Table 7.**
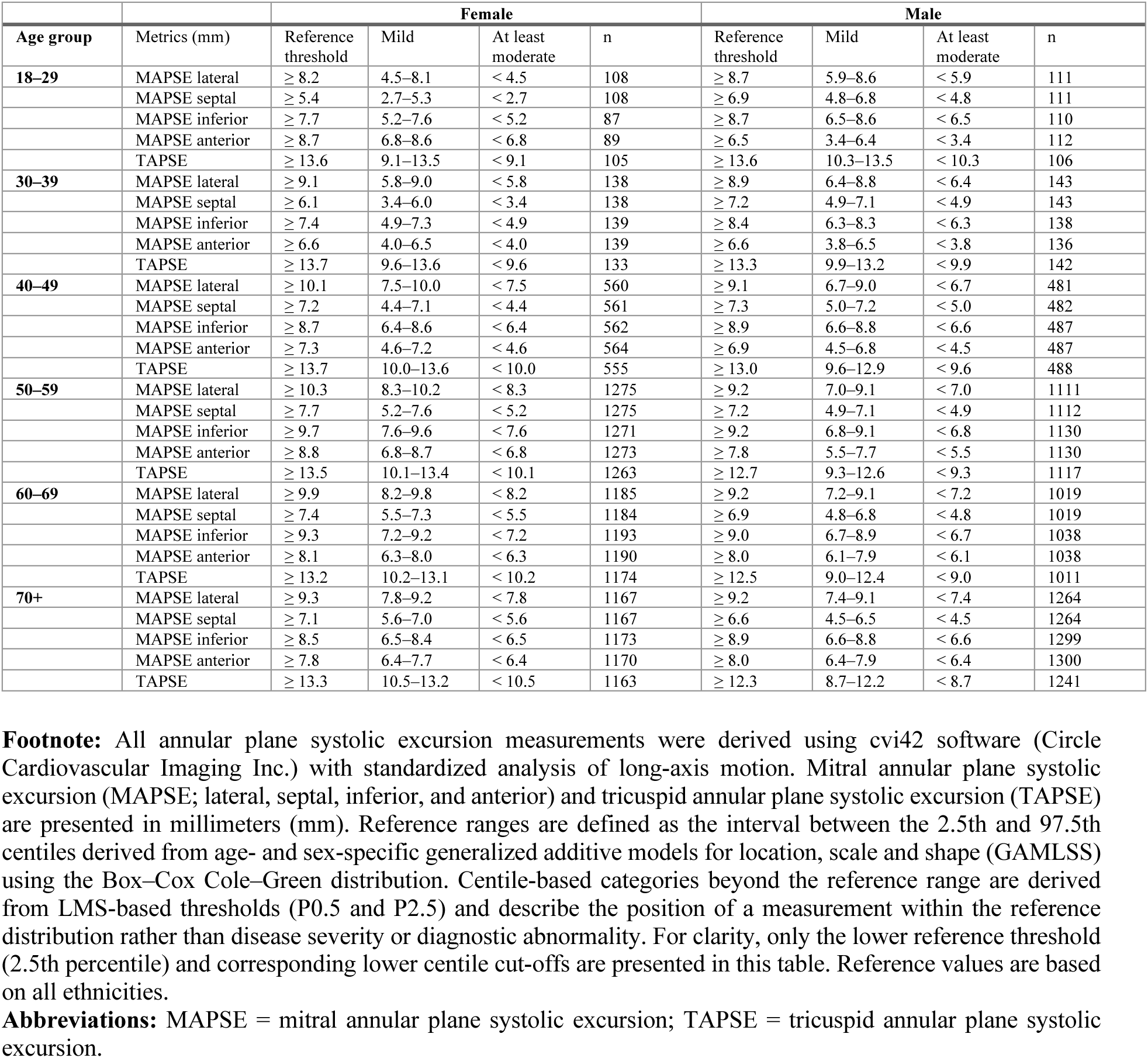
Annular plane systolic excursion metrics reference ranges and centile-based categories stratified by sex and age for all ethnicities using cvi42 (Circle Cardiovascular Imaging Inc.)

Rapid strain metrics demonstrated consistent patterns across parameters (**Supplementary Figure 8–11; Table 8–9**). Women exhibited higher values than men for LV long-axis strain and LV atrioventricular junction strain across all age groups, with these parameters remaining largely similar across age groups. In contrast, atrial rapid strain parameters, particularly right atrial long-axis and atrioventricular junction measures, were lower in older age groups.

**Table 8.**
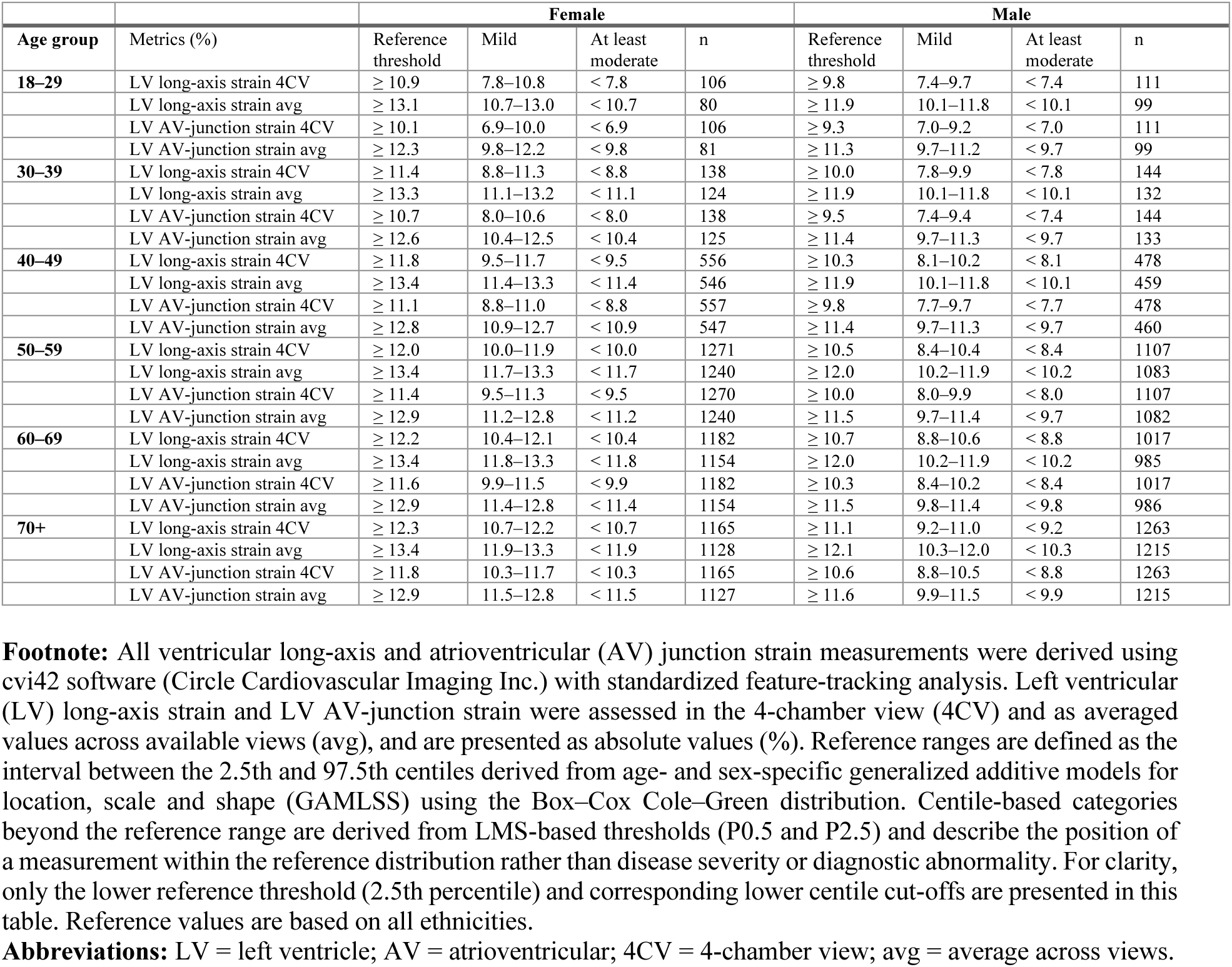
Ventricular long-axis and atrioventricular junction strain metrics reference ranges and centile-based categories stratified by sex and age for all ethnicities using cvi42 (Circle Cardiovascular Imaging Inc.)

**Table 9.**
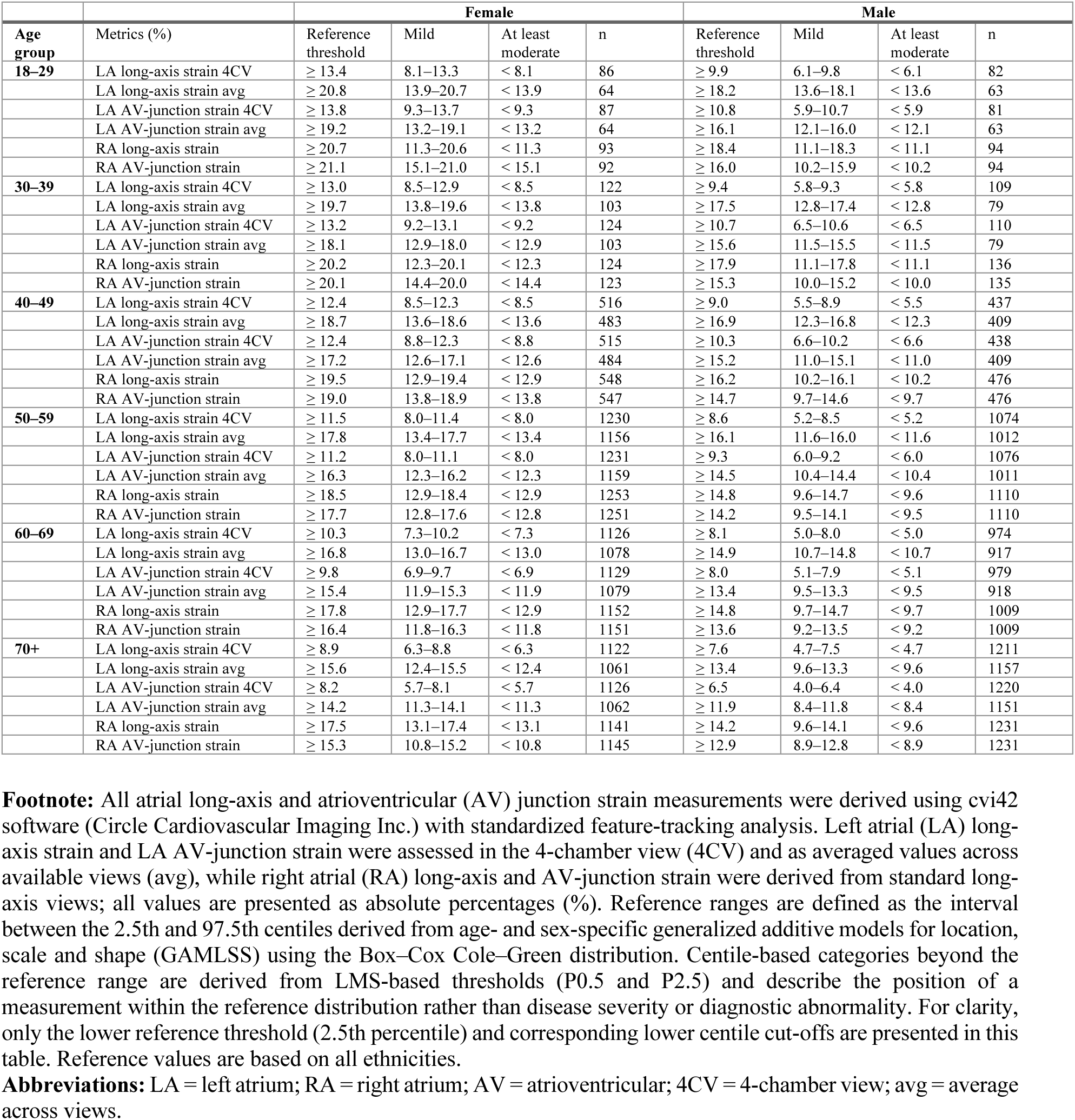
Atrial long-axis and atrioventricular junction strain metrics reference ranges and centile-based categories stratified by sex and age for all ethnicities using cvi42 (Circle Cardiovascular Imaging Inc.)

### Ethnic differences in strain metrics across age and sex

Ethnicity-specific analyses across all modalities (cvi42, suiteHEART, and displacement/rapid strain metrics) demonstrated overall consistent patterns, with only small and heterogeneous differences compared with the White reference group (**Supplementary Figures 12–16**).

For cvi42-derived ventricular strain, values were largely comparable across ethnic groups (**Supplementary Figure 12**). Observed differences were small and without a consistent directional pattern. Black participants showed lower LV GLS and RV strain parameters in the 50–69 age group, whereas Chinese participants exhibited higher radial and circumferential strain in selected age strata below 60 years. South Asian and Mixed/Other groups demonstrated minimal differences overall.

suiteHEART analyses showed broadly similar ethnicity-specific findings and extended the assessment to atrial strain (**Supplementary Figure 13**). Ventricular and atrial strain parameters remained broadly similar across ethnicities, with only minor, non-systematic variations. Annular plane systolic excursion parameters (MAPSE and TAPSE) demonstrated the greatest stability, with no relevant differences across ethnic groups (**Supplementary Figure 14**). Rapid strain metrics showed similarly high consistency across ethnic groups (**Supplementary Figures 15–16**). Small, age- and sex-specific variations were observed, including higher LV long-axis and atrioventricular junction strain values in South Asian men and lower values in older Black participants, but without consistent patterns.

### Scanner vendor and magnetic field strength effects on strain metrics

Using cvi42, LV GLS was similar at 3 T and 1.5 T (p=0.808). In contrast, LV GCS and GRS were slightly, but significantly higher at 3 T. For the RV, GLS was only slightly higher at 3 T, whereas GCS and particularly GRS were higher at 3 T (all p≤0.013). Scanner vendor-related differences were minimal for GLS. Circumferential strain showed small vendor-related differences, while the largest differences were observed for radial strain, with higher values for Philips and lower values for GE compared with Siemens. RV GLS showed no vendor dependence, whereas RV GCS and especially RV GRS were higher for Philips. Overall, cvi42-derived measures were most sensitive for radial strain, while longitudinal strain was relatively robust to field-strength and vendor differences (**Supplementary Figure 17**).

For suiteHEART, magnetic field strength showed parameter-specific effects (**Supplementary Figure 18**). At 3 T, LV GLS and RV GLS were significantly lower, whereas LV GCS and LV GRS were higher; however, these differences were small in magnitude and likely of limited clinical relevance. RV GCS was slightly higher at 3 T. In contrast, RV GRS was substantially lower at 3 T, representing the most pronounced field strength–related difference. Atrial strain exhibited a differential pattern, with significantly lower LA GLS, while RA GLS remained unchanged. Vendor effects were more pronounced and less uniform. Both Philips and GE showed lower LV GLS and higher LV GCS compared with Siemens, while LV GRS varied across vendors. Notably, RV GRS was lower for Philips and GE compared with Siemens, representing the most pronounced vendor-related difference. In addition, LA GLS was lower for both Philips and GE, highlighting its sensitivity to vendor-related variability. Overall, vendor effects were most relevant for RV GRS and LA GLS, whereas other parameters showed smaller or less consistent differences (**Supplementary Figure 18**).

Magnetic field strength had limited impact on annular plane systolic excursion parameters, with similar values for lateral MAPSE and TAPSE at 1.5 T and 3 T. Rapid strain parameters were generally lower at 3 T for LV long-axis and AV-junction strain and for LA strain, whereas RA strain was higher, although this difference was likely of limited clinical relevance (**Supplementary Figure 19**).

Vendor effects for displacement measures were consistent, with lower MAPSE and TAPSE for Philips and partly for GE. For rapid strain, both vendors showed lower LV and LA values, most pronounced for LA parameters, while RA strain showed no consistent vendor dependence. Overall, displacement measures were relatively stable, whereas scanner-related differences in rapid strain were mainly parameter-specific and most evident for atrial measures (**Supplementary Figure 19**).

### Comparison of software solutions

Agreement between cvi42 and suiteHEART varied by strain component (**Supplementary Figure 20; Supplementary Table 3**). For longitudinal strain, suiteHEART values were higher than cvi42 (LV GLS bias +2.7, 95% limits of agreement [−0.4, 5.8], r = 0.72; RV GLS bias +1.4 [−7.0, 9.7], r = 0.39). For circumferential strain, suiteHEART values were lower than cvi42 (LV GCS bias −2.8 [−7.7, 2.2], r = 0.48; RV GCS bias −2.6 [−10.8, 5.6], r = 0.39). For radial strain, suiteHEART values were several-fold higher than cvi42 (LV GRS 97.3 vs 31.1, bias +66.2, r = 0.66; RV GRS 275.0 vs 26.0, bias +249.0, r = −0.01).

### Reference ranges and centile-based categories for CMR-derived strain parameters across age, sex, and ethnicity

Age- and sex-specific reference ranges and centile-based categories for CMR-derived strain parameters are summarized in **Tables 3–9**.

For ventricular global strain derived with cvi42 (**Table 3**), LV GLS, LV GCS, LV GRS, and RV GLS are presented across all age groups stratified by sex. Reference ranges for ventricular global strain rate parameters derived with cvi42 are shown in **Table 4**, including systolic and diastolic strain rate values for LV and RV metrics.

For ventricular global strain derived with suiteHEART (**Table 5**), LV and RV strain parameters are reported across age and sex groups. Atrial strain reference ranges derived with suiteHEART are presented in **Table 6**, including LA and RA GLS.

Reference ranges for annular plane systolic excursion metrics are presented in **Table 7**, including MAPSE (lateral, septal, inferior, anterior) and TAPSE across age and sex groups.

**Tables 8 and 9** summarize reference ranges for rapid strain parameters derived with cvi42. **Table 8** includes left ventricular long-axis and atrioventricular junction strain metrics, while **Table 9** presents corresponding atrial long-axis and atrioventricular junction strain parameters. Ethnicity-specific reference ranges for all strain, strain rate, displacement, velocity, and rapid strain parameters are provided in the **Supplementary Tables (Supplementary Tables 6–51)**, with corresponding visualization in **Supplementary Figures 12–16**.

### Prognostic associations of centile-based CMR strain categories

Centile-based CMR strain categories showed the strongest associations with incident heart failure. Values in the at least moderately out-of-range lower-tail category (<0.5th centile) were associated with markedly higher heart failure risk across LV GLS, LV GCS, and LV GRS, with hazard ratios ranging from 9.86 for LV GLS (95% CI: 7.00–13.90) to 12.38 for LV GRS (95% CI: 8.49–18.06). Lateral MAPSE and selected RV strain metrics were also associated with incident heart failure in this category (RV GCS HR 4.72, 95% CI 2.33–9.58; RV GRS HR 6.27, 95% CI 3.64–10.79; lateral MAPSE HR 5.82, 95% CI 3.81–8.89).

Associations with myocardial infarction were less pronounced but remained evident for LV strain metrics. In the at least moderately out-of-range category, LV GLS, LV GCS, and LV GRS were associated with higher myocardial infarction risk (HRs 3.31, 2.88, and 2.83, respectively). Mild reductions in LV GCS and LV GRS were also associated with higher risk (HRs 3.17 and 3.00, respectively). Among RV parameters, only at least moderately out-of-range RV GCS showed a significant association (HR 2.84, 95% CI 1.17–6.90).

For stroke, significant associations were mainly observed for LV strain metrics. LV GLS was associated with stroke risk in both the mildly out-of-range (HR 2.18, 95% CI 1.15–4.14) and at least moderately out-of-range categories (HR 2.28, 95% CI 1.19–4.34), whereas LV GCS and LV GRS showed significant associations only in the at least moderately out-of-range category (HR 3.77, 95% CI 1.91–7.45 and HR 3.70, 95% CI 1.93–7.07, respectively). Lateral MAPSE was also associated with stroke in the at least moderately out-of-range category (HR 2.27, 95% CI 1.16–4.47)

Associations with all-cause death were more modest. Significant associations in the at least moderately out-of-range category were observed for LV GLS (HR 1.60, 95% CI 1.01–2.55), LV GRS (HR 2.11, 95% CI 1.28–3.50), RV GLS (HR 2.21, 95% CI 1.24–3.93), and lateral MAPSE (HR 1.94, 95% CI 1.22–3.08), while mild reductions were not significantly associated with all-cause death.

Overall, centile-based CMR strain categories identified participants at increased risk of incident clinical outcomes, with the most consistent and largest effect sizes observed for incident heart failure. Detailed hazard ratios, confidence intervals, p-values, model sample sizes, and event counts are presented in **Table 10**, **Figure 3 and 4**.

**Figure 3.**
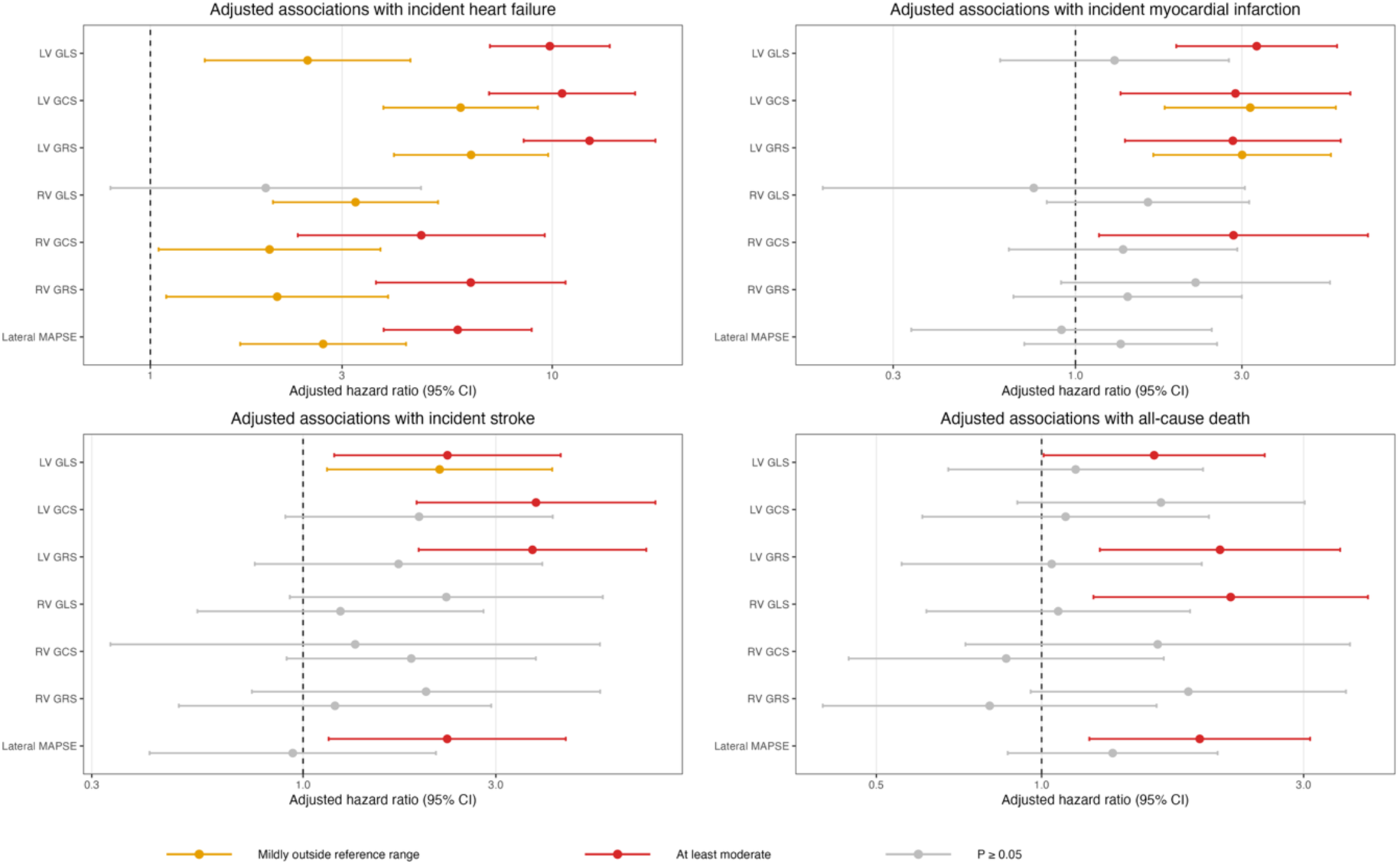
Associations between cardiovascular magnetic resonance feature tracking (CMR-FT) strain categories derived using cvi42 and incident clinical outcomes. Forest plots show adjusted hazard ratios (HRs) and 95% confidence intervals (CIs) for incident heart failure, myocardial infarction, stroke, and all-cause death according to centile-based cardiovascular magnetic resonance feature tracking (CMR-FT) strain categories derived using cvi42 (Circle Cardiovascular Imaging Inc.). HRs were derived from Cox proportional hazards models adjusted for age, sex, and body mass index at imaging, using the within-reference-range category as the reference group. Mildly outside reference range denotes values outside the reference range but above the at-least-moderate threshold. Models were fitted separately for each CMR metric. The vertical dashed line indicates HR = 1. Grey markers indicate associations with P ≥ 0.05. **Abbreviations:** BMI = body mass index; CI = confidence interval; CMR-FT = cardiovascular magnetic resonance feature tracking; GCS = global circumferential strain; GLS = global longitudinal strain; GRS = global radial strain; HF = heart failure; HR = hazard ratio; LV = left ventricular; MAPSE = mitral annular plane systolic excursion; MI = myocardial infarction; RV = right ventricular.

**Figure 4.**
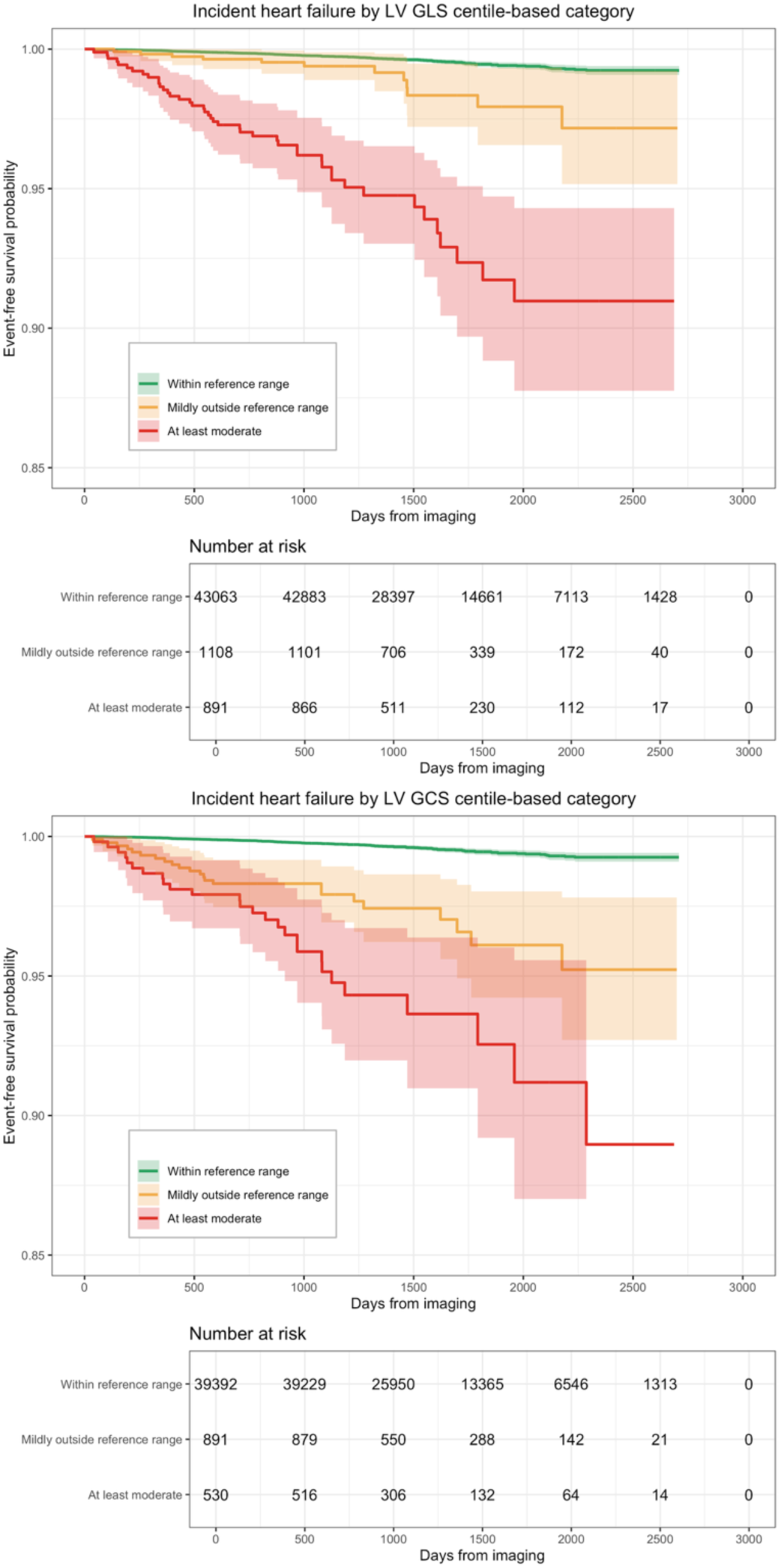
Kaplan–Meier curves for incident heart failure according to centile-based cardiovascular magnetic resonance feature tracking (CMR-FT) left ventricular strain categories. Kaplan–Meier curves showing event-free survival for incident heart failure according to centile-based left ventricular (LV) global circumferential strain (GCS) and global longitudinal strain (GLS) categories derived from cardiovascular magnetic resonance feature tracking (CMR-FT) using cvi42 (Circle Cardiovascular Imaging Inc.). Participants were classified as within reference range, mildly outside reference range, or at least moderately outside reference range according to age- and sex-specific centile thresholds. Shaded areas represent 95% confidence intervals. Numbers at risk are shown below each panel. **Abbreviations:** GCS = global circumferential strain; GLS = global longitudinal strain; LV = left ventricular.

**Table 10.**
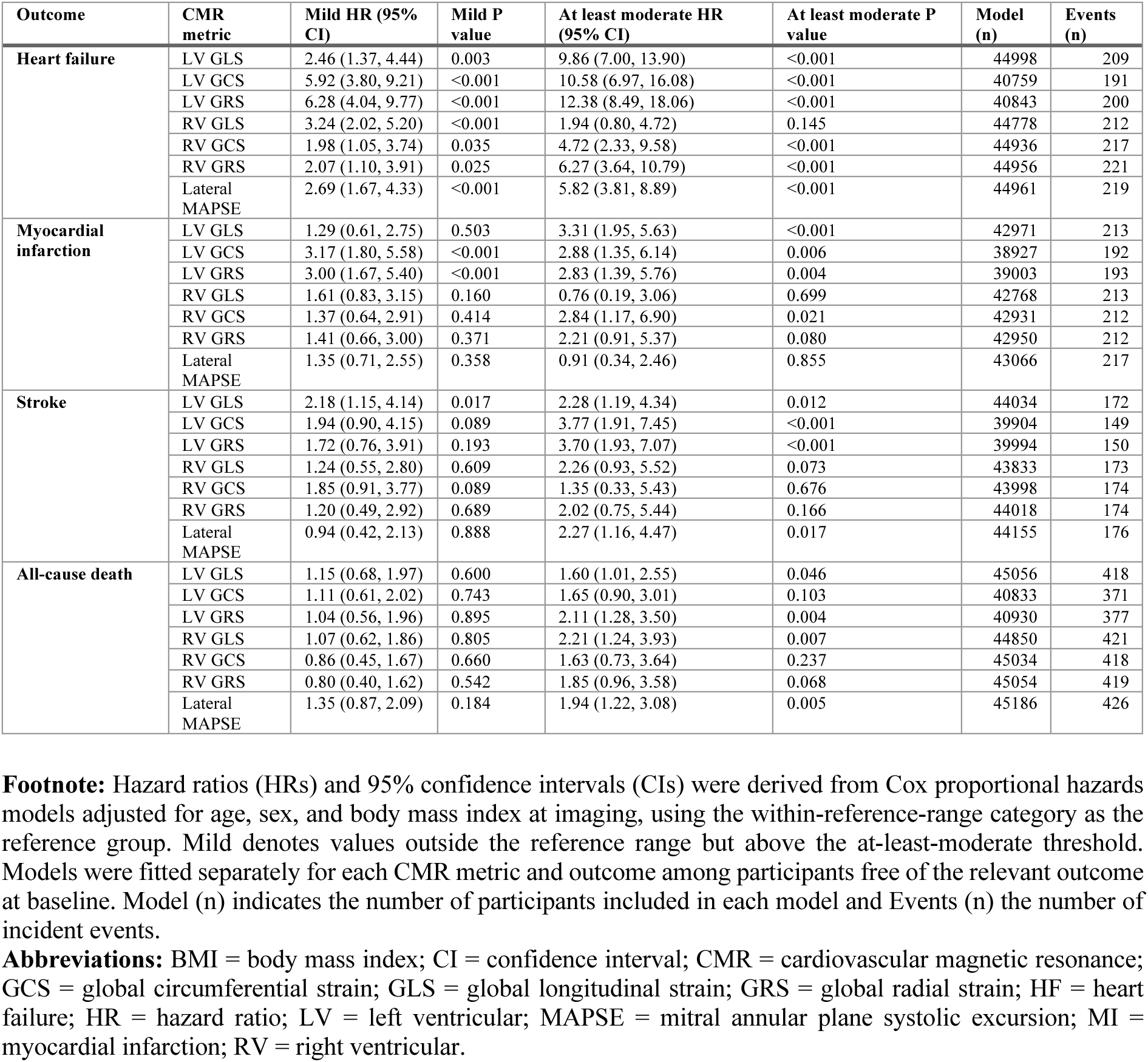
Adjusted associations between centile-based CMR strain categories and incident clinical outcomes.

| Outcome | CMR metric | Mild HR (95% CI) | Mild P value | At least moderate HR (95% CI) | At least moderate P value | Model (n) | Events (n) |
| --- | --- | --- | --- | --- | --- | --- | --- |
| Heart failure | LV GLS | 2.46 (1.37, 4.44) | 0.003 | 9.86 (7.00, 13.90) | <0.001 | 44998 | 209 |
|  | LV GCS | 5.92 (3.80, 9.21) | <0.001 | 10.58 (6.97, 16.08) | <0.001 | 40759 | 191 |
|  | LV GRS | 6.28 (4.04, 9.77) | <0.001 | 12.38 (8.49, 18.06) | <0.001 | 40843 | 200 |
|  | RV GLS | 3.24 (2.02, 5.20) | <0.001 | 1.94 (0.80, 4.72) | 0.145 | 44778 | 212 |
|  | RV GCS | 1.98 (1.05, 3.74) | 0.035 | 4.72 (2.33, 9.58) | <0.001 | 44936 | 217 |
|  | RV GRS | 2.07 (1.10, 3.91) | 0.025 | 6.27 (3.64, 10.79) | <0.001 | 44956 | 221 |
|  | Lateral MAPSE | 2.69 (1.67, 4.33) | <0.001 | 5.82 (3.81, 8.89) | <0.001 | 44961 | 219 |
| Myocardial infarction | LV GLS | 1.29 (0.61, 2.75) | 0.503 | 3.31 (1.95, 5.63) | <0.001 | 42971 | 213 |
|  | LV GCS | 3.17 (1.80, 5.58) | <0.001 | 2.88 (1.35, 6.14) | 0.006 | 38927 | 192 |
|  | LV GRS | 3.00 (1.67, 5.40) | <0.001 | 2.83 (1.39, 5.76) | 0.004 | 39003 | 193 |
|  | RV GLS | 1.61 (0.83, 3.15) | 0.160 | 0.76 (0.19, 3.06) | 0.699 | 42768 | 213 |
|  | RV GCS | 1.37 (0.64, 2.91) | 0.414 | 2.84 (1.17, 6.90) | 0.021 | 42931 | 212 |
|  | RV GRS | 1.41 (0.66, 3.00) | 0.371 | 2.21 (0.91, 5.37) | 0.080 | 42950 | 212 |
|  | Lateral MAPSE | 1.35 (0.71, 2.55) | 0.358 | 0.91 (0.34, 2.46) | 0.855 | 43066 | 217 |
| Stroke | LV GLS | 2.18 (1.15, 4.14) | 0.017 | 2.28 (1.19, 4.34) | 0.012 | 44034 | 172 |
|  | LV GCS | 1.94 (0.90, 4.15) | 0.089 | 3.77 (1.91, 7.45) | <0.001 | 39904 | 149 |
|  | LV GRS | 1.72 (0.76, 3.91) | 0.193 | 3.70 (1.93, 7.07) | <0.001 | 39994 | 150 |
|  | RV GLS | 1.24 (0.55, 2.80) | 0.609 | 2.26 (0.93, 5.52) | 0.073 | 43833 | 173 |
|  | RV GCS | 1.85 (0.91, 3.77) | 0.089 | 1.35 (0.33, 5.43) | 0.676 | 43998 | 174 |
|  | RV GRS | 1.20 (0.49, 2.92) | 0.689 | 2.02 (0.75, 5.44) | 0.166 | 44018 | 174 |
|  | Lateral MAPSE | 0.94 (0.42, 2.13) | 0.888 | 2.27 (1.16, 4.47) | 0.017 | 44155 | 176 |
| All-cause death | LV GLS | 1.15 (0.68, 1.97) | 0.600 | 1.60 (1.01, 2.55) | 0.046 | 45056 | 418 |
|  | LV GCS | 1.11 (0.61, 2.02) | 0.743 | 1.65 (0.90, 3.01) | 0.103 | 40833 | 371 |
|  | LV GRS | 1.04 (0.56, 1.96) | 0.895 | 2.11 (1.28, 3.50) | 0.004 | 40930 | 377 |
|  | RV GLS | 1.07 (0.62, 1.86) | 0.805 | 2.21 (1.24, 3.93) | 0.007 | 44850 | 421 |
|  | RV GCS | 0.86 (0.45, 1.67) | 0.660 | 1.63 (0.73, 3.64) | 0.237 | 45034 | 418 |
|  | RV GRS | 0.80 (0.40, 1.62) | 0.542 | 1.85 (0.96, 3.58) | 0.068 | 45054 | 419 |
|  | Lateral MAPSE | 1.35 (0.87, 2.09) | 0.184 | 1.94 (1.22, 3.08) | 0.005 | 45186 | 426 |

**Footnote:** Hazard ratios (HRs) and 95% confidence intervals (CIs) were derived from Cox proportional hazards models adjusted for age, sex, and body mass index at imaging, using the within-reference-range category as the reference group. Mild denotes values outside the reference range but above the at-least-moderate threshold. Models were fitted separately for each CMR metric and outcome among participants free of the relevant outcome at baseline. Model (n) indicates the number of participants included in each model and Events (n) the number of incident events.
**Abbreviations:** BMI = body mass index; CI = confidence interval; CMR = cardiovascular magnetic resonance; GCS = global circumferential strain; GLS = global longitudinal strain; GRS = global radial strain; HF = heart failure; HR = hazard ratio; LV = left ventricular; MAPSE = mitral annular plane systolic excursion; MI = myocardial infarction; RV = right ventricular.

## Discussion

### Overall findings

In this large multicenter healthy cohort, age and sex emerged as the dominant biological determinants of CMR-derived deformation metrics across ventricular, atrial, rapid strain, and annular plane systolic excursion parameters. Ethnicity-related differences were modest and substantially smaller. Technical factors, including magnetic field strength, scanner vendor, exerted parameter-specific effects, with the greatest susceptibility observed for radial strain. Longitudinal strain and annular plane systolic excursion metrics appeared comparatively robust across acquisition settings, whereas radial strain was more sensitive to technical variation.

### Sex-related differences

Sex-related differences were consistent across most deformation metrics, with women demonstrating higher absolute ventricular and atrial strain values than men. This pattern was observed across both software platforms and extended to rapid strain parameters, supporting the concept of sex-specific myocardial functional phenotypes. Sex-related differences appeared broadly preserved across age groups, without clear attenuation in older women. This may reflect the healthy cohort design and persistent sex-related differences in cardiac size, geometry, and loading conditions. However, menopausal status and hormone replacement therapy were not available and could therefore not be assessed directly. These findings are consistent with prior CMR studies reporting higher strain magnitudes in women [11,17,35].

### Differences across age groups

Age-related differences in ventricular strain were generally modest and parameter specific. In agreement with prior studies, longitudinal strain remained largely stable across age groups, whereas circumferential and radial strain demonstrated greater variability, with some studies reporting higher strain magnitude in older participants [11,17,19]. The relative preservation of GLS in older age groups may reflect the healthy reference cohort design and lower GLS in older individuals may be more apparent in cohorts with higher cardiovascular risk burden or established disease. For the RV, longitudinal deformation remained stable, while circumferential and radial components showed modest, age-group differences, in line with meta-analysis data [36].

### Atrial strain

Atrial strain showed clear age- and sex-specific patterns. Women consistently exhibited higher LA and RA strain values. Potential contributors include sex-related differences in cardiac size, geometry, and loading conditions, although these mechanisms were not directly tested in the present study. In line with our findings, Gao et al. reported higher RA reservoir, conduit, and booster strain values in women compared with men [37].

Across age groups, LA strain demonstrated the most consistent differences, with lower values in older participants. This finding has been reported across multiple CMR and echocardiographic studies, reflecting progressive alterations in atrial compliance and reservoir function across age groups [19,38]. In our cohort, this was particularly evident for LA GLS, with a pronounced decrease in the lower reference threshold with advancing age.

In contrast, RA strain showed only limited differences across age groups. This may suggest that RA deformation is less consistently related to age than LA deformation. However, technical factors, including thinner atrial walls, complex RA geometry, and greater tracking variability, may also contribute. Therefore, RA strain findings should be interpreted cautiously.

### Annular plane systolic excursion measurements

Excursion-based metrics showed only modest sex-related differences. MAPSE demonstrated small but anatomical-region-specific variations between sexes, whereas TAPSE values were largely comparable, indicating limited sex dependency of displacement-based measures. Across age groups, MAPSE values were lower in older participants, particularly affecting anterior and septal components, while TAPSE remained comparatively stable with only mild reductions at older ages.

These findings are consistent with prior CMR studies demonstrating an overall age-dependent decrease in cardiac valve plane displacement with marked regional variability and minimal sex-related differences [39,40].

These findings can be interpreted in the context of structural remodeling at the atrioventricular junction. Prior CMR data have demonstrated progressive changes in annular geometry with age, including enlargement of the tricuspid annulus and alterations in mitral leaflet tethering [41]. Such structural adaptations likely contribute to the observed reduction in longitudinal shortening, linking annular remodeling to functional impairment ^42,43^.

The relative preservation of TAPSE despite structural changes of the tricuspid annulus suggests that right ventricular longitudinal mechanics may be maintained through compensatory mechanisms or distinct myocardial aging. Differences in exposure to systemic afterload and other systemic stressors, compared with the left heart, may also contribute, although this remains speculative.

### Ethnicity-related differences

Ethnicity-related differences in cardiac structure and function have been reported previously, particularly in smaller or more selective cohorts. In a healthy UK Biobank sample, Parke et al. observed ethnic differences between South Asian and White European participants, including higher circumferential strain and partly higher longitudinal strain in South Asian individuals, although these effects were not uniform across sexes or strain components [13]. Our study extends this previous work in several respects. We included a larger number of South Asian participants from UK Biobank and focused primarily on deriving age- and sex-specific reference ranges rather than solely comparing South Asian and White participants. We also examined additional ethnic groups, including Black, Chinese, and Mixed/Other participants. The inclusion of the Singapore cohort further increased the representation of Chinese participants, although other ethnic groups were available predominantly from UK Biobank. Across modalities, ethnicity-related differences were modest and heterogeneous. Ventricular and atrial strain parameters were broadly comparable across ethnic groups, with no consistent pattern of deviation. Rapid strain, MAPSE, and TAPSE measures also demonstrated high stability across ethnic groups.

Overall, ethnicity-related variation was small supporting the use of unified reference ranges for most deformation metrics while acknowledging that subtle subgroup differences may require further investigation.

### Influence of magnetic field strength and scanner vendor

Magnetic field strength showed parameter-specific effects on CMR-derived deformation metrics in our study. In line with previous reports, no relevant differences were observed for GLS and only minor effects for GCS between 1.5 T and 3 T [42,43]. However, particularly radial strain values were higher at 3 T, with the most pronounced differences observed for RV GRS across both software solutions (cvi42 and suiteHEART). These findings extend previous meta-analytic observations. Yang et al. identified field strength as a contributor to variability in LV GLS [14]. Our data additionally demonstrate higher radial strain at 3 T, suggesting that magnetic field strength-related effects may become more apparent in larger cohorts and under standardized analysis conditions. The marked susceptibility of radial strain may be explained by its greater dependence on transmural tracking and endocardial border definition, rendering it particularly sensitive to image characteristics such as signal-to-noise ratio, spatial resolution, and susceptibility artifacts, which differ between 1.5 T and 3 T acquisitions. These findings have direct implications for both clinical interpretation and multicenter research, as radial strain values should be interpreted with caution across different field strengths, and pooling without harmonization may introduce systematic bias.

Scanner vendor–related differences were parameter- and software-dependent. Across both platforms, radial strain was consistently the most affected parameter, with the largest differences, highlighting its susceptibility to technical factors.

Importantly, RV GRS showed the largest software-related discrepancy, with several-fold higher values using suiteHEART than cvi42 and essentially no correlation between platforms. This finding indicates that RV radial strain is highly dependent on the post-processing pipeline and should not be used interchangeably across software solutions. Software-specific reference ranges and consistent use of the same platform are therefore essential.

### Prognostic relevance of centile-based strain categories

The age- and sex-specific centile categories showed prognostic associations with subsequent clinical outcomes, most consistently for incident heart failure. LV GLS, GCS, and GRS values below the reference range were associated with higher heart failure risk, with the largest effect estimates below the 0.5th centile. To our knowledge, this is the first study to demonstrate the prognostic relevance of FT-CMR strain using age- and sex-specific, centile-based categories derived from a healthy reference population. Whereas prior work established the prognostic value of CMR-derived strain as a continuous measure[9], our findings suggest that classifying individuals relative to their expected reference distribution captures clinically meaningful risk beyond uniform strain thresholds.

These associations are consistent with previous studies of CMR-derived myocardial deformation [6,9]. Significant associations were also observed for selected RV strain measures and lateral MAPSE, suggesting that both ventricular deformation and longitudinal annular displacement may reflect early functional impairment. Findings for myocardial infarction, stroke, and all-cause death were less consistent across parameters and centile categories.

The relatively small number of events within the extreme centile categories likely contributed to wider confidence intervals. Further validation in independent cohorts, and assessment of incremental prognostic value beyond conventional CMR measures and established cardiovascular risk factors, are required.

### Clinical implications

In this large, multicenter cohort of healthy individuals, GLS emerged as the most robust and reproducible strain parameter across different scanner settings and vendors. These findings support the use of GLS as the preferred strain metric for both clinical routine and multicenter research settings. Its relative stability suggests that GLS is less susceptible to technical variability, thereby facilitating comparability across institutions and enhancing its suitability for longitudinal follow-up and large-scale cohort studies.

These findings highlight the ongoing need for standardization in CMR strain analysis. The implementation of harmonized acquisition protocols and post-processing pipelines remains essential to minimize technical variability. The present study addresses one component of this need by providing software-specific reference ranges derived independently with cvi42 and suiteHEART. Further development and validation of vendor-specific reference values may additionally improve the accuracy and clinical interpretability of strain measurements. Collectively, such efforts are critical to enable reliable integration of strain metrics into clinical decision-making and to ensure consistency in multicenter research environments.

### Strengths

This study includes a large, multicenter cohort with broad age and ethnic representation. A major strength is the comprehensive range of deformation parameters covered, including ventricular and atrial strain, strain rate, rapid strain, and annular plane systolic excursion metrics. Multiple software platforms were analyzed, allowing assessment across two independent post-processing solutions. Rigorous quality control supports robustness of the derived reference values. In addition, the LMS-based centile framework enables robust estimation of age- and sex-specific reference ranges and centile-based categories. The approach uses routinely acquired cine CMR images and does not require dedicated strain sequences. It can therefore be integrated into existing clinical and research workflows and provide additional functional information from standard cine images with relatively efficient post-processing.

### Limitations

This study has several limitations. Although multiple ethnic groups were included, some were underrepresented, and younger adults were less well represented than middle-aged and older participants. Future studies with larger and more balanced multi-ethnic cohorts and greater representation of younger age groups are needed to validate and refine ethnicity-specific reference ranges.

The derivation of centile-based categories for out-of-range values – particularly the “at least moderate” range – should be interpreted with caution. These thresholds are based on degree of deviation from the reference range calculated from a healthy population, and are distinct from grading of severity within specific disease entities (e.g. heart failure).

The exploration of technical confounders, including scanner vendor, magnetic field strength, and software is complex. Although systematic effects were identified, residual interactions between acquisition and post-processing factors cannot be fully disentangled and may influence the observed variability.

## Conclusion

This large multicenter study provides age- and sex-specific reference ranges for CMR-derived myocardial deformation using an LMS-based approach. Age and sex and not ethnicity were the main determinants. GLS emerged as the most robust parameter across scanner settings, supporting its use in clinical practice and multicenter studies.

Importantly, the application of centile-based thresholds enables a standardized, clinically applicable categorization of myocardial function, including graded classification of out-of-range values. This approach facilitates more consistent interpretation and may support risk stratification and clinical decision-making. However, these centile-based categories reflect statistical deviation from the healthy reference distribution rather than disease-specific severity grades, and their clinical interpretation may benefit from further evaluation in relevant patient populations.

## Supporting information

Supplementary_material_reference_values_strain_HHC

## Data Availability

All data produced in the present study are available upon reasonable request to the authors

https://healthy-hearts.org.uk

## Abbreviations

AV: Atrioventricular
AVJS: Atrioventricular Junction Strain
BSA: Body Surface Area
CMR: Cardiovascular Magnetic Resonance
FT-CMR: Feature-Tracking Cardiovascular Magnetic Resonance
GCD: Global Circumferential Displacement
GCS: Global Circumferential Strain
GCSR: Global Circumferential Strain Rate
GLD: Global Longitudinal Displacement
GLS: Global Longitudinal Strain
GLSR: Global Longitudinal Strain Rate
GLV: Global Longitudinal Velocity
GRD: Global Radial Displacement
GRS: Global Radial Strain
GRSR: Global Radial Strain Rate
GRV: Global Radial Velocity
HHC: Healthy Hearts Consortium
LAS: Long Axis Strain
LA: Left Atrial
LV: Left Ventricular
MAPSE: Mitral Annular Plane Systolic Excursion
QC: Quality Control
RA: Right Atrial
RV: Right Ventricular
TAPSE: Tricuspid Annular Plane Systolic Excursion

## Additional information

### Ethics statement

This study was conducted in accordance with the principles of the Declaration of Helsinki. Analysis of the UK Biobank dataset was performed under approval from the NHS National Research Ethics Service (Ref 11/NW/0382; extension Ref 21/NW/0157), with written informed consent obtained from all participants. For all other contributing cohorts, local ethical approval and participant consent procedures were confirmed by the respective institutions.

---

**Central illustration.**
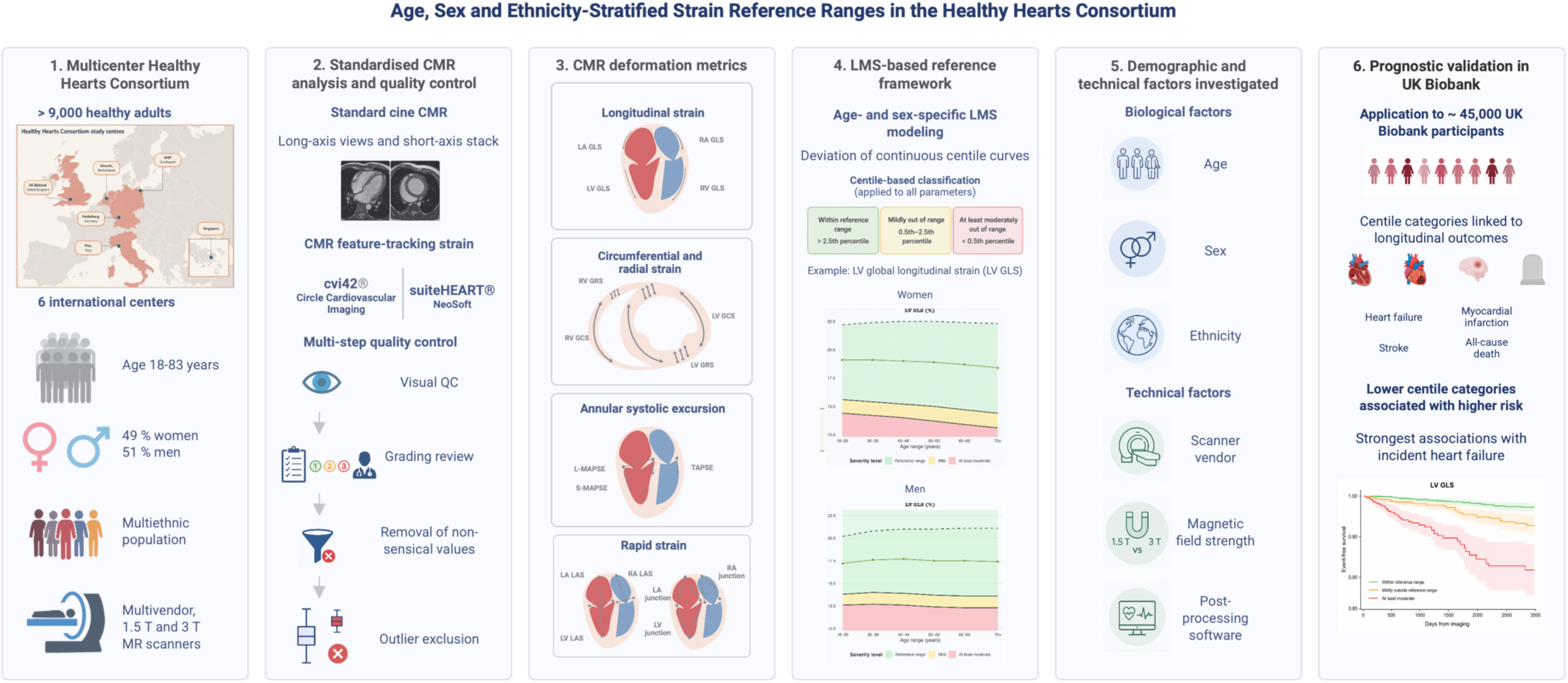
**(1)** The Healthy Hearts Consortium included more than 9,000 healthy adults aged 18–83 years from six international centers, comprising a multiethnic population examined using multivendor 1.5-T and 3-T CMR scanners. **(2)** Standard cine CMR long-axis and short-axis images were analyzed using cvi42 and suiteHEART. A multistep quality-control procedure included visual assessment, grading review, removal of non-physiological values, and outlier exclusion. **(3)** Derived deformation parameters included ventricular and atrial longitudinal, circumferential, and radial strain, annular plane systolic excursion, and rapid strain metrics. **(4)** Age- and sex-specific continuous centile curves were generated using LMS modeling and translated into clinically interpretable categories of within reference range, mildly out of range, and at least moderately out of range. **(5)** Biological and technical determinants were investigated, including age, sex, ethnicity, scanner vendor, magnetic field strength, and post-processing software. **(6)** The resulting centile categories were applied to approximately 45,000 UK Biobank participants and assessed in relation to incident heart failure, myocardial infarction, stroke, and all-cause mortality. Lower centile categories were associated with higher cardiovascular risk, with the strongest associations observed for incident heart failure. **Abbreviations:** CMR = cardiovascular magnetic resonance; GCS = global circumferential strain; GLS = global longitudinal strain; GRS = global radial strain; LA = left atrial; LAS = long-axis strain; LMS = lambda-mu-sigma; LV = left ventricular; MAPSE = mitral annular plane systolic excursion; QC = quality control; RA = right atrial; RV = right ventricular; TAPSE = tricuspid annular plane systolic excursion.

