## Supplementary_material_reference_values_strain_HHC for "Age-, sex-, and ethnicity-stratified CMR strain reference ranges and centile-based categories from the Healthy Hearts Consortium"

**Supplementary Figure 1 Flowchart of the quality control process for strain values derived using cvi42 (Circle Cardiovascular Imaging Inc.)**

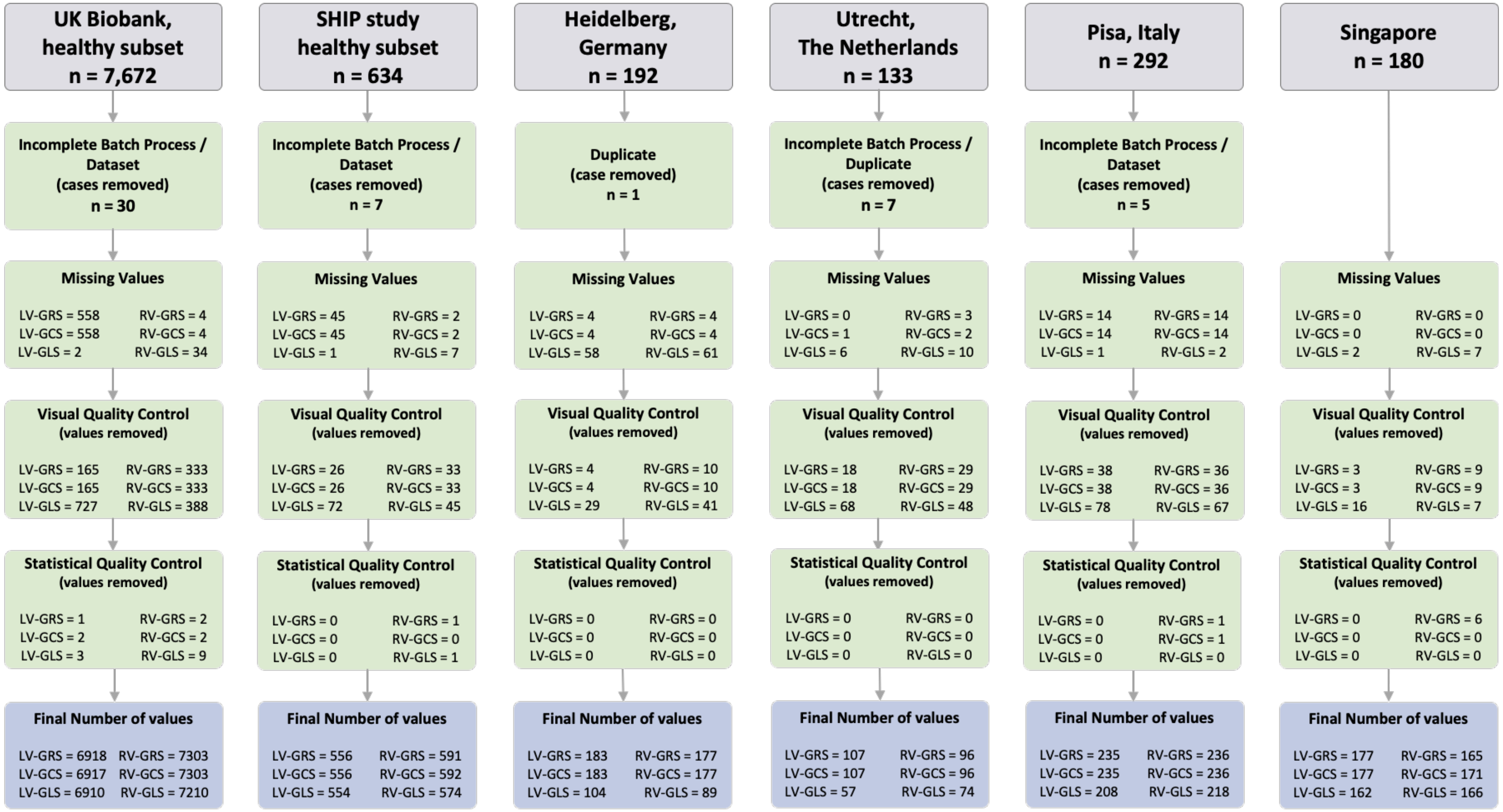

**Footnote**: Incomplete batch processing, dataset exclusions, and duplicate case removal were performed prior to analysis. Missing values indicate non-available strain measurements. Visual quality control refers to manual exclusion due to insufficient image quality or tracking errors. Statistical quality control corresponds to exclusion of statistical outliers according to pre-specified criteria. Final numbers represent valid measurements per parameter after sequential processing steps.

**Abbreviations:** LV = left ventricle; RV = right ventricle; GCS = global circumferential strain; GLS = global longitudinal strain; GRS = global radial strain.

**Supplementary Figure 2 Flowchart for the quality control for the strain values derived by suiteHEART (NeoSoft)**

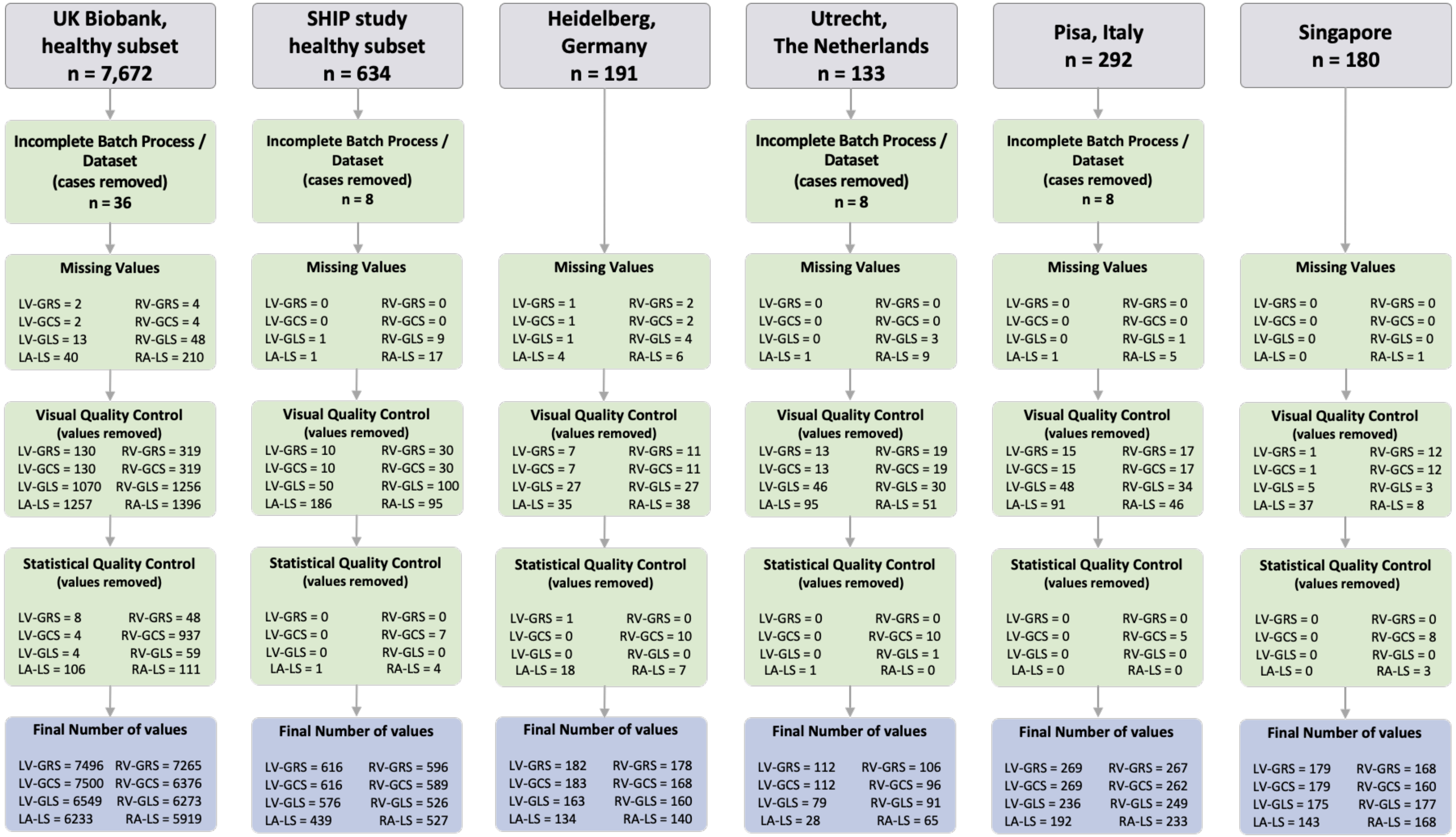

**Footnote:** Incomplete batch processes, dataset exclusions, and duplicate cases were applied prior to analysis. Missing values represent non-available strain and long-axis strain measurements. Visual quality control refers to manual exclusion due to insufficient image quality or tracking errors. Statistical quality control corresponds to exclusion of statistical outliers according to pre-specified criteria. Final numbers represent valid measurements per parameter after sequential processing steps.

**Abbreviations:** LA = left atrium; LS = longitudinal strain; LV = left ventricle; RA = right atrium; RV = right ventricle; GCS = global circumferential strain; GLS = global longitudinal strain; GRS = global radial strain.

**Supplementary Figure 3 Flowchart for the quality control for the annular plane systolic excursion and rapid longitudinal shortening values derived by cvi42 (Circle Cardiovascular Imaging Inc)**

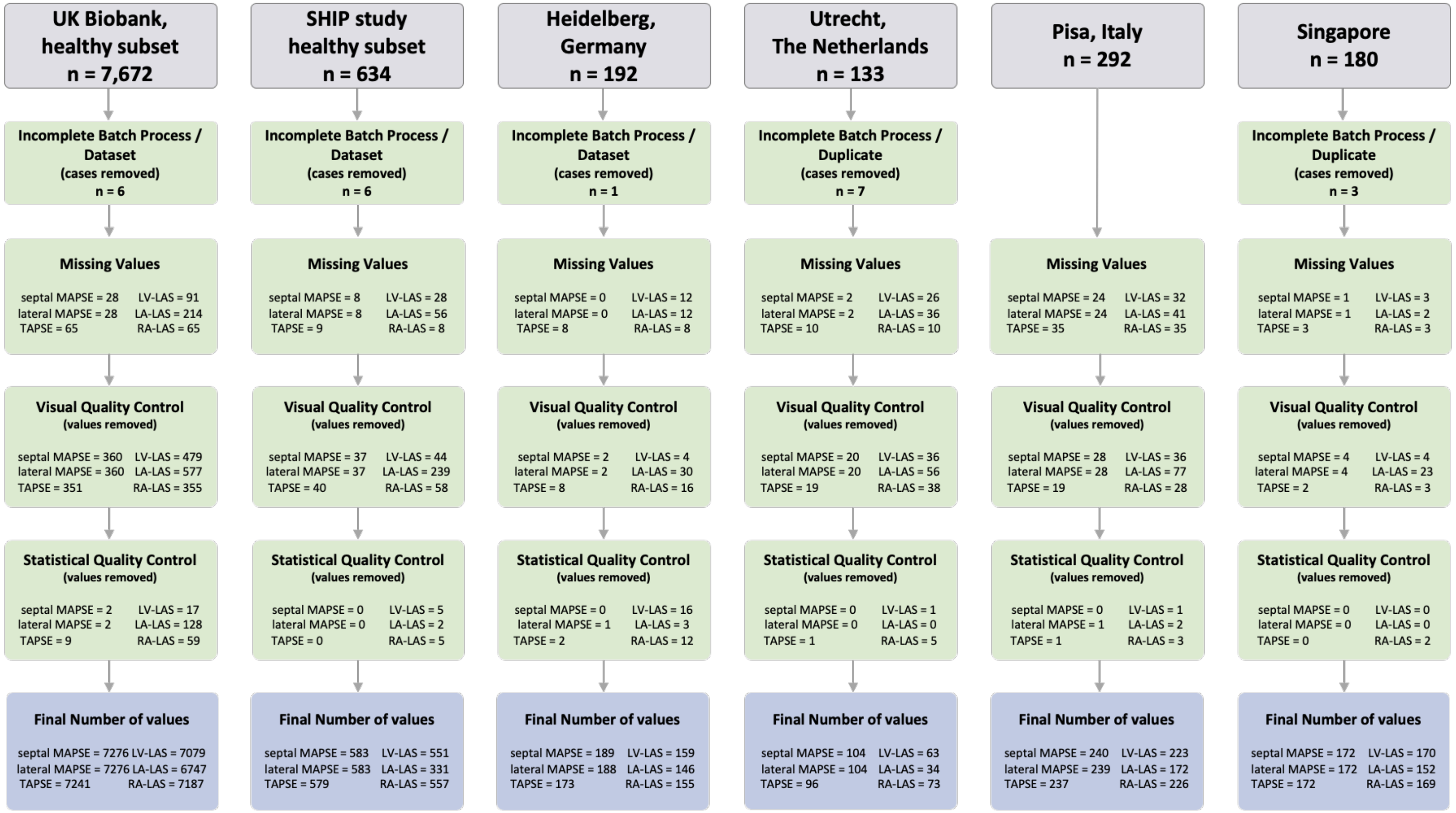

**Footnote:** Incomplete batch processing, dataset exclusions, and duplicate case removal were performed prior to analysis. Missing values indicate non-available annular plane excursion and rapid longitudinal shortening measurements. Visual quality control refers to manual exclusion due to insufficient image quality or tracking errors. Statistical quality control corresponds to exclusion of statistical outliers according to pre-specified criteria. Final numbers represent valid measurements per parameter after sequential processing steps.
**Abbreviations:** AVJS = atrioventricular junction shortening; LA = left atrium; LS = long-axis shortening; LV = left ventricle; RA = right atrium; MAPSE = mitral annular plane systolic excursion; TAPSE = tricuspid annular plane systolic excursion.

### Supplementary Figure 4 Age-specific reference ranges and centile-based categories for atrial and ventricular strain metrics in females (*all ethnicities*), derived by suiteHEART (NeoSoft)

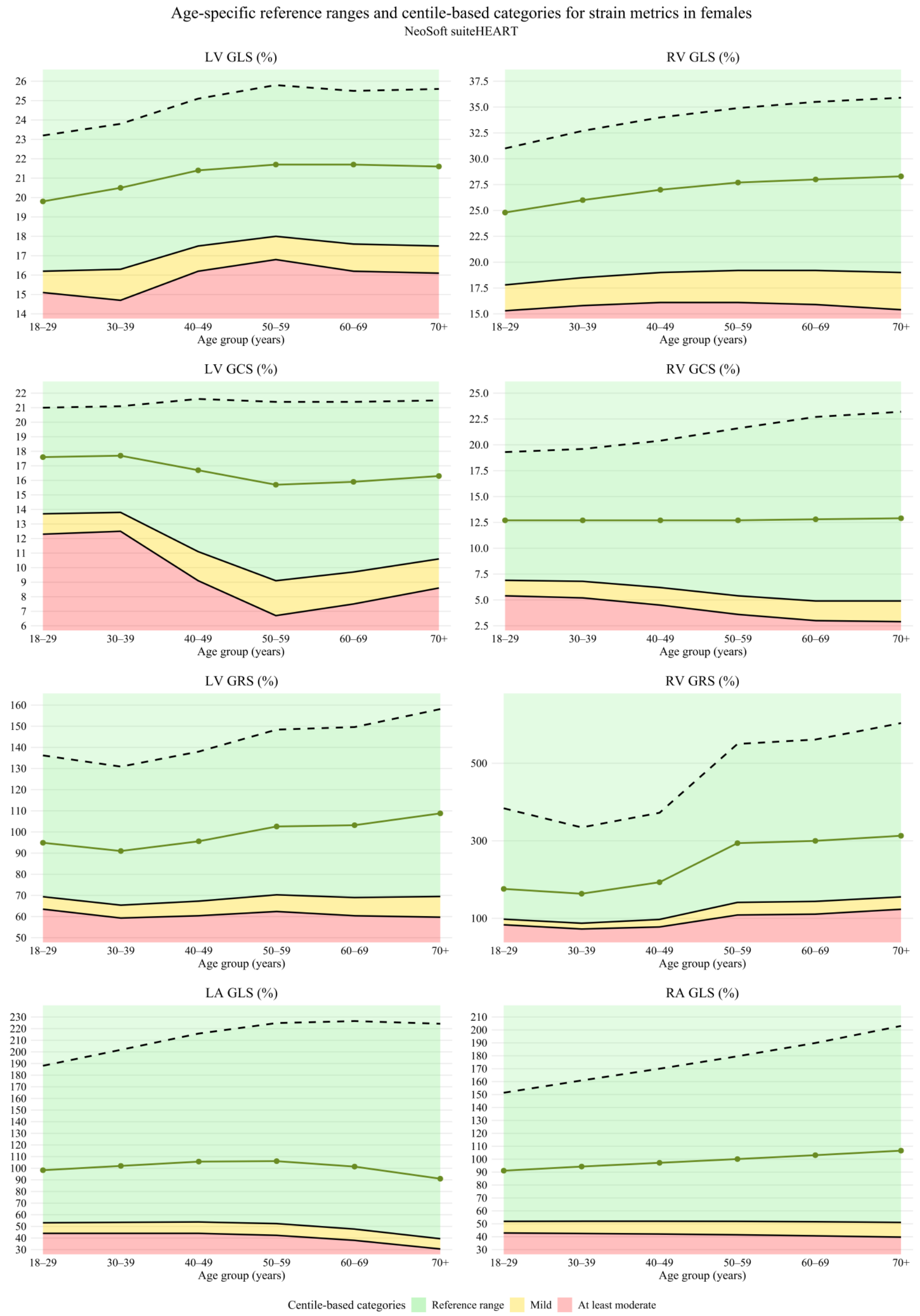

**Footnote:** Age-specific reference ranges and centile-based categories for left ventricular (LV), right ventricular (RV), left atrial (LA), and right atrial (RA) strain metrics derived from CMR feature tracking (suiteHEART by NeoSoft) in females. Solid lines represent median values across age groups, dashed lines indicate upper reference limits, and shaded areas depict centile-based categories: green (reference range, ≥P2.5), yellow (mild reduction, <P2.5), and red (at least moderate reduction, <P0.5). Age-related trends are shown for LV and RV global longitudinal (GLS), circumferential (GCS), and radial strain (GRS), as well as atrial longitudinal strain (LA GLS, RA GLS).

**Abbreviations:** LA = left atrium; LV = left ventricle; RA = right atrium; RV = right ventricle; GCS = global circumferential strain; GLS = global longitudinal strain; GRS = global radial strain.

### Supplementary Figure 5 Age-specific reference ranges and centile-based categories for atrial and ventricular strain metrics in males (*all ethnicities*), derived by suiteHEART (NeoSoft)

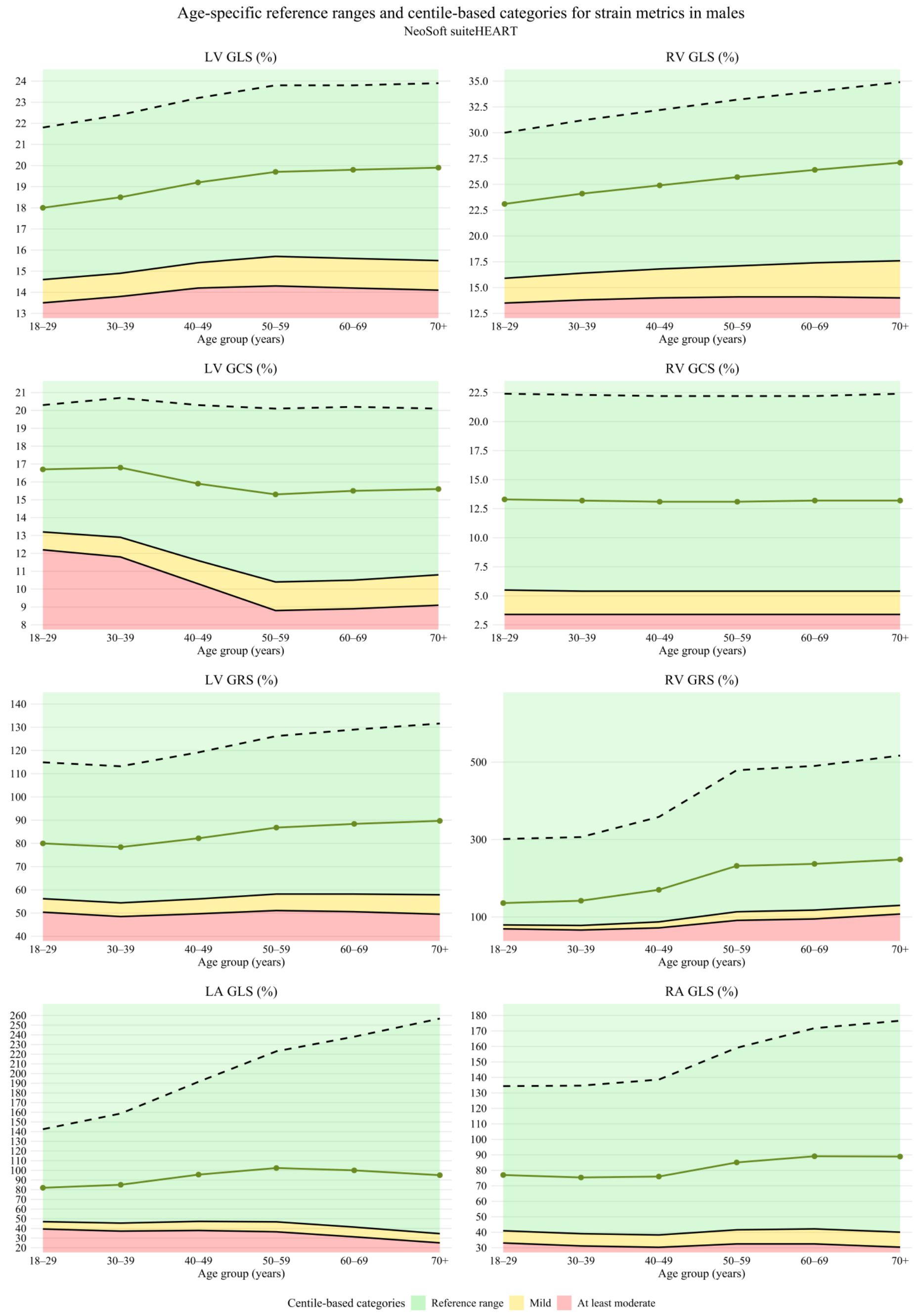

**Footnote:** Age-specific reference ranges and centile-based categories for left ventricular (LV), right ventricular (RV), left atrial (LA), and right atrial (RA) strain metrics derived from CMR feature tracking (suiteHEART by NeoSoft) in males. Solid lines represent median values across age groups, dashed lines indicate upper reference limits, and shaded areas depict centile-based categories: green (reference range, ≥P2.5), yellow (mild reduction, <P2.5), and red (at least moderate reduction, <P0.5). Age-related trends are shown for LV and RV global longitudinal (GLS), circumferential (GCS), and radial strain (GRS), as well as atrial longitudinal strain (LA GLS, RA GLS).

**Abbreviations:** LA = left atrium; LV = left ventricle; RA = right atrium; RV = right ventricle; GCS = global circumferential strain; GLS = global longitudinal strain; GRS = global radial strain.

**Supplementary Figure 6 Age-specific reference ranges and centile-based categories for MAPSE and TAPSE metrics in females (*all ethnicities*), using cvi42 (Circle Cardiovascular Imaging Inc.)**

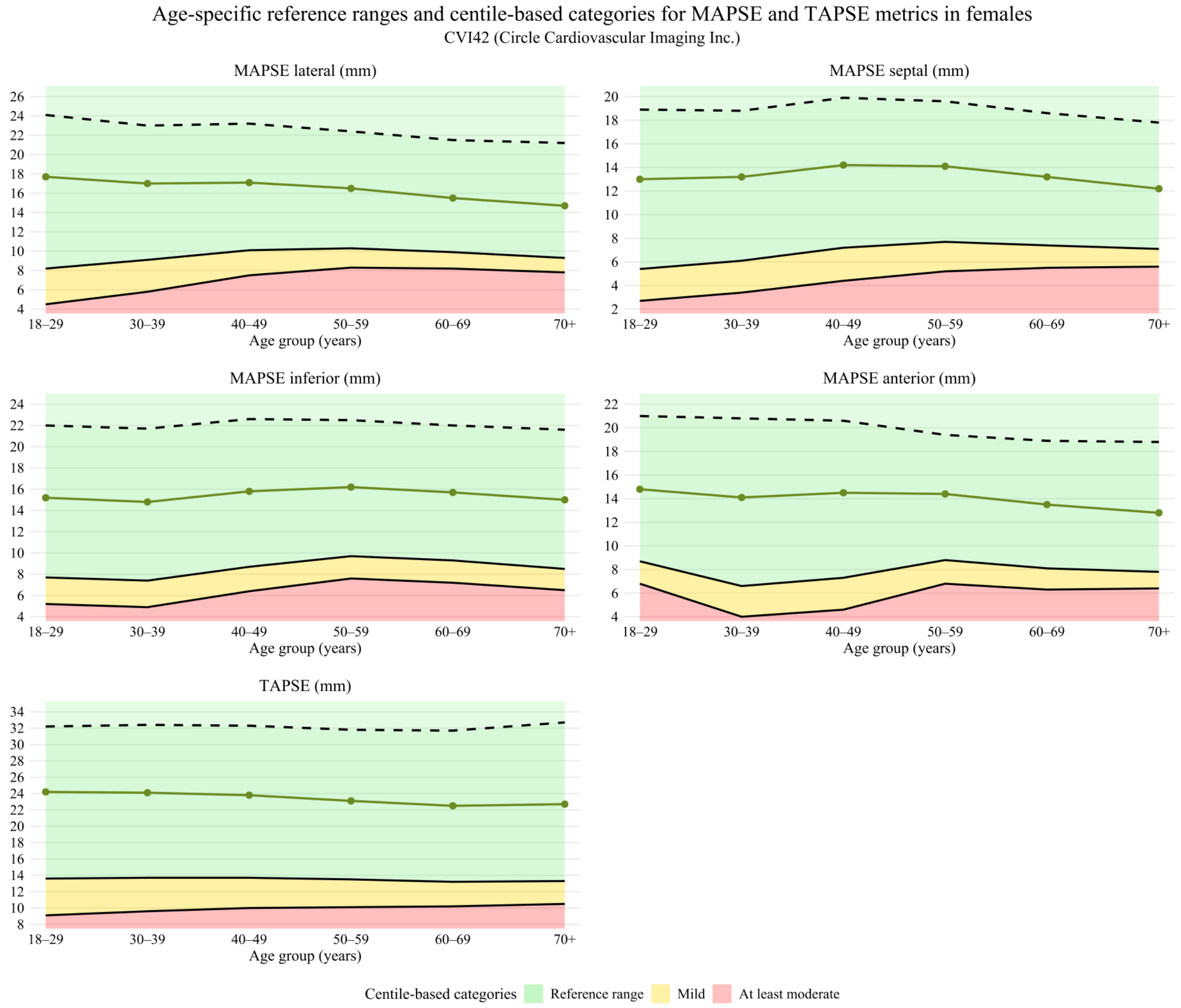

**Footnote:** Age-specific reference ranges and centile-based categories for mitral annular plane systolic excursion (MAPSE; lateral, septal, inferior, anterior) and tricuspid annular plane systolic excursion (TAPSE) derived by cvi42 (Circle Cardiovascular Imaging Inc.) in females. Solid lines represent median values across age groups, dashed lines indicate upper reference limits, and shaded areas depict centile-based categories: green (reference range, ≥P2.5), yellow (mild reduction, <P2.5), and red (at least moderate reduction, <P0.5). Age-related trends across annular displacement metrics are shown. **Abbreviations:** MAPSE = mitral annular plane systolic excursion; TAPSE = tricuspid annular plane systolic excursion.

**Supplementary Figure 7 Age-specific reference ranges and centile-based categories for MAPSE and TAPSE metrics in males (*all ethnicities*), using cvi42 (Circle Cardiovascular Imaging Inc.)**

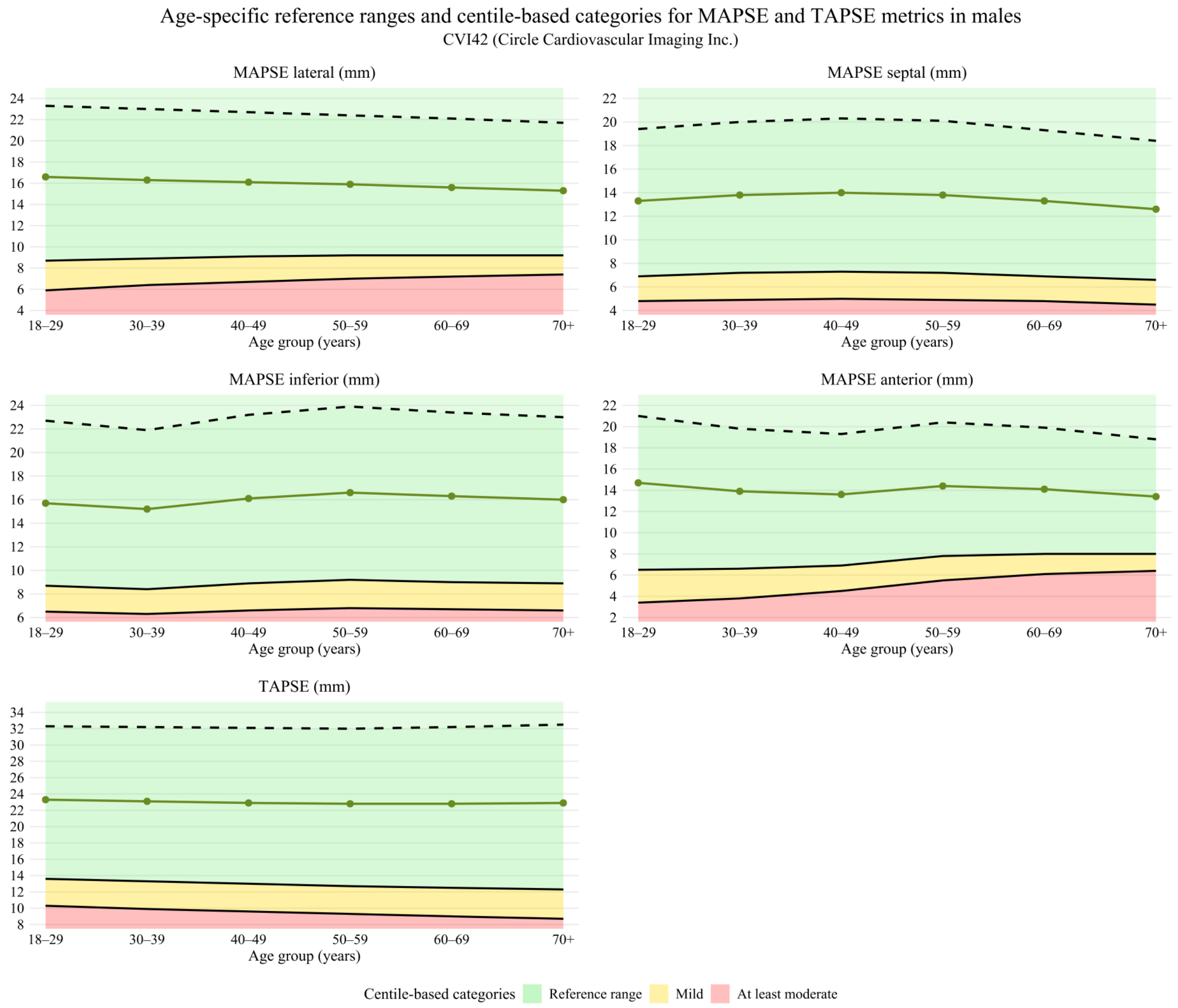

**Footnote:** Age-specific reference ranges and centile-based categories for mitral annular plane systolic excursion (MAPSE; lateral, septal, inferior, anterior) and tricuspid annular plane systolic excursion (TAPSE) derived by cvi42 (Circle Cardiovascular Imaging Inc.) in males. Solid lines represent median values across age groups, dashed lines indicate upper reference limits, and shaded areas depict centile-based categories: green (reference range, ≥P2.5), yellow (mild reduction, <P2.5), and red (at least moderate reduction, <P0.5). Age-related trends across annular displacement metrics are shown. **Abbreviations:** MAPSE = mitral annular plane systolic excursion; TAPSE = tricuspid annular plane systolic excursion.

**Supplementary Figure 8 Age-specific reference ranges and centile-based categories for LV rapid strain metrics in females (*all ethnicities*), using cvi42 (Circle Cardiovascular Imaging Inc.)**

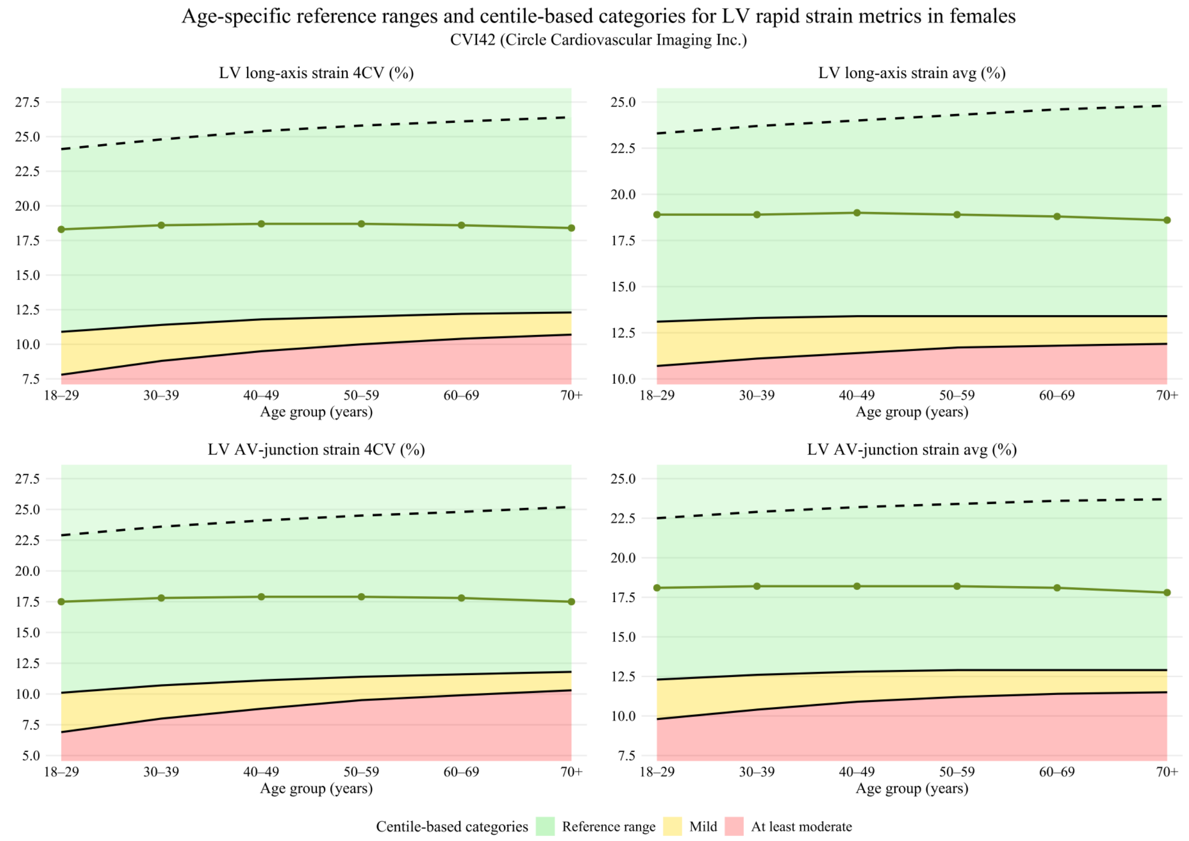

**Footnote:** Age-specific reference ranges and centile-based categories for left ventricular (LV) rapid strain metrics derived by cvi42 (Circle Cardiovascular Imaging Inc.) in females, including LV long-axis strain (4CV and average) and LV atrioventricular (AV) junction strain (4CV and average). Solid lines represent median values across age groups, dashed lines indicate upper reference limits, and shaded areas depict centile-based categories: green (reference range, ≥P2.5), yellow (mild reduction, <P2.5), and red (at least moderate reduction, <P0.5). Age-related trends are illustrated across all LV rapid strain parameters. **Abbreviations:** AV = atrioventricular; 4CV = four-chamber view; LV = left ventricle.

**Supplementary Figure 9 Age-specific reference ranges and centile-based categories for LV rapid strain metrics in males (*all ethnicities*), using cvi42 (Circle Cardiovascular Imaging Inc.)**

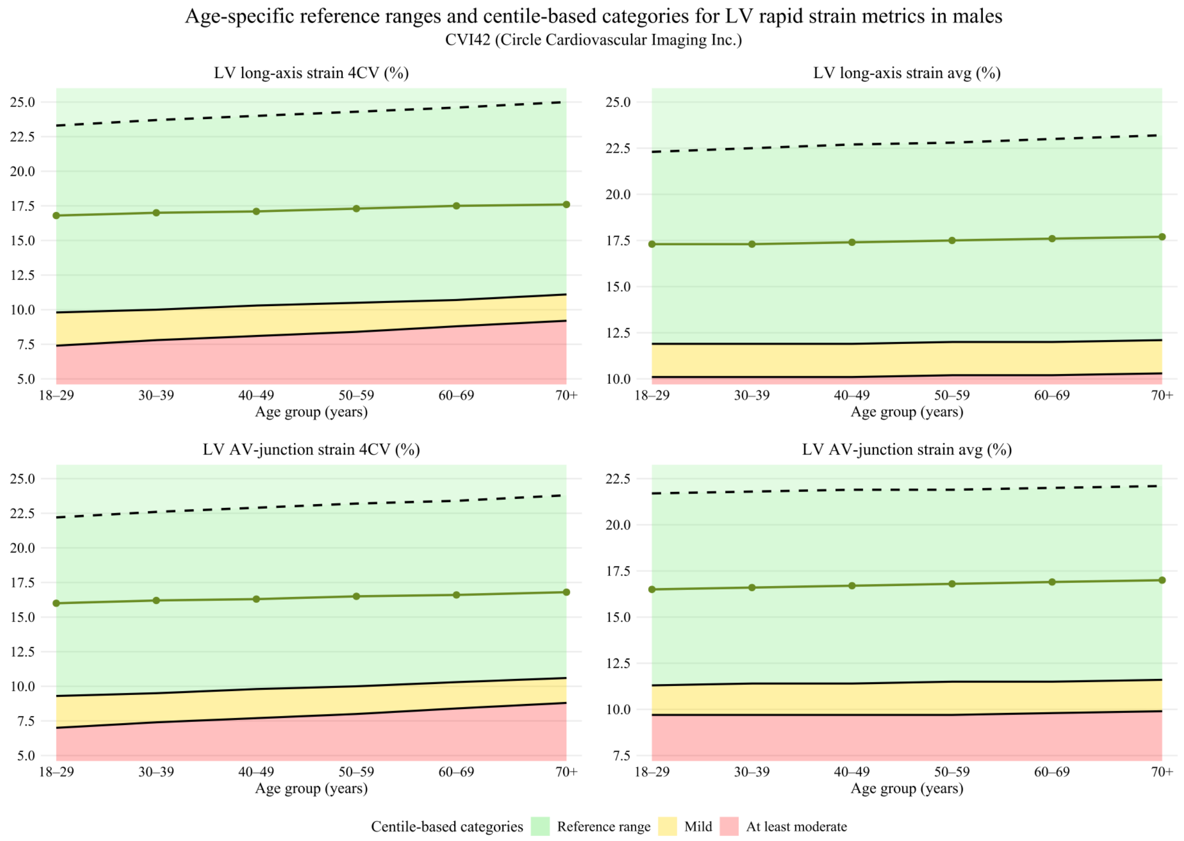

**Footnote:** Age-specific reference ranges and centile-based categories for left ventricular (LV) rapid strain metrics derived by cvi42 (Circle Cardiovascular Imaging Inc.) in males, including LV long-axis strain (4CV and average) and LV atrioventricular (AV) junction strain (4CV and average). Solid lines represent median values across age groups, dashed lines indicate upper reference limits, and shaded areas depict centile-based categories: green (reference range, ≥P2.5), yellow (mild reduction, <P2.5), and red (at least moderate reduction, <P0.5). Age-related trends are illustrated across all LV rapid strain parameters.
**Abbreviations:** AV = atrioventricular; 4CV = four-chamber view; LV = left ventricle.

**Supplementary Figure 10 Age-specific reference ranges and centile-based categories for atrial rapid strain metrics in females (*all ethnicities*), using cvi42 (Circle Cardiovascular Imaging Inc.)**

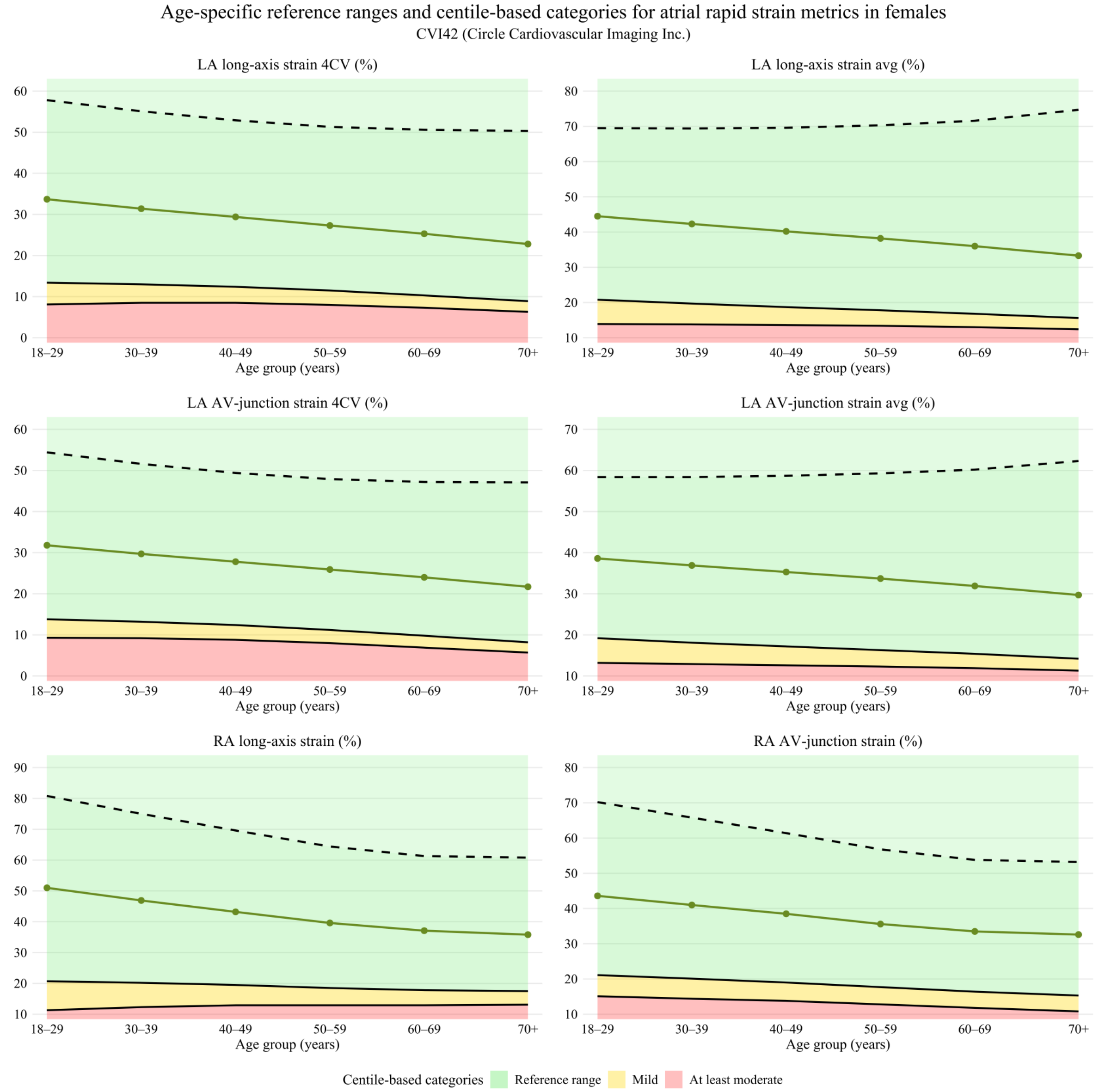

**Footnote:**
Age-specific reference ranges and centile-based categories for left atrial (LA) and right atrial (RA) rapid strain metrics derived by cvi42 (Circle Cardiovascular Imaging Inc.) in females, including LA and RA long-axis strain and atrioventricular (AV) junction strain (4CV and average where applicable). Solid lines represent median values across age groups, dashed lines indicate upper reference limits, and shaded areas depict centile-based categories: green (reference range, ≥P2.5), yellow (mild reduction, <P2.5), and red (at least moderate reduction, <P0.5). Age-related trends are shown for atrial deformation parameters.
**Abbreviations:** 4CV = four-chamber view; AV = atrioventricular; LA = left atrium; RA = right atrium.

**Supplementary Figure 11 Age-specific reference ranges and centile-based categories for atrial rapid strain metrics in males (*all ethnicities*), using cvi42 (Circle Cardiovascular Imaging Inc.)**

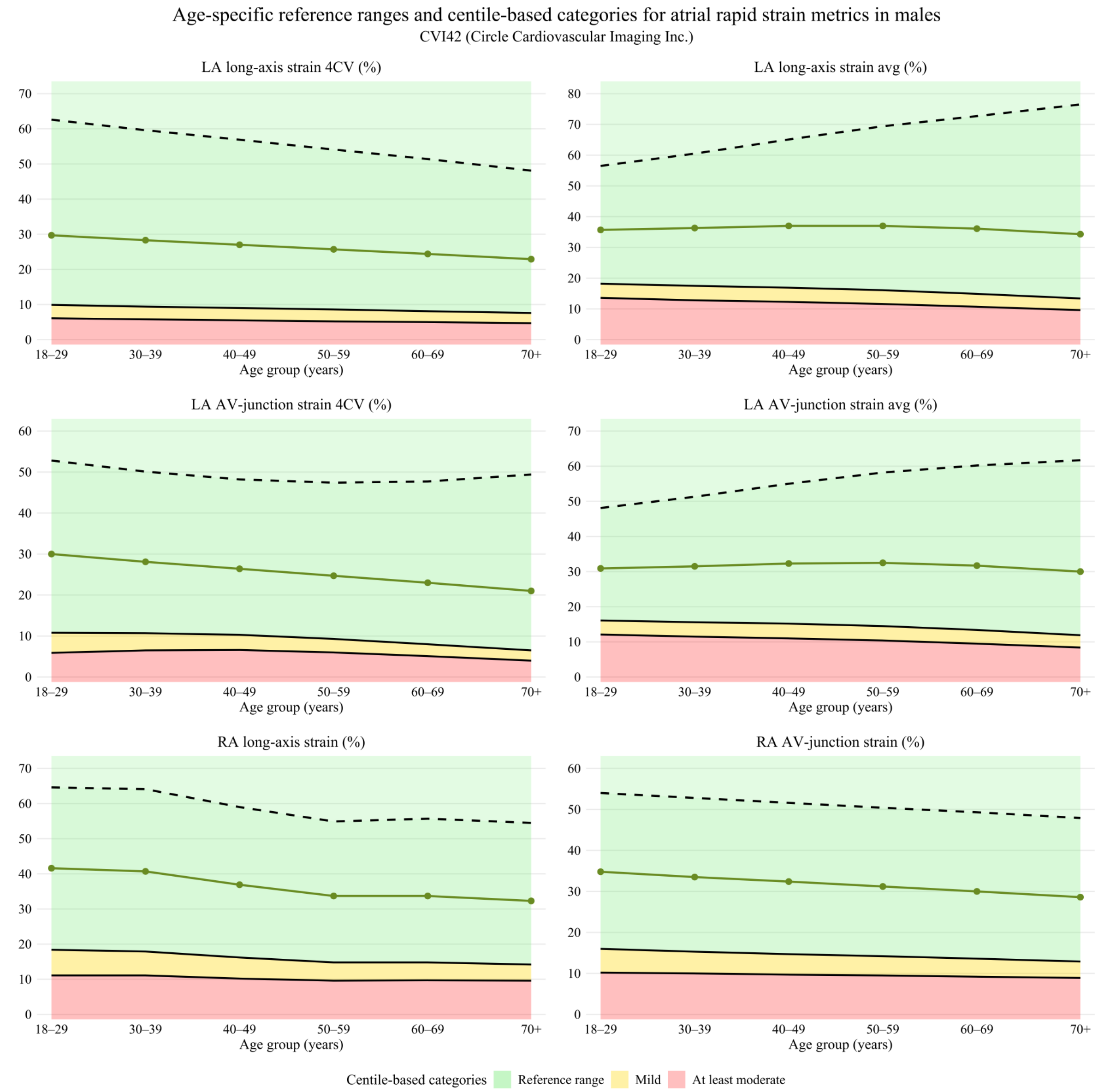

**Footnote:**

Age-specific reference ranges and centile-based categories for left atrial (LA) and right atrial (RA) rapid strain metrics derived by cvi42 (Circle Cardiovascular Imaging Inc.) in males, including LA and RA long-axis strain and atrioventricular (AV) junction strain (4CV and average where applicable). Solid lines represent median values across age groups, dashed lines indicate upper reference limits, and shaded areas depict centile-based categories: green (reference range, ≥P2.5), yellow (mild reduction, <P2.5), and red (at least moderate reduction, <P0.5). Age-related trends are shown for atrial deformation parameters.
**Abbreviations:** 4CV = four-chamber view; AV = atrioventricular; LA = left atrium; RA = right atrium.

**Supplementary Figure 12 Ethnicity-specific differences in global ventricular strain metrics across age and sex groups using cvi42 (Circle Cardiovascular Imaging Inc.)**

**Asian vs. White ethnicity**

| Metric | 18-29 | | 30-39 | | 40-49 | | 50-59 | | 60-69 | | >70 | |
|---|---|---|---|---|---|---|---|---|---|---|---|---|
| | Women | Men | Women | Men | Women | Men | Women | Men | Women | Men | Women | Men |
| LV GLS (%) | n<5 | n<5 | n<5 | n<5 | = | = | = | = | = | = | = | = |
| LV GRS (%) | n<5 | n<5 | n<5 | n<5 | = | = | = | = | = | = | = | = |
| LV GCS (%) | n<5 | n<5 | n<5 | n<5 | = | = | = | = | = | = | = | = |
| RV GLS (%) | n<5 | n<5 | n<5 | n<5 | = | = | = | = | = | = | = | = |
| RV GRS (%) | n<5 | n<5 | n<5 | n<5 | = | = | = | ↓ | = | = | = | = |
| RV GCS (%) | n<5 | n<5 | n<5 | n<5 | = | = | = | ↓ | = | = | = | = |

**Black vs. White ethnicity**

| Metric | 18-29 | | 30-39 | | 40-49 | | 50-59 | | 60-69 | | >70 | |
|---|---|---|---|---|---|---|---|---|---|---|---|---|
| | Women | Men | Women | Men | Women | Men | Women | Men | Women | Men | Women | Men |
| LV GLS (%) | n<5 | n<5 | n<5 | n<5 | = | = | ↓ | ↓ | ↓ | ↓ | = | = |
| LV GRS (%) | n<5 | n<5 | n<5 | n<5 | = | = | = | = | = | = | = | = |
| LV GCS (%) | n<5 | n<5 | n<5 | n<5 | = | = | = | = | = | = | = | = |
| RV GLS (%) | n<5 | n<5 | n<5 | n<5 | = | = | ↓ | ↓ | ↓ | ↓ | = | = |
| RV GRS (%) | n<5 | n<5 | n<5 | n<5 | = | = | ↓ | ↓ | ↓ | ↓ | = | = |
| RV GCS (%) | n<5 | n<5 | n<5 | n<5 | = | = | ↓ | ↓ | ↓ | ↓ | = | = |

**Chinese vs. White ethnicity**

| Metric | 18-29 | | 30-39 | | 40-49 | | 50-59 | | 60-69 | | >70 | |
|---|---|---|---|---|---|---|---|---|---|---|---|---|
| | Women | Men | Women | Men | Women | Men | Women | Men | Women | Men | Women | Men |
| LV GLS (%) | = | = | = | = | = | = | = | = | = | = | = | n<5 |
| LV GRS (%) | ↑ | ↑ | ↑ | = | ↑ | = | ↑ | = | = | = | = | n<5 |
| LV GCS (%) | = | = | ↑ | = | ↑ | = | ↑ | = | = | = | = | n<5 |
| RV GLS (%) | = | = | = | = | = | = | = | = | = | = | = | n<5 |
| RV GRS (%) | = | = | ↑ | = | = | = | ↑ | = | = | = | = | n<5 |
| RV GCS (%) | = | = | = | = | = | = | = | = | = | = | = | n<5 |

**Mixed/Other vs. White ethnicity**

| Metric | 18-29 | | 30-39 | | 40-49 | | 50-59 | | 60-69 | | >70 | |
|---|---|---|---|---|---|---|---|---|---|---|---|---|
| | Women | Men | Women | Men | Women | Men | Women | Men | Women | Men | Women | Men |
| LV GLS (%) | n<5 | n<5 | n<5 | n<5 | = | = | = | = | = | = | = | = |
| LV GRS (%) | n<5 | n<5 | n<5 | n<5 | = | = | = | = | = | = | = | = |
| LV GCS (%) | n<5 | n<5 | n<5 | n<5 | = | = | = | = | = | = | = | = |
| RV GLS (%) | n<5 | n<5 | n<5 | n<5 | = | = | = | = | = | = | = | = |
| RV GRS (%) | n<5 | n<5 | n<5 | n<5 | = | = | = | = | = | ↓ | = | = |
| RV GCS (%) | n<5 | n<5 | n<5 | n<5 | = | = | = | = | = | = | = | = |

**Footnote:** Comparison of left ventricular (LV) and right ventricular (RV) global longitudinal (GLS), circumferential (GCS), and radial strain (GRS) between different ethnic groups (South Asian, Black, Chinese, and Mixed/Other) and the White reference population, stratified by age group and sex, derived from CMR feature tracking using CVI42 (Circle Cardiovascular Imaging Inc.). Arrows indicate direction of difference relative to the White group (↑ higher, ↓ lower), while "=" denotes no relevant difference. Cells marked "n<5" indicate insufficient sample size for reliable comparison. Group comparisons were performed using Wilcoxon rank-sum tests with adjustment for multiple testing.

**Abbreviations:** LV = left ventricle; RV = right ventricle; GCS = global circumferential strain; GLS = global longitudinal strain; GRS = global radial strain.

**Supplementary Figure 13 Ethnicity-specific differences in global ventricular and atrial strain metrics across age and sex groups using suiteHEART (NeoSoft)**

**Asian vs. White ethnicity**

| Metric | 18-29 | | 30-39 | | 40-49 | | 50-59 | | 60-69 | | >70 | |
|---|---|---|---|---|---|---|---|---|---|---|---|---|
| | Women | Men | Women | Men | Women | Men | Women | Men | Women | Men | Women | Men |
| LV GLS (%) | n<5 | n<5 | n<5 | n<5 | = | ↑ | = | ↑ | ↑ | ↑ | = | = |
| LV GRS (%) | n<5 | n<5 | n<5 | n<5 | = | ↑ | = | ↑ | = | = | = | = |
| LV GCS (%) | n<5 | n<5 | n<5 | n<5 | = | = | = | = | = | = | = | = |
| RV GLS (%) | n<5 | n<5 | n<5 | n<5 | = | = | = | = | = | = | = | = |
| RV GRS (%) | n<5 | n<5 | n<5 | n<5 | = | = | = | ↑ | = | ↑ | = | = |
| RV GCS (%) | n<5 | n<5 | n<5 | n<5 | = | = | = | ↓ | = | = | = | = |
| LA GLS (%) | n<5 | n<5 | n<5 | n<5 | = | = | = | = | = | = | = | = |
| RA GLS (%) | n<5 | n<5 | n<5 | n<5 | = | = | = | = | = | = | = | = |

**Black vs. White ethnicity**

| Metric | 18-29 | | 30-39 | | 40-49 | | 50-59 | | 60-69 | | >70 | |
|---|---|---|---|---|---|---|---|---|---|---|---|---|
| | Women | Men | Women | Men | Women | Men | Women | Men | Women | Men | Women | Men |
| LV GLS (%) | n<5 | n<5 | n<5 | n<5 | = | = | = | = | = | = | = | = |
| LV GRS (%) | n<5 | n<5 | n<5 | n<5 | = | = | = | = | = | = | = | = |
| LV GCS (%) | n<5 | n<5 | n<5 | n<5 | = | = | = | = | = | = | = | = |
| RV GLS (%) | n<5 | n<5 | n<5 | n<5 | = | = | = | = | = | = | = | = |
| RV GRS (%) | n<5 | n<5 | n<5 | n<5 | = | = | ↑ | = | = | = | = | = |
| RV GCS (%) | n<5 | n<5 | n<5 | n<5 | = | = | = | = | = | = | = | = |
| LA GLS (%) | n<5 | n<5 | n<5 | n<5 | = | = | = | = | = | = | = | = |
| RA GLS (%) | n<5 | n<5 | n<5 | n<5 | = | = | = | = | = | = | ↓ | = |

**Chinese vs. White ethnicity**

| Metric | 18-29 | | 30-39 | | 40-49 | | 50-59 | | 60-69 | | >70 | |
|---|---|---|---|---|---|---|---|---|---|---|---|---|
| | Women | Men | Women | Men | Women | Men | Women | Men | Women | Men | Women | Men |
| LV GLS (%) | = | = | = | = | = | = | = | = | = | = | = | = |
| LV GRS (%) | = | = | = | = | = | = | = | = | = | = | = | n<5 |
| LV GCS (%) | = | = | = | = | ↑ | ↑ | = | = | = | = | = | n<5 |
| RV GLS (%) | = | = | = | = | = | = | = | = | = | = | = | = |
| RV GRS (%) | = | = | = | = | = | ↓ | = | = | = | = | = | n<5 |
| RV GCS (%) | = | = | = | = | = | = | = | = | = | = | = | n<5 |
| LA GLS (%) | = | = | = | = | = | = | = | = | = | = | = | n<5 |
| RA GLS (%) | = | = | = | = | = | = | = | = | = | = | = | n<5 |

**Mixed/Other vs. White ethnicity**

| Metric | 18-29 | | 30-39 | | 40-49 | | 50-59 | | 60-69 | | >70 | |
|---|---|---|---|---|---|---|---|---|---|---|---|---|
| | Women | Men | Women | Men | Women | Men | Women | Men | Women | Men | Women | Men |
| LV GLS (%) | n<5 | n<5 | n<5 | n<5 | = | = | = | = | = | = | = | = |
| LV GRS (%) | n<5 | n<5 | n<5 | n<5 | = | = | = | = | = | = | = | = |
| LV GCS (%) | n<5 | n<5 | n<5 | n<5 | = | = | = | = | = | = | = | = |
| RV GLS (%) | n<5 | n<5 | n<5 | n<5 | = | = | = | = | = | = | = | = |
| RV GRS (%) | n<5 | n<5 | n<5 | n<5 | = | = | = | = | = | = | = | = |
| RV GCS (%) | n<5 | n<5 | n<5 | n<5 | = | = | = | = | = | = | = | = |
| LA GLS (%) | n<5 | n<5 | n<5 | n<5 | = | = | ↑ | = | = | = | = | = |
| RA GLS (%) | n<5 | n<5 | n<5 | n<5 | = | = | = | = | = | = | = | = |

**Footnote:** Comparison of left ventricular (LV), right ventricular (RV), left atrial (LA), and right atrial (RA) strain metrics, including global longitudinal (GLS), circumferential (GCS), and radial strain (GRS) where applicable, between different ethnic groups (South Asian, Black, Chinese, and Mixed/Other) and the White reference population, stratified by age group and sex, derived from CMR feature tracking using suiteHEART (NeoSoft). Arrows indicate direction of difference relative to the White group (↑ higher, ↓ lower), while "=" denotes no relevant difference. Cells marked "n<5" indicate insufficient sample size for reliable comparison. Group comparisons were performed using Wilcoxon rank-sum tests with adjustment for multiple testing.
**Abbreviations:** LA = left atrium; LV = left ventricle; RA = right atrium; RV = right ventricle; GCS = global circumferential strain; GLS = global longitudinal strain; GRS = global radial strain.

**Supplementary Figure 14 Ethnicity-specific differences in MAPSE and TAPSE across age and sex groups using cvi42 (Circle Cardiovascular Imaging Inc.)**

**Asian vs. White ethnicity**

| Metric | 18-29 | | 30-39 | | 40-49 | | 50-59 | | 60-69 | | >70 | |
|---|---|---|---|---|---|---|---|---|---|---|---|---|
| | Women | Men | Women | Men | Women | Men | Women | Men | Women | Men | Women | Men |
| MAPSE lateral (mm) | n<5 | n<5 | n<5 | n<5 | = | = | = | = | = | = | = | = |
| MAPSE septal (mm) | n<5 | n<5 | n<5 | n<5 | = | = | = | = | = | = | = | = |
| MAPSE inferior (mm) | n<5 | n<5 | n<5 | n<5 | = | = | = | = | = | = | = | = |
| MAPSE anterior (mm) | n<5 | n<5 | n<5 | n<5 | = | = | = | = | = | = | = | = |
| TAPSE (mm) | n<5 | n<5 | n<5 | n<5 | = | = | = | = | = | = | = | = |

**Black vs. White ethnicity**

| Metric | 18-29 | | 30-39 | | 40-49 | | 50-59 | | 60-69 | | >70 | |
|---|---|---|---|---|---|---|---|---|---|---|---|---|
| | Women | Men | Women | Men | Women | Men | Women | Men | Women | Men | Women | Men |
| MAPSE lateral (mm) | n<5 | n<5 | n<5 | n<5 | = | = | = | = | = | = | = | = |
| MAPSE septal (mm) | n<5 | n<5 | n<5 | n<5 | = | = | = | = | = | = | = | = |
| MAPSE inferior (mm) | n<5 | n<5 | n<5 | n<5 | = | = | = | = | = | = | = | = |
| MAPSE anterior (mm) | n<5 | n<5 | n<5 | n<5 | = | = | = | = | = | = | = | = |
| TAPSE (mm) | n<5 | n<5 | n<5 | n<5 | = | = | = | = | = | = | = | = |

**Chinese vs. White ethnicity**

| Metric | 18-29 | | 30-39 | | 40-49 | | 50-59 | | 60-69 | | >70 | |
|---|---|---|---|---|---|---|---|---|---|---|---|---|
| | Women | Men | Women | Men | Women | Men | Women | Men | Women | Men | Women | Men |
| MAPSE lateral (mm) | = | = | = | = | = | = | = | = | = | = | = | n<5 |
| MAPSE septal (mm) | = | = | = | = | = | = | = | = | = | = | = | n<5 |
| MAPSE inferior (mm) | = | = | = | = | = | = | = | = | = | = | = | n<5 |
| MAPSE anterior (mm) | = | = | = | = | = | = | = | = | = | = | = | n<5 |
| TAPSE (mm) | = | = | = | = | = | = | = | = | = | = | = | n<5 |

**Mixed/Other vs. White ethnicity**

| Metric | 18-29 | | 30-39 | | 40-49 | | 50-59 | | 60-69 | | >70 | |
|---|---|---|---|---|---|---|---|---|---|---|---|---|
| | Women | Men | Women | Men | Women | Men | Women | Men | Women | Men | Women | Men |
| MAPSE lateral (mm) | n<5 | n<5 | n<5 | n<5 | = | = | = | = | = | = | = | = |
| MAPSE septal (mm) | n<5 | n<5 | n<5 | n<5 | = | = | = | = | = | = | = | = |
| MAPSE inferior (mm) | n<5 | n<5 | n<5 | n<5 | = | = | ↑ | = | = | = | = | = |
| MAPSE anterior (mm) | n<5 | n<5 | n<5 | n<5 | = | = | = | ↑ | = | = | = | = |
| TAPSE (mm) | n<5 | n<5 | n<5 | n<5 | = | = | = | = | = | = | = | = |

**Footnote:** Comparison of mitral annular plane systolic excursion (MAPSE; lateral, septal, inferior, anterior) and tricuspid annular plane systolic excursion (TAPSE) between different ethnic groups (South Asian, Black, Chinese, and Mixed/Other) and the White reference population, stratified by age group and sex, using cvi42 (Circle Cardiovascular Imaging Inc.). Arrows indicate direction of difference relative to the White group (↑ higher, ↓ lower), while "=" denotes no relevant difference. Cells marked "n<5" indicate insufficient sample size for reliable comparison. Group comparisons were performed using Wilcoxon rank-sum tests with adjustment for multiple testing.

**Abbreviations:** MAPSE = mitral annular plane systolic excursion; TAPSE = tricuspid annular plane systolic excursion.

**Supplementary Figure 15 Ethnicity-specific differences in left ventricular long-axis and atrioventricular junction strain across age and sex groups using cvi42 (Circle Cardiovascular Imaging Inc.)**

**Asian vs. White ethnicity**

| Metric | 18-29 | | 30-39 | | 40-49 | | 50-59 | | 60-69 | | >70 | |
|---|---|---|---|---|---|---|---|---|---|---|---|---|
| | Women | Men | Women | Men | Women | Men | Women | Men | Women | Men | Women | Men |
| LV long-axis strain 4CV (%) | n<5 | n<5 | n<5 | n<5 | = | = | = | = | = | = | = | = |
| LV long-axis strain avg (%) | n<5 | n<5 | n<5 | n<5 | = | ↑ | = | = | = | = | = | = |
| LV AV-junction strain 4CV (%) | n<5 | n<5 | n<5 | n<5 | = | = | = | = | = | = | = | = |
| LV AV-junction strain avg (%) | n<5 | n<5 | n<5 | n<5 | = | ↑ | = | = | = | = | = | = |

**Black vs. White ethnicity**

| Metric | 18-29 | | 30-39 | | 40-49 | | 50-59 | | 60-69 | | >70 | |
|---|---|---|---|---|---|---|---|---|---|---|---|---|
| | Women | Men | Women | Men | Women | Men | Women | Men | Women | Men | Women | Men |
| LV long-axis strain 4CV (%) | n<5 | n<5 | n<5 | n<5 | = | = | = | = | = | = | ↓ | = |
| LV long-axis strain avg (%) | n<5 | n<5 | n<5 | n<5 | = | = | = | = | ↓ | = | = | = |
| LV AV-junction strain 4CV (%) | n<5 | n<5 | n<5 | n<5 | = | = | = | = | = | = | ↓ | = |
| LV AV-junction strain avg (%) | n<5 | n<5 | n<5 | n<5 | = | = | = | = | ↓ | = | = | = |

**Chinese vs. White ethnicity**

| Metric | 18-29 | | 30-39 | | 40-49 | | 50-59 | | 60-69 | | >70 | |
|---|---|---|---|---|---|---|---|---|---|---|---|---|
| | Women | Men | Women | Men | Women | Men | Women | Men | Women | Men | Women | Men |
| LV long-axis strain 4CV (%) | = | = | = | = | = | = | = | = | = | = | = | n<5 |
| LV long-axis strain avg (%) | = | = | = | = | = | = | = | = | = | = | = | n<5 |
| LV AV-junction strain 4CV (%) | = | = | = | = | = | = | = | = | = | = | = | n<5 |
| LV AV-junction strain avg (%) | = | = | = | = | = | = | = | = | = | = | = | n<5 |

**Mixed/Other vs. White ethnicity**

| Metric | 18-29 | | 30-39 | | 40-49 | | 50-59 | | 60-69 | | >70 | |
|---|---|---|---|---|---|---|---|---|---|---|---|---|
| | Women | Men | Women | Men | Women | Men | Women | Men | Women | Men | Women | Men |
| LV long-axis strain 4CV (%) | n<5 | n<5 | n<5 | n<5 | = | = | = | = | = | = | = | = |
| LV long-axis strain avg (%) | n<5 | n<5 | n<5 | n<5 | = | = | = | = | = | = | = | = |
| LV AV-junction strain 4CV (%) | n<5 | n<5 | n<5 | n<5 | = | = | = | = | = | = | = | = |
| LV AV-junction strain avg (%) | n<5 | n<5 | n<5 | n<5 | = | = | = | = | = | = | = | = |

**Footnote:** Comparison of left ventricular (LV) long-axis strain (4-chamber view and average) and LV atrioventricular (AV) junction strain (4-chamber view and average) between different ethnic groups (South Asian, Black, Chinese, and Mixed/Other) and the White reference population, stratified by age group and sex, derived from rapid strain analysis using cvi42 (Circle Cardiovascular Imaging Inc.). Arrows indicate direction of difference relative to the White group (↑ higher, ↓ lower), while "=" denotes no relevant difference. Cells marked "n<5" indicate insufficient sample size for reliable comparison. Group comparisons were performed using Wilcoxon rank-sum tests with adjustment for multiple testing.

**Abbreviations:** 4CV = four-chamber view; AV = atrioventricular; LV = left ventricle.

### Supplementary Figure 16 Ethnicity-specific differences in atrial long-axis and atrioventricular junction strain across age and sex groups using cvi42 (Circle Cardiovascular Imaging Inc.)

**Asian vs. White ethnicity**

| Metric | 18-29 | | 30-39 | | 40-49 | | 50-59 | | 60-69 | | >70 | |
|---|---|---|---|---|---|---|---|---|---|---|---|---|
| | Women | Men | Women | Men | Women | Men | Women | Men | Women | Men | Women | Men |
| LA long-axis strain 4CV (%) | n<5 | n<5 | n<5 | n<5 | = | = | = | = | = | = | = | = |
| LA long-axis strain avg (%) | n<5 | n<5 | n<5 | n<5 | = | = | = | ↑ | = | = | = | = |
| LA AV-junction strain 4CV (%) | n<5 | n<5 | n<5 | n<5 | = | = | = | = | = | = | = | = |
| LA AV-junction strain avg (%) | n<5 | n<5 | n<5 | n<5 | = | = | = | ↑ | = | = | = | = |
| RA long-axis strain (%) | n<5 | n<5 | n<5 | n<5 | = | = | = | = | = | = | = | = |
| RA AV-junction strain (%) | n<5 | n<5 | n<5 | n<5 | = | = | = | ↑ | = | = | = | = |

**Black vs. White ethnicity**

| Metric | 18-29 | | 30-39 | | 40-49 | | 50-59 | | 60-69 | | >70 | |
|---|---|---|---|---|---|---|---|---|---|---|---|---|
| | Women | Men | Women | Men | Women | Men | Women | Men | Women | Men | Women | Men |
| LA long-axis strain 4CV (%) | n<5 | n<5 | n<5 | n<5 | = | = | = | = | = | = | = | = |
| LA long-axis strain avg (%) | n<5 | n<5 | n<5 | n<5 | = | = | = | = | = | = | = | = |
| LA AV-junction strain 4CV (%) | n<5 | n<5 | n<5 | n<5 | = | = | = | = | = | = | = | = |
| LA AV-junction strain avg (%) | n<5 | n<5 | n<5 | n<5 | = | = | = | = | = | = | = | = |
| RA long-axis strain (%) | n<5 | n<5 | n<5 | n<5 | = | = | = | = | = | = | = | = |
| RA AV-junction strain (%) | n<5 | n<5 | n<5 | n<5 | = | = | = | = | = | = | = | = |

**Chinese vs. White ethnicity**

| Metric | 18-29 | | 30-39 | | 40-49 | | 50-59 | | 60-69 | | >70 | |
|---|---|---|---|---|---|---|---|---|---|---|---|---|
| | Women | Men | Women | Men | Women | Men | Women | Men | Women | Men | Women | Men |
| LA long-axis strain 4CV (%) | = | = | = | = | = | = | = | = | = | = | = | n<5 |
| LA long-axis strain avg (%) | = | = | = | = | = | = | = | = | = | = | = | n<5 |
| LA AV-junction strain 4CV (%) | = | = | = | = | = | = | = | = | = | = | = | n<5 |
| LA AV-junction strain avg (%) | = | = | = | = | = | = | = | = | = | = | = | n<5 |
| RA long-axis strain (%) | = | = | = | = | ↑ | = | ↑ | = | = | = | = | n<5 |
| RA AV-junction strain (%) | = | = | = | = | ↑ | = | ↑ | = | = | = | = | n<5 |

**Mixed/Other vs. White ethnicity**

| Metric | 18-29 | | 30-39 | | 40-49 | | 50-59 | | 60-69 | | >70 | |
|---|---|---|---|---|---|---|---|---|---|---|---|---|
| | Women | Men | Women | Men | Women | Men | Women | Men | Women | Men | Women | Men |
| LA long-axis strain 4CV (%) | n<5 | n<5 | n<5 | n<5 | = | = | = | = | = | = | = | = |
| LA long-axis strain avg (%) | n<5 | n<5 | n<5 | n<5 | = | = | = | = | = | = | = | = |
| LA AV-junction strain 4CV (%) | n<5 | n<5 | n<5 | n<5 | = | = | = | = | = | = | = | = |
| LA AV-junction strain avg (%) | n<5 | n<5 | n<5 | n<5 | = | = | = | = | = | = | = | = |
| RA long-axis strain (%) | n<5 | n<5 | n<5 | n<5 | = | = | = | = | = | = | = | = |
| RA AV-junction strain (%) | n<5 | n<5 | n<5 | n<5 | = | = | = | = | = | = | = | = |

**Footnote:** Comparison of left atrial (LA) and right atrial (RA) long-axis strain (4-chamber view and average where applicable) and atrioventricular (AV) junction strain between different ethnic groups (South Asian, Black, Chinese, and Mixed/Other) and the White reference population, stratified by age group and sex, derived from rapid strain analysis using cvi42 (Circle Cardiovascular Imaging Inc.). Arrows indicate direction of difference relative to the White group (↑ higher, ↓ lower), while "=" denotes no relevant difference. Cells marked "n<5" indicate insufficient sample size for reliable comparison. Group comparisons were performed using Wilcoxon rank-sum tests with adjustment for multiple testing.

**Abbreviations:** 4CV = four-chamber view; AV = atrioventricular; LA = left atrium; RA = right atrium.

### Supplementary Figure 17 Magnet strength and scanner vendor–specific differences in strain parameters using cvi42 (Circle Cardiovascular Imaging Inc.)

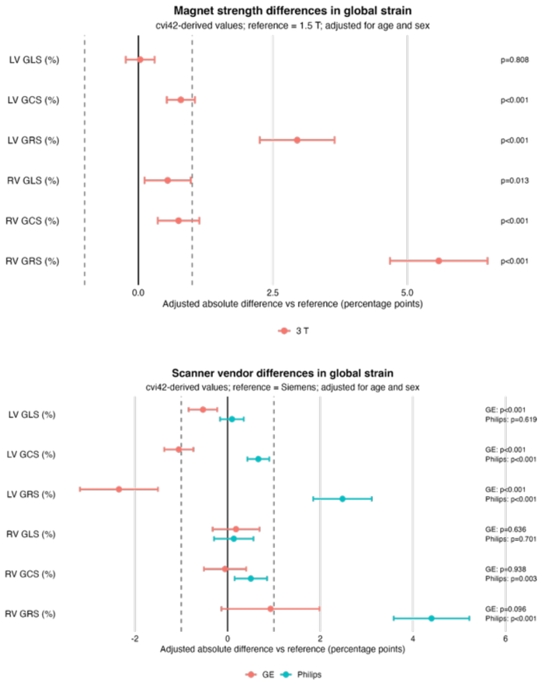

**Footnote:**
Combined analysis of strain parameter differences according to magnet strength and scanner vendor, using 1.5T and Siemens scanners as reference categories, derived from CMR feature tracking using cvi42 (Circle Cardiovascular Imaging Inc.). Points represent effect estimates and horizontal bars indicate 95% confidence intervals. Vertical solid lines denote no difference relative to the reference category, while dashed lines indicate predefined equivalence margins where applicable. Positive values reflect higher strain values and negative values indicate lower values compared with the respective reference. Statistical comparisons were performed using regression-based models with adjustment for relevant covariates; confidence intervals crossing the reference line indicate no statistically significant difference.
**Abbreviations:** LV = left ventricle; RV = right ventricle; GCS = global circumferential strain; GLS = global longitudinal strain; GRS = global radial strain.

**Supplementary Figure 18 Magnet strength and scanner vendor–specific differences in strain parameters using suiteHEART (NeoSoft)**

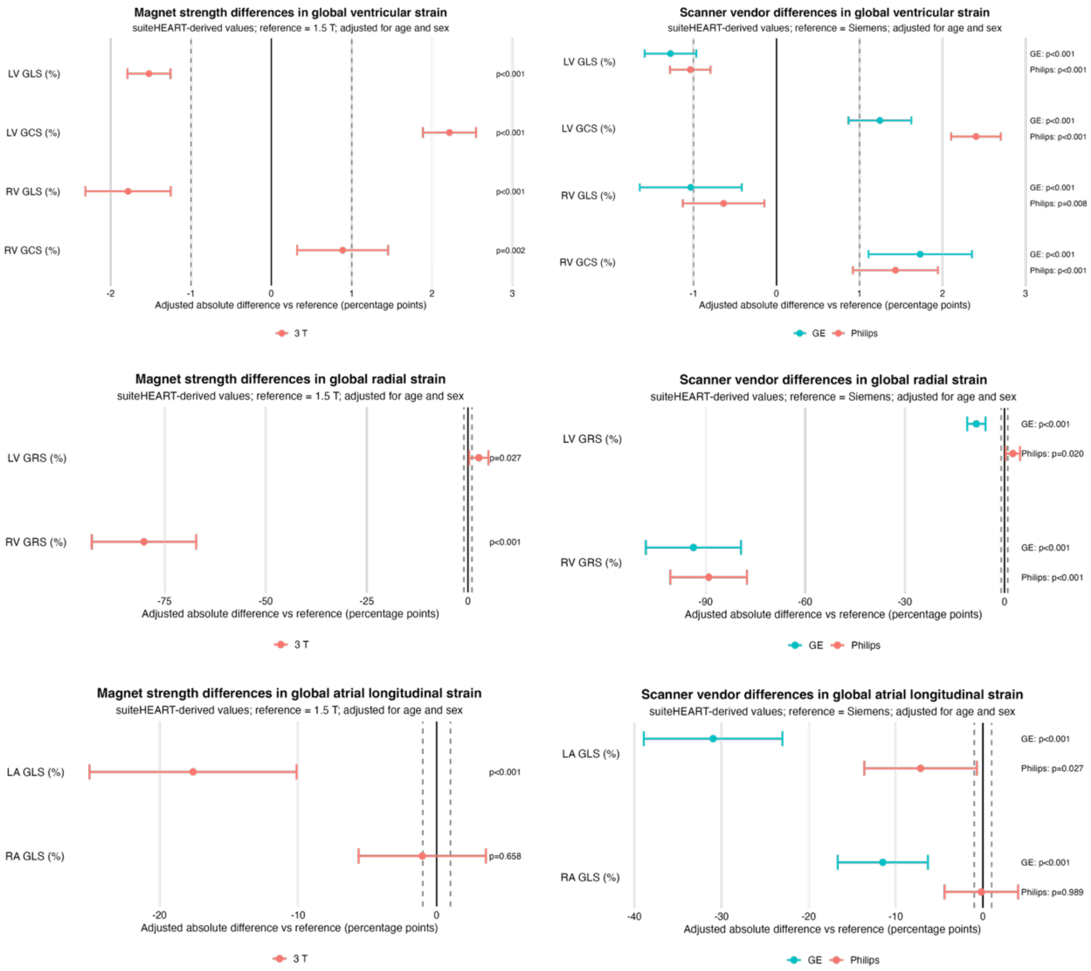

**Footnote:**
Combined analysis of strain parameter differences according to magnet strength and scanner vendor, using 1.5T and Siemens scanners as reference categories, derived from CMR feature tracking using suiteHEART (NeoSoft). Points represent effect estimates and horizontal bars indicate 95% confidence intervals. Vertical solid lines denote no difference relative to the reference category, while dashed lines indicate predefined equivalence margins where applicable. Positive values reflect higher strain values and negative values indicate lower values compared with the respective reference. Statistical comparisons were performed using regression-based models with adjustment for relevant covariates; confidence intervals crossing the reference line indicate no statistically significant difference.
**Abbreviations:** LA, left atrium; LV = left ventricle; RV = right ventricle; RA, right atrium; GCS = global circumferential strain; GLS = global longitudinal strain; GRS = global radial strain.

### Supplementary Figure 19 Magnet strength– and scanner vendor–related differences in MAPSE, TAPSE, and ventricular and atrial rapid strain parameters across age and sex groups using cvi42 (Circle Cardiovascular Imaging Inc.)

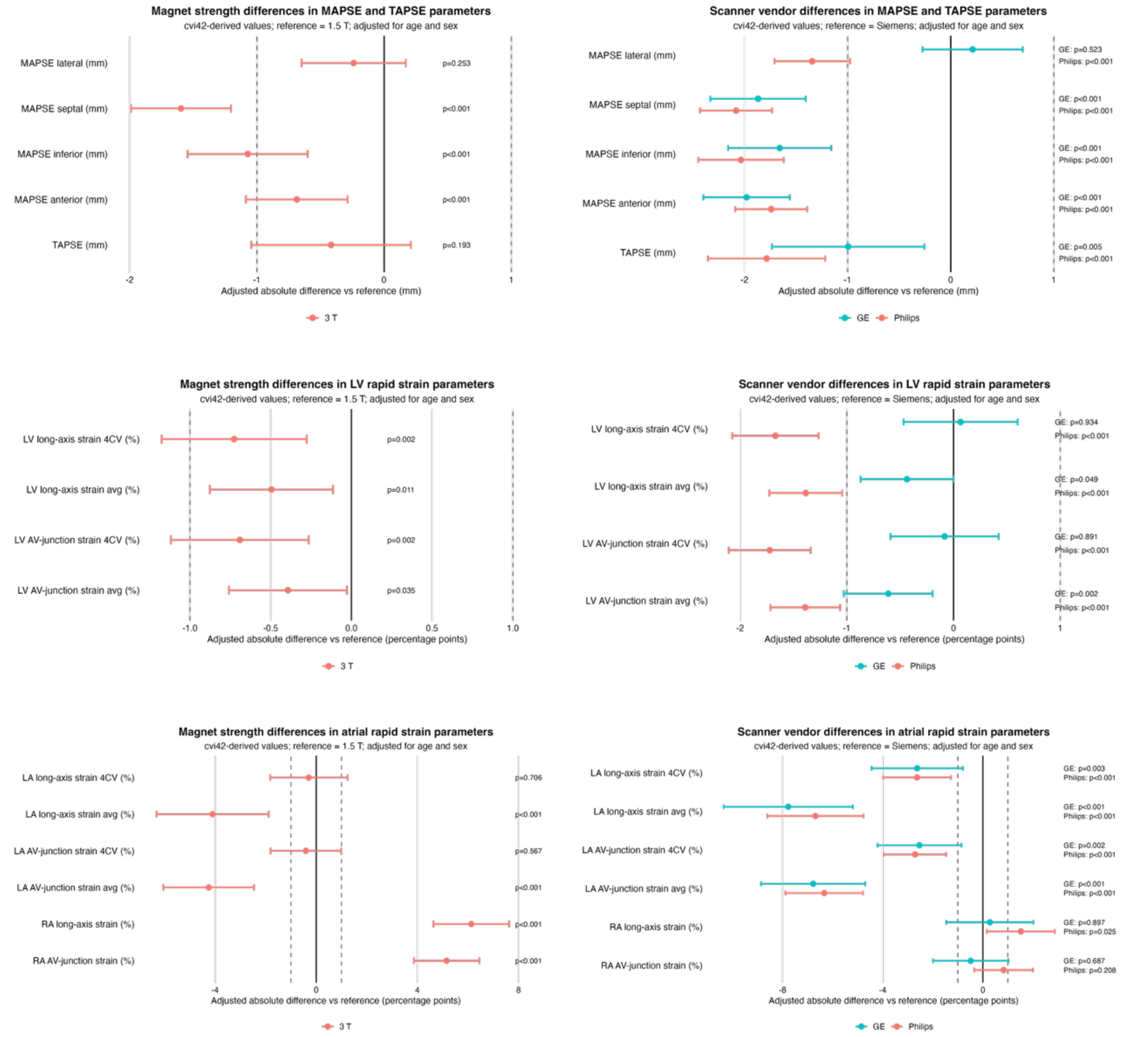

**Footnote:**
Combined analysis of MAPSE, TAPSE, and rapid strain parameters including left ventricular (LV) long-axis strain and atrioventricular (AV) junction strain, as well as left atrial (LA) and right atrial (RA) long-axis and AV junction strain, according to magnet strength and scanner vendor. Siemens scanners and 1.5T were used as reference categories. All measurements were derived from rapid strain analysis using cvi42 (Circle Cardiovascular Imaging Inc.). Points represent adjusted effect estimates and horizontal bars indicate 95% confidence intervals. Solid vertical lines denote no difference relative to the reference category, while dashed lines indicate predefined equivalence margins, where applicable. Positive values indicate higher values and negative values indicate lower values compared with the respective reference. Statistical comparisons were performed using multivariable regression models adjusted for age and sex; confidence intervals crossing the reference line indicate no statistically significant difference.

**Abbreviations:** AV = atrioventricular; LA = left atrium; LV = left ventricle; RA = right atrium; RV = right ventricle; MAPSE = mitral annular plane systolic excursion; TAPSE = tricuspid annular plane systolic excursion.

### Supplementary Figure 20 Agreement between suiteHEART (NeoSoft) and cvi42 (Circle Cardiovascular Imaging Inc.) for ventricular strain measurements (Bland–Altman analysis)

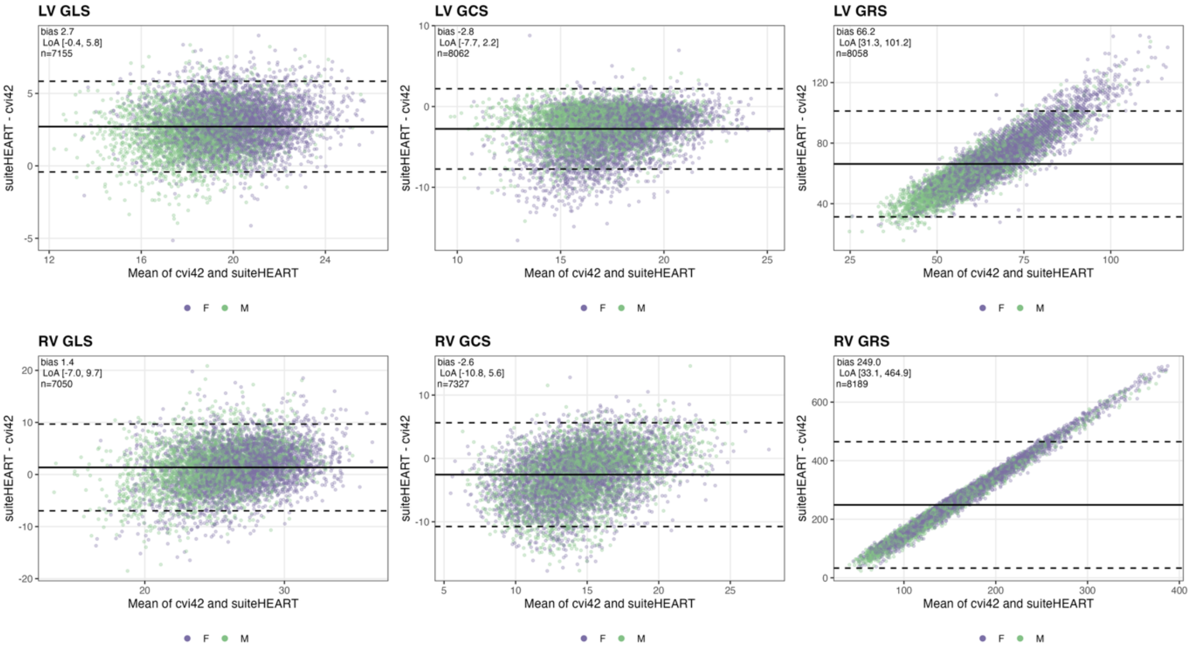

**Footnote:**
Bland–Altman plots comparing left ventricular (LV) and right ventricular (RV) global longitudinal (GLS), circumferential (GCS), and radial (GRS) strain derived from the same examinations using two post-processing software platforms, cvi42 (Circle Cardiovascular Imaging Inc.) and suiteHEART (NeoSoft Inc.). For each metric, the difference between the two measurements (suiteHEART − cvi42) is plotted against the mean of the two measurements. The solid horizontal line denotes the mean difference (bias), and the dashed horizontal lines denote the 95% limits of agreement (bias ± 1.96·SD of the differences), assuming approximately normally distributed differences. The bias, 95% limits of agreement (LoA), and number of paired observations (n) are annotated in each panel. Points are coloured by sex. All strain values are presented as absolute values.
**Abbreviations**: LV = left ventricle; RV = right ventricle; GCS = global circumferential strain; GLS = global longitudinal strain; GRS = global radial strain; F = female; M = male; LoA = limits of agreement; SD = standard deviation.

Supplementary Table 14 Left and right ventricular global strain reference ranges and centile-based categories stratified by sex and age using cvi42 (Circle Cardiovascular Imaging Inc.), *in a South Asian adult population* .40

Supplementary Table 15 Left and right ventricular global strain rate reference ranges and centile-based categories stratified by sex and age using cvi42 (Circle Cardiovascular Imaging Inc.), *in a South Asian adult population* ....41

Supplementary Table 16 Left ventricular global displacement reference ranges and centile-based categories stratified by sex and age using cvi42 (Circle Cardiovascular Imaging Inc.), *in a South Asian adult population* .42

**Supplementary Table 1 Baseline characteristics of cohorts after quality control using suiteHEART (NeoSoft)**

| | **Whole cohort (n=8,992)** | **Women (n=4,597)** | **Men (n=4,395)** |
|---|---|---|---|
| **Age, years** | | | |
| 18–29 | 247 (2.7%) | 120 (2.6%) | 127 (2.9%) |
| 30–39 | 303 (3.4%) | 154 (3.4%) | 149 (3.4%) |
| 40–49 | 1,075 (12.0%) | 571 (12.4%) | 504 (11.5%) |
| 50–59 | 2,475 (27.5%) | 1,308 (28.5%) | 1,167 (26.6%) |
| 60–69 | 2,299 (25.6%) | 1,217 (26.5%) | 1,082 (24.6%) |
| >70 | 2,593 (28.8%) | 1,227 (26.7%) | 1,366 (31.1%) |
| **Ethnicity** | | | |
| White | 7,345 (81.7%) | 3,730 (81.1%) | 3,615 (82.3%) |
| South Asian | 502 (5.6%) | 186 (4.1%) | 316 (7.2%) |
| Black | 334 (3.7%) | 188 (4.1%) | 146 (3.3%) |
| Chinese | 335 (3.7%) | 188 (4.1%) | 147 (3.3%) |
| Other/Mixed | 476 (5.3%) | 305 (6.6%) | 171 (3.9%) |
| **Cohort** | | | |
| UK Biobank | 7,600 (84.5%) | 3,891 (84.6%) | 3,709 (84.4%) |
| SHIP | 626 (7.0%) | 328 (7.1%) | 298 (6.8%) |
| Pisa | 283 (3.2%) | 144 (3.1%) | 139 (3.2%) |
| Heidelberg | 188 (2.1%) | 84 (1.8%) | 104 (2.4%) |
| Singapore | 180 (2.0%) | 89 (1.9%) | 91 (2.1%) |
| Utrecht | 115 (1.3%) | 61 (1.3%) | 54 (1.2%) |
| **Scanner Vendor** | | | |
| Siemens | 8,226 (91.5%) | 4,219 (91.8%) | 4,007 (91.2%) |
| Philips | 483 (5.4%) | 234 (5.1%) | 249 (5.7%) |
| GE | 283 (3.2%) | 144 (3.1%) | 139 (3.2%) |
| **Magnet Strength** | | | |
| 1.5 T | 8,696 (96.7%) | 4,446 (96.7%) | 4,250 (96.7%) |
| 3 T | 296 (3.3%) | 151 (3.3%) | 145 (3.3%) |

**Footnote:** Values are n (%). SHIP = Study of Health in Pomerania; T = Tesla.

**Supplementary Table 2 Baseline characteristics of the study population after quality control of annular plane systolic excursion and rapid longitudinal shortening metrics derived using cvi42 (Circle Cardiovascular Imaging Inc)**

| | Whole cohort (n=8,984) | Women (n=4,598) | Men (n=4,386) |
|---|---|---|---|
| **Age, years** | | | |
| 18–29 | 240 (2.7%) | 115 (2.5%) | 125 (2.9%) |
| 30–39 | 304 (3.4%) | 153 (3.3%) | 151 (3.4%) |
| 40–49 | 1,078 (12.0%) | 573 (12.5%) | 505 (11.5%) |
| 50–59 | 2,476 (27.6%) | 1,309 (28.5%) | 1,167 (26.6%) |
| 60–69 | 2,302 (25.6%) | 1,225 (26.6%) | 1,077 (24.6%) |
| >70 | 2,584 (28.8%) | 1,223 (26.6%) | 1,361 (31.0%) |
| **Ethnicity** | | | |
| White | 7,342 (81.7%) | 3,731 (81.1%) | 3,611 (82.3%) |
| South Asian | 503 (5.6%) | 186 (4.1%) | 317 (7.2%) |
| Black | 335 (3.7%) | 190 (4.1%) | 145 (3.3%) |
| Chinese | 331 (3.7%) | 187 (4.1%) | 144 (3.3%) |
| Other/Mixed | 473 (5.3%) | 304 (6.6%) | 169 (3.9%) |
| **Cohort** | | | |
| UK Biobank | 7,596 (84.6%) | 3,895 (84.7%) | 3,701 (84.4%) |
| SHIP | 623 (6.9%) | 328 (7.1%) | 295 (6.7%) |
| Pisa | 288 (3.2%) | 145 (3.2%) | 143 (3.3%) |
| Heidelberg | 191 (2.1%) | 86 (1.9%) | 105 (2.4%) |
| Singapore | 177 (2.0%) | 88 (1.9%) | 89 (2.0%) |
| Utrecht | 109 (1.2%) | 56 (1.2%) | 53 (1.2%) |
| **Scanner Vendor** | | | |
| Siemens | 8,219 (91.5%) | 4,223 (91.8%) | 3,996 (91.1%) |
| Philips | 477 (5.3%) | 230 (5.0%) | 247 (5.6%) |
| GE | 288 (3.2%) | 145 (3.2%) | 143 (3.3%) |
| **Magnet Strength** | | | |
| 1.5 T | 8,698 (96.8%) | 4,454 (96.9%) | 4,244 (96.8%) |
| 3 T | 286 (3.2%) | 144 (3.1%) | 142 (3.2%) |

**Footnote:** Values are n (%). SHIP: Study of Health in Pomerania; T: Tesla.

### Supplementary Table 3 Agreement between suiteHEART (NeoSoft) and cvi42 (Circle Cardiovascular Imaging Inc.) for ventricular strain measurements

| Metric | n | cvi42 mean ± SD | suiteHEART mean ± SD | Bias (suiteHEART − cvi42) | 95% limits of agreement | Relative bias, % | Pearson r |
|---|---|---|---|---|---|---|---|
| **LV GLS** | 7,155 | 18.0 ± 2.0 | 20.7 ± 2.2 | +2.7 | −0.4 to 5.8 | +14.0 | 0.72 |
| **LV GCS** | 8,062 | 18.6 ± 2.2 | 15.8 ± 2.7 | −2.8 | −7.7 to 2.2 | −16.1 | 0.48 |
| **LV GRS** | 8,058 | 31.1 ± 5.8 | 97.3 ± 21.1 | +66.2 | 31.3 to 101.2 | +103.2 | 0.66 |
| **RV GLS** | 7,050 | 25.4 ± 3.3 | 26.8 ± 4.3 | +1.4 | −7.0 to 9.7 | +5.2 | 0.39 |
| **RV GCS** | 7,327 | 15.7 ± 3.0 | 13.2 ± 4.3 | −2.6 | −10.8 to 5.6 | −17.7 | 0.39 |
| **RV GRS** | 8,189 | 26.0 ± 7.1 | 275.0 ± 109.8 | +249.0 | 33.1 to 464.9 | +165.5 | −0.01 |

**Footnote:** Paired ventricular strain measurements derived from the same examinations using two post-processing software platforms, cvi42 (Circle Cardiovascular Imaging Inc.) and suiteHEART (NeoSoft Inc.). For each metric the bias was defined as the mean of the paired differences (suiteHEART − cvi42), and the 95% limits of agreement as the bias ± 1.96·SD of the differences, assuming approximately normally distributed differences. Relative bias is expressed as a percentage of the mean cvi42 value. The linear association between platforms is summarised by the Pearson correlation coefficient (r). n denotes the number of paired observations. All strain values are presented as absolute values.
**Abbreviations**: LV = left ventricle; RV = right ventricle; GCS = global circumferential strain; GLS = global longitudinal strain; GRS = global radial strain; SD = standard deviation.

**Supplementary Table 4 Left ventricular global displacement reference ranges and centile-based categories stratified by sex and age for all ethnicities using cvi42 (Circle Cardiovascular Imaging Inc.)**

| | | Female | | | | Male | | | |
|---|---|---|---|---|---|---|---|---|---|
| **Age group** | Metrics (mm) | Reference threshold | Mild | At least moderate | n | Reference threshold | Mild | At least moderate | n |
| **18–29** | LV GLD | ≥ 3.5 | 2.9–3.4 | < 2.9 | 76 | ≥ 2.5 | 1.9–2.4 | < 1.9 | 93 |
| | LV GCD | ≥ 0.8 | 0.6–0.7 | < 0.6 | 89 | ≥ 0.8 | 0.6–0.7 | < 0.6 | 104 |
| | LV GRD | ≥ 4.0 | 3.7–3.9 | < 3.7 | 105 | ≥ 4.0 | 3.6–3.9 | < 3.6 | 120 |
| **30–39** | LV GLD | ≥ 3.5 | 2.8–3.4 | < 2.8 | 125 | ≥ 2.7 | 2.1–2.6 | < 2.1 | 125 |
| | LV GCD | ≥ 0.8 | 0.5–0.7 | < 0.5 | 122 | ≥ 0.8 | 0.6–0.7 | < 0.6 | 130 |
| | LV GRD | ≥ 4.0 | 3.7–3.9 | < 3.7 | 140 | ≥ 3.9 | 3.5–3.8 | < 3.5 | 137 |
| **40–49** | LV GLD | ≥ 3.3 | 2.6–3.2 | < 2.6 | 530 | ≥ 2.7 | 2.1–2.6 | < 2.1 | 454 |
| | LV GCD | ≥ 0.7 | 0.5–0.6 | < 0.5 | 459 | ≥ 0.8 | 0.5–0.7 | < 0.5 | 424 |
| | LV GRD | ≥ 4.0 | 3.7–3.9 | < 3.7 | 521 | ≥ 3.9 | 3.5–3.8 | < 3.5 | 461 |
| **50–59** | LV GLD | ≥ 2.9 | 2.3–2.8 | < 2.3 | 1206 | ≥ 2.4 | 1.8–2.3 | < 1.8 | 1028 |
| | LV GCD | ≥ 0.7 | 0.5–0.6 | < 0.5 | 940 | ≥ 0.8 | 0.5–0.7 | < 0.5 | 971 |
| | LV GRD | ≥ 4.0 | 3.6–3.9 | < 3.6 | 1187 | ≥ 3.8 | 3.4–3.7 | < 3.4 | 1056 |
| **60–69** | LV GLD | ≥ 2.7 | 2.1–2.6 | < 2.1 | 1132 | ≥ 2.3 | 1.8–2.2 | < 1.8 | 954 |
| | LV GCD | ≥ 0.7 | 0.5–0.6 | < 0.5 | 856 | ≥ 0.8 | 0.5–0.7 | < 0.5 | 852 |
| | LV GRD | ≥ 3.9 | 3.5–3.8 | < 3.5 | 1129 | ≥ 3.8 | 3.3–3.7 | < 3.3 | 975 |
| **70+** | LV GLD | ≥ 2.4 | 1.8–2.3 | < 1.8 | 1104 | ≥ 2.3 | 1.8–2.2 | < 1.8 | 1169 |
| | LV GCD | ≥ 0.6 | 0.5–0.5 | < 0.5 | 808 | ≥ 0.8 | 0.5–0.7 | < 0.5 | 1052 |
| | LV GRD | ≥ 3.9 | 3.4–3.8 | < 3.4 | 1130 | ≥ 3.7 | 3.3–3.6 | < 3.3 | 1216 |

**Footnote:** All displacement measurements were derived using cvi42 software (Circle Cardiovascular Imaging Inc.) with standardized feature-tracking analysis. Global longitudinal displacement (GLD), global circumferential displacement (GCD), and global radial displacement (GRD) are presented in millimetres (mm). Reference ranges are defined as the interval between the 2.5th and 97.5th centiles derived from age- and sex-specific generalized additive models for location, scale and shape (GAMLSS) using the Box–Cox Cole–Green distribution. Centile-based categories beyond the reference range are derived from LMS-based thresholds (P0.5 and P2.5) and describe the position of a measurement within the reference distribution rather than disease severity or diagnostic abnormality. For clarity, only the lower reference threshold (2.5th percentile) and corresponding lower centile cut-offs are presented in this table. Reference values are based on all ethnicities.
**Abbreviations**: LV = left ventricle; GLD = global longitudinal displacement; GCD = global circumferential displacement; GRD = global radial displacement.

**Supplementary Table 5 Left ventricular global velocity reference ranges and centile-based categories stratified by sex and age for all ethnicities using cvi42 (Circle Cardiovascular Imaging Inc.)**

| | | Female | | | | Male | | | |
|---|---|---|---|---|---|---|---|---|---|
| **Age group** | Metrics (%/s) | Reference threshold | Mild | At least moderate | n | Reference threshold | Mild | At least moderate | n |
| **18–29** | LV sysGLV | ≥ 16.4 | 13.3–16.3 | < 13.3 | 74 | ≥ 14.0 | 10.8–13.9 | < 10.8 | 90 |
| | LV sysGCV | ≥ 11.8 | 8.1–11.7 | < 8.1 | 91 | ≥ 12.9 | 9.0–12.8 | < 9.0 | 111 |
| | LV sysGRV | ≥ 20.9 | 18.7–20.8 | < 18.7 | 103 | ≥ 21.7 | 19.0–21.6 | < 19.0 | 120 |
| | LV diaGLV | ≥ 13.7 | 6.6–13.6 | < 6.6 | 73 | ≥ 10.9 | 5.3–10.8 | < 5.3 | 92 |
| | LV diaGCV | ≥ 5.9 | 3.9–5.8 | < 3.9 | 73 | ≥ 5.1 | 3.8–5.0 | < 3.8 | 80 |
| | LV diaGRV | ≥ 15.0 | 12.0–14.9 | < 12.0 | 105 | ≥ 14.7 | 11.4–14.6 | < 11.4 | 118 |
| **30–39** | LV sysGLV | ≥ 19.5 | 16.4–19.4 | < 16.4 | 119 | ≥ 15.4 | 11.6–15.3 | < 11.6 | 118 |
| | LV sysGCV | ≥ 11.2 | 7.9–11.1 | < 7.9 | 127 | ≥ 12.4 | 8.8–12.3 | < 8.8 | 130 |
| | LV sysGRV | ≥ 21.4 | 19.4–21.3 | < 19.4 | 140 | ≥ 22.6 | 20.5–22.5 | < 20.5 | 136 |
| | LV diaGLV | ≥ 16.8 | 9.2–16.7 | < 9.2 | 119 | ≥ 14.8 | 10.5–14.7 | < 10.5 | 125 |
| | LV diaGCV | ≥ 6.1 | 4.2–6.0 | < 4.2 | 92 | ≥ 5.1 | 3.8–5.0 | < 3.8 | 101 |
| | LV diaGRV | ≥ 16.5 | 13.9–16.4 | < 13.9 | 138 | ≥ 16.1 | 13.5–16.0 | < 13.5 | 136 |
| **40–49** | LV sysGLV | ≥ 18.2 | 15.6–18.1 | < 15.6 | 519 | ≥ 15.6 | 12.4–15.5 | < 12.4 | 437 |
| | LV sysGCV | ≥ 10.6 | 7.7–10.5 | < 7.7 | 466 | ≥ 12.0 | 8.6–11.9 | < 8.6 | 438 |
| | LV sysGRV | ≥ 21.0 | 19.2–20.9 | < 19.2 | 519 | ≥ 22.4 | 20.5–22.3 | < 20.5 | 460 |
| | LV diaGLV | ≥ 18.4 | 13.8–18.3 | < 13.8 | 525 | ≥ 15.6 | 12.3–15.5 | < 12.3 | 453 |
| | LV diaGCV | ≥ 6.6 | 4.9–6.5 | < 4.9 | 356 | ≥ 5.1 | 3.8–5.0 | < 3.8 | 375 |
| | LV diaGRV | ≥ 17.8 | 15.3–17.7 | < 15.3 | 520 | ≥ 16.2 | 13.9–16.1 | < 13.9 | 457 |
| **50–59** | LV sysGLV | ≥ 15.3 | 13.2–15.2 | < 13.2 | 1200 | ≥ 14.8 | 12.5–14.7 | < 12.5 | 1017 |
| | LV sysGCV | ≥ 9.9 | 7.3–9.8 | < 7.3 | 989 | ≥ 11.6 | 8.4–11.5 | < 8.4 | 979 |
| | LV sysGRV | ≥ 19.9 | 18.3–19.8 | < 18.3 | 1186 | ≥ 21.5 | 19.6–21.4 | < 19.6 | 1053 |
| | LV diaGLV | ≥ 16.8 | 13.9–16.7 | < 13.9 | 1201 | ≥ 14.5 | 12.0–14.4 | < 12.0 | 1025 |
| | LV diaGCV | ≥ 5.6 | 4.2–5.5 | < 4.2 | 787 | ≥ 5.2 | 3.9–5.1 | < 3.9 | 831 |
| | LV diaGRV | ≥ 17.2 | 15.0–17.1 | < 15.0 | 1184 | ≥ 16.0 | 13.9–15.9 | < 13.9 | 1048 |
| **60–69** | LV sysGLV | ≥ 14.8 | 12.8–14.7 | < 12.8 | 1127 | ≥ 15.2 | 13.1–15.1 | < 13.1 | 950 |
| | LV sysGCV | ≥ 9.4 | 7.1–9.3 | < 7.1 | 851 | ≥ 11.2 | 8.2–11.1 | < 8.2 | 857 |
| | LV sysGRV | ≥ 19.0 | 17.5–18.9 | < 17.5 | 1129 | ≥ 20.2 | 18.3–20.1 | < 18.3 | 973 |
| | LV diaGLV | ≥ 16.3 | 13.7–16.2 | < 13.7 | 1128 | ≥ 14.6 | 12.1–14.5 | < 12.1 | 947 |
| | LV diaGCV | ≥ 5.4 | 4.1–5.3 | < 4.1 | 732 | ≥ 5.3 | 4.0–5.2 | < 4.0 | 754 |
| | LV diaGRV | ≥ 16.3 | 14.3–16.2 | < 14.3 | 1126 | ≥ 15.9 | 14.0–15.8 | < 14.0 | 974 |
| **70+** | LV sysGLV | ≥ 13.9 | 11.8–13.8 | < 11.8 | 1100 | ≥ 15.2 | 12.9–15.1 | < 12.9 | 1164 |
| | LV sysGCV | ≥ 9.1 | 7.0–9.0 | < 7.0 | 742 | ≥ 10.6 | 7.9–10.5 | < 7.9 | 965 |
| | LV sysGRV | ≥ 18.4 | 16.8–18.3 | < 16.8 | 1130 | ≥ 19.4 | 17.6–19.3 | < 17.6 | 1215 |
| | LV diaGLV | ≥ 15.8 | 12.9–15.7 | < 12.9 | 1099 | ≥ 14.1 | 11.1–14.0 | < 11.1 | 1161 |
| | LV diaGCV | ≥ 5.2 | 3.9–5.1 | < 3.9 | 696 | ≥ 5.4 | 4.1–5.3 | < 4.1 | 968 |
| | LV diaGRV | ≥ 16.1 | 14.2–16.0 | < 14.2 | 1126 | ≥ 15.2 | 13.4–15.1 | < 13.4 | 1210 |

**Footnote:** All velocity measurements were derived using cvi42 software (Circle Cardiovascular Imaging Inc.) with standardized feature-tracking analysis. Global longitudinal velocity (GLV), global circumferential velocity (GCV), and global radial velocity (GRV) are presented in %/s. Reference ranges are defined as the interval between the 2.5th and 97.5th centiles derived from age- and sex-specific generalized additive models for location, scale and shape (GAMLSS) using the Box–Cox Cole–Green distribution. Centile-based categories beyond the reference range are derived from LMS-based thresholds (P0.5 and P2.5) and describe the position of a measurement within the reference distribution rather than disease severity or diagnostic abnormality. For clarity, only the lower reference threshold (2.5th percentile) and corresponding lower centile cut-offs are presented in this table. Reference values are based on all ethnicities.
**Abbreviations**: LV = left ventricle; GLV = global longitudinal velocity; GCV = global circumferential velocity; GRV = global radial velocity; sys = systolic; dia = diastolic.

**Supplementary Table 6 Left and right ventricular global strain reference ranges and centile-based categories stratified by sex and age using cvi42 (Circle Cardiovascular Imaging Inc.), *in a White adult population***

| | | Female | | | | Male | | | |
|---|---|---|---|---|---|---|---|---|---|
| **Age group** | Metrics (%) | Reference threshold | Mild | At least moderate | n | Reference threshold | Mild | At least moderate | n |
| **18–29** | LV GLS | ≥ 15.4 | 14.3–15.3 | < 14.3 | 64 | ≥ 13.7 | 12.4–13.6 | < 12.4 | 75 |
| | LV GCS | ≥ 14.9 | 13.9–14.8 | < 13.9 | 90 | ≥ 13.5 | 12.3–13.4 | < 12.3 | 101 |
| | LV GRS | ≥ 21.9 | 20.0–21.8 | < 20.0 | 90 | ≥ 19.2 | 17.2–19.1 | < 17.2 | 101 |
| | RV GLS | ≥ 18.3 | 15.8–18.2 | < 15.8 | 75 | ≥ 16.7 | 14.2–16.6 | < 14.2 | 80 |
| | RV GCS | ≥ 9.0 | 7.4–8.9 | < 7.4 | 86 | ≥ 8.9 | 7.0–8.8 | < 7.0 | 101 |
| | RV GRS | ≥ 13.4 | 11.0–13.3 | < 11.0 | 86 | ≥ 13.3 | 10.9–13.2 | < 10.9 | 101 |
| **30–39** | LV GLS | ≥ 15.4 | 14.2–15.3 | < 14.2 | 108 | ≥ 14.0 | 12.7–13.9 | < 12.7 | 110 |
| | LV GCS | ≥ 14.9 | 13.8–14.8 | < 13.8 | 122 | ≥ 13.8 | 12.7–13.7 | < 12.7 | 120 |
| | LV GRS | ≥ 22.0 | 19.8–21.9 | < 19.8 | 122 | ≥ 19.8 | 17.7–19.7 | < 17.7 | 120 |
| | RV GLS | ≥ 19.2 | 16.8–19.1 | < 16.8 | 109 | ≥ 17.4 | 14.9–17.3 | < 14.9 | 120 |
| | RV GCS | ≥ 9.2 | 7.6–9.1 | < 7.6 | 125 | ≥ 9.0 | 7.2–8.9 | < 7.2 | 120 |
| | RV GRS | ≥ 13.9 | 11.5–13.8 | < 11.5 | 125 | ≥ 13.5 | 10.9–13.4 | < 10.9 | 120 |
| **40–49** | LV GLS | ≥ 15.3 | 14.1–15.2 | < 14.1 | 484 | ≥ 13.9 | 12.6–13.8 | < 12.6 | 402 |
| | LV GCS | ≥ 14.9 | 13.6–14.8 | < 13.6 | 472 | ≥ 13.8 | 12.6–13.7 | < 12.6 | 406 |
| | LV GRS | ≥ 21.9 | 19.5–21.8 | < 19.5 | 472 | ≥ 19.8 | 17.5–19.7 | < 17.5 | 406 |
| | RV GLS | ≥ 19.8 | 17.5–19.7 | < 17.5 | 493 | ≥ 17.8 | 15.4–17.7 | < 15.4 | 412 |
| | RV GCS | ≥ 9.2 | 7.6–9.1 | < 7.6 | 499 | ≥ 9.3 | 7.4–9.2 | < 7.4 | 427 |
| | RV GRS | ≥ 13.4 | 11.1–13.3 | < 11.1 | 499 | ≥ 13.7 | 11.1–13.6 | < 11.1 | 427 |
| **50–59** | LV GLS | ≥ 15.1 | 13.9–15.0 | < 13.9 | 844 | ≥ 13.7 | 12.4–13.6 | < 12.4 | 733 |
| | LV GCS | ≥ 14.9 | 13.5–14.8 | < 13.5 | 833 | ≥ 13.5 | 12.1–13.4 | < 12.1 | 759 |
| | LV GRS | ≥ 21.9 | 19.2–21.8 | < 19.2 | 832 | ≥ 19.3 | 16.8–19.2 | < 16.8 | 759 |
| | RV GLS | ≥ 20.1 | 17.8–20.0 | < 17.8 | 876 | ≥ 18.0 | 15.6–17.9 | < 15.6 | 777 |
| | RV GCS | ≥ 9.4 | 7.8–9.3 | < 7.8 | 882 | ≥ 9.8 | 7.9–9.7 | < 7.9 | 817 |
| | RV GRS | ≥ 13.6 | 11.1–13.5 | < 11.1 | 881 | ≥ 14.3 | 11.6–14.2 | < 11.6 | 817 |
| **60–69** | LV GLS | ≥ 14.8 | 13.6–14.7 | < 13.6 | 859 | ≥ 13.6 | 12.3–13.5 | < 12.3 | 717 |
| | LV GCS | ≥ 15.0 | 13.4–14.9 | < 13.4 | 858 | ≥ 13.6 | 12.1–13.5 | < 12.1 | 731 |
| | LV GRS | ≥ 21.9 | 18.9–21.8 | < 18.9 | 859 | ≥ 19.4 | 16.7–19.3 | < 16.7 | 731 |
| | RV GLS | ≥ 19.9 | 17.3–19.8 | < 17.3 | 875 | ≥ 18.0 | 15.4–17.9 | < 15.4 | 751 |
| | RV GCS | ≥ 9.8 | 8.0–9.7 | < 8.0 | 893 | ≥ 10.2 | 8.3–10.1 | < 8.3 | 786 |
| | RV GRS | ≥ 14.2 | 11.6–14.1 | < 11.6 | 893 | ≥ 15.1 | 12.2–15.0 | < 12.2 | 786 |
| **70+** | LV GLS | ≥ 14.5 | 13.2–14.4 | < 13.2 | 1006 | ≥ 13.6 | 12.3–13.5 | < 12.3 | 1087 |
| | LV GCS | ≥ 15.1 | 13.4–15.0 | < 13.4 | 1025 | ≥ 13.7 | 12.1–13.6 | < 12.1 | 1136 |
| | LV GRS | ≥ 22.1 | 18.6–22.0 | < 18.6 | 1025 | ≥ 19.5 | 16.5–19.4 | < 16.5 | 1137 |
| | RV GLS | ≥ 19.2 | 16.0–19.1 | < 16.0 | 1053 | ≥ 17.7 | 14.8–17.6 | < 14.8 | 1157 |
| | RV GCS | ≥ 9.8 | 7.9–9.7 | < 7.9 | 1047 | ≥ 10.4 | 8.5–10.3 | < 8.5 | 1209 |
| | RV GRS | ≥ 14.1 | 11.2–14.0 | < 11.2 | 1047 | ≥ 15.4 | 12.4–15.3 | < 12.4 | 1209 |

**Footnote:** All strain measurements were derived using cvi42 software (Circle Cardiovascular Imaging Inc.) with standardized feature-tracking analysis. Global longitudinal strain (GLS), global circumferential strain (GCS), and global radial strain (GRS) are presented as absolute values (%). Reference ranges are defined as the interval between the 2.5th and 97.5th centiles derived from age- and sex-specific generalized additive models for location, scale and shape (GAMLSS) using the Box–Cox Cole–Green distribution. Centile-based categories beyond the reference range are derived from LMS-based thresholds (P0.5 and P2.5) and describe the position of a measurement within the reference distribution rather than disease severity or diagnostic abnormality. For clarity, only the lower reference threshold (2.5th percentile) and corresponding lower centile cut-offs are presented in this table. Reference values are derived from a White adult population.

**Abbreviations**: LV = left ventricle; RV = right ventricle; GLS = global longitudinal strain; GCS = global circumferential strain; GRS = global radial strain.

### Supplementary Table 7 Left and right ventricular global strain rate reference ranges and centile-based categories stratified by sex and age using cvi42 (Circle Cardiovascular Imaging Inc.), *in a White adult population*

| | | **Female** | | | | **Male** | | | |
|---|---|---|---|---|---|---|---|---|---|
| **Age group** | Metrics ($s^{-1}$) | Reference threshold | Mild | At least moderate | n | Reference threshold | Mild | At least moderate | n |
| **18–29** | LV sysGLSR | ≥ 0.71 | 0.66–0.71 | < 0.66 | 62 | ≥ 0.63 | 0.59–0.63 | < 0.59 | 73 |
| | LV sysGCSR | ≥ 0.74 | 0.68–0.74 | < 0.68 | 90 | ≥ 0.71 | 0.63–0.71 | < 0.63 | 101 |
| | LV sysGRSR | ≥ 1.15 | 1.02–1.15 | < 1.02 | 90 | ≥ 1.04 | 0.90–1.04 | < 0.90 | 101 |
| | LV diaGLSR | ≥ 0.56 | 0.44–0.56 | < 0.44 | 63 | ≥ 0.40 | 0.26–0.40 | < 0.26 | 75 |
| | LV diaGCSR | ≥ 0.52 | 0.42–0.52 | < 0.42 | 90 | ≥ 0.45 | 0.36–0.45 | < 0.36 | 98 |
| | LV diaGRSR | ≥ 0.94 | 0.77–0.94 | < 0.77 | 89 | ≥ 0.81 | 0.68–0.81 | < 0.68 | 97 |
| | RV sysGLSR | ≥ 0.86 | 0.75–0.86 | < 0.75 | 67 | ≥ 0.82 | 0.75–0.82 | < 0.75 | 73 |
| | RV sysGCSR | ≥ 0.42 | 0.34–0.42 | < 0.34 | 85 | ≥ 0.42 | 0.33–0.42 | < 0.33 | 101 |
| | RV sysGRSR | ≥ 0.66 | 0.52–0.66 | < 0.52 | 86 | ≥ 0.65 | 0.49–0.65 | < 0.49 | 100 |
| | RV diaGLSR | ≥ 0.80 | 0.69–0.80 | < 0.69 | 73 | ≥ 0.70 | 0.59–0.70 | < 0.59 | 80 |
| | RV diaGCSR | ≥ 0.41 | 0.32–0.41 | < 0.32 | 83 | ≥ 0.43 | 0.36–0.43 | < 0.36 | 86 |
| | RV diaGRSR | ≥ 0.71 | 0.56–0.71 | < 0.56 | 82 | ≥ 0.71 | 0.58–0.71 | < 0.58 | 86 |
| **30–39** | LV sysGLSR | ≥ 0.72 | 0.67–0.72 | < 0.67 | 103 | ≥ 0.64 | 0.60–0.64 | < 0.60 | 105 |
| | LV sysGCSR | ≥ 0.75 | 0.68–0.75 | < 0.68 | 122 | ≥ 0.73 | 0.66–0.73 | < 0.66 | 119 |
| | LV sysGRSR | ≥ 1.13 | 1.00–1.13 | < 1.00 | 122 | ≥ 1.12 | 1.00–1.12 | < 1.00 | 120 |
| | LV diaGLSR | ≥ 0.65 | 0.55–0.65 | < 0.55 | 107 | ≥ 0.53 | 0.45–0.53 | < 0.45 | 109 |
| | LV diaGCSR | ≥ 0.57 | 0.48–0.57 | < 0.48 | 121 | ≥ 0.49 | 0.41–0.49 | < 0.41 | 119 |
| | LV diaGRSR | ≥ 0.96 | 0.79–0.96 | < 0.79 | 120 | ≥ 0.86 | 0.74–0.86 | < 0.74 | 118 |
| | RV sysGLSR | ≥ 0.91 | 0.81–0.91 | < 0.81 | 100 | ≥ 0.83 | 0.76–0.83 | < 0.76 | 113 |
| | RV sysGCSR | ≥ 0.45 | 0.37–0.45 | < 0.37 | 125 | ≥ 0.47 | 0.38–0.47 | < 0.38 | 119 |
| | RV sysGRSR | ≥ 0.67 | 0.54–0.67 | < 0.54 | 125 | ≥ 0.69 | 0.54–0.69 | < 0.54 | 120 |
| | RV diaGLSR | ≥ 0.83 | 0.71–0.83 | < 0.71 | 109 | ≥ 0.72 | 0.61–0.72 | < 0.61 | 118 |
| | RV diaGCSR | ≥ 0.44 | 0.36–0.44 | < 0.36 | 122 | ≥ 0.44 | 0.38–0.44 | < 0.38 | 109 |
| | RV diaGRSR | ≥ 0.74 | 0.60–0.74 | < 0.60 | 119 | ≥ 0.74 | 0.61–0.74 | < 0.61 | 106 |
| **40–49** | LV sysGLSR | ≥ 0.70 | 0.65–0.70 | < 0.65 | 478 | ≥ 0.63 | 0.58–0.63 | < 0.58 | 391 |
| | LV sysGCSR | ≥ 0.73 | 0.67–0.73 | < 0.67 | 471 | ≥ 0.73 | 0.67–0.73 | < 0.67 | 406 |
| | LV sysGRSR | ≥ 1.10 | 0.97–1.10 | < 0.97 | 472 | ≥ 1.11 | 0.99–1.11 | < 0.99 | 405 |
| | LV diaGLSR | ≥ 0.68 | 0.59–0.68 | < 0.59 | 481 | ≥ 0.58 | 0.52–0.58 | < 0.52 | 402 |
| | LV diaGCSR | ≥ 0.61 | 0.52–0.61 | < 0.52 | 471 | ≥ 0.51 | 0.44–0.51 | < 0.44 | 403 |
| | LV diaGRSR | ≥ 0.97 | 0.80–0.97 | < 0.80 | 471 | ≥ 0.82 | 0.70–0.82 | < 0.70 | 397 |
| | RV sysGLSR | ≥ 0.89 | 0.81–0.89 | < 0.81 | 451 | ≥ 0.84 | 0.76–0.84 | < 0.76 | 390 |
| | RV sysGCSR | ≥ 0.46 | 0.39–0.46 | < 0.39 | 498 | ≥ 0.51 | 0.42–0.51 | < 0.42 | 424 |
| | RV sysGRSR | ≥ 0.67 | 0.55–0.67 | < 0.55 | 498 | ≥ 0.73 | 0.58–0.73 | < 0.58 | 426 |
| | RV diaGLSR | ≥ 0.86 | 0.73–0.86 | < 0.73 | 492 | ≥ 0.74 | 0.63–0.74 | < 0.63 | 411 |
| | RV diaGCSR | ≥ 0.46 | 0.38–0.46 | < 0.38 | 489 | ≥ 0.46 | 0.39–0.46 | < 0.39 | 411 |
| | RV diaGRSR | ≥ 0.77 | 0.63–0.77 | < 0.63 | 486 | ≥ 0.76 | 0.62–0.76 | < 0.62 | 398 |
| **50–59** | LV sysGLSR | ≥ 0.66 | 0.62–0.66 | < 0.62 | 838 | ≥ 0.62 | 0.57–0.62 | < 0.57 | 727 |
| | LV sysGCSR | ≥ 0.71 | 0.64–0.71 | < 0.64 | 833 | ≥ 0.72 | 0.66–0.72 | < 0.66 | 757 |
| | LV sysGRSR | ≥ 1.07 | 0.94–1.07 | < 0.94 | 833 | ≥ 1.04 | 0.90–1.04 | < 0.90 | 759 |
| | LV diaGLSR | ≥ 0.65 | 0.57–0.65 | < 0.57 | 844 | ≥ 0.57 | 0.51–0.57 | < 0.51 | 732 |
| | LV diaGCSR | ≥ 0.60 | 0.52–0.60 | < 0.52 | 832 | ≥ 0.52 | 0.45–0.52 | < 0.45 | 753 |
| | LV diaGRSR | ≥ 0.93 | 0.77–0.93 | < 0.77 | 830 | ≥ 0.74 | 0.62–0.74 | < 0.62 | 746 |
| | RV sysGLSR | ≥ 0.88 | 0.79–0.88 | < 0.79 | 838 | ≥ 0.85 | 0.77–0.85 | < 0.77 | 756 |
| | RV sysGCSR | ≥ 0.48 | 0.41–0.48 | < 0.41 | 878 | ≥ 0.54 | 0.45–0.54 | < 0.45 | 816 |
| | RV sysGRSR | ≥ 0.69 | 0.57–0.69 | < 0.57 | 880 | ≥ 0.78 | 0.62–0.78 | < 0.62 | 815 |
| | RV diaGLSR | ≥ 0.88 | 0.74–0.88 | < 0.74 | 876 | ≥ 0.76 | 0.64–0.76 | < 0.64 | 773 |
| | RV diaGCSR | ≥ 0.46 | 0.39–0.46 | < 0.39 | 862 | ≥ 0.46 | 0.40–0.46 | < 0.40 | 748 |
| | RV diaGRSR | ≥ 0.77 | 0.64–0.77 | < 0.64 | 852 | ≥ 0.77 | 0.63–0.77 | < 0.63 | 726 |
| **60–69** | LV sysGLSR | ≥ 0.65 | 0.60–0.65 | < 0.60 | 858 | ≥ 0.62 | 0.56–0.62 | < 0.56 | 713 |
| | LV sysGCSR | ≥ 0.69 | 0.63–0.69 | < 0.63 | 859 | ≥ 0.70 | 0.64–0.70 | < 0.64 | 728 |
| | LV sysGRSR | ≥ 1.04 | 0.91–1.04 | < 0.91 | 859 | ≥ 0.99 | 0.85–0.99 | < 0.85 | 731 |
| | LV diaGLSR | ≥ 0.66 | 0.58–0.66 | < 0.58 | 857 | ≥ 0.56 | 0.49–0.56 | < 0.49 | 717 |
| | LV diaGCSR | ≥ 0.58 | 0.50–0.58 | < 0.50 | 857 | ≥ 0.52 | 0.46–0.52 | < 0.46 | 730 |
| | LV diaGRSR | ≥ 0.88 | 0.72–0.88 | < 0.72 | 851 | ≥ 0.71 | 0.60–0.71 | < 0.60 | 725 |
| | RV sysGLSR | ≥ 0.89 | 0.80–0.89 | < 0.80 | 843 | ≥ 0.86 | 0.77–0.86 | < 0.77 | 742 |
| | RV sysGCSR | ≥ 0.52 | 0.44–0.52 | < 0.44 | 889 | ≥ 0.57 | 0.48–0.57 | < 0.48 | 785 |
| | RV sysGRSR | ≥ 0.74 | 0.61–0.74 | < 0.61 | 891 | ≥ 0.82 | 0.65–0.82 | < 0.65 | 786 |
| | RV diaGLSR | ≥ 0.88 | 0.72–0.88 | < 0.72 | 876 | ≥ 0.76 | 0.65–0.76 | < 0.65 | 744 |
| | RV diaGCSR | ≥ 0.46 | 0.40–0.46 | < 0.40 | 865 | ≥ 0.46 | 0.40–0.46 | < 0.40 | 725 |
| | RV diaGRSR | ≥ 0.76 | 0.63–0.76 | < 0.63 | 853 | ≥ 0.77 | 0.62–0.77 | < 0.62 | 682 |
| **70+** | LV sysGLSR | ≥ 0.63 | 0.58–0.63 | < 0.58 | 1003 | ≥ 0.60 | 0.53–0.60 | < 0.53 | 1086 |
| | LV sysGCSR | ≥ 0.67 | 0.61–0.67 | < 0.61 | 1025 | ≥ 0.66 | 0.59–0.66 | < 0.59 | 1136 |
| | LV sysGRSR | ≥ 1.01 | 0.88–1.01 | < 0.88 | 1024 | ≥ 0.95 | 0.82–0.95 | < 0.82 | 1136 |
| | LV diaGLSR | ≥ 0.65 | 0.57–0.65 | < 0.57 | 1006 | ≥ 0.54 | 0.46–0.54 | < 0.46 | 1088 |
| | LV diaGCSR | ≥ 0.58 | 0.50–0.58 | < 0.50 | 1019 | ≥ 0.51 | 0.45–0.51 | < 0.45 | 1135 |
| | LV diaGRSR | ≥ 0.83 | 0.69–0.83 | < 0.69 | 1014 | ≥ 0.69 | 0.58–0.69 | < 0.58 | 1113 |
| | RV sysGLSR | ≥ 0.90 | 0.81–0.90 | < 0.81 | 1018 | ≥ 0.85 | 0.75–0.85 | < 0.75 | 1143 |
| | RV sysGCSR | ≥ 0.55 | 0.47–0.55 | < 0.47 | 1040 | ≥ 0.57 | 0.49–0.57 | < 0.49 | 1204 |
| | RV sysGRSR | ≥ 0.77 | 0.63–0.77 | < 0.63 | 1043 | ≥ 0.82 | 0.66–0.82 | < 0.66 | 1206 |
| | RV diaGLSR | ≥ 0.85 | 0.67–0.85 | < 0.67 | 1046 | ≥ 0.74 | 0.62–0.74 | < 0.62 | 1146 |
| | RV diaGCSR | ≥ 0.46 | 0.39–0.46 | < 0.39 | 998 | ≥ 0.45 | 0.38–0.45 | < 0.38 | 1105 |
| | RV diaGRSR | ≥ 0.73 | 0.60–0.73 | < 0.60 | 988 | ≥ 0.74 | 0.59–0.74 | < 0.59 | 1036 |

**Footnote:** All strain rate measurements were derived using cvi42 software (Circle Cardiovascular Imaging Inc.) with standardized feature-tracking analysis. Systolic and diastolic global longitudinal strain rate (GLSR), global circumferential strain rate (GCSR), and global radial strain rate (GRSR) are presented in $s^{-1}$. Reference ranges are defined as the interval between the 2.5th and 97.5th centiles derived from age- and sex-specific generalized additive models for location, scale and shape (GAMLSS) using the Box–Cox Cole–Green distribution. Centile-based categories beyond the reference range are derived from LMS-based thresholds (P0.5 and P2.5) and describe the position of a measurement within the reference distribution rather than disease severity or diagnostic abnormality. For clarity, only the lower reference threshold (2.5th percentile) and corresponding lower centile cut-offs are presented in this table. Reference values are derived from a White adult population.

**Abbreviations**: LV = left ventricle; RV = right ventricle; GLSR = global longitudinal strain rate; GCSR = global circumferential strain rate; GRSR = global radial strain rate; sys = systolic; dia = diastolic.

**Supplementary Table 8 Left ventricular global displacement reference ranges and centile-based categories stratified by sex and age using cvi42 (Circle Cardiovascular Imaging Inc.), *in a White adult population***

| | | Female | | | | Male | | | |
|---|---|---|---|---|---|---|---|---|---|
| **Age group** | Metrics (mm) | Reference threshold | Mild | At least moderate | n | Reference threshold | Mild | At least moderate | n |
| **18–29** | LV GLD | ≥ 3.4 | 2.7–3.3 | < 2.7 | 64 | ≥ 2.4 | 1.7–2.3 | < 1.7 | 75 |
| | LV GCD | ≥ 0.8 | 0.5–0.7 | < 0.5 | 76 | ≥ 0.8 | 0.6–0.7 | < 0.6 | 87 |
| | LV GRD | ≥ 3.9 | 3.6–3.8 | < 3.6 | 90 | ≥ 3.9 | 3.5–3.8 | < 3.5 | 101 |
| **30–39** | LV GLD | ≥ 3.4 | 2.7–3.3 | < 2.7 | 108 | ≥ 2.7 | 2.0–2.6 | < 2.0 | 110 |
| | LV GCD | ≥ 0.7 | 0.5–0.6 | < 0.5 | 106 | ≥ 0.8 | 0.5–0.7 | < 0.5 | 114 |
| | LV GRD | ≥ 3.9 | 3.6–3.8 | < 3.6 | 122 | ≥ 3.9 | 3.5–3.8 | < 3.5 | 120 |
| **40–49** | LV GLD | ≥ 3.3 | 2.6–3.2 | < 2.6 | 484 | ≥ 2.6 | 2.0–2.5 | < 2.0 | 402 |
| | LV GCD | ≥ 0.7 | 0.5–0.6 | < 0.5 | 411 | ≥ 0.8 | 0.5–0.7 | < 0.5 | 369 |
| | LV GRD | ≥ 4.0 | 3.6–3.9 | < 3.6 | 472 | ≥ 3.9 | 3.5–3.8 | < 3.5 | 406 |
| **50–59** | LV GLD | ≥ 2.9 | 2.2–2.8 | < 2.2 | 844 | ≥ 2.3 | 1.8–2.2 | < 1.8 | 732 |
| | LV GCD | ≥ 0.7 | 0.5–0.6 | < 0.5 | 668 | ≥ 0.8 | 0.5–0.7 | < 0.5 | 708 |
| | LV GRD | ≥ 4.0 | 3.6–3.9 | < 3.6 | 833 | ≥ 3.8 | 3.4–3.7 | < 3.4 | 759 |
| **60–69** | LV GLD | ≥ 2.6 | 2.0–2.5 | < 2.0 | 859 | ≥ 2.3 | 1.7–2.2 | < 1.7 | 718 |
| | LV GCD | ≥ 0.7 | 0.5–0.6 | < 0.5 | 658 | ≥ 0.8 | 0.5–0.7 | < 0.5 | 650 |
| | LV GRD | ≥ 3.9 | 3.5–3.8 | < 3.5 | 859 | ≥ 3.8 | 3.3–3.7 | < 3.3 | 731 |
| **70+** | LV GLD | ≥ 2.4 | 1.8–2.3 | < 1.8 | 1005 | ≥ 2.3 | 1.7–2.2 | < 1.7 | 1089 |
| | LV GCD | ≥ 0.7 | 0.5–0.6 | < 0.5 | 730 | ≥ 0.8 | 0.5–0.7 | < 0.5 | 987 |
| | LV GRD | ≥ 3.9 | 3.4–3.8 | < 3.4 | 1025 | ≥ 3.7 | 3.2–3.6 | < 3.2 | 1137 |

**Footnote:** All displacement measurements were derived using cvi42 software (Circle Cardiovascular Imaging Inc.) with standardized feature-tracking analysis. Global longitudinal displacement (GLD), global circumferential displacement (GCD), and global radial displacement (GRD) are presented in millimetres (mm). Reference ranges are defined as the interval between the 2.5th and 97.5th centiles derived from age- and sex-specific generalized additive models for location, scale and shape (GAMLSS) using the Box–Cox Cole–Green distribution. Centile-based categories beyond the reference range are derived from LMS-based thresholds (P0.5 and P2.5) and describe the position of a measurement within the reference distribution rather than disease severity or diagnostic abnormality. For clarity, only the lower reference threshold (2.5th percentile) and corresponding lower centile cut-offs are presented in this table. Reference values are derived from a White adult population.
**Abbreviations**: LV = left ventricle; GLD = global longitudinal displacement; GCD = global circumferential displacement; GRD = global radial displacement.

### Supplementary Table 9 Left ventricular global velocity reference ranges and centile-based categories stratified by sex and age using cvi42 (Circle Cardiovascular Imaging Inc.), *in a White adult population*

| | | Female | | | | Male | | | |
|---|---|---|---|---|---|---|---|---|---|
| **Age group** | Metrics (%/s) | Reference threshold | Mild | At least moderate | n | Reference threshold | Mild | At least moderate | n |
| **18–29** | LV sysGLV | ≥ 16.1 | 12.7–16.0 | < 12.7 | 62 | ≥ 13.4 | 10.0–13.3 | < 10.0 | 72 |
| | LV sysGCV | ≥ 10.1 | 6.7–10.0 | < 6.7 | 76 | ≥ 11.8 | 8.0–11.7 | < 8.0 | 93 |
| | LV sysGRV | ≥ 20.8 | 18.5–20.7 | < 18.5 | 89 | ≥ 21.2 | 18.3–21.1 | < 18.3 | 101 |
| | LV diaGLV | ≥ 13.5 | 6.4–13.4 | < 6.4 | 61 | ≥ 10.7 | 5.6–10.6 | < 5.6 | 74 |
| | LV diaGCV | ≥ 4.9 | 2.5–4.8 | < 2.5 | 61 | ≥ 4.8 | 3.5–4.7 | < 3.5 | 64 |
| | LV diaGRV | ≥ 14.7 | 11.8–14.6 | < 11.8 | 90 | ≥ 14.7 | 11.5–14.6 | < 11.5 | 99 |
| **30–39** | LV sysGLV | ≥ 19.2 | 16.0–19.1 | < 16.0 | 102 | ≥ 15.2 | 11.1–15.1 | < 11.1 | 103 |
| | LV sysGCV | ≥ 11.5 | 8.5–11.4 | < 8.5 | 109 | ≥ 11.8 | 8.2–11.7 | < 8.2 | 113 |
| | LV sysGRV | ≥ 21.2 | 19.2–21.1 | < 19.2 | 122 | ≥ 22.6 | 20.4–22.5 | < 20.4 | 119 |
| | LV diaGLV | ≥ 16.5 | 8.9–16.4 | < 8.9 | 103 | ≥ 14.6 | 10.3–14.5 | < 10.3 | 110 |
| | LV diaGCV | ≥ 5.9 | 3.9–5.8 | < 3.9 | 77 | ≥ 4.9 | 3.6–4.8 | < 3.6 | 86 |
| | LV diaGRV | ≥ 16.3 | 13.6–16.2 | < 13.6 | 121 | ≥ 15.7 | 13.1–15.6 | < 13.1 | 119 |
| **40–49** | LV sysGLV | ≥ 18.1 | 15.5–18.0 | < 15.5 | 473 | ≥ 15.3 | 11.9–15.2 | < 11.9 | 385 |
| | LV sysGCV | ≥ 10.4 | 7.6–10.3 | < 7.6 | 420 | ≥ 11.8 | 8.3–11.7 | < 8.3 | 385 |
| | LV sysGRV | ≥ 20.9 | 19.0–20.8 | < 19.0 | 470 | ≥ 22.8 | 20.9–22.7 | < 20.9 | 405 |
| | LV diaGLV | ≥ 18.2 | 13.5–18.1 | < 13.5 | 479 | ≥ 15.3 | 11.9–15.2 | < 11.9 | 401 |
| | LV diaGCV | ≥ 6.2 | 4.5–6.1 | < 4.5 | 319 | ≥ 5.0 | 3.7–4.9 | < 3.7 | 328 |
| | LV diaGRV | ≥ 17.7 | 15.2–17.6 | < 15.2 | 471 | ≥ 15.9 | 13.6–15.8 | < 13.6 | 402 |
| **50–59** | LV sysGLV | ≥ 15.1 | 13.1–15.0 | < 13.1 | 838 | ≥ 14.7 | 12.2–14.6 | < 12.2 | 721 |
| | LV sysGCV | ≥ 9.7 | 7.1–9.6 | < 7.1 | 692 | ≥ 11.6 | 8.3–11.5 | < 8.3 | 714 |
| | LV sysGRV | ≥ 19.9 | 18.3–19.8 | < 18.3 | 832 | ≥ 22.2 | 20.4–22.1 | < 20.4 | 756 |
| | LV diaGLV | ≥ 16.6 | 13.6–16.5 | < 13.6 | 841 | ≥ 14.0 | 11.4–13.9 | < 11.4 | 730 |
| | LV diaGCV | ≥ 5.8 | 4.3–5.7 | < 4.3 | 560 | ≥ 5.1 | 3.8–5.0 | < 3.8 | 598 |
| | LV diaGRV | ≥ 17.1 | 14.9–17.0 | < 14.9 | 831 | ≥ 15.8 | 13.7–15.7 | < 13.7 | 751 |
| **60–69** | LV sysGLV | ≥ 14.6 | 12.6–14.5 | < 12.6 | 854 | ≥ 15.2 | 13.0–15.1 | < 13.0 | 714 |
| | LV sysGCV | ≥ 9.5 | 7.1–9.4 | < 7.1 | 659 | ≥ 11.2 | 8.2–11.1 | < 8.2 | 652 |
| | LV sysGRV | ≥ 19.2 | 17.6–19.1 | < 17.6 | 859 | ≥ 20.9 | 19.0–20.8 | < 19.0 | 729 |
| | LV diaGLV | ≥ 16.1 | 13.6–16.0 | < 13.6 | 855 | ≥ 14.3 | 11.7–14.2 | < 11.7 | 711 |
| | LV diaGCV | ≥ 5.3 | 4.0–5.2 | < 4.0 | 560 | ≥ 5.2 | 3.9–5.1 | < 3.9 | 571 |
| | LV diaGRV | ≥ 16.2 | 14.2–16.1 | < 14.2 | 856 | ≥ 15.7 | 13.7–15.6 | < 13.7 | 730 |
| **70+** | LV sysGLV | ≥ 13.9 | 11.7–13.8 | < 11.7 | 1001 | ≥ 15.1 | 12.9–15.0 | < 12.9 | 1086 |
| | LV sysGCV | ≥ 8.9 | 6.8–8.8 | < 6.8 | 673 | ≥ 10.5 | 7.8–10.4 | < 7.8 | 912 |
| | LV sysGRV | ≥ 18.5 | 16.9–18.4 | < 16.9 | 1025 | ≥ 19.3 | 17.5–19.2 | < 17.5 | 1136 |
| | LV diaGLV | ≥ 16.0 | 13.2–15.9 | < 13.2 | 1000 | ≥ 14.1 | 11.1–14.0 | < 11.1 | 1082 |
| | LV diaGCV | ≥ 5.2 | 3.8–5.1 | < 3.8 | 634 | ≥ 5.3 | 4.0–5.2 | < 4.0 | 908 |
| | LV diaGRV | ≥ 16.1 | 14.2–16.0 | < 14.2 | 1021 | ≥ 15.2 | 13.4–15.1 | < 13.4 | 1131 |

**Footnote:** All velocity measurements were derived using cvi42 software (Circle Cardiovascular Imaging Inc.) with standardized feature-tracking analysis. Global longitudinal velocity (GLV), global circumferential velocity (GCV), and global radial velocity (GRV) are presented in %/s. Systolic and diastolic measurements are indicated by the prefixes sys and dia, respectively. Reference ranges are defined as the interval between the 2.5th and 97.5th centiles derived from age- and sex-specific generalized additive models for location, scale and shape (GAMLSS) using the Box–Cox Cole–Green distribution. Centile-based categories beyond the reference range are derived from LMS-based thresholds (P0.5 and P2.5) and describe the position of a measurement within the reference distribution rather than disease severity or diagnostic abnormality. For clarity, only the lower reference threshold (2.5th percentile) and corresponding lower centile cut-offs are presented in this table. Reference values are derived from a White adult population.

**Abbreviations**: LV = left ventricle; GLV = global longitudinal velocity; GCV = global circumferential velocity; GRV = global radial velocity; sys = systolic; dia = diastolic.

**Supplementary Table 10 Left and right ventricular global strain reference ranges and centile-based categories stratified by sex and age using cvi42 (Circle Cardiovascular Imaging Inc.), *in a Black adult population***

| | | Female | | | | Male | | | |
|---|---|---|---|---|---|---|---|---|---|
| **Age group** | Metrics (%) | Reference threshold | Mild | At least moderate | n | Reference threshold | Mild | At least moderate | n |
| **40–49** | LV GLS | ≥ 14.9 | 14.1–14.8 | < 14.1 | 10 | | | | |
| | LV GCS | ≥ 15.2 | 14.5–15.1 | < 14.5 | 10 | | | | |
| | LV GRS | ≥ 22.5 | 21.0–22.4 | < 21.0 | 10 | | | | |
| | RV GLS | ≥ 17.6 | 15.7–17.5 | < 15.7 | 10 | | | | |
| | RV GCS | | | | | | | | |
| | RV GRS | | | | | | | | |
| **50–59** | LV GLS | ≥ 14.4 | 13.4–14.3 | < 13.4 | 89 | ≥ 13.0 | 11.7–12.9 | < 11.7 | 62 |
| | LV GCS | ≥ 15.1 | 14.2–15.0 | < 14.2 | 87 | ≥ 13.6 | 12.5–13.5 | < 12.5 | 67 |
| | LV GRS | ≥ 22.3 | 20.5–22.2 | < 20.5 | 87 | ≥ 19.4 | 17.3–19.3 | < 17.3 | 67 |
| | RV GLS | ≥ 17.6 | 15.7–17.5 | < 15.7 | 88 | ≥ 15.2 | 12.5–15.1 | < 12.5 | 72 |
| | RV GCS | ≥ 9.2 | 8.0–9.1 | < 8.0 | 90 | ≥ 7.9 | 5.7–7.8 | < 5.7 | 76 |
| | RV GRS | ≥ 12.9 | 11.2–12.8 | < 11.2 | 90 | ≥ 11.2 | 8.4–11.1 | < 8.4 | 76 |
| **60–69** | LV GLS | ≥ 13.4 | 12.1–13.3 | < 12.1 | 59 | ≥ 12.3 | 10.3–12.2 | < 10.3 | 40 |
| | LV GCS | ≥ 14.6 | 13.2–14.5 | < 13.2 | 64 | ≥ 10.9 | 7.6–10.8 | < 7.6 | 45 |
| | LV GRS | ≥ 21.5 | 18.9–21.4 | < 18.9 | 64 | ≥ 13.0 | 7.5–12.9 | < 7.5 | 45 |
| | RV GLS | ≥ 17.7 | 15.8–17.6 | < 15.8 | 60 | ≥ 14.6 | 11.5–14.5 | < 11.5 | 44 |
| | RV GCS | ≥ 9.0 | 7.8–8.9 | < 7.8 | 61 | ≥ 8.4 | 6.5–8.3 | < 6.5 | 46 |
| | RV GRS | ≥ 12.7 | 10.8–12.6 | < 10.8 | 61 | ≥ 12.2 | 9.7–12.1 | < 9.7 | 46 |
| **70+** | LV GLS | ≥ 12.2 | 10.4–12.1 | < 10.4 | 15 | ≥ 11.4 | 8.6–11.3 | < 8.6 | 12 |
| | LV GCS | ≥ 13.5 | 10.7–13.4 | < 10.7 | 16 | ≥ 12.7 | 10.1–12.6 | < 10.1 | 12 |
| | LV GRS | ≥ 19.6 | 15.0–19.5 | < 15.0 | 16 | ≥ 15.8 | 10.5–15.7 | < 10.5 | 12 |
| | RV GLS | ≥ 17.8 | 15.9–17.7 | < 15.9 | 16 | ≥ 14.5 | 11.0–14.4 | < 11.0 | 10 |
| | RV GCS | ≥ 8.8 | 7.5–8.7 | < 7.5 | 15 | ≥ 8.8 | 7.2–8.7 | < 7.2 | 14 |
| | RV GRS | ≥ 12.4 | 10.4–12.3 | < 10.4 | 15 | ≥ 13.4 | 11.1–13.3 | < 11.1 | 14 |

**Footnote:** All strain measurements were derived using cvi42 software (Circle Cardiovascular Imaging Inc.) with standardized feature-tracking analysis. Global longitudinal strain (GLS), global circumferential strain (GCS), and global radial strain (GRS) are presented as absolute values (%). Reference ranges are defined as the interval between the 2.5th and 97.5th centiles derived from age- and sex-specific generalized additive models for location, scale and shape (GAMLSS) using the Box–Cox Cole–Green distribution. Centile-based categories beyond the reference range are derived from LMS-based thresholds (P0.5 and P2.5) and describe the position of a measurement within the reference distribution rather than disease severity or diagnostic abnormality. For clarity, only the lower reference threshold (2.5th percentile) and corresponding lower centile cut-offs are presented in this table. Reference values are derived from a Black adult population.
**Abbreviations**: LV = left ventricle; RV = right ventricle; GLS = global longitudinal strain; GCS = global circumferential strain; GRS = global radial strain.

**Supplementary Table 11 Left and right ventricular global strain rate reference ranges and centile-based categories stratified by sex and age using cvi42 (Circle Cardiovascular Imaging Inc.),** ***in a Black adult population***

| | | Female | | | | Male | | | |
|---|---|---|---|---|---|---|---|---|---|
| **Age group** | Metrics (s⁻¹) | Reference threshold | Mild | At least moderate | n | Reference threshold | Mild | At least moderate | n |
| **40–49** | LV sysGLSR | ≥ 0.61 | 0.56–0.61 | < 0.56 | 10 | | | | |
| | LV sysGCSR | ≥ 0.74 | 0.68–0.74 | < 0.68 | 10 | | | | |
| | LV sysGRSR | ≥ 1.14 | 1.06–1.14 | < 1.06 | 10 | | | | |
| | LV diaGLSR | ≥ 0.61 | 0.56–0.61 | < 0.56 | 10 | | | | |
| | LV diaGCSR | ≥ 0.60 | 0.53–0.60 | < 0.53 | 10 | | | | |
| | LV diaGRSR | ≥ 1.02 | 0.91–1.02 | < 0.91 | 10 | | | | |
| | RV sysGLSR | ≥ 0.76 | 0.68–0.76 | < 0.68 | 10 | | | | |
| | RV sysGCSR | | | | | | | | |
| | RV sysGRSR | | | | | | | | |
| | RV diaGLSR | ≥ 0.85 | 0.76–0.85 | < 0.76 | 10 | | | | |
| | RV diaGCSR | | | | | | | | |
| | RV diaGRSR | | | | | | | | |
| **50–59** | LV sysGLSR | ≥ 0.61 | 0.55–0.61 | < 0.55 | 89 | ≥ 0.61 | 0.56–0.61 | < 0.56 | 62 |
| | LV sysGCSR | ≥ 0.72 | 0.66–0.72 | < 0.66 | 87 | ≥ 0.70 | 0.63–0.70 | < 0.63 | 67 |
| | LV sysGRSR | ≥ 1.11 | 1.02–1.11 | < 1.02 | 87 | ≥ 0.99 | 0.85–0.99 | < 0.85 | 67 |
| | LV diaGLSR | ≥ 0.62 | 0.57–0.62 | < 0.57 | 89 | ≥ 0.57 | 0.50–0.57 | < 0.50 | 61 |
| | LV diaGCSR | ≥ 0.59 | 0.53–0.59 | < 0.53 | 86 | ≥ 0.49 | 0.41–0.49 | < 0.41 | 67 |
| | LV diaGRSR | ≥ 0.96 | 0.84–0.96 | < 0.84 | 87 | ≥ 0.75 | 0.62–0.75 | < 0.62 | 66 |
| | RV sysGLSR | ≥ 0.75 | 0.67–0.75 | < 0.67 | 86 | ≥ 0.75 | 0.67–0.75 | < 0.67 | 68 |
| | RV sysGCSR | ≥ 0.46 | 0.39–0.46 | < 0.39 | 89 | ≥ 0.45 | 0.36–0.45 | < 0.36 | 76 |
| | RV sysGRSR | ≥ 0.63 | 0.52–0.63 | < 0.52 | 91 | ≥ 0.61 | 0.46–0.61 | < 0.46 | 76 |
| | RV diaGLSR | ≥ 0.82 | 0.72–0.82 | < 0.72 | 88 | ≥ 0.67 | 0.54–0.67 | < 0.54 | 72 |
| | RV diaGCSR | ≥ 0.45 | 0.39–0.45 | < 0.39 | 87 | ≥ 0.48 | 0.43–0.48 | < 0.43 | 71 |
| | RV diaGRSR | ≥ 0.73 | 0.63–0.73 | < 0.63 | 90 | ≥ 0.77 | 0.65–0.77 | < 0.65 | 73 |
| **60–69** | LV sysGLSR | ≥ 0.59 | 0.54–0.59 | < 0.54 | 58 | ≥ 0.56 | 0.50–0.56 | < 0.50 | 40 |
| | LV sysGCSR | ≥ 0.69 | 0.63–0.69 | < 0.63 | 64 | ≥ 0.66 | 0.58–0.66 | < 0.58 | 45 |
| | LV sysGRSR | ≥ 1.04 | 0.92–1.04 | < 0.92 | 64 | ≥ 0.93 | 0.79–0.93 | < 0.79 | 45 |
| | LV diaGLSR | ≥ 0.64 | 0.58–0.64 | < 0.58 | 59 | ≥ 0.56 | 0.51–0.56 | < 0.51 | 40 |
| | LV diaGCSR | ≥ 0.58 | 0.51–0.58 | < 0.51 | 64 | ≥ 0.51 | 0.43–0.51 | < 0.43 | 45 |
| | LV diaGRSR | ≥ 0.88 | 0.74–0.88 | < 0.74 | 64 | ≥ 0.72 | 0.59–0.72 | < 0.59 | 45 |
| | RV sysGLSR | ≥ 0.75 | 0.66–0.75 | < 0.66 | 59 | ≥ 0.71 | 0.59–0.71 | < 0.59 | 45 |
| | RV sysGCSR | ≥ 0.50 | 0.43–0.50 | < 0.43 | 61 | ≥ 0.44 | 0.33–0.44 | < 0.33 | 46 |
| | RV sysGRSR | ≥ 0.66 | 0.55–0.66 | < 0.55 | 61 | ≥ 0.61 | 0.44–0.61 | < 0.44 | 46 |
| | RV diaGLSR | ≥ 0.76 | 0.62–0.76 | < 0.62 | 60 | ≥ 0.64 | 0.49–0.64 | < 0.49 | 45 |
| | RV diaGCSR | ≥ 0.43 | 0.37–0.43 | < 0.37 | 58 | ≥ 0.42 | 0.35–0.42 | < 0.35 | 43 |
| | RV diaGRSR | ≥ 0.71 | 0.60–0.71 | < 0.60 | 56 | ≥ 0.60 | 0.43–0.60 | < 0.43 | 43 |
| **70+** | LV sysGLSR | ≥ 0.57 | 0.52–0.57 | < 0.52 | 15 | ≥ 0.52 | 0.44–0.52 | < 0.44 | 12 |
| | LV sysGCSR | ≥ 0.66 | 0.60–0.66 | < 0.60 | 16 | ≥ 0.62 | 0.55–0.62 | < 0.55 | 12 |
| | LV sysGRSR | ≥ 0.94 | 0.78–0.94 | < 0.78 | 16 | ≥ 0.89 | 0.75–0.89 | < 0.75 | 12 |
| | LV diaGLSR | ≥ 0.65 | 0.60–0.65 | < 0.60 | 15 | ≥ 0.56 | 0.51–0.56 | < 0.51 | 11 |
| | LV diaGCSR | ≥ 0.57 | 0.50–0.57 | < 0.50 | 16 | ≥ 0.53 | 0.45–0.53 | < 0.45 | 12 |
| | LV diaGRSR | ≥ 0.86 | 0.71–0.86 | < 0.71 | 16 | ≥ 0.69 | 0.57–0.69 | < 0.57 | 12 |
| | RV sysGLSR | ≥ 0.73 | 0.64–0.73 | < 0.64 | 16 | ≥ 0.71 | 0.50–0.71 | < 0.50 | 10 |
| | RV sysGCSR | ≥ 0.54 | 0.47–0.54 | < 0.47 | 15 | ≥ 0.47 | 0.37–0.47 | < 0.37 | 14 |
| | RV sysGRSR | ≥ 0.69 | 0.57–0.69 | < 0.57 | 15 | ≥ 0.61 | 0.43–0.61 | < 0.43 | 14 |
| | RV diaGLSR | ≥ 0.65 | 0.46–0.65 | < 0.46 | 16 | ≥ 0.61 | 0.42–0.61 | < 0.42 | 10 |
| | RV diaGCSR | ≥ 0.42 | 0.35–0.42 | < 0.35 | 14 | ≥ 0.33 | 0.23–0.33 | < 0.23 | 12 |
| | RV diaGRSR | ≥ 0.69 | 0.58–0.69 | < 0.58 | 14 | ≥ 0.53 | 0.37–0.53 | < 0.37 | 13 |

**Footnote:** All strain rate measurements were derived using CVI42 software (Circle Cardiovascular Imaging Inc.) with standardized feature-tracking analysis. Systolic and diastolic global longitudinal strain rate (GLSR), global circumferential strain rate (GCSR), and global radial strain rate (GRSR) are presented in $s^{-1}$. Reference ranges are defined as the interval between the 2.5th and 97.5th centiles derived from age- and sex-specific generalized additive models for location, scale and shape (GAMLSS) using the Box–Cox Cole–Green distribution. Centile-based categories beyond the reference range are derived from LMS-based thresholds (P0.5 and P2.5) and describe the position of a measurement within the reference distribution rather than disease severity or diagnostic abnormality. For clarity, only the lower reference threshold (2.5th percentile) and corresponding lower centile cut-offs are presented in this table. Reference values are derived from a Black adult population.
**Abbreviations**: LV = left ventricle; RV = right ventricle; GLSR = global longitudinal strain rate; GCSR = global circumferential strain rate; GRSR = global radial strain rate; sys = systolic; dia = diastolic.

**Supplementary Table 12 Left ventricular global displacement reference ranges and centile-based categories stratified by sex and age using cvi42 (Circle Cardiovascular Imaging Inc.), *in a Black adult population***

| | | Female | | | | Male | | | |
|---|---|---|---|---|---|---|---|---|---|
| **Age group** | Metrics (mm) | Reference threshold | Mild | At least moderate | n | Reference threshold | Mild | At least moderate | n |
| **40–49** | LV GLD | ≥ 2.5 | 1.7–2.4 | < 1.7 | 10 | | | | |
| | LV GRD | ≥ 4.1 | 3.9–4.0 | < 3.9 | 10 | | | | |
| **50–59** | LV GLD | ≥ 3.0 | 2.5–2.9 | < 2.5 | 89 | ≥ 2.8 | 2.5–2.7 | < 2.5 | 62 |
| | LV GCD | ≥ 0.7 | 0.5–0.6 | < 0.5 | 56 | ≥ 0.7 | 0.6–0.6 | < 0.6 | 55 |
| | LV GRD | ≥ 4.1 | 3.8–4.0 | < 3.8 | 87 | ≥ 3.9 | 3.6–3.8 | < 3.6 | 67 |
| **60–69** | LV GLD | ≥ 3.2 | 2.9–3.1 | < 2.9 | 59 | ≥ 2.6 | 2.1–2.5 | < 2.1 | 40 |
| | LV GCD | ≥ 0.6 | 0.5–0.5 | < 0.5 | 36 | ≥ 0.7 | 0.5–0.6 | < 0.5 | 34 |
| | LV GRD | ≥ 3.9 | 3.4–3.8 | < 3.4 | 64 | ≥ 3.9 | 3.5–3.8 | < 3.5 | 45 |
| **70+** | LV GLD | ≥ 3.3 | 3.1–3.2 | < 3.1 | 15 | ≥ 2.3 | 1.7–2.2 | < 1.7 | 12 |
| | LV GCD | | | | | ≥ 0.6 | 0.4–0.5 | < 0.4 | 10 |
| | LV GRD | ≥ 3.6 | 2.7–3.5 | < 2.7 | 16 | ≥ 3.8 | 3.4–3.7 | < 3.4 | 12 |

**Footnote:** All displacement measurements were derived using cvi42 software (Circle Cardiovascular Imaging Inc.) with standardized feature-tracking analysis. Global longitudinal displacement (GLD), global circumferential displacement (GCD), and global radial displacement (GRD) are presented in millimetres (mm). Reference ranges are defined as the interval between the 2.5th and 97.5th centiles derived from age- and sex-specific generalized additive models for location, scale and shape (GAMLSS) using the Box–Cox Cole–Green distribution. Centile-based categories beyond the reference range are derived from LMS-based thresholds (P0.5 and P2.5) and describe the position of a measurement within the reference distribution rather than disease severity or diagnostic abnormality. For clarity, only the lower reference threshold (2.5th percentile) and corresponding lower centile cut-offs are presented in this table. Reference values are derived from a Black adult population.
**Abbreviations**: LV = left ventricle; GLD = global longitudinal displacement; GCD = global circumferential displacement; GRD = global radial displacement.

**Supplementary Table 13 Left ventricular global velocity reference ranges and centile-based categories stratified by sex and age using cvi42 (Circle Cardiovascular Imaging Inc.),** ***in a Black adult population***

| | | Female | | | | Male | | | |
|---|---|---|---|---|---|---|---|---|---|
| **Age group** | Metrics (%/s) | Reference threshold | Mild | At least moderate | n | Reference threshold | Mild | At least moderate | n |
| **40–49** | LV sysGLV | ≥ 14.3 | 11.9–14.2 | < 11.9 | 10 | | | | |
| | LV sysGCV | | | | | | | | |
| | LV sysGRV | ≥ 21.1 | 19.6–21.0 | < 19.6 | 10 | | | | |
| | LV diaGLV | ≥ 16.0 | 12.8–15.9 | < 12.8 | 10 | | | | |
| | LV diaGCV | | | | | | | | |
| | LV diaGRV | ≥ 17.1 | 15.0–17.0 | < 15.0 | 10 | | | | |
| **50–59** | LV sysGLV | ≥ 14.9 | 12.9–14.8 | < 12.9 | 89 | ≥ 14.3 | 12.6–14.2 | < 12.6 | 62 |
| | LV sysGCV | ≥ 9.9 | 7.5–9.8 | < 7.5 | 65 | ≥ 11.4 | 8.3–11.3 | < 8.3 | 56 |
| | LV sysGRV | ≥ 20.4 | 18.8–20.3 | < 18.8 | 87 | ≥ 21.5 | 19.8–21.4 | < 19.8 | 67 |
| | LV diaGLV | ≥ 17.1 | 14.7–17.0 | < 14.7 | 87 | ≥ 17.0 | 15.3–16.9 | < 15.3 | 62 |
| | LV diaGCV | ≥ 4.8 | 2.7–4.7 | < 2.7 | 46 | ≥ 5.9 | 5.1–5.8 | < 5.1 | 47 |
| | LV diaGRV | ≥ 16.9 | 14.9–16.8 | < 14.9 | 86 | ≥ 16.0 | 14.3–15.9 | < 14.3 | 67 |
| **60–69** | LV sysGLV | ≥ 15.3 | 13.8–15.2 | < 13.8 | 59 | ≥ 14.3 | 12.5–14.2 | < 12.5 | 40 |
| | LV sysGCV | ≥ 9.1 | 6.9–9.0 | < 6.9 | 39 | ≥ 8.5 | 4.3–8.4 | < 4.3 | 34 |
| | LV sysGRV | ≥ 19.3 | 17.7–19.2 | < 17.7 | 64 | ≥ 19.6 | 17.6–19.5 | < 17.6 | 45 |
| | LV diaGLV | ≥ 18.1 | 16.3–18.0 | < 16.3 | 59 | ≥ 15.9 | 13.7–15.8 | < 13.7 | 40 |
| | LV diaGCV | ≥ 4.5 | 2.6–4.4 | < 2.6 | 28 | ≥ 5.8 | 4.9–5.7 | < 4.9 | 23 |
| | LV diaGRV | ≥ 16.5 | 14.7–16.4 | < 14.7 | 64 | ≥ 16.1 | 14.6–16.0 | < 14.6 | 45 |
| **70+** | LV sysGLV | ≥ 15.3 | 14.1–15.2 | < 14.1 | 15 | ≥ 14.3 | 12.3–14.2 | < 12.3 | 11 |
| | LV sysGCV | | | | | | | | |
| | LV sysGRV | ≥ 18.3 | 16.6–18.2 | < 16.6 | 16 | ≥ 17.8 | 15.3–17.7 | < 15.3 | 12 |
| | LV diaGLV | ≥ 18.6 | 17.2–18.5 | < 17.2 | 15 | ≥ 14.5 | 11.5–14.4 | < 11.5 | 11 |
| | LV diaGCV | | | | | | | | |
| | LV diaGRV | ≥ 16.1 | 14.5–16.0 | < 14.5 | 16 | ≥ 16.1 | 14.8–16.0 | < 14.8 | 12 |

**Footnote**: All velocity measurements were derived using cvi42 software (Circle Cardiovascular Imaging Inc.) with standardized feature-tracking analysis. Systolic and diastolic global longitudinal velocity (GLV), global circumferential velocity (GCV), and global radial velocity (GRV) are presented in %/s. Reference ranges are defined as the interval between the 2.5th and 97.5th centiles derived from age- and sex-specific generalized additive models for location, scale and shape (GAMLSS) using the Box–Cox Cole–Green distribution. Centile-based categories beyond the reference range are derived from LMS-based thresholds (P0.5 and P2.5) and describe the position of a measurement within the reference distribution rather than disease severity or diagnostic abnormality. For clarity, only the lower reference threshold (2.5th percentile) and corresponding lower centile cut-offs are presented in this table. Reference values are derived from a Black adult population.
**Abbreviations**: LV = left ventricle; GLV = global longitudinal velocity; GCV = global circumferential velocity; GRV = global radial velocity; sys = systolic; dia = diastolic.

**Supplementary Table 14 Left and right ventricular global strain reference ranges and centile-based categories stratified by sex and age using cvi42 (Circle Cardiovascular Imaging Inc.), *in a South Asian adult population***

| | | Female | | | | Male | | | |
|---|---|---|---|---|---|---|---|---|---|
| **Age group** | Metrics (%) | Reference threshold | Mild | At least moderate | n | Reference threshold | Mild | At least moderate | n |
| **40–49** | LV GLS | | | | | ≥ 14.7 | 13.4–14.6 | < 13.4 | 19 |
| | LV GCS | | | | | ≥ 14.2 | 13.2–14.1 | < 13.2 | 20 |
| | LV GRS | | | | | ≥ 20.4 | 18.4–20.3 | < 18.4 | 20 |
| | RV GLS | | | | | ≥ 16.6 | 13.3–16.5 | < 13.3 | 19 |
| | RV GCS | | | | | ≥ 9.0 | 7.3–8.9 | < 7.3 | 20 |
| | RV GRS | | | | | ≥ 13.1 | 10.8–13.0 | < 10.8 | 20 |
| **50–59** | LV GLS | ≥ 14.5 | 12.7–14.4 | < 12.7 | 77 | ≥ 14.2 | 13.0–14.1 | < 13.0 | 130 |
| | LV GCS | ≥ 15.5 | 14.3–15.4 | < 14.3 | 78 | ≥ 14.2 | 12.9–14.1 | < 12.9 | 124 |
| | LV GRS | ≥ 23.1 | 20.7–23.0 | < 20.7 | 78 | ≥ 20.4 | 18.0–20.3 | < 18.0 | 124 |
| | RV GLS | ≥ 18.9 | 16.5–18.8 | < 16.5 | 80 | ≥ 17.4 | 14.7–17.3 | < 14.7 | 131 |
| | RV GCS | ≥ 10.8 | 9.7–10.7 | < 9.7 | 79 | ≥ 9.4 | 7.8–9.3 | < 7.8 | 129 |
| | RV GRS | ≥ 15.5 | 13.6–15.4 | < 13.6 | 79 | ≥ 13.7 | 11.4–13.6 | < 11.4 | 129 |
| **60–69** | LV GLS | ≥ 15.0 | 13.2–14.9 | < 13.2 | 57 | ≥ 14.1 | 12.9–14.0 | < 12.9 | 110 |
| | LV GCS | ≥ 15.5 | 13.9–15.4 | < 13.9 | 54 | ≥ 14.0 | 12.3–13.9 | < 12.3 | 109 |
| | LV GRS | ≥ 23.1 | 20.0–23.0 | < 20.0 | 54 | ≥ 20.0 | 17.1–19.9 | < 17.1 | 109 |
| | RV GLS | ≥ 19.4 | 17.3–19.3 | < 17.3 | 59 | ≥ 18.1 | 15.8–18.0 | < 15.8 | 112 |
| | RV GCS | ≥ 10.3 | 8.8–10.2 | < 8.8 | 61 | ≥ 9.8 | 8.3–9.7 | < 8.3 | 120 |
| | RV GRS | ≥ 14.7 | 12.4–14.6 | < 12.4 | 61 | ≥ 14.3 | 12.1–14.2 | < 12.1 | 120 |
| **70+** | LV GLS | ≥ 15.3 | 13.6–15.2 | < 13.6 | 29 | ≥ 14.4 | 13.3–14.3 | < 13.3 | 33 |
| | LV GCS | ≥ 15.4 | 13.1–15.3 | < 13.1 | 29 | ≥ 13.5 | 10.9–13.4 | < 10.9 | 34 |
| | LV GRS | ≥ 23.0 | 18.7–22.9 | < 18.7 | 29 | ≥ 19.2 | 15.2–19.1 | < 15.2 | 34 |
| | RV GLS | ≥ 19.9 | 18.0–19.8 | < 18.0 | 28 | ≥ 18.5 | 16.6–18.4 | < 16.6 | 36 |
| | RV GCS | ≥ 9.5 | 7.7–9.4 | < 7.7 | 31 | ≥ 10.3 | 8.9–10.2 | < 8.9 | 39 |
| | RV GRS | ≥ 13.4 | 10.5–13.3 | < 10.5 | 31 | ≥ 15.1 | 12.9–15.0 | < 12.9 | 39 |

**Footnote**: All strain measurements were derived using cvi42 software (Circle Cardiovascular Imaging Inc.) with standardized feature-tracking analysis. Global longitudinal strain (GLS), global circumferential strain (GCS), and global radial strain (GRS) are presented as absolute values (%). Reference ranges are defined as the interval between the 2.5th and 97.5th centiles derived from age- and sex-specific generalized additive models for location, scale and shape (GAMLSS) using the Box–Cox Cole–Green distribution. Centile-based categories beyond the reference range are derived from LMS-based thresholds (P0.5 and P2.5) and describe the position of a measurement within the reference distribution rather than disease severity or diagnostic abnormality. For clarity, only the lower reference threshold (2.5th percentile) and corresponding lower centile cut-offs are presented in this table. Reference values are derived from a South Asian adult population.
**Abbreviations**: LV = left ventricle; RV = right ventricle; GLS = global longitudinal strain; GCS = global circumferential strain; GRS = global radial strain.

**Supplementary Table 15 Left and right ventricular global strain rate reference ranges and centile-based categories stratified by sex and age using cvi42 (Circle Cardiovascular Imaging Inc.), *in a South Asian adult population***

| | | Female | | | | Male | | | |
|---|---|---|---|---|---|---|---|---|---|
| **Age group** | Metrics ($s^{-1}$) | Reference threshold | Mild | At least moderate | n | Reference threshold | Mild | At least moderate | n |
| **40–49** | LV sysGLSR | | | | | ≥ 0.63 | 0.56–0.63 | < 0.56 | 19 |
| | LV sysGCSR | | | | | ≥ 0.75 | 0.70–0.75 | < 0.70 | 20 |
| | LV sysGRSR | | | | | ≥ 1.13 | 1.04–1.13 | < 1.04 | 20 |
| | LV diaGLSR | | | | | ≥ 0.70 | 0.64–0.70 | < 0.64 | 19 |
| | LV diaGCSR | | | | | ≥ 0.62 | 0.57–0.62 | < 0.57 | 20 |
| | LV diaGRSR | | | | | ≥ 0.97 | 0.87–0.97 | < 0.87 | 20 |
| | RV sysGLSR | | | | | ≥ 0.77 | 0.67–0.77 | < 0.67 | 19 |
| | RV sysGCSR | | | | | ≥ 0.48 | 0.40–0.48 | < 0.40 | 20 |
| | RV sysGRSR | | | | | ≥ 0.71 | 0.61–0.71 | < 0.61 | 20 |
| | RV diaGLSR | | | | | ≥ 0.73 | 0.60–0.73 | < 0.60 | 19 |
| | RV diaGCSR | | | | | ≥ 0.43 | 0.37–0.43 | < 0.37 | 20 |
| | RV diaGRSR | | | | | ≥ 0.67 | 0.54–0.67 | < 0.54 | 20 |
| **50–59** | LV sysGLSR | ≥ 0.69 | 0.65–0.69 | < 0.65 | 77 | ≥ 0.63 | 0.57–0.63 | < 0.57 | 130 |
| | LV sysGCSR | ≥ 0.74 | 0.68–0.74 | < 0.68 | 78 | ≥ 0.72 | 0.66–0.72 | < 0.66 | 124 |
| | LV sysGRSR | ≥ 1.13 | 1.00–1.13 | < 1.00 | 78 | ≥ 1.08 | 0.97–1.08 | < 0.97 | 124 |
| | LV diaGLSR | ≥ 0.74 | 0.66–0.74 | < 0.66 | 77 | ≥ 0.64 | 0.57–0.64 | < 0.57 | 130 |
| | LV diaGCSR | ≥ 0.66 | 0.57–0.66 | < 0.57 | 78 | ≥ 0.61 | 0.57–0.61 | < 0.57 | 124 |
| | LV diaGRSR | ≥ 0.98 | 0.76–0.98 | < 0.76 | 78 | ≥ 0.90 | 0.79–0.90 | < 0.79 | 124 |
| | RV sysGLSR | ≥ 0.86 | 0.78–0.86 | < 0.78 | 71 | ≥ 0.80 | 0.70–0.80 | < 0.70 | 128 |
| | RV sysGCSR | ≥ 0.50 | 0.44–0.50 | < 0.44 | 79 | ≥ 0.50 | 0.42–0.50 | < 0.42 | 129 |
| | RV sysGRSR | ≥ 0.75 | 0.65–0.75 | < 0.65 | 79 | ≥ 0.73 | 0.62–0.73 | < 0.62 | 129 |
| | RV diaGLSR | ≥ 0.86 | 0.71–0.86 | < 0.71 | 80 | ≥ 0.74 | 0.61–0.74 | < 0.61 | 131 |
| | RV diaGCSR | ≥ 0.50 | 0.45–0.50 | < 0.45 | 78 | ≥ 0.44 | 0.38–0.44 | < 0.38 | 126 |
| | RV diaGRSR | ≥ 0.85 | 0.75–0.85 | < 0.75 | 79 | ≥ 0.69 | 0.56–0.69 | < 0.56 | 124 |
| **60–69** | LV sysGLSR | ≥ 0.68 | 0.64–0.68 | < 0.64 | 56 | ≥ 0.62 | 0.57–0.62 | < 0.57 | 110 |
| | LV sysGCSR | ≥ 0.67 | 0.60–0.67 | < 0.60 | 54 | ≥ 0.67 | 0.61–0.67 | < 0.61 | 109 |
| | LV sysGRSR | ≥ 1.06 | 0.91–1.06 | < 0.91 | 54 | ≥ 1.00 | 0.87–1.00 | < 0.87 | 109 |
| | LV diaGLSR | ≥ 0.70 | 0.62–0.70 | < 0.62 | 57 | ≥ 0.63 | 0.55–0.63 | < 0.55 | 110 |
| | LV diaGCSR | ≥ 0.58 | 0.48–0.58 | < 0.48 | 54 | ≥ 0.61 | 0.55–0.61 | < 0.55 | 109 |
| | LV diaGRSR | ≥ 0.86 | 0.65–0.86 | < 0.65 | 54 | ≥ 0.81 | 0.69–0.81 | < 0.69 | 109 |
| | RV sysGLSR | ≥ 0.84 | 0.74–0.84 | < 0.74 | 53 | ≥ 0.83 | 0.74–0.83 | < 0.74 | 106 |
| | RV sysGCSR | ≥ 0.48 | 0.39–0.48 | < 0.39 | 61 | ≥ 0.53 | 0.44–0.53 | < 0.44 | 120 |
| | RV sysGRSR | ≥ 0.69 | 0.57–0.69 | < 0.57 | 61 | ≥ 0.75 | 0.62–0.75 | < 0.62 | 120 |
| | RV diaGLSR | ≥ 0.86 | 0.70–0.86 | < 0.70 | 59 | ≥ 0.75 | 0.63–0.75 | < 0.63 | 112 |
| | RV diaGCSR | ≥ 0.45 | 0.39–0.45 | < 0.39 | 59 | ≥ 0.45 | 0.38–0.45 | < 0.38 | 109 |
| | RV diaGRSR | ≥ 0.71 | 0.59–0.71 | < 0.59 | 57 | ≥ 0.73 | 0.60–0.73 | < 0.60 | 109 |
| **70+** | LV sysGLSR | ≥ 0.68 | 0.65–0.68 | < 0.65 | 29 | ≥ 0.63 | 0.58–0.63 | < 0.58 | 33 |
| | LV sysGCSR | ≥ 0.62 | 0.53–0.62 | < 0.53 | 29 | ≥ 0.62 | 0.55–0.62 | < 0.55 | 34 |
| | LV sysGRSR | ≥ 1.04 | 0.88–1.04 | < 0.88 | 29 | ≥ 0.88 | 0.71–0.88 | < 0.71 | 34 |
| | LV diaGLSR | ≥ 0.67 | 0.58–0.67 | < 0.58 | 29 | ≥ 0.71 | 0.62–0.71 | < 0.62 | 33 |
| | LV diaGCSR | ≥ 0.57 | 0.49–0.57 | < 0.49 | 28 | ≥ 0.60 | 0.53–0.60 | < 0.53 | 34 |
| | LV diaGRSR | ≥ 0.74 | 0.55–0.74 | < 0.55 | 29 | ≥ 0.70 | 0.55–0.70 | < 0.55 | 34 |
| | RV sysGLSR | ≥ 0.80 | 0.69–0.80 | < 0.69 | 26 | ≥ 0.87 | 0.78–0.87 | < 0.78 | 36 |
| | RV sysGCSR | ≥ 0.42 | 0.30–0.42 | < 0.30 | 30 | ≥ 0.56 | 0.46–0.56 | < 0.46 | 39 |
| | RV sysGRSR | ≥ 0.60 | 0.45–0.60 | < 0.45 | 31 | ≥ 0.77 | 0.62–0.77 | < 0.62 | 39 |
| | RV diaGLSR | ≥ 0.86 | 0.69–0.86 | < 0.69 | 28 | ≥ 0.77 | 0.64–0.77 | < 0.64 | 35 |
| | RV diaGCSR | ≥ 0.39 | 0.33–0.39 | < 0.33 | 30 | ≥ 0.42 | 0.35–0.42 | < 0.35 | 36 |
| | RV diaGRSR | ≥ 0.53 | 0.38–0.53 | < 0.38 | 28 | ≥ 0.79 | 0.65–0.79 | < 0.65 | 34 |

**Footnote**: All strain rate measurements were derived using cvi42 software (Circle Cardiovascular Imaging Inc.) with standardized feature-tracking analysis. Systolic and diastolic global longitudinal strain rate (GLSR), global circumferential strain rate (GCSR), and global radial strain rate (GRSR) are presented in $s^{-1}$. Reference ranges are defined as the interval between the 2.5th and 97.5th centiles derived from age- and sex-specific generalized additive models for location, scale and shape (GAMLSS) using the Box–Cox Cole–Green distribution. Centile-based categories beyond the reference range are derived from LMS-based thresholds (P0.5 and P2.5) and describe the position of a measurement within the reference distribution rather than disease severity or diagnostic abnormality. For clarity, only the lower reference threshold (2.5th percentile) and corresponding lower centile cut-offs are presented in this table. Reference values are derived from an South Asian adult population.

**Abbreviations**: LV = left ventricle; RV = right ventricle; GLSR = global longitudinal strain rate; GCSR = global circumferential strain rate; GRSR = global radial strain rate; sys = systolic; dia = diastolic.

**Supplementary Table 16 Left ventricular global displacement reference ranges and centile-based categories stratified by sex and age using cvi42 (Circle Cardiovascular Imaging Inc.),** ***in a South Asian adult population***

| | | **Female** | | | | **Male** | | | |
|---|---|---|---|---|---|---|---|---|---|
| **Age group** | Metrics (mm) | Reference threshold | Mild | At least moderate | n | Reference threshold | Mild | At least moderate | n |
| **40–49** | LV GLD | | | | | ≥ 2.8 | 2.2–2.7 | < 2.2 | 19 |
| | LV GCD | | | | | ≥ 0.7 | 0.5–0.6 | < 0.5 | 20 |
| | LV GRD | | | | | ≥ 3.7 | 3.3–3.6 | < 3.3 | 20 |
| **50–59** | LV GLD | ≥ 3.3 | 2.8–3.2 | < 2.8 | 77 | ≥ 2.8 | 2.2–2.7 | < 2.2 | 130 |
| | LV GCD | ≥ 0.7 | 0.4–0.6 | < 0.4 | 64 | ≥ 0.7 | 0.5–0.6 | < 0.5 | 112 |
| | LV GRD | ≥ 3.9 | 3.6–3.8 | < 3.6 | 78 | ≥ 3.7 | 3.3–3.6 | < 3.3 | 124 |
| **60–69** | LV GLD | ≥ 3.0 | 2.4–2.9 | < 2.4 | 57 | ≥ 2.7 | 2.2–2.6 | < 2.2 | 110 |
| | LV GCD | ≥ 0.6 | 0.4–0.5 | < 0.4 | 43 | ≥ 0.7 | 0.5–0.6 | < 0.5 | 90 |
| | LV GRD | ≥ 3.9 | 3.5–3.8 | < 3.5 | 54 | ≥ 3.7 | 3.2–3.6 | < 3.2 | 109 |
| **70+** | LV GLD | ≥ 2.7 | 2.1–2.6 | < 2.1 | 29 | ≥ 2.6 | 2.2–2.5 | < 2.2 | 33 |
| | LV GCD | ≥ 0.5 | 0.3–0.4 | < 0.3 | 27 | ≥ 0.8 | 0.6–0.7 | < 0.6 | 26 |
| | LV GRD | ≥ 3.8 | 3.3–3.7 | < 3.3 | 29 | ≥ 3.6 | 3.1–3.5 | < 3.1 | 34 |

**Footnote**: All displacement measurements were derived using cvi42 software (Circle Cardiovascular Imaging Inc.) with standardized feature-tracking analysis. Global longitudinal displacement (GLD), global circumferential displacement (GCD), and global radial displacement (GRD) are presented in millimetres (mm). Reference ranges are defined as the interval between the 2.5th and 97.5th centiles derived from age- and sex-specific generalized additive models for location, scale and shape (GAMLSS) using the Box–Cox Cole–Green distribution. Centile-based categories beyond the reference range are derived from LMS-based thresholds (P0.5 and P2.5) and describe the position of a measurement within the reference distribution rather than disease severity or diagnostic abnormality. For clarity, only the lower reference threshold (2.5th percentile) and corresponding lower centile cut-offs are presented in this table. Reference values are derived from an South Asian adult population.

**Abbreviations**: LV = left ventricle; GLD = global longitudinal displacement; GCD = global circumferential displacement; GRD = global radial displacement.

**Supplementary Table 17 Left ventricular global velocity reference ranges and centile-based categories stratified by sex and age using cvi42 (Circle Cardiovascular Imaging Inc.), *in a South Asian adult population***

| | | Female | | | | Male | | | |
|---|---|---|---|---|---|---|---|---|---|
| **Age group** | Metrics (%/s) | Reference threshold | Mild | At least moderate | n | Reference threshold | Mild | At least moderate | n |
| **40–49** | LV sysGLV | | | | | ≥ 15.8 | 14.1–15.7 | < 14.1 | 19 |
| | LV sysGCV | | | | | ≥ 12.9 | 10.0–12.8 | < 10.0 | 20 |
| | LV sysGRV | | | | | ≥ 20.6 | 18.7–20.5 | < 18.7 | 20 |
| | LV diaGLV | | | | | ≥ 16.4 | 13.5–16.3 | < 13.5 | 19 |
| | LV diaGCV | | | | | ≥ 5.3 | 4.3–5.2 | < 4.3 | 20 |
| | LV diaGRV | | | | | ≥ 17.3 | 15.6–17.2 | < 15.6 | 20 |
| **50–59** | LV sysGLV | ≥ 16.8 | 15.0–16.7 | < 15.0 | 77 | ≥ 15.3 | 13.4–15.2 | < 13.4 | 130 |
| | LV sysGCV | ≥ 11.1 | 8.6–11.0 | < 8.6 | 62 | ≥ 12.1 | 9.6–12.0 | < 9.6 | 114 |
| | LV sysGRV | ≥ 19.5 | 18.1–19.4 | < 18.1 | 78 | ≥ 19.7 | 17.8–19.6 | < 17.8 | 124 |
| | LV diaGLV | ≥ 19.2 | 16.7–19.1 | < 16.7 | 77 | ≥ 15.8 | 13.5–15.7 | < 13.5 | 130 |
| | LV diaGCV | ≥ 4.7 | 3.2–4.6 | < 3.2 | 60 | ≥ 5.2 | 4.2–5.1 | < 4.2 | 107 |
| | LV diaGRV | ≥ 17.8 | 15.7–17.7 | < 15.7 | 78 | ≥ 17.2 | 15.6–17.1 | < 15.6 | 124 |
| **60–69** | LV sysGLV | ≥ 15.9 | 14.0–15.8 | < 14.0 | 57 | ≥ 15.1 | 12.9–15.0 | < 12.9 | 110 |
| | LV sysGCV | ≥ 10.0 | 7.7–9.9 | < 7.7 | 37 | ≥ 12.3 | 9.9–12.2 | < 9.9 | 93 |
| | LV sysGRV | ≥ 17.6 | 16.0–17.5 | < 16.0 | 54 | ≥ 18.6 | 16.7–18.5 | < 16.7 | 109 |
| | LV diaGLV | ≥ 17.7 | 15.1–17.6 | < 15.1 | 57 | ≥ 15.6 | 13.3–15.5 | < 13.3 | 110 |
| | LV diaGCV | ≥ 4.7 | 3.2–4.6 | < 3.2 | 38 | ≥ 5.1 | 4.0–5.0 | < 4.0 | 81 |
| | LV diaGRV | ≥ 15.6 | 13.4–15.5 | < 13.4 | 54 | ≥ 17.1 | 15.5–17.0 | < 15.5 | 109 |
| **70+** | LV sysGLV | ≥ 13.7 | 11.8–13.6 | < 11.8 | 29 | ≥ 15.5 | 12.7–15.4 | < 12.7 | 33 |
| | LV sysGCV | ≥ 9.5 | 7.4–9.4 | < 7.4 | 23 | ≥ 13.4 | 11.0–13.3 | < 11.0 | 21 |
| | LV sysGRV | ≥ 15.6 | 13.7–15.5 | < 13.7 | 29 | ≥ 17.4 | 15.5–17.3 | < 15.5 | 34 |
| | LV diaGLV | ≥ 16.6 | 14.0–16.5 | < 14.0 | 29 | ≥ 15.9 | 12.8–15.8 | < 12.8 | 33 |
| | LV diaGCV | ≥ 4.9 | 3.4–4.8 | < 3.4 | 22 | ≥ 4.9 | 3.7–4.8 | < 3.7 | 25 |
| | LV diaGRV | ≥ 13.2 | 11.0–13.1 | < 11.0 | 29 | ≥ 17.0 | 15.4–16.9 | < 15.4 | 34 |

**Footnote**: All velocity measurements were derived using cvi42 software (Circle Cardiovascular Imaging Inc.) with standardized feature-tracking analysis. Systolic and diastolic global longitudinal velocity (GLV), global circumferential velocity (GCV), and global radial velocity (GRV) are presented in %/s. Reference ranges are defined as the interval between the 2.5th and 97.5th centiles derived from age- and sex-specific generalized additive models for location, scale and shape (GAMLSS) using the Box–Cox Cole–Green distribution. Centile-based categories beyond the reference range are derived from LMS-based thresholds (P0.5 and P2.5) and describe the position of a measurement within the reference distribution rather than disease severity or diagnostic abnormality. For clarity, only the lower reference threshold (2.5th percentile) and corresponding lower centile cut-offs are presented in this table. Reference values are derived from an South Asian adult population.

**Abbreviations**: LV = left ventricle; GLV = global longitudinal velocity; GCV = global circumferential velocity; GRV = global radial velocity; sys = systolic; dia = diastolic.

**Supplementary Table 18 Left and right ventricular global strain reference ranges and centile-based categories stratified by sex and age using cvi42 (Circle Cardiovascular Imaging Inc.), *in a Chinese adult population***

| | | Female | | | | Male | | | |
|---|---|---|---|---|---|---|---|---|---|
| **Age group** | Metrics (%) | Reference threshold | Mild | At least moderate | n | Reference threshold | Mild | At least moderate | n |
| **18–29** | LV GLS | ≥ 18.1 | 17.2–18.0 | < 17.2 | 12 | ≥ 14.9 | 13.5–14.8 | < 13.5 | 18 |
| | LV GCS | ≥ 15.5 | 13.5–15.4 | < 13.5 | 15 | ≥ 14.6 | 12.1–14.5 | < 12.1 | 19 |
| | LV GRS | ≥ 24.1 | 20.3–24.0 | < 20.3 | 15 | ≥ 22.9 | 18.9–22.8 | < 18.9 | 19 |
| | RV GLS | ≥ 22.3 | 21.0–22.2 | < 21.0 | 14 | ≥ 19.9 | 18.3–19.8 | < 18.3 | 18 |
| | RV GCS | ≥ 7.8 | 5.1–7.7 | < 5.1 | 14 | ≥ 10.5 | 9.2–10.4 | < 9.2 | 18 |
| | RV GRS | ≥ 17.1 | 13.8–17.0 | < 13.8 | 14 | ≥ 20.1 | 17.9–20.0 | < 17.9 | 18 |
| **30–39** | LV GLS | ≥ 17.3 | 16.2–17.2 | < 16.2 | 17 | ≥ 14.7 | 13.4–14.6 | < 13.4 | 15 |
| | LV GCS | ≥ 15.5 | 13.4–15.4 | < 13.4 | 18 | ≥ 14.4 | 12.0–14.3 | < 12.0 | 17 |
| | LV GRS | ≥ 23.9 | 19.8–23.8 | < 19.8 | 18 | ≥ 22.4 | 19.0–22.3 | < 19.0 | 17 |
| | RV GLS | ≥ 22.0 | 20.5–21.9 | < 20.5 | 17 | ≥ 19.5 | 17.8–19.4 | < 17.8 | 15 |
| | RV GCS | ≥ 8.4 | 5.9–8.3 | < 5.9 | 16 | ≥ 10.2 | 8.7–10.1 | < 8.7 | 16 |
| | RV GRS | ≥ 15.8 | 12.6–15.7 | < 12.6 | 16 | ≥ 18.7 | 16.3–18.6 | < 16.3 | 16 |
| **40–49** | LV GLS | ≥ 16.3 | 14.8–16.2 | < 14.8 | 21 | ≥ 14.4 | 13.4–14.3 | < 13.4 | 19 |
| | LV GCS | ≥ 15.5 | 13.4–15.4 | < 13.4 | 24 | ≥ 14.4 | 12.5–14.3 | < 12.5 | 20 |
| | LV GRS | ≥ 23.6 | 19.3–23.5 | < 19.3 | 24 | ≥ 22.0 | 19.1–21.9 | < 19.1 | 20 |
| | RV GLS | ≥ 21.6 | 19.5–21.5 | < 19.5 | 22 | ≥ 19.0 | 17.0–18.9 | < 17.0 | 19 |
| | RV GCS | ≥ 8.9 | 6.8–8.8 | < 6.8 | 24 | ≥ 9.8 | 8.0–9.7 | < 8.0 | 18 |
| | RV GRS | ≥ 14.5 | 11.3–14.4 | < 11.3 | 24 | ≥ 17.2 | 14.6–17.1 | < 14.6 | 18 |
| **50–59** | LV GLS | ≥ 15.1 | 13.3–15.0 | < 13.3 | 63 | ≥ 14.2 | 13.3–14.1 | < 13.3 | 43 |
| | LV GCS | ≥ 15.5 | 13.3–15.4 | < 13.3 | 65 | ≥ 14.6 | 13.2–14.5 | < 13.2 | 47 |
| | LV GRS | ≥ 23.2 | 18.8–23.1 | < 18.8 | 65 | ≥ 21.6 | 19.2–21.5 | < 19.2 | 47 |
| | RV GLS | ≥ 20.8 | 17.8–20.7 | < 17.8 | 65 | ≥ 18.5 | 16.1–18.4 | < 16.1 | 47 |
| | RV GCS | ≥ 9.4 | 7.6–9.3 | < 7.6 | 66 | ≥ 9.3 | 7.1–9.2 | < 7.1 | 46 |
| | RV GRS | ≥ 13.8 | 10.6–13.7 | < 10.6 | 65 | ≥ 15.5 | 12.6–15.4 | < 12.6 | 46 |
| **60–69** | LV GLS | ≥ 13.8 | 11.7–13.7 | < 11.7 | 50 | ≥ 14.0 | 13.2–13.9 | < 13.2 | 33 |
| | LV GCS | ≥ 15.5 | 13.2–15.4 | < 13.2 | 49 | ≥ 14.6 | 13.7–14.5 | < 13.7 | 36 |
| | LV GRS | ≥ 22.9 | 18.2–22.8 | < 18.2 | 49 | ≥ 21.3 | 19.2–21.2 | < 19.2 | 36 |
| | RV GLS | ≥ 19.8 | 15.3–19.7 | < 15.3 | 51 | ≥ 17.8 | 15.0–17.7 | < 15.0 | 32 |
| | RV GCS | ≥ 9.9 | 8.3–9.8 | < 8.3 | 50 | ≥ 8.6 | 5.9–8.5 | < 5.9 | 36 |
| | RV GRS | ≥ 13.8 | 10.7–13.7 | < 10.7 | 47 | ≥ 13.7 | 10.4–13.6 | < 10.4 | 35 |
| **70+** | LV GLS | ≥ 12.7 | 10.5–12.6 | < 10.5 | 12 | | | | |
| | LV GCS | ≥ 15.5 | 13.2–15.4 | < 13.2 | 12 | | | | |
| | LV GRS | ≥ 22.6 | 17.6–22.5 | < 17.6 | 12 | | | | |
| | RV GLS | ≥ 19.1 | 14.2–19.0 | < 14.2 | 12 | | | | |
| | RV GCS | ≥ 10.3 | 8.8–10.2 | < 8.8 | 12 | | | | |
| | RV GRS | ≥ 14.1 | 11.3–14.0 | < 11.3 | 11 | | | | |

**Footnote**: All strain measurements were derived using cvi42 software (Circle Cardiovascular Imaging Inc.) with standardized feature-tracking analysis. Global longitudinal strain (GLS), global circumferential strain (GCS), and global radial strain (GRS) are presented as absolute values (%). Reference ranges are defined as the interval between the 2.5th and 97.5th centiles derived from age- and sex-specific generalized additive models for location, scale and shape (GAMLSS) using the Box–Cox Cole–Green distribution. Centile-based categories beyond the reference range are derived from LMS-based thresholds (P0.5 and P2.5) and describe the position of a measurement within the reference distribution rather than disease severity or diagnostic abnormality. For clarity, only the lower reference threshold (2.5th percentile) and corresponding lower centile cut-offs are presented in this table. Reference values are derived from a Chinese adult population.
**Abbreviations**: LV = left ventricle; RV = right ventricle; GLS = global longitudinal strain; GCS = global circumferential strain; GRS = global radial strain.

**Supplementary Table 19 Left and right ventricular global strain rate reference ranges and centile-based categories stratified by sex and age using cvi42 (Circle Cardiovascular Imaging Inc.),** ***in a Chinese adult population***

| | | Female | | | | Male | | | |
|---|---|---|---|---|---|---|---|---|---|
| **Age group** | Metrics ($s^{-1}$) | Reference threshold | Mild | At least moderate | n | Reference threshold | Mild | At least moderate | n |
| **18–29** | LV sysGLSR | ≥ 0.79 | 0.75–0.79 | < 0.75 | 12 | ≥ 0.63 | 0.56–0.63 | < 0.56 | 18 |
| | LV sysGCSR | ≥ 0.82 | 0.77–0.82 | < 0.77 | 13 | ≥ 0.80 | 0.75–0.80 | < 0.75 | 19 |
| | LV sysGRSR | ≥ 1.28 | 1.10–1.28 | < 1.10 | 14 | ≥ 1.24 | 1.12–1.24 | < 1.12 | 19 |
| | LV diaGLSR | ≥ 0.70 | 0.49–0.70 | < 0.49 | 12 | ≥ 0.54 | 0.40–0.54 | < 0.40 | 18 |
| | LV diaGCSR | ≥ 0.68 | 0.52–0.68 | < 0.52 | 15 | ≥ 0.62 | 0.54–0.62 | < 0.54 | 19 |
| | LV diaGRSR | ≥ 1.27 | 0.97–1.27 | < 0.97 | 15 | ≥ 1.10 | 0.91–1.10 | < 0.91 | 19 |
| | RV sysGLSR | ≥ 0.85 | 0.78–0.85 | < 0.78 | 14 | ≥ 0.85 | 0.78–0.85 | < 0.78 | 18 |
| | RV sysGCSR | ≥ 0.39 | 0.32–0.39 | < 0.32 | 14 | ≥ 0.53 | 0.48–0.53 | < 0.48 | 18 |
| | RV sysGRSR | ≥ 0.63 | 0.44–0.63 | < 0.44 | 14 | ≥ 0.95 | 0.82–0.95 | < 0.82 | 18 |
| | RV diaGLSR | ≥ 0.98 | 0.89–0.98 | < 0.89 | 14 | ≥ 0.77 | 0.55–0.77 | < 0.55 | 18 |
| | RV diaGCSR | ≥ 0.37 | 0.33–0.37 | < 0.33 | 11 | ≥ 0.40 | 0.32–0.40 | < 0.32 | 15 |
| | RV diaGRSR | ≥ 0.83 | 0.70–0.83 | < 0.70 | 11 | ≥ 0.85 | 0.68–0.85 | < 0.68 | 17 |
| **30–39** | LV sysGLSR | ≥ 0.76 | 0.72–0.76 | < 0.72 | 17 | ≥ 0.64 | 0.58–0.64 | < 0.58 | 15 |
| | LV sysGCSR | ≥ 0.79 | 0.74–0.79 | < 0.74 | 18 | ≥ 0.77 | 0.71–0.77 | < 0.71 | 17 |
| | LV sysGRSR | ≥ 1.25 | 1.09–1.25 | < 1.09 | 18 | ≥ 1.21 | 1.10–1.21 | < 1.10 | 17 |
| | LV diaGLSR | ≥ 0.69 | 0.50–0.69 | < 0.50 | 17 | ≥ 0.56 | 0.45–0.56 | < 0.45 | 15 |
| | LV diaGCSR | ≥ 0.67 | 0.54–0.67 | < 0.54 | 18 | ≥ 0.61 | 0.53–0.61 | < 0.53 | 17 |
| | LV diaGRSR | ≥ 1.23 | 0.97–1.23 | < 0.97 | 17 | ≥ 1.09 | 0.95–1.09 | < 0.95 | 17 |
| | RV sysGLSR | ≥ 0.87 | 0.79–0.87 | < 0.79 | 17 | ≥ 0.86 | 0.78–0.86 | < 0.78 | 15 |
| | RV sysGCSR | ≥ 0.42 | 0.35–0.42 | < 0.35 | 16 | ≥ 0.53 | 0.47–0.53 | < 0.47 | 16 |
| | RV sysGRSR | ≥ 0.68 | 0.51–0.68 | < 0.51 | 16 | ≥ 0.91 | 0.77–0.91 | < 0.77 | 15 |
| | RV diaGLSR | ≥ 0.99 | 0.89–0.99 | < 0.89 | 17 | ≥ 0.80 | 0.61–0.80 | < 0.61 | 15 |
| | RV diaGCSR | ≥ 0.40 | 0.35–0.40 | < 0.35 | 14 | ≥ 0.39 | 0.32–0.39 | < 0.32 | 14 |
| | RV diaGRSR | ≥ 0.82 | 0.70–0.82 | < 0.70 | 12 | ≥ 0.79 | 0.63–0.79 | < 0.63 | 13 |
| **40–49** | LV sysGLSR | ≥ 0.73 | 0.69–0.73 | < 0.69 | 21 | ≥ 0.65 | 0.60–0.65 | < 0.60 | 19 |
| | LV sysGCSR | ≥ 0.77 | 0.71–0.77 | < 0.71 | 24 | ≥ 0.75 | 0.68–0.75 | < 0.68 | 20 |
| | LV sysGRSR | ≥ 1.23 | 1.08–1.23 | < 1.08 | 24 | ≥ 1.19 | 1.08–1.19 | < 1.08 | 20 |
| | LV diaGLSR | ≥ 0.69 | 0.55–0.69 | < 0.55 | 21 | ≥ 0.58 | 0.49–0.58 | < 0.49 | 19 |
| | LV diaGCSR | ≥ 0.66 | 0.55–0.66 | < 0.55 | 24 | ≥ 0.59 | 0.52–0.59 | < 0.52 | 20 |
| | LV diaGRSR | ≥ 1.18 | 0.97–1.18 | < 0.97 | 24 | ≥ 1.03 | 0.93–1.03 | < 0.93 | 20 |
| | RV sysGLSR | ≥ 0.89 | 0.82–0.89 | < 0.82 | 21 | ≥ 0.86 | 0.78–0.86 | < 0.78 | 18 |
| | RV sysGCSR | ≥ 0.45 | 0.38–0.45 | < 0.38 | 23 | ≥ 0.53 | 0.45–0.53 | < 0.45 | 18 |
| | RV sysGRSR | ≥ 0.72 | 0.57–0.72 | < 0.57 | 24 | ≥ 0.87 | 0.72–0.87 | < 0.72 | 18 |
| | RV diaGLSR | ≥ 0.99 | 0.86–0.99 | < 0.86 | 22 | ≥ 0.82 | 0.67–0.82 | < 0.67 | 19 |
| | RV diaGCSR | ≥ 0.42 | 0.37–0.42 | < 0.37 | 22 | ≥ 0.39 | 0.32–0.39 | < 0.32 | 16 |
| | RV diaGRSR | ≥ 0.81 | 0.69–0.81 | < 0.69 | 22 | ≥ 0.73 | 0.58–0.73 | < 0.58 | 14 |
| **50–59** | LV sysGLSR | ≥ 0.70 | 0.66–0.70 | < 0.66 | 63 | ≥ 0.65 | 0.61–0.65 | < 0.61 | 43 |
| | LV sysGCSR | ≥ 0.74 | 0.69–0.74 | < 0.69 | 65 | ≥ 0.73 | 0.67–0.73 | < 0.67 | 47 |
| | LV sysGRSR | ≥ 1.20 | 1.06–1.20 | < 1.06 | 65 | ≥ 1.16 | 1.05–1.16 | < 1.05 | 47 |
| | LV diaGLSR | ≥ 0.69 | 0.59–0.69 | < 0.59 | 63 | ≥ 0.60 | 0.53–0.60 | < 0.53 | 43 |
| | LV diaGCSR | ≥ 0.64 | 0.55–0.64 | < 0.55 | 65 | ≥ 0.57 | 0.51–0.57 | < 0.51 | 47 |
| | LV diaGRSR | ≥ 1.12 | 0.94–1.12 | < 0.94 | 65 | ≥ 0.94 | 0.84–0.94 | < 0.84 | 47 |
| | RV sysGLSR | ≥ 0.91 | 0.84–0.91 | < 0.84 | 58 | ≥ 0.86 | 0.78–0.86 | < 0.78 | 46 |
| | RV sysGCSR | ≥ 0.49 | 0.41–0.49 | < 0.41 | 66 | ≥ 0.52 | 0.42–0.52 | < 0.42 | 46 |
| | RV sysGRSR | ≥ 0.75 | 0.62–0.75 | < 0.62 | 66 | ≥ 0.82 | 0.67–0.82 | < 0.67 | 46 |
| | RV diaGLSR | ≥ 0.96 | 0.81–0.96 | < 0.81 | 65 | ≥ 0.86 | 0.73–0.86 | < 0.73 | 47 |
| | RV diaGCSR | ≥ 0.45 | 0.40–0.45 | < 0.40 | 65 | ≥ 0.39 | 0.32–0.39 | < 0.32 | 43 |
| | RV diaGRSR | ≥ 0.79 | 0.68–0.79 | < 0.68 | 65 | ≥ 0.67 | 0.53–0.67 | < 0.53 | 44 |
| **60–69** | LV sysGLSR | ≥ 0.67 | 0.62–0.67 | < 0.62 | 50 | ≥ 0.65 | 0.61–0.65 | < 0.61 | 33 |
| | LV sysGCSR | ≥ 0.72 | 0.66–0.72 | < 0.66 | 49 | ≥ 0.72 | 0.66–0.72 | < 0.66 | 36 |
| | LV sysGRSR | ≥ 1.18 | 1.05–1.18 | < 1.05 | 49 | ≥ 1.14 | 1.03–1.14 | < 1.03 | 36 |
| | LV diaGLSR | ≥ 0.68 | 0.61–0.68 | < 0.61 | 50 | ≥ 0.62 | 0.56–0.62 | < 0.56 | 33 |
| | LV diaGCSR | ≥ 0.63 | 0.55–0.63 | < 0.55 | 49 | ≥ 0.55 | 0.49–0.55 | < 0.49 | 36 |
| | LV diaGRSR | ≥ 1.05 | 0.90–1.05 | < 0.90 | 49 | ≥ 0.81 | 0.68–0.81 | < 0.68 | 36 |
| | RV sysGLSR | ≥ 0.94 | 0.86–0.94 | < 0.86 | 44 | ≥ 0.86 | 0.77–0.86 | < 0.77 | 32 |
| | RV sysGCSR | ≥ 0.52 | 0.44–0.52 | < 0.44 | 50 | ≥ 0.49 | 0.37–0.49 | < 0.37 | 36 |
| | RV sysGRSR | ≥ 0.78 | 0.66–0.78 | < 0.66 | 48 | ≥ 0.77 | 0.61–0.77 | < 0.61 | 36 |
| | RV diaGLSR | ≥ 0.91 | 0.71–0.91 | < 0.71 | 50 | ≥ 0.89 | 0.78–0.89 | < 0.78 | 31 |
| | RV diaGCSR | ≥ 0.47 | 0.42–0.47 | < 0.42 | 48 | ≥ 0.38 | 0.31–0.38 | < 0.31 | 30 |
| | RV diaGRSR | ≥ 0.78 | 0.67–0.78 | < 0.67 | 43 | ≥ 0.62 | 0.49–0.62 | < 0.49 | 26 |
| **70+** | LV sysGLSR | ≥ 0.65 | 0.60–0.65 | < 0.60 | 12 | | | | |
| | LV sysGCSR | ≥ 0.70 | 0.64–0.70 | < 0.64 | 12 | | | | |
| | LV sysGRSR | ≥ 1.15 | 1.03–1.15 | < 1.03 | 12 | | | | |
| | LV diaGLSR | ≥ 0.68 | 0.62–0.68 | < 0.62 | 12 | | | | |
| | LV diaGCSR | ≥ 0.62 | 0.55–0.62 | < 0.55 | 12 | | | | |
| | LV diaGRSR | ≥ 0.99 | 0.86–0.99 | < 0.86 | 12 | | | | |
| | RV sysGLSR | ≥ 0.95 | 0.88–0.95 | < 0.88 | 11 | | | | |
| | RV sysGCSR | ≥ 0.55 | 0.47–0.55 | < 0.47 | 12 | | | | |
| | RV sysGRSR | ≥ 0.80 | 0.69–0.80 | < 0.69 | 11 | | | | |
| | RV diaGLSR | ≥ 0.84 | 0.58–0.84 | < 0.58 | 12 | | | | |
| | RV diaGCSR | ≥ 0.49 | 0.44–0.49 | < 0.44 | 12 | | | | |
| | RV diaGRSR | ≥ 0.77 | 0.66–0.77 | < 0.66 | 10 | | | | |

**Footnote**: All strain rate measurements were derived using cvi42 software (Circle Cardiovascular Imaging Inc.) with standardized feature-tracking analysis. Systolic and diastolic global longitudinal strain rate (GLSR), global circumferential strain rate (GCSR), and global radial strain rate (GRSR) are presented in $s^{-1}$. Reference ranges are defined as the interval between the 2.5th and 97.5th centiles derived from age- and sex-specific generalized additive models for location, scale and shape (GAMLSS) using the Box–Cox Cole–Green distribution. Centile-based categories beyond the reference range are derived from LMS-based thresholds (P0.5 and P2.5) and describe the position of a measurement within the reference distribution rather than disease severity or diagnostic abnormality. For clarity, only the lower reference threshold (2.5th percentile) and corresponding lower centile cut-offs are presented in this table. Reference values are derived from a Chinese adult population.

**Abbreviations**: LV = left ventricle; RV = right ventricle; GLSR = global longitudinal strain rate; GCSR = global circumferential strain rate; GRSR = global radial strain rate; sys = systolic; dia = diastolic.

**Supplementary Table 20 Left ventricular global displacement reference ranges and centile-based categories stratified by sex and age using cvi42 (Circle Cardiovascular Imaging Inc.), *in a Chinese adult population***

| | | Female | | | | Male | | | |
|---|---|---|---|---|---|---|---|---|---|
| **Age group** | Metrics (mm) | Reference threshold | Mild | At least moderate | n | Reference threshold | Mild | At least moderate | n |
| **18–29** | LV GLD | ≥ 4.2 | 3.5–4.1 | < 3.5 | 12 | ≥ 3.0 | 2.2–2.9 | < 2.2 | 18 |
| | LV GCD | ≥ 1.3 | 1.0–1.2 | < 1.0 | 13 | ≥ 1.1 | 0.9–1.0 | < 0.9 | 17 |
| | LV GRD | ≥ 4.1 | 3.7–4.0 | < 3.7 | 15 | ≥ 4.4 | 4.0–4.3 | < 4.0 | 19 |
| **30–39** | LV GLD | ≥ 3.9 | 3.2–3.8 | < 3.2 | 17 | ≥ 2.5 | 1.7–2.4 | < 1.7 | 15 |
| | LV GCD | ≥ 1.1 | 0.8–1.0 | < 0.8 | 16 | ≥ 1.0 | 0.8–0.9 | < 0.8 | 16 |
| | LV GRD | ≥ 4.2 | 3.9–4.1 | < 3.9 | 18 | ≥ 4.2 | 3.8–4.1 | < 3.8 | 17 |
| **40–49** | LV GLD | ≥ 3.5 | 2.9–3.4 | < 2.9 | 21 | ≥ 2.0 | 1.2–1.9 | < 1.2 | 19 |
| | LV GCD | ≥ 0.9 | 0.7–0.8 | < 0.7 | 24 | ≥ 0.9 | 0.7–0.8 | < 0.7 | 20 |
| | LV GRD | ≥ 4.3 | 4.0–4.2 | < 4.0 | 24 | ≥ 4.0 | 3.6–3.9 | < 3.6 | 20 |
| **50–59** | LV GLD | ≥ 3.2 | 2.5–3.1 | < 2.5 | 63 | ≥ 2.3 | 1.4–2.2 | < 1.4 | 43 |
| | LV GCD | ≥ 0.8 | 0.5–0.7 | < 0.5 | 56 | ≥ 0.9 | 0.7–0.8 | < 0.7 | 43 |
| | LV GRD | ≥ 4.3 | 3.9–4.2 | < 3.9 | 65 | ≥ 3.9 | 3.5–3.8 | < 3.5 | 47 |
| **60–69** | LV GLD | ≥ 2.8 | 2.2–2.7 | < 2.2 | 50 | ≥ 3.0 | 2.6–2.9 | < 2.6 | 33 |
| | LV GCD | ≥ 0.6 | 0.3–0.5 | < 0.3 | 41 | ≥ 0.8 | 0.6–0.7 | < 0.6 | 31 |
| | LV GRD | ≥ 4.1 | 3.7–4.0 | < 3.7 | 49 | ≥ 3.7 | 3.3–3.6 | < 3.3 | 36 |
| **70+** | LV GLD | ≥ 2.6 | 1.9–2.5 | < 1.9 | 12 | | | | |
| | LV GCD | | | | | | | | |
| | LV GRD | ≥ 3.9 | 3.4–3.8 | < 3.4 | 12 | | | | |

**Footnote**: All displacement measurements were derived using cvi42 software (Circle Cardiovascular Imaging Inc.) with standardized feature-tracking analysis. Global longitudinal displacement (GLD), global circumferential displacement (GCD), and global radial displacement (GRD) are presented in millimetres (mm). Reference ranges are defined as the interval between the 2.5th and 97.5th centiles derived from age- and sex-specific generalized additive models for location, scale and shape (GAMLSS) using the Box–Cox Cole–Green distribution. Centile-based categories beyond the reference range are derived from LMS-based thresholds (P0.5 and P2.5) and describe the position of a measurement within the reference distribution rather than disease severity or diagnostic abnormality. For clarity, only the lower reference threshold (2.5th percentile) and corresponding lower centile cut-offs are presented in this table. Reference values are derived from a Chinese adult population.
**Abbreviations**: LV = left ventricle; GLD = global longitudinal displacement; GCD = global circumferential displacement; GRD = global radial displacement.

**Supplementary Table 21 Left ventricular global velocity reference ranges and centile-based categories stratified by sex and age using cvi42 (Circle Cardiovascular Imaging Inc.),** ***in a Chinese adult population***

| | | Female | | | | Male | | | |
|---|---|---|---|---|---|---|---|---|---|
| **Age group** | Metrics (%/s) | Reference threshold | Mild | At least moderate | n | Reference threshold | Mild | At least moderate | n |
| **18–29** | LV sysGLV | ≥ 19.8 | 16.7–19.7 | < 16.7 | 12 | ≥ 16.3 | 12.2–16.2 | < 12.2 | 18 |
| | LV sysGCV | ≥ 16.6 | 13.4–16.5 | < 13.4 | 15 | ≥ 17.2 | 13.9–17.1 | < 13.9 | 18 |
| | LV sysGRV | ≥ 22.3 | 20.9–22.2 | < 20.9 | 14 | ≥ 24.2 | 22.8–24.1 | < 22.8 | 19 |
| | LV diaGLV | ≥ 21.1 | 13.6–21.0 | < 13.6 | 12 | ≥ 19.5 | 14.5–19.4 | < 14.5 | 18 |
| | LV diaGCV | ≥ 9.6 | 7.5–9.5 | < 7.5 | 12 | ≥ 7.6 | 6.0–7.5 | < 6.0 | 16 |
| | LV diaGRV | ≥ 18.1 | 13.2–18.0 | < 13.2 | 15 | ≥ 20.9 | 18.9–20.8 | < 18.9 | 19 |
| **30–39** | LV sysGLV | ≥ 18.6 | 15.9–18.5 | < 15.9 | 17 | ≥ 17.0 | 13.9–16.9 | < 13.9 | 15 |
| | LV sysGCV | ≥ 14.8 | 11.5–14.7 | < 11.5 | 18 | ≥ 15.9 | 12.7–15.8 | < 12.7 | 17 |
| | LV sysGRV | ≥ 21.6 | 20.3–21.5 | < 20.3 | 18 | ≥ 23.0 | 21.5–22.9 | < 21.5 | 17 |
| | LV diaGLV | ≥ 20.0 | 13.7–19.9 | < 13.7 | 16 | ≥ 14.7 | 9.7–14.6 | < 9.7 | 15 |
| | LV diaGCV | ≥ 8.5 | 6.5–8.4 | < 6.5 | 15 | ≥ 7.3 | 5.8–7.2 | < 5.8 | 15 |
| | LV diaGRV | ≥ 18.4 | 14.4–18.3 | < 14.4 | 17 | ≥ 19.5 | 17.6–19.4 | < 17.6 | 17 |
| **40–49** | LV sysGLV | ≥ 17.4 | 14.9–17.3 | < 14.9 | 21 | ≥ 17.2 | 14.8–17.1 | < 14.8 | 19 |
| | LV sysGCV | ≥ 13.0 | 9.6–12.9 | < 9.6 | 23 | ≥ 14.6 | 11.5–14.5 | < 11.5 | 18 |
| | LV sysGRV | ≥ 20.9 | 19.6–20.8 | < 19.6 | 24 | ≥ 21.8 | 20.1–21.7 | < 20.1 | 20 |
| | LV diaGLV | ≥ 18.9 | 14.0–18.8 | < 14.0 | 21 | ≥ 8.2 | 4.0–8.1 | < 4.0 | 19 |
| | LV diaGCV | ≥ 7.4 | 5.4–7.3 | < 5.4 | 21 | ≥ 7.0 | 5.6–6.9 | < 5.6 | 16 |
| | LV diaGRV | ≥ 18.6 | 15.6–18.5 | < 15.6 | 24 | ≥ 18.1 | 16.2–18.0 | < 16.2 | 20 |
| **50–59** | LV sysGLV | ≥ 16.1 | 13.9–16.0 | < 13.9 | 63 | ≥ 17.2 | 15.3–17.1 | < 15.3 | 43 |
| | LV sysGCV | ≥ 11.1 | 7.6–11.0 | < 7.6 | 59 | ≥ 13.3 | 10.3–13.2 | < 10.3 | 41 |
| | LV sysGRV | ≥ 20.2 | 18.9–20.1 | < 18.9 | 65 | ≥ 20.9 | 19.2–20.8 | < 19.2 | 47 |
| | LV diaGLV | ≥ 17.8 | 14.1–17.7 | < 14.1 | 63 | ≥ 14.2 | 11.3–14.1 | < 11.3 | 42 |
| | LV diaGCV | ≥ 6.3 | 4.4–6.2 | < 4.4 | 48 | ≥ 6.6 | 5.4–6.5 | < 5.4 | 35 |
| | LV diaGRV | ≥ 18.6 | 16.4–18.5 | < 16.4 | 65 | ≥ 16.7 | 14.9–16.6 | < 14.9 | 47 |
| **60–69** | LV sysGLV | ≥ 14.9 | 12.9–14.8 | < 12.9 | 50 | ≥ 17.1 | 15.4–17.0 | < 15.4 | 33 |
| | LV sysGCV | ≥ 9.2 | 5.6–9.1 | < 5.6 | 38 | ≥ 12.1 | 9.3–12.0 | < 9.3 | 32 |
| | LV sysGRV | ≥ 19.5 | 18.3–19.4 | < 18.3 | 49 | ≥ 20.2 | 18.8–20.1 | < 18.8 | 36 |
| | LV diaGLV | ≥ 16.6 | 13.7–16.5 | < 13.7 | 50 | ≥ 19.3 | 18.0–19.2 | < 18.0 | 33 |
| | LV diaGCV | ≥ 5.3 | 3.5–5.2 | < 3.5 | 41 | ≥ 6.3 | 5.1–6.2 | < 5.1 | 29 |
| | LV diaGRV | ≥ 18.3 | 16.8–18.2 | < 16.8 | 49 | ≥ 15.3 | 13.6–15.2 | < 13.6 | 36 |
| **70+** | LV sysGLV | ≥ 13.8 | 12.1–13.7 | < 12.1 | 12 | | | | |
| | LV sysGCV | | | | | | | | |
| | LV sysGRV | ≥ 18.9 | 17.7–18.8 | < 17.7 | 12 | | | | |
| | LV diaGLV | ≥ 15.5 | 13.1–15.4 | < 13.1 | 12 | | | | |
| | LV diaGCV | | | | | | | | |
| | LV diaGRV | ≥ 17.9 | 16.7–17.8 | < 16.7 | 12 | | | | |

**Footnote**: All velocity measurements were derived using cvi42 software (Circle Cardiovascular Imaging Inc.) with standardized feature-tracking analysis. Systolic and diastolic global longitudinal velocity (GLV), global circumferential velocity (GCV), and global radial velocity (GRV) are presented in %/s. Reference ranges are defined as the interval between the 2.5th and 97.5th centiles derived from age- and sex-specific generalized additive models for location, scale and shape (GAMLSS) using the Box–Cox Cole–Green distribution. Centile-based categories beyond the reference range are derived from LMS-based thresholds (P0.5 and P2.5) and describe the position of a measurement within the reference distribution rather than disease severity or diagnostic abnormality. For clarity, only the lower reference threshold (2.5th percentile) and corresponding lower centile cut-offs are presented in this table. Reference values are derived from a Chinese adult population.
**Abbreviations**: LV = left ventricle; GLV = global longitudinal velocity; GCV = global circumferential velocity; GRV = global radial velocity; sys = systolic; dia = diastolic.

### Supplementary Table 22 Left and right ventricular global strain reference ranges and centile-based categories stratified by sex and age using cvi42 (Circle Cardiovascular Imaging Inc.), *in a Mixed adult population*

| | | Female | | | | Male | | | |
|---|---|---|---|---|---|---|---|---|---|
| **Age group** | Metrics (%) | Reference threshold | Mild | At least moderate | n | Reference threshold | Mild | At least moderate | n |
| **50–59** | LV GLS | ≥ 15.4 | 14.2–15.3 | < 14.2 | 133 | ≥ 13.4 | 11.8–13.3 | < 11.8 | 61 |
| | LV GCS | ≥ 15.8 | 14.7–15.7 | < 14.7 | 124 | ≥ 14.5 | 13.5–14.4 | < 13.5 | 59 |
| | LV GRS | ≥ 23.8 | 21.5–23.7 | < 21.5 | 124 | ≥ 21.2 | 19.1–21.1 | < 19.1 | 59 |
| | RV GLS | ≥ 18.7 | 15.8–18.6 | < 15.8 | 135 | ≥ 16.8 | 13.8–16.7 | < 13.8 | 63 |
| | RV GCS | ≥ 8.8 | 6.9–8.7 | < 6.9 | 131 | ≥ 9.5 | 7.8–9.4 | < 7.8 | 64 |
| | RV GRS | ≥ 12.6 | 10.0–12.5 | < 10.0 | 131 | ≥ 13.6 | 11.2–13.5 | < 11.2 | 64 |
| **60–69** | LV GLS | ≥ 15.3 | 14.2–15.2 | < 14.2 | 107 | ≥ 13.5 | 11.9–13.4 | < 11.9 | 53 |
| | LV GCS | ≥ 15.5 | 14.1–15.4 | < 14.1 | 103 | ≥ 14.4 | 13.3–14.3 | < 13.3 | 54 |
| | LV GRS | ≥ 23.0 | 20.3–22.9 | < 20.3 | 103 | ≥ 20.9 | 18.8–20.8 | < 18.8 | 54 |
| | RV GLS | ≥ 18.4 | 15.1–18.3 | < 15.1 | 108 | ≥ 17.5 | 14.4–17.4 | < 14.4 | 57 |
| | RV GCS | ≥ 9.6 | 7.9–9.5 | < 7.9 | 107 | ≥ 9.6 | 7.9–9.5 | < 7.9 | 61 |
| | RV GRS | ≥ 13.7 | 11.2–13.6 | < 11.2 | 107 | ≥ 13.8 | 11.3–13.7 | < 11.3 | 61 |
| **70+** | LV GLS | ≥ 14.6 | 13.4–14.5 | < 13.4 | 43 | ≥ 13.4 | 11.8–13.3 | < 11.8 | 31 |
| | LV GCS | ≥ 14.8 | 12.8–14.7 | < 12.8 | 48 | ≥ 14.4 | 13.3–14.3 | < 13.3 | 29 |
| | LV GRS | ≥ 21.7 | 18.0–21.6 | < 18.0 | 48 | ≥ 20.8 | 18.7–20.7 | < 18.7 | 29 |
| | RV GLS | ≥ 18.0 | 14.2–17.9 | < 14.2 | 49 | ≥ 17.2 | 14.1–17.1 | < 14.1 | 31 |
| | RV GCS | ≥ 10.4 | 8.9–10.3 | < 8.9 | 48 | ≥ 9.6 | 7.9–9.5 | < 7.9 | 32 |
| | RV GRS | ≥ 14.7 | 12.4–14.6 | < 12.4 | 48 | ≥ 14.0 | 11.4–13.9 | < 11.4 | 32 |

**Footnote**: All strain measurements were derived using cvi42 software (Circle Cardiovascular Imaging Inc.) with standardized feature-tracking analysis. Global longitudinal strain (GLS), global circumferential strain (GCS), and global radial strain (GRS) are presented as absolute values (%). Reference ranges are defined as the interval between the 2.5th and 97.5th centiles derived from age- and sex-specific generalized additive models for location, scale and shape (GAMLSS) using the Box–Cox Cole–Green distribution. Centile-based categories beyond the reference range are derived from LMS-based thresholds (P0.5 and P2.5) and describe the position of a measurement within the reference distribution rather than disease severity or diagnostic abnormality. For clarity, only the lower reference threshold (2.5th percentile) and corresponding lower centile cut-offs are presented in this table. Reference values are derived from a Mixed adult population.

**Abbreviations**: LV = left ventricle; RV = right ventricle; GLS = global longitudinal strain; GCS = global circumferential strain; GRS = global radial strain.

### Supplementary Table 23 Left and right ventricular global strain rate reference ranges and centile-based categories stratified by sex and age using cvi42 (Circle Cardiovascular Imaging Inc.), *in a Mixed adult population*

| | | Female | | | | Male | | | |
|---|---|---|---|---|---|---|---|---|---|
| **Age group** | Metrics ($s^{-1}$) | Reference threshold | Mild | At least moderate | n | Reference threshold | Mild | At least moderate | n |
| **50–59** | LV sysGLSR | ≥ 0.66 | 0.62–0.66 | < 0.62 | 133 | ≥ 0.61 | 0.55–0.61 | < 0.55 | 61 |
| | LV sysGCSR | ≥ 0.73 | 0.68–0.73 | < 0.68 | 124 | ≥ 0.73 | 0.66–0.73 | < 0.66 | 59 |
| | LV sysGRSR | ≥ 1.14 | 1.03–1.14 | < 1.03 | 124 | ≥ 1.08 | 0.95–1.08 | < 0.95 | 59 |
| | LV diaGLSR | ≥ 0.66 | 0.58–0.66 | < 0.58 | 133 | ≥ 0.58 | 0.52–0.58 | < 0.52 | 61 |
| | LV diaGCSR | ≥ 0.64 | 0.59–0.64 | < 0.59 | 124 | ≥ 0.54 | 0.48–0.54 | < 0.48 | 59 |
| | LV diaGRSR | ≥ 1.01 | 0.85–1.01 | < 0.85 | 124 | ≥ 0.83 | 0.70–0.83 | < 0.70 | 59 |
| | RV sysGLSR | ≥ 0.82 | 0.72–0.82 | < 0.72 | 130 | ≥ 0.83 | 0.74–0.83 | < 0.74 | 61 |
| | RV sysGCSR | ≥ 0.50 | 0.43–0.50 | < 0.43 | 130 | ≥ 0.55 | 0.49–0.55 | < 0.49 | 64 |
| | RV sysGRSR | ≥ 0.70 | 0.57–0.70 | < 0.57 | 131 | ≥ 0.77 | 0.66–0.77 | < 0.66 | 64 |
| | RV diaGLSR | ≥ 0.83 | 0.70–0.83 | < 0.70 | 134 | ≥ 0.81 | 0.72–0.81 | < 0.72 | 63 |
| | RV diaGCSR | ≥ 0.49 | 0.43–0.49 | < 0.43 | 126 | ≥ 0.45 | 0.39–0.45 | < 0.39 | 61 |
| | RV diaGRSR | ≥ 0.75 | 0.60–0.75 | < 0.60 | 125 | ≥ 0.79 | 0.69–0.79 | < 0.69 | 61 |
| **60–69** | LV sysGLSR | ≥ 0.65 | 0.60–0.65 | < 0.60 | 107 | ≥ 0.63 | 0.57–0.63 | < 0.57 | 53 |
| | LV sysGCSR | ≥ 0.70 | 0.65–0.70 | < 0.65 | 103 | ≥ 0.70 | 0.64–0.70 | < 0.64 | 54 |
| | LV sysGRSR | ≥ 1.09 | 0.98–1.09 | < 0.98 | 103 | ≥ 1.02 | 0.90–1.02 | < 0.90 | 53 |
| | LV diaGLSR | ≥ 0.63 | 0.54–0.63 | < 0.54 | 107 | ≥ 0.60 | 0.53–0.60 | < 0.53 | 53 |
| | LV diaGCSR | ≥ 0.62 | 0.56–0.62 | < 0.56 | 103 | ≥ 0.54 | 0.48–0.54 | < 0.48 | 54 |
| | LV diaGRSR | ≥ 0.90 | 0.75–0.90 | < 0.75 | 103 | ≥ 0.77 | 0.65–0.77 | < 0.65 | 52 |
| | RV sysGLSR | ≥ 0.87 | 0.79–0.87 | < 0.79 | 102 | ≥ 0.86 | 0.77–0.86 | < 0.77 | 54 |
| | RV sysGCSR | ≥ 0.51 | 0.44–0.51 | < 0.44 | 106 | ≥ 0.57 | 0.50–0.57 | < 0.50 | 61 |
| | RV sysGRSR | ≥ 0.75 | 0.64–0.75 | < 0.64 | 107 | ≥ 0.80 | 0.69–0.80 | < 0.69 | 61 |
| | RV diaGLSR | ≥ 0.89 | 0.75–0.89 | < 0.75 | 108 | ≥ 0.80 | 0.71–0.80 | < 0.71 | 57 |
| | RV diaGCSR | ≥ 0.46 | 0.41–0.46 | < 0.41 | 101 | ≥ 0.45 | 0.39–0.45 | < 0.39 | 55 |
| | RV diaGRSR | ≥ 0.72 | 0.60–0.72 | < 0.60 | 100 | ≥ 0.76 | 0.66–0.76 | < 0.66 | 50 |
| **70+** | LV sysGLSR | ≥ 0.60 | 0.55–0.60 | < 0.55 | 43 | ≥ 0.62 | 0.57–0.62 | < 0.57 | 31 |
| | LV sysGCSR | ≥ 0.63 | 0.56–0.63 | < 0.56 | 48 | ≥ 0.67 | 0.61–0.67 | < 0.61 | 29 |
| | LV sysGRSR | ≥ 0.95 | 0.78–0.95 | < 0.78 | 48 | ≥ 0.98 | 0.86–0.98 | < 0.86 | 29 |
| | LV diaGLSR | ≥ 0.58 | 0.48–0.58 | < 0.48 | 43 | ≥ 0.63 | 0.57–0.63 | < 0.57 | 31 |
| | LV diaGCSR | ≥ 0.61 | 0.53–0.61 | < 0.53 | 48 | ≥ 0.56 | 0.50–0.56 | < 0.50 | 29 |
| | LV diaGRSR | ≥ 0.78 | 0.65–0.78 | < 0.65 | 48 | ≥ 0.69 | 0.59–0.69 | < 0.59 | 28 |
| | RV sysGLSR | ≥ 0.93 | 0.86–0.93 | < 0.86 | 45 | ≥ 0.85 | 0.76–0.85 | < 0.76 | 31 |
| | RV sysGCSR | ≥ 0.51 | 0.45–0.51 | < 0.45 | 48 | ≥ 0.58 | 0.51–0.58 | < 0.51 | 32 |
| | RV sysGRSR | ≥ 0.72 | 0.60–0.72 | < 0.60 | 48 | ≥ 0.82 | 0.71–0.82 | < 0.71 | 32 |
| | RV diaGLSR | ≥ 1.06 | 0.93–1.06 | < 0.93 | 49 | ≥ 0.83 | 0.74–0.83 | < 0.74 | 31 |
| | RV diaGCSR | ≥ 0.43 | 0.38–0.43 | < 0.38 | 46 | ≥ 0.41 | 0.36–0.41 | < 0.36 | 29 |
| | RV diaGRSR | ≥ 0.68 | 0.59–0.68 | < 0.59 | 45 | ≥ 0.71 | 0.62–0.71 | < 0.62 | 29 |

**Footnote**: All strain rate measurements were derived using cvi42 software (Circle Cardiovascular Imaging Inc.) with standardized feature-tracking analysis. Systolic and diastolic global longitudinal strain rate (GLSR), global circumferential strain rate (GCSR), and global radial strain rate (GRSR) are presented in $s^{-1}$. Reference ranges are defined as the interval between the 2.5th and 97.5th centiles derived from age- and sex-specific generalized additive models for location, scale and shape (GAMLSS) using the Box–Cox Cole–Green distribution. Centile-based categories beyond the reference range are derived from LMS-based thresholds (P0.5 and P2.5) and describe the position of a measurement within the reference distribution rather than disease severity or diagnostic abnormality. For clarity, only the lower reference threshold (2.5th percentile) and corresponding lower centile cut-offs are presented in this table. Reference values are derived from a Mixed adult population.
**Abbreviations**: LV = left ventricle; RV = right ventricle; GLSR = global longitudinal strain rate; GCSR = global circumferential strain rate; GRSR = global radial strain rate; sys = systolic; dia = diastolic.

**Supplementary Table 24 Left ventricular global displacement reference ranges and centile-based categories stratified by sex and age using cvi42 (Circle Cardiovascular Imaging Inc.), *in a Mixed adult population***

| | | Female | | | | Male | | | |
|---|---|---|---|---|---|---|---|---|---|
| **Age group** | Metrics (mm) | Reference threshold | Mild | At least moderate | n | Reference threshold | Mild | At least moderate | n |
| **50–59** | LV GLD | ≥ 3.1 | 2.5–3.0 | < 2.5 | 133 | ≥ 2.8 | 2.1–2.7 | < 2.1 | 61 |
| | LV GCD | ≥ 0.7 | 0.5–0.6 | < 0.5 | 96 | ≥ 0.8 | 0.6–0.7 | < 0.6 | 53 |
| | LV GRD | ≥ 4.1 | 3.9–4.0 | < 3.9 | 124 | ≥ 4.1 | 3.8–4.0 | < 3.8 | 59 |
| **60–69** | LV GLD | ≥ 2.7 | 2.0–2.6 | < 2.0 | 107 | ≥ 2.5 | 1.9–2.4 | < 1.9 | 53 |
| | LV GCD | ≥ 0.6 | 0.4–0.5 | < 0.4 | 78 | ≥ 0.8 | 0.6–0.7 | < 0.6 | 47 |
| | LV GRD | ≥ 4.1 | 3.8–4.0 | < 3.8 | 103 | ≥ 4.1 | 3.8–4.0 | < 3.8 | 54 |
| **70+** | LV GLD | ≥ 2.2 | 1.4–2.1 | < 1.4 | 43 | ≥ 2.7 | 2.0–2.6 | < 2.0 | 31 |
| | LV GCD | ≥ 0.6 | 0.4–0.5 | < 0.4 | 36 | ≥ 0.8 | 0.6–0.7 | < 0.6 | 26 |
| | LV GRD | ≥ 3.8 | 3.4–3.7 | < 3.4 | 48 | ≥ 3.9 | 3.7–3.8 | < 3.7 | 29 |

**Footnote**: All displacement measurements were derived using cvi42 software (Circle Cardiovascular Imaging Inc.) with standardized feature-tracking analysis. Global longitudinal displacement (GLD), global circumferential displacement (GCD), and global radial displacement (GRD) are presented in millimetres (mm). Reference ranges are defined as the interval between the 2.5th and 97.5th centiles derived from age- and sex-specific generalized additive models for location, scale and shape (GAMLSS) using the Box–Cox Cole–Green distribution. Centile-based categories beyond the reference range are derived from LMS-based thresholds (P0.5 and P2.5) and describe the position of a measurement within the reference distribution rather than disease severity or diagnostic abnormality. For clarity, only the lower reference threshold (2.5th percentile) and corresponding lower centile cut-offs are presented in this table. Reference values are derived from a Mixed adult population.
**Abbreviations**: LV = left ventricle; GLD = global longitudinal displacement; GCD = global circumferential displacement; GRD = global radial displacement.

### Supplementary Table 25 Left ventricular global velocity reference ranges and centile-based categories stratified by sex and age using cvi42 (Circle Cardiovascular Imaging Inc.), *in a Mixed adult population*

| | | Female | | | | Male | | | |
|---|---|---|---|---|---|---|---|---|---|
| **Age group** | Metrics (%/s) | Reference threshold | Mild | At least moderate | n | Reference threshold | Mild | At least moderate | n |
| **50–59** | LV sysGLV | ≥ 16.3 | 14.2–16.2 | < 14.2 | 133 | ≥ 16.1 | 14.2–16.0 | < 14.2 | 61 |
| | LV sysGCV | ≥ 12.1 | 9.5–12.0 | < 9.5 | 111 | ≥ 13.1 | 9.9–13.0 | < 9.9 | 54 |
| | LV sysGRV | ≥ 20.3 | 19.0–20.2 | < 19.0 | 124 | ≥ 21.2 | 19.4–21.1 | < 19.4 | 59 |
| | LV diaGLV | ≥ 17.6 | 14.4–17.5 | < 14.4 | 133 | ≥ 16.0 | 13.2–15.9 | < 13.2 | 61 |
| | LV diaGCV | ≥ 5.8 | 4.4–5.7 | < 4.4 | 73 | ≥ 5.5 | 4.4–5.4 | < 4.4 | 44 |
| | LV diaGRV | ≥ 17.8 | 15.9–17.7 | < 15.9 | 124 | ≥ 16.6 | 15.0–16.5 | < 15.0 | 59 |
| **60–69** | LV sysGLV | ≥ 15.5 | 13.5–15.4 | < 13.5 | 107 | ≥ 15.8 | 13.9–15.7 | < 13.9 | 53 |
| | LV sysGCV | ≥ 10.7 | 8.3–10.6 | < 8.3 | 78 | ≥ 12.1 | 9.1–12.0 | < 9.1 | 46 |
| | LV sysGRV | ≥ 19.2 | 17.9–19.1 | < 17.9 | 103 | ≥ 20.2 | 18.5–20.1 | < 18.5 | 54 |
| | LV diaGLV | ≥ 16.2 | 12.8–16.1 | < 12.8 | 107 | ≥ 15.1 | 12.5–15.0 | < 12.5 | 53 |
| | LV diaGCV | ≥ 6.0 | 4.8–5.9 | < 4.8 | 65 | ≥ 6.0 | 4.8–5.9 | < 4.8 | 50 |
| | LV diaGRV | ≥ 17.2 | 15.6–17.1 | < 15.6 | 103 | ≥ 16.2 | 14.6–16.1 | < 14.6 | 54 |
| **70+** | LV sysGLV | ≥ 14.6 | 12.8–14.5 | < 12.8 | 43 | ≥ 16.4 | 14.4–16.3 | < 14.4 | 30 |
| | LV sysGCV | ≥ 9.3 | 7.2–9.2 | < 7.2 | 32 | ≥ 12.8 | 9.7–12.7 | < 9.7 | 23 |
| | LV sysGRV | ≥ 17.7 | 16.3–17.6 | < 16.3 | 48 | ≥ 18.9 | 17.3–18.8 | < 17.3 | 29 |
| | LV diaGLV | ≥ 12.0 | 7.5–11.9 | < 7.5 | 43 | ≥ 16.8 | 13.8–16.7 | < 13.8 | 31 |
| | LV diaGCV | ≥ 6.1 | 5.1–6.0 | < 5.1 | 27 | ≥ 6.1 | 4.9–6.0 | < 4.9 | 23 |
| | LV diaGRV | ≥ 16.5 | 15.0–16.4 | < 15.0 | 48 | ≥ 16.1 | 14.5–16.0 | < 14.5 | 29 |

**Footnote**: All velocity measurements were derived using cvi42 software (Circle Cardiovascular Imaging Inc.) with standardized feature-tracking analysis. Systolic and diastolic global longitudinal velocity (GLV), global circumferential velocity (GCV), and global radial velocity (GRV) are presented in %/s. Reference ranges are defined as the interval between the 2.5th and 97.5th centiles derived from age- and sex-specific generalized additive models for location, scale and shape (GAMLSS) using the Box–Cox Cole–Green distribution. Centile-based categories beyond the reference range are derived from LMS-based thresholds (P0.5 and P2.5) and describe the position of a measurement within the reference distribution rather than disease severity or diagnostic abnormality. For clarity, only the lower reference threshold (2.5th percentile) and corresponding lower centile cut-offs are presented in this table. Reference values are derived from a Mixed adult population.
**Abbreviations**: LV = left ventricle; GLV = global longitudinal velocity; GCV = global circumferential velocity; GRV = global radial velocity; sys = systolic; dia = diastolic.

### Supplementary Table 26 Left and right ventricular global strain reference ranges and centile-based categories, by age group using suiteHEART, *in a White adult population*

| | | Female | | | | Male | | | |
|---|---|---|---|---|---|---|---|---|---|
| **Age group** | Metrics (%) | Reference threshold | Mild | At least moderate | n | Reference threshold | Mild | At least moderate | n |
| **18–29** | LV GLS | ≥ 15.9 | 14.7–15.8 | < 14.7 | 78 | ≥ 14.5 | 13.6–14.4 | < 13.6 | 92 |
| | LV GCS | ≥ 14.2 | 13.0–14.1 | < 13.0 | 103 | ≥ 13.1 | 12.1–13.0 | < 12.1 | 104 |
| | LV GRS | ≥ 68.6 | 62.5–68.5 | < 62.5 | 103 | ≥ 54.6 | 49.0–54.5 | < 49.0 | 104 |
| | RV GLS | ≥ 17.6 | 15.1–17.5 | < 15.1 | 83 | ≥ 15.6 | 13.3–15.5 | < 13.3 | 93 |
| | RV GCS | ≥ 6.8 | 5.1–6.7 | < 5.1 | 92 | ≥ 7.7 | 5.7–7.6 | < 5.7 | 100 |
| | RV GRS | ≥ 94.8 | 80.2–94.7 | < 80.2 | 96 | ≥ 75.0 | 63.3–74.9 | < 63.3 | 103 |
| **30–39** | LV GLS | ≥ 16.6 | 15.3–16.5 | < 15.3 | 121 | ≥ 15.0 | 13.9–14.9 | < 13.9 | 116 |
| | LV GCS | ≥ 13.7 | 12.3–13.6 | < 12.3 | 133 | ≥ 12.9 | 11.8–12.8 | < 11.8 | 131 |
| | LV GRS | ≥ 64.4 | 58.2–64.3 | < 58.2 | 132 | ≥ 55.6 | 49.5–55.5 | < 49.5 | 131 |
| | RV GLS | ≥ 18.3 | 15.6–18.2 | < 15.6 | 121 | ≥ 16.2 | 13.6–16.1 | < 13.6 | 116 |
| | RV GCS | ≥ 7.2 | 5.6–7.1 | < 5.6 | 123 | ≥ 6.9 | 4.8–6.8 | < 4.8 | 127 |
| | RV GRS | ≥ 85.9 | 71.2–85.8 | < 71.2 | 129 | ≥ 74.5 | 62.1–74.4 | < 62.1 | 130 |
| **40–49** | LV GLS | ≥ 17.4 | 16.1–17.3 | < 16.1 | 487 | ≥ 15.3 | 14.2–15.2 | < 14.2 | 404 |
| | LV GCS | ≥ 11.0 | 9.0–10.9 | < 9.0 | 518 | ≥ 11.6 | 10.3–11.5 | < 10.3 | 439 |
| | LV GRS | ≥ 66.6 | 59.7–66.5 | < 59.7 | 518 | ≥ 56.4 | 49.9–56.3 | < 49.9 | 439 |
| | RV GLS | ≥ 18.9 | 15.9–18.8 | < 15.9 | 476 | ≥ 16.7 | 13.9–16.6 | < 13.9 | 388 |
| | RV GCS | ≥ 6.1 | 4.2–6.0 | < 4.2 | 451 | ≥ 6.2 | 4.0–6.1 | < 4.0 | 390 |
| | RV GRS | ≥ 102.2 | 82.7–102.1 | < 82.7 | 506 | ≥ 88.0 | 72.4–87.9 | < 72.4 | 426 |
| **50–59** | LV GLS | ≥ 17.7 | 16.4–17.6 | < 16.4 | 816 | ≥ 15.6 | 14.3–15.5 | < 14.3 | 729 |
| | LV GCS | ≥ 9.3 | 6.9–9.2 | < 6.9 | 909 | ≥ 10.5 | 9.0–10.4 | < 9.0 | 821 |
| | LV GRS | ≥ 70.1 | 62.3–70.0 | < 62.3 | 909 | ≥ 57.1 | 50.1–57.0 | < 50.1 | 820 |
| | RV GLS | ≥ 19.2 | 16.0–19.1 | < 16.0 | 837 | ≥ 17.2 | 14.1–17.1 | < 14.1 | 669 |
| | RV GCS | ≥ 5.5 | 3.6–5.4 | < 3.6 | 794 | ≥ 5.6 | 3.5–5.5 | < 3.5 | 728 |
| | RV GRS | ≥ 142.9 | 113.2–142.8 | < 113.2 | 893 | ≥ 114.0 | 93.1–113.9 | < 93.1 | 800 |
| **60–69** | LV GLS | ≥ 17.6 | 16.3–17.5 | < 16.3 | 812 | ≥ 15.6 | 14.3–15.5 | < 14.3 | 687 |
| | LV GCS | ≥ 9.9 | 7.7–9.8 | < 7.7 | 920 | ≥ 10.7 | 9.0–10.6 | < 9.0 | 807 |
| | LV GRS | ≥ 69.1 | 60.7–69.0 | < 60.7 | 918 | ≥ 57.6 | 50.1–57.5 | < 50.1 | 806 |
| | RV GLS | ≥ 19.2 | 15.8–19.1 | < 15.8 | 802 | ≥ 17.5 | 14.1–17.4 | < 14.1 | 610 |
| | RV GCS | ≥ 5.0 | 3.1–4.9 | < 3.1 | 768 | ≥ 5.3 | 3.3–5.2 | < 3.3 | 721 |
| | RV GRS | ≥ 145.5 | 114.5–145.4 | < 114.5 | 879 | ≥ 117.3 | 96.0–117.2 | < 96.0 | 788 |
| **70+** | LV GLS | ≥ 17.5 | 16.2–17.4 | < 16.2 | 936 | ≥ 15.6 | 14.1–15.5 | < 14.1 | 1038 |
| | LV GCS | ≥ 10.7 | 8.7–10.6 | < 8.7 | 1099 | ≥ 10.7 | 9.0–10.6 | < 9.0 | 1246 |
| | LV GRS | ≥ 70.5 | 60.9–70.4 | < 60.9 | 1099 | ≥ 58.0 | 49.8–57.9 | < 49.8 | 1246 |
| | RV GLS | ≥ 19.1 | 15.5–19.0 | < 15.5 | 927 | ≥ 17.6 | 13.9–17.5 | < 13.9 | 900 |
| | RV GCS | ≥ 4.8 | 2.8–4.7 | < 2.8 | 917 | ≥ 5.3 | 3.3–5.2 | < 3.3 | 1099 |
| | RV GRS | ≥ 156.2 | 124.0–156.1 | < 124.0 | 1049 | ≥ 128.2 | 105.8–128.1 | < 105.8 | 1207 |

**Footnote**: All strain measurements were derived using suiteHEART software with standardized feature-tracking analysis. Global longitudinal strain (GLS), global circumferential strain (GCS), and global radial strain (GRS) are presented as absolute values (%). Reference ranges are defined as the interval between the 2.5th and 97.5th centiles derived from age- and sex-specific generalized additive models for location, scale and shape (GAMLSS) using the Box–Cox Cole–Green distribution. Centile-based categories beyond the reference range are derived from LMS-based thresholds (P0.5 and P2.5) and describe the position of a measurement within the reference distribution rather than disease severity or diagnostic abnormality. For clarity, only the lower reference threshold (2.5th percentile) and corresponding lower centile cut-offs are presented in this table. Reference values are derived from a White adult population.

**Abbreviations**: LV = left ventricle; RV = right ventricle; GLS = global longitudinal strain; GCS = global circumferential strain; GRS = global radial strain.

### Supplementary Table 27 Left and right atrial global strain reference ranges and centile-based categories, by age group using suiteHEART, *in a White adult population*

| | | Female | | | | Male | | | |
|---|---|---|---|---|---|---|---|---|---|
| **Age group** | Metrics (%) | Reference threshold | Mild | At least moderate | n | Reference threshold | Mild | At least moderate | n |
| **18–29** | LA GLS | ≥ 51.8 | 42.6–51.7 | < 42.6 | 57 | ≥ 45.1 | 37.3–45.0 | < 37.3 | 64 |
| | RA GLS | ≥ 52.3 | 44.8–52.2 | < 44.8 | 76 | ≥ 39.1 | 31.3–39.0 | < 31.3 | 81 |
| **30–39** | LA GLS | ≥ 52.4 | 42.8–52.3 | < 42.8 | 85 | ≥ 46.1 | 37.3–46.0 | < 37.3 | 87 |
| | RA GLS | ≥ 52.8 | 44.4–52.7 | < 44.4 | 107 | ≥ 38.4 | 30.4–38.3 | < 30.4 | 111 |
| **40–49** | LA GLS | ≥ 52.7 | 42.7–52.6 | < 42.7 | 445 | ≥ 46.7 | 37.2–46.6 | < 37.2 | 361 |
| | RA GLS | ≥ 52.9 | 43.7–52.8 | < 43.7 | 473 | ≥ 39.0 | 30.6–38.9 | < 30.6 | 393 |
| **50–59** | LA GLS | ≥ 51.2 | 41.1–51.1 | < 41.1 | 804 | ≥ 45.3 | 35.1–45.2 | < 35.1 | 675 |
| | RA GLS | ≥ 52.6 | 42.5–52.5 | < 42.5 | 798 | ≥ 40.8 | 31.7–40.7 | < 31.7 | 678 |
| **60–69** | LA GLS | ≥ 46.9 | 37.1–46.8 | < 37.1 | 770 | ≥ 40.9 | 30.7–40.8 | < 30.7 | 628 |
| | RA GLS | ≥ 51.9 | 40.8–51.8 | < 40.8 | 735 | ≥ 41.5 | 32.0–41.4 | < 32.0 | 580 |
| **70+** | LA GLS | ≥ 39.2 | 30.3–39.1 | < 30.3 | 864 | ≥ 34.0 | 24.4–33.9 | < 24.4 | 911 |
| | RA GLS | ≥ 50.5 | 37.9–50.4 | < 37.9 | 810 | ≥ 40.1 | 30.4–40.0 | < 30.4 | 780 |

**Footnote**: All strain measurements were derived using suiteHEART software with standardized feature-tracking analysis. Left atrial global longitudinal strain (LA GLS) and right atrial global longitudinal strain (RA GLS) are presented as absolute values (%). Reference ranges are defined as the interval between the 2.5th and 97.5th centiles derived from age- and sex-specific generalized additive models for location, scale and shape (GAMLSS) using the Box–Cox Cole–Green distribution. Centile-based categories beyond the reference range are derived from LMS-based thresholds (P0.5 and P2.5) and describe the position of a measurement within the reference distribution rather than disease severity or diagnostic abnormality. For clarity, only the lower reference threshold (2.5th percentile) and corresponding lower centile cut-offs are presented in this table. Reference values are derived from a White adult population.
**Abbreviations**: LA = left atrium; RA = right atrium; GLS = global longitudinal strain.

### Supplementary Table 28 Left and right ventricular global strain reference ranges and centile-based categories, by age group using suiteHEART, *in a Black adult population*

| | | Female | | | | Male | | | |
|---|---|---|---|---|---|---|---|---|---|
| **Age group** | Metrics (%) | Reference threshold | Mild | At least moderate | n | Reference threshold | Mild | At least moderate | n |
| **40–49** | LV GLS | ≥ 18.3 | 17.2–18.2 | < 17.2 | 10 | | | | |
| | LV GCS | ≥ 8.1 | 5.0–8.0 | < 5.0 | 10 | | | | |
| | LV GRS | ≥ 69.7 | 62.6–69.6 | < 62.6 | 10 | | | | |
| | RV GLS | | | | | | | | |
| | RV GCS | | | | | | | | |
| | RV GRS | ≥ 148.9 | 104.3–148.8 | < 104.3 | 10 | | | | |
| **50–59** | LV GLS | ≥ 17.8 | 16.7–17.7 | < 16.7 | 84 | ≥ 15.2 | 13.8–15.1 | < 13.8 | 64 |
| | LV GCS | ≥ 9.0 | 6.6–8.9 | < 6.6 | 94 | ≥ 10.4 | 9.1–10.3 | < 9.1 | 76 |
| | LV GRS | ≥ 69.1 | 61.6–69.0 | < 61.6 | 94 | ≥ 61.7 | 55.7–61.6 | < 55.7 | 76 |
| | RV GLS | ≥ 19.2 | 16.6–19.1 | < 16.6 | 82 | ≥ 15.2 | 11.3–15.1 | < 11.3 | 60 |
| | RV GCS | ≥ 4.6 | 3.0–4.5 | < 3.0 | 68 | ≥ 5.2 | 2.9–5.1 | < 2.9 | 61 |
| | RV GRS | ≥ 160.3 | 121.5–160.2 | < 121.5 | 92 | ≥ 147.9 | 127.8–147.8 | < 127.8 | 73 |
| **60–69** | LV GLS | ≥ 17.0 | 16.0–16.9 | < 16.0 | 53 | ≥ 14.9 | 13.5–14.8 | < 13.5 | 43 |
| | LV GCS | ≥ 10.2 | 8.6–10.1 | < 8.6 | 65 | ≥ 7.9 | 5.3–7.8 | < 5.3 | 47 |
| | LV GRS | ≥ 68.0 | 59.7–67.9 | < 59.7 | 65 | ≥ 56.6 | 47.8–56.5 | < 47.8 | 47 |
| | RV GLS | ≥ 17.9 | 14.5–17.8 | < 14.5 | 55 | ≥ 12.6 | 7.8–12.5 | < 7.8 | 37 |
| | RV GCS | ≥ 4.6 | 3.1–4.5 | < 3.1 | 45 | ≥ 5.8 | 4.3–5.7 | < 4.3 | 38 |
| | RV GRS | ≥ 174.1 | 141.5–174.0 | < 141.5 | 58 | ≥ 143.1 | 116.4–143.0 | < 116.4 | 44 |
| **70+** | LV GLS | ≥ 16.1 | 15.2–16.0 | < 15.2 | 18 | ≥ 14.6 | 13.2–14.5 | < 13.2 | 12 |
| | LV GCS | ≥ 11.1 | 9.9–11.0 | < 9.9 | 18 | ≥ 10.2 | 7.9–10.1 | < 7.9 | 14 |
| | LV GRS | ≥ 66.7 | 57.4–66.6 | < 57.4 | 18 | ≥ 48.6 | 34.2–48.5 | < 34.2 | 14 |
| | RV GLS | ≥ 16.1 | 11.4–16.0 | < 11.4 | 16 | ≥ 15.4 | 10.9–15.3 | < 10.9 | 10 |
| | RV GCS | ≥ 4.6 | 3.1–4.5 | < 3.1 | 15 | ≥ 6.1 | 5.0–6.0 | < 5.0 | 14 |
| | RV GRS | ≥ 184.8 | 156.5–184.7 | < 156.5 | 17 | ≥ 134.8 | 98.2–134.7 | < 98.2 | 14 |

**Footnote**: All strain measurements were derived using suiteHEART software with standardized feature-tracking analysis. Global longitudinal strain (GLS), global circumferential strain (GCS), and global radial strain (GRS) are presented as absolute values (%). Reference ranges are defined as the interval between the 2.5th and 97.5th centiles derived from age- and sex-specific generalized additive models for location, scale and shape (GAMLSS) using the Box–Cox Cole–Green distribution. Centile-based categories beyond the reference range are derived from LMS-based thresholds (P0.5 and P2.5) and describe the position of a measurement within the reference distribution rather than disease severity or diagnostic abnormality. For clarity, only the lower reference threshold (2.5th percentile) and corresponding lower centile cut-offs are presented in this table. Reference values are derived from a Black adult population.
**Abbreviations**: LV = left ventricle; RV = right ventricle; GLS = global longitudinal strain; GCS = global circumferential strain; GRS = global radial strain.

### Supplementary Table 29 Left and right atrial global strain reference ranges and centile-based categories, by age group using suiteHEART, *in a Black adult population*

| | | Female | | | | Male | | | |
|---|---|---|---|---|---|---|---|---|---|
| **Age group** | Metrics (%) | Reference threshold | Mild | At least moderate | n | Reference threshold | Mild | At least moderate | n |
| **40–49** | LA GLS | | | | | | | | |
| | RA GLS | ≥ 38.1 | 27.1–38.0 | < 27.1 | 10 | | | | |
| **50–59** | LA GLS | ≥ 55.8 | 48.3–55.7 | < 48.3 | 88 | ≥ 46.3 | 36.7–46.2 | < 36.7 | 71 |
| | RA GLS | ≥ 43.6 | 34.3–43.5 | < 34.3 | 85 | ≥ 46.2 | 40.0–46.1 | < 40.0 | 68 |
| **60–69** | LA GLS | ≥ 47.4 | 39.8–47.3 | < 39.8 | 58 | ≥ 43.5 | 34.4–43.4 | < 34.4 | 43 |
| | RA GLS | ≥ 50.2 | 42.6–50.1 | < 42.6 | 55 | ≥ 39.4 | 30.9–39.3 | < 30.9 | 39 |
| **70+** | LA GLS | ≥ 39.1 | 31.5–39.0 | < 31.5 | 17 | ≥ 61.6 | 48.7–61.5 | < 48.7 | 11 |
| | RA GLS | ≥ 55.1 | 48.5–55.0 | < 48.5 | 15 | ≥ 30.4 | 18.0–30.3 | < 18.0 | 10 |

**Footnote**: All strain measurements were derived using suiteHEART software with standardized feature-tracking analysis. Left atrial global longitudinal strain (LA GLS) and right atrial global longitudinal strain (RA GLS) are presented as absolute values (%). Reference ranges are defined as the interval between the 2.5th and 97.5th centiles derived from age- and sex-specific generalized additive models for location, scale and shape (GAMLSS) using the Box–Cox Cole–Green distribution. Centile-based categories beyond the reference range are derived from LMS-based thresholds (P0.5 and P2.5) and describe the position of a measurement within the reference distribution rather than disease severity or diagnostic abnormality. For clarity, only the lower reference threshold (2.5th percentile) and corresponding lower centile cut-offs are presented in this table. Reference values are derived from a Black adult population.
**Abbreviations**: LA = left atrium; RA = right atrium; GLS = global longitudinal strain.

**Supplementary Table 30 Left and right ventricular global strain reference ranges and centile-based categories, by age group using suiteHEART, *in a South Asian adult population***

| | | Female | | | | Male | | | |
|---|---|---|---|---|---|---|---|---|---|
| **Age group** | Metrics (%) | Reference threshold | Mild | At least moderate | n | Reference threshold | Mild | At least moderate | n |
| **40–49** | LV GLS | ≥ 17.5 | 14.8–17.4 | < 14.8 | 10 | ≥ 17.4 | 16.1–17.3 | < 16.1 | 20 |
| | LV GCS | | | | | ≥ 10.1 | 8.8–10.0 | < 8.8 | 20 |
| | LV GRS | | | | | ≥ 60.9 | 52.5–60.8 | < 52.5 | 20 |
| | RV GLS | ≥ 18.3 | 14.3–18.2 | < 14.3 | 10 | ≥ 16.6 | 13.3–16.5 | < 13.3 | 19 |
| | RV GCS | | | | | ≥ 4.8 | 3.2–4.7 | < 3.2 | 19 |
| | RV GRS | | | | | ≥ 147.1 | 126.5–147.0 | < 126.5 | 20 |
| **50–59** | LV GLS | ≥ 18.0 | 16.3–17.9 | < 16.3 | 76 | ≥ 16.7 | 15.3–16.6 | < 15.3 | 124 |
| | LV GCS | ≥ 8.3 | 5.4–8.2 | < 5.4 | 81 | ≥ 10.3 | 9.1–10.2 | < 9.1 | 132 |
| | LV GRS | ≥ 71.9 | 63.3–71.8 | < 63.3 | 81 | ≥ 61.1 | 52.6–61.0 | < 52.6 | 132 |
| | RV GLS | ≥ 18.9 | 15.4–18.8 | < 15.4 | 75 | ≥ 17.4 | 14.3–17.3 | < 14.3 | 122 |
| | RV GCS | ≥ 4.5 | 2.4–4.4 | < 2.4 | 68 | ≥ 4.6 | 3.0–4.5 | < 3.0 | 103 |
| | RV GRS | ≥ 184.8 | 158.1–184.7 | < 158.1 | 77 | ≥ 150.0 | 129.2–149.9 | < 129.2 | 130 |
| **60–69** | LV GLS | ≥ 18.6 | 17.5–18.5 | < 17.5 | 52 | ≥ 16.2 | 14.6–16.1 | < 14.6 | 102 |
| | LV GCS | ≥ 9.3 | 7.0–9.2 | < 7.0 | 62 | ≥ 10.6 | 9.4–10.5 | < 9.4 | 118 |
| | LV GRS | ≥ 71.5 | 61.4–71.4 | < 61.4 | 62 | ≥ 62.5 | 53.8–62.4 | < 53.8 | 117 |
| | RV GLS | ≥ 19.6 | 16.6–19.5 | < 16.6 | 53 | ≥ 18.3 | 15.5–18.2 | < 15.5 | 105 |
| | RV GCS | ≥ 4.7 | 2.6–4.6 | < 2.6 | 46 | ≥ 4.4 | 2.7–4.3 | < 2.7 | 103 |
| | RV GRS | ≥ 177.6 | 149.2–177.5 | < 149.2 | 57 | ≥ 153.6 | 132.6–153.5 | < 132.6 | 118 |
| **70+** | LV GLS | ≥ 19.3 | 18.5–19.2 | < 18.5 | 24 | ≥ 16.1 | 14.2–16.0 | < 14.2 | 34 |
| | LV GCS | ≥ 10.3 | 8.4–10.2 | < 8.4 | 31 | ≥ 10.9 | 9.7–10.8 | < 9.7 | 39 |
| | LV GRS | ≥ 70.6 | 58.3–70.5 | < 58.3 | 31 | ≥ 65.9 | 56.8–65.8 | < 56.8 | 39 |
| | RV GLS | ≥ 20.3 | 17.7–20.2 | < 17.7 | 25 | ≥ 19.3 | 16.7–19.2 | < 16.7 | 32 |
| | RV GCS | ≥ 5.0 | 2.8–4.9 | < 2.8 | 22 | ≥ 4.3 | 2.6–4.2 | < 2.6 | 35 |
| | RV GRS | ≥ 169.7 | 139.0–169.6 | < 139.0 | 29 | ≥ 157.7 | 136.3–157.6 | < 136.3 | 38 |

**Footnote**: All strain measurements were derived using suiteHEART software with standardized feature-tracking analysis. Global longitudinal strain (GLS), global circumferential strain (GCS), and global radial strain (GRS) are presented as absolute values (%). Reference ranges are defined as the interval between the 2.5th and 97.5th centiles derived from age- and sex-specific generalized additive models for location, scale and shape (GAMLSS) using the Box–Cox Cole–Green distribution. Centile-based categories beyond the reference range are derived from LMS-based thresholds (P0.5 and P2.5) and describe the position of a measurement within the reference distribution rather than disease severity or diagnostic abnormality. For clarity, only the lower reference threshold (2.5th percentile) and corresponding lower centile cut-offs are presented in this table. Reference values are derived from a South Asian adult population.

**Abbreviations**: LV = left ventricle; RV = right ventricle; GLS = global longitudinal strain; GCS = global circumferential strain; GRS = global radial strain.

### Supplementary Table 31 Left and right atrial global strain reference ranges and centile-based categories, by age group using suiteHEART, *in a South Asian adult population*

| | | Female | | | | Male | | | |
|---|---|---|---|---|---|---|---|---|---|
| **Age group** | Metrics (%) | Reference threshold | Mild | At least moderate | n | Reference threshold | Mild | At least moderate | n |
| **40–49** | LA GLS | | | | | ≥ 59.7 | 48.7–59.6 | < 48.7 | 19 |
| | RA GLS | ≥ 64.3 | 56.3–64.2 | < 56.3 | 10 | ≥ 40.8 | 28.5–40.7 | < 28.5 | 19 |
| **50–59** | LA GLS | ≥ 63.4 | 54.0–63.3 | < 54.0 | 76 | ≥ 53.0 | 42.3–52.9 | < 42.3 | 120 |
| | RA GLS | ≥ 58.6 | 48.6–58.5 | < 48.6 | 75 | ≥ 42.7 | 31.0–42.6 | < 31.0 | 124 |
| **60–69** | LA GLS | ≥ 56.3 | 47.9–56.2 | < 47.9 | 54 | ≥ 44.9 | 34.7–44.8 | < 34.7 | 102 |
| | RA GLS | ≥ 45.3 | 29.9–45.2 | < 29.9 | 47 | ≥ 45.1 | 34.1–45.0 | < 34.1 | 103 |
| **70+** | LA GLS | ≥ 48.8 | 41.5–48.7 | < 41.5 | 23 | ≥ 36.5 | 27.0–36.4 | < 27.0 | 29 |
| | RA GLS | ≥ 25.4 | 8.6–25.3 | < 8.6 | 22 | ≥ 47.8 | 37.5–47.7 | < 37.5 | 26 |

**Footnote**: All strain measurements were derived using suiteHEART software with standardized feature-tracking analysis. Left atrial global longitudinal strain (LA GLS) and right atrial global longitudinal strain (RA GLS) are presented as absolute values (%). Reference ranges are defined as the interval between the 2.5th and 97.5th centiles derived from age- and sex-specific generalized additive models for location, scale and shape (GAMLSS) using the Box–Cox Cole–Green distribution. Centile-based categories beyond the reference range are derived from LMS-based thresholds (P0.5 and P2.5) and describe the position of a measurement within the reference distribution rather than disease severity or diagnostic abnormality. For clarity, only the lower reference threshold (2.5th percentile) and corresponding lower centile cut-offs are presented in this table. Reference values are derived from a South Asian adult population.
**Abbreviations**: LA = left atrium; RA = right atrium; GLS = global longitudinal strain.

### Supplementary Table 32 Left and right ventricular global strain reference ranges and centile-based categories, by age group using suiteHEART, *in a Chinese adult population*

| | | Female | | | | Male | | | |
|---|---|---|---|---|---|---|---|---|---|
| **Age group** | Metrics (%) | Reference threshold | Mild | At least moderate | n | Reference threshold | Mild | At least moderate | n |
| **18–29** | LV GLS | ≥ 17.4 | 16.0–17.3 | < 16.0 | 15 | ≥ 15.3 | 14.3–15.2 | < 14.3 | 19 |
| | LV GCS | ≥ 10.7 | 7.3–10.6 | < 7.3 | 16 | ≥ 13.8 | 11.9–13.7 | < 11.9 | 19 |
| | LV GRS | ≥ 73.7 | 67.7–73.6 | < 67.7 | 16 | ≥ 63.8 | 57.9–63.7 | < 57.9 | 19 |
| | RV GLS | ≥ 19.2 | 16.8–19.1 | < 16.8 | 16 | ≥ 18.3 | 16.4–18.2 | < 16.4 | 19 |
| | RV GCS | ≥ 7.8 | 6.8–7.7 | < 6.8 | 15 | ≥ 6.7 | 4.7–6.6 | < 4.7 | 16 |
| | RV GRS | ≥ 91.5 | 70.4–91.4 | < 70.4 | 15 | ≥ 51.8 | 38.9–51.7 | < 38.9 | 17 |
| **30–39** | LV GLS | ≥ 17.6 | 16.1–17.5 | < 16.1 | 17 | ≥ 15.3 | 14.2–15.2 | < 14.2 | 17 |
| | LV GCS | ≥ 9.8 | 6.6–9.7 | < 6.6 | 17 | ≥ 13.1 | 11.2–13.0 | < 11.2 | 17 |
| | LV GRS | ≥ 74.8 | 68.3–74.7 | < 68.3 | 17 | ≥ 62.0 | 56.1–61.9 | < 56.1 | 17 |
| | RV GLS | ≥ 19.2 | 16.6–19.1 | < 16.6 | 17 | ≥ 18.4 | 16.4–18.3 | < 16.4 | 17 |
| | RV GCS | ≥ 7.4 | 6.1–7.3 | < 6.1 | 13 | ≥ 6.4 | 4.5–6.3 | < 4.5 | 15 |
| | RV GRS | ≥ 81.4 | 61.0–81.3 | < 61.0 | 16 | ≥ 60.0 | 45.1–59.9 | < 45.1 | 15 |
| **40–49** | LV GLS | ≥ 17.7 | 16.3–17.6 | < 16.3 | 23 | ≥ 15.2 | 13.9–15.1 | < 13.9 | 20 |
| | LV GCS | ≥ 9.2 | 6.1–9.1 | < 6.1 | 24 | ≥ 12.4 | 10.7–12.3 | < 10.7 | 20 |
| | LV GRS | ≥ 74.9 | 67.8–74.8 | < 67.8 | 24 | ≥ 60.2 | 54.1–60.1 | < 54.1 | 20 |
| | RV GLS | ≥ 19.2 | 16.3–19.1 | < 16.3 | 24 | ≥ 18.4 | 16.3–18.3 | < 16.3 | 20 |
| | RV GCS | ≥ 6.7 | 5.2–6.6 | < 5.2 | 22 | ≥ 6.2 | 4.4–6.1 | < 4.4 | 19 |
| | RV GRS | ≥ 80.5 | 58.5–80.4 | < 58.5 | 24 | ≥ 68.7 | 51.6–68.6 | < 51.6 | 19 |
| **50–59** | LV GLS | ≥ 17.9 | 16.5–17.8 | < 16.5 | 63 | ≥ 15.0 | 13.6–14.9 | < 13.6 | 44 |
| | LV GCS | ≥ 8.8 | 5.8–8.7 | < 5.8 | 66 | ≥ 11.8 | 10.3–11.7 | < 10.3 | 49 |
| | LV GRS | ≥ 72.6 | 64.1–72.5 | < 64.1 | 66 | ≥ 58.3 | 52.1–58.2 | < 52.1 | 49 |
| | RV GLS | ≥ 19.1 | 16.0–19.0 | < 16.0 | 63 | ≥ 18.4 | 16.2–18.3 | < 16.2 | 44 |
| | RV GCS | ≥ 5.8 | 4.1–5.7 | < 4.1 | 53 | ≥ 6.0 | 4.3–5.9 | < 4.3 | 46 |
| | RV GRS | ≥ 86.0 | 60.3–85.9 | < 60.3 | 64 | ≥ 77.3 | 58.1–77.2 | < 58.1 | 48 |
| **60–69** | LV GLS | ≥ 18.1 | 16.8–18.0 | < 16.8 | 44 | ≥ 14.8 | 13.3–14.7 | < 13.3 | 35 |
| | LV GCS | ≥ 8.5 | 5.6–8.4 | < 5.6 | 51 | ≥ 11.3 | 10.0–11.2 | < 10.0 | 37 |
| | LV GRS | ≥ 66.7 | 55.7–66.6 | < 55.7 | 52 | ≥ 56.4 | 50.1–56.3 | < 50.1 | 37 |
| | RV GLS | ≥ 18.9 | 15.6–18.8 | < 15.6 | 48 | ≥ 18.4 | 16.1–18.3 | < 16.1 | 35 |
| | RV GCS | ≥ 4.6 | 2.7–4.5 | < 2.7 | 46 | ≥ 5.8 | 4.3–5.7 | < 4.3 | 31 |
| | RV GRS | ≥ 91.8 | 61.8–91.7 | < 61.8 | 51 | ≥ 85.9 | 64.6–85.8 | < 64.6 | 36 |
| **70+** | LV GLS | ≥ 18.3 | 17.0–18.2 | < 17.0 | 11 | | | | |
| | LV GCS | ≥ 8.3 | 5.5–8.2 | < 5.5 | 12 | | | | |
| | LV GRS | ≥ 58.9 | 44.3–58.8 | < 44.3 | 12 | | | | |
| | RV GLS | ≥ 18.7 | 15.2–18.6 | < 15.2 | 11 | | | | |
| | RV GCS | | | | | | | | |
| | RV GRS | ≥ 95.7 | 62.0–95.6 | < 62.0 | 12 | | | | |

**Footnote**: All strain measurements were derived using suiteHEART software with standardized feature-tracking analysis. Global longitudinal strain (GLS), global circumferential strain (GCS), and global radial strain (GRS) are presented as absolute values (%). Reference ranges are defined as the interval between the 2.5th and 97.5th centiles derived from age- and sex-specific generalized additive models for location, scale and shape (GAMLSS) using the Box–Cox Cole–Green distribution. Centile-based categories beyond the reference range are derived from LMS-based thresholds (P0.5 and P2.5) and describe the position of a measurement within the reference distribution rather than disease severity or diagnostic abnormality. For clarity, only the lower reference threshold (2.5th percentile) and corresponding lower centile cut-offs are presented in this table. Reference values are derived from a Chinese adult population.

**Abbreviations**: LV = left ventricle; RV = right ventricle; GLS = global longitudinal strain; GCS = global circumferential strain; GRS = global radial strain.

### Supplementary Table 33 Left and right atrial global strain reference ranges and centile-based categories, by age group using suiteHEART, *in a Chinese adult population*

| | | Female | | | | Male | | | |
|---|---|---|---|---|---|---|---|---|---|
| **Age group** | Metrics (%) | Reference threshold | Mild | At least moderate | n | Reference threshold | Mild | At least moderate | n |
| **18–29** | LA GLS | ≥ 67.3 | 61.5–67.2 | < 61.5 | 14 | ≥ 43.0 | 35.0–42.9 | < 35.0 | 15 |
| | RA GLS | ≥ 66.4 | 57.3–66.3 | < 57.3 | 16 | ≥ 49.3 | 44.6–49.2 | < 44.6 | 18 |
| **30–39** | LA GLS | ≥ 64.5 | 58.0–64.4 | < 58.0 | 16 | ≥ 37.5 | 30.6–37.4 | < 30.6 | 14 |
| | RA GLS | ≥ 63.5 | 54.3–63.4 | < 54.3 | 17 | ≥ 49.1 | 43.8–49.0 | < 43.8 | 16 |
| **40–49** | LA GLS | ≥ 60.8 | 53.5–60.7 | < 53.5 | 20 | ≥ 39.2 | 32.0–39.1 | < 32.0 | 17 |
| | RA GLS | ≥ 60.5 | 51.2–60.4 | < 51.2 | 22 | ≥ 48.2 | 42.1–48.1 | < 42.1 | 20 |
| **50–59** | LA GLS | ≥ 56.2 | 48.1–56.1 | < 48.1 | 59 | ≥ 50.1 | 40.9–50.0 | < 40.9 | 39 |
| | RA GLS | ≥ 57.5 | 48.0–57.4 | < 48.0 | 61 | ≥ 46.6 | 39.5–46.5 | < 39.5 | 44 |
| **60–69** | LA GLS | ≥ 50.5 | 41.4–50.4 | < 41.4 | 43 | ≥ 58.8 | 48.0–58.7 | < 48.0 | 30 |
| | RA GLS | ≥ 54.5 | 44.9–54.4 | < 44.9 | 47 | ≥ 43.9 | 35.6–43.8 | < 35.6 | 31 |
| **70+** | LA GLS | | | | | | | | |
| | RA GLS | ≥ 52.0 | 42.2–51.9 | < 42.2 | 11 | | | | |

**Footnote**: All strain measurements were derived using suiteHEART software with standardized feature-tracking analysis. Left atrial global longitudinal strain (LA GLS) and right atrial global longitudinal strain (RA GLS) are presented as absolute values (%). Reference ranges are defined as the interval between the 2.5th and 97.5th centiles derived from age- and sex-specific generalized additive models for location, scale and shape (GAMLSS) using the Box–Cox Cole–Green distribution. Centile-based categories beyond the reference range are derived from LMS-based thresholds (P0.5 and P2.5) and describe the position of a measurement within the reference distribution rather than disease severity or diagnostic abnormality. For clarity, only the lower reference threshold (2.5th percentile) and corresponding lower centile cut-offs are presented in this table. Reference values are derived from a Chinese adult population.
**Abbreviations**: LA = left atrium; RA = right atrium; GLS = global longitudinal strain.

### Supplementary Table 34 Left and right ventricular global strain reference ranges and centile-based categories, by age group using suiteHEART, *in a Mixed adult population*

| | | Female | | | | Male | | | |
|---|---|---|---|---|---|---|---|---|---|
| **Age group** | Metrics (%) | Reference threshold | Mild | At least moderate | n | Reference threshold | Mild | At least moderate | n |
| **50–59** | LV GLS | ≥ 18.4 | 17.4–18.3 | < 17.4 | 125 | ≥ 16.2 | 14.7–16.1 | < 14.7 | 62 |
| | LV GCS | ≥ 8.5 | 6.0–8.4 | < 6.0 | 136 | ≥ 10.2 | 8.3–10.1 | < 8.3 | 67 |
| | LV GRS | ≥ 74.9 | 67.6–74.8 | < 67.6 | 136 | ≥ 64.8 | 58.5–64.7 | < 58.5 | 67 |
| | RV GLS | ≥ 20.5 | 18.2–20.4 | < 18.2 | 122 | ≥ 16.2 | 12.6–16.1 | < 12.6 | 57 |
| | RV GCS | ≥ 5.0 | 3.6–4.9 | < 3.6 | 120 | | | | |
| | RV GRS | ≥ 179.5 | 151.4–179.4 | < 151.4 | 136 | ≥ 141.6 | 120.5–141.5 | < 120.5 | 65 |
| **60–69** | LV GLS | ≥ 18.2 | 17.1–18.1 | < 17.1 | 93 | ≥ 16.2 | 14.7–16.1 | < 14.7 | 53 |
| | LV GCS | ≥ 8.2 | 4.9–8.1 | < 4.9 | 110 | ≥ 10.1 | 8.2–10.0 | < 8.2 | 62 |
| | LV GRS | ≥ 71.2 | 61.6–71.1 | < 61.6 | 110 | ≥ 62.6 | 56.5–62.5 | < 56.5 | 62 |
| | RV GLS | ≥ 19.1 | 16.0–19.0 | < 16.0 | 100 | ≥ 17.0 | 13.1–16.9 | < 13.1 | 53 |
| | RV GCS | ≥ 4.7 | 3.1–4.6 | < 3.1 | 88 | | | | |
| | RV GRS | ≥ 168.5 | 137.6–168.4 | < 137.6 | 106 | ≥ 147.6 | 125.6–147.5 | < 125.6 | 59 |
| **70+** | LV GLS | ≥ 17.9 | 16.6–17.8 | < 16.6 | 41 | ≥ 16.7 | 15.1–16.6 | < 15.1 | 29 |
| | LV GCS | ≥ 10.9 | 8.9–10.8 | < 8.9 | 49 | ≥ 10.3 | 8.4–10.2 | < 8.4 | 35 |
| | LV GRS | ≥ 60.2 | 44.0–60.1 | < 44.0 | 49 | ≥ 61.6 | 55.6–61.5 | < 55.6 | 35 |
| | RV GLS | ≥ 17.0 | 12.4–16.9 | < 12.4 | 43 | ≥ 16.1 | 12.5–16.0 | < 12.5 | 30 |
| | RV GCS | ≥ 4.2 | 2.4–4.1 | < 2.4 | 42 | | | | |
| | RV GRS | ≥ 153.7 | 119.4–153.6 | < 119.4 | 45 | ≥ 146.5 | 124.7–146.4 | < 124.7 | 34 |

**Footnote**: All strain measurements were derived using suiteHEART software with standardized feature-tracking analysis. Global longitudinal strain (GLS), global circumferential strain (GCS), and global radial strain (GRS) are presented as absolute values (%). Reference ranges are defined as the interval between the 2.5th and 97.5th centiles derived from age- and sex-specific generalized additive models for location, scale and shape (GAMLSS) using the Box–Cox Cole–Green distribution. Centile-based categories beyond the reference range are derived from LMS-based thresholds (P0.5 and P2.5) and describe the position of a measurement within the reference distribution rather than disease severity or diagnostic abnormality. For clarity, only the lower reference threshold (2.5th percentile) and corresponding lower centile cut-offs are presented in this table. Reference values are derived from a Mixed adult population.
**Abbreviations**: LV = left ventricle; RV = right ventricle; GLS = global longitudinal strain; GCS = global circumferential strain; GRS = global radial strain.

### Supplementary Table 35 Left and right atrial global strain reference ranges and centile-based categories, by age group using suiteHEART, *in a Mixed adult population*

| | | Female | | | | Male | | | |
|---|---|---|---|---|---|---|---|---|---|
| **Age group** | Metrics (%) | Reference threshold | Mild | At least moderate | n | Reference threshold | Mild | At least moderate | n |
| **50–59** | LA GLS | ≥ 57.5 | 45.2–57.4 | < 45.2 | 123 | ≥ 55.3 | 45.8–55.2 | < 45.8 | 58 |
| | RA GLS | ≥ 54.6 | 45.9–54.5 | < 45.9 | 122 | ≥ 45.8 | 36.1–45.7 | < 36.1 | 62 |
| **60–69** | LA GLS | ≥ 48.6 | 37.6–48.5 | < 37.6 | 87 | ≥ 48.6 | 40.2–48.5 | < 40.2 | 52 |
| | RA GLS | ≥ 53.7 | 44.6–53.6 | < 44.6 | 94 | ≥ 44.7 | 35.3–44.6 | < 35.3 | 50 |
| **70+** | LA GLS | ≥ 39.4 | 29.9–39.3 | < 29.9 | 42 | ≥ 45.8 | 37.9–45.7 | < 37.9 | 29 |
| | RA GLS | ≥ 55.1 | 45.2–55.0 | < 45.2 | 39 | ≥ 43.6 | 34.4–43.5 | < 34.4 | 28 |

**Footnote**: All strain measurements were derived using suiteHEART software with standardized feature-tracking analysis. Left atrial global longitudinal strain (LA GLS) and right atrial global longitudinal strain (RA GLS) are presented as absolute values (%). Reference ranges are defined as the interval between the 2.5th and 97.5th centiles derived from age- and sex-specific generalized additive models for location, scale and shape (GAMLSS) using the Box–Cox Cole–Green distribution. Centile-based categories beyond the reference range are derived from LMS-based thresholds (P0.5 and P2.5) and describe the position of a measurement within the reference distribution rather than disease severity or diagnostic abnormality. For clarity, only the lower reference threshold (2.5th percentile) and corresponding lower centile cut-offs are presented in this table. Reference values are derived from a Mixed adult population.
**Abbreviations**: LA = left atrium; RA = right atrium; GLS = global longitudinal strain.

### Supplementary Table 36 Annular plane systolic excursion metrics reference ranges and centile-based categories, by age group using cvi42 (Circle Cardiovascular Imaging Inc), *in a White adult population*

| | | Female | | | | Male | | | |
|---|---|---|---|---|---|---|---|---|---|
| **Age group** | Metrics (mm) | Reference threshold | Mild | At least moderate | n | Reference threshold | Mild | At least moderate | n |
| **18–29** | MAPSE lateral | ≥ 8.2 | 4.5–8.1 | < 4.5 | 108 | ≥ 8.7 | 5.9–8.6 | < 5.9 | 111 |
| | MAPSE septal | ≥ 5.4 | 2.7–5.3 | < 2.7 | 108 | ≥ 6.9 | 4.8–6.8 | < 4.8 | 111 |
| | MAPSE inferior | ≥ 7.7 | 5.2–7.6 | < 5.2 | 87 | ≥ 8.7 | 6.5–8.6 | < 6.5 | 110 |
| | MAPSE anterior | ≥ 8.7 | 6.8–8.6 | < 6.8 | 89 | ≥ 6.5 | 3.4–6.4 | < 3.4 | 112 |
| | TAPSE | ≥ 13.6 | 9.1–13.5 | < 9.1 | 105 | ≥ 13.6 | 10.3–13.5 | < 10.3 | 106 |
| **30–39** | MAPSE lateral | ≥ 9.1 | 5.8–9.0 | < 5.8 | 138 | ≥ 8.9 | 6.4–8.8 | < 6.4 | 143 |
| | MAPSE septal | ≥ 6.1 | 3.4–6.0 | < 3.4 | 138 | ≥ 7.2 | 4.9–7.1 | < 4.9 | 143 |
| | MAPSE inferior | ≥ 7.4 | 4.9–7.3 | < 4.9 | 139 | ≥ 8.4 | 6.3–8.3 | < 6.3 | 138 |
| | MAPSE anterior | ≥ 6.6 | 4.0–6.5 | < 4.0 | 139 | ≥ 6.6 | 3.8–6.5 | < 3.8 | 136 |
| | TAPSE | ≥ 13.7 | 9.6–13.6 | < 9.6 | 133 | ≥ 13.3 | 9.9–13.2 | < 9.9 | 142 |
| **40–49** | MAPSE lateral | ≥ 10.1 | 7.5–10.0 | < 7.5 | 560 | ≥ 9.1 | 6.7–9.0 | < 6.7 | 481 |
| | MAPSE septal | ≥ 7.2 | 4.4–7.1 | < 4.4 | 561 | ≥ 7.3 | 5.0–7.2 | < 5.0 | 482 |
| | MAPSE inferior | ≥ 8.7 | 6.4–8.6 | < 6.4 | 562 | ≥ 8.9 | 6.6–8.8 | < 6.6 | 487 |
| | MAPSE anterior | ≥ 7.3 | 4.6–7.2 | < 4.6 | 564 | ≥ 6.9 | 4.5–6.8 | < 4.5 | 487 |
| | TAPSE | ≥ 13.7 | 10.0–13.6 | < 10.0 | 555 | ≥ 13.0 | 9.6–12.9 | < 9.6 | 488 |
| **50–59** | MAPSE lateral | ≥ 10.3 | 8.3–10.2 | < 8.3 | 1275 | ≥ 9.2 | 7.0–9.1 | < 7.0 | 1111 |
| | MAPSE septal | ≥ 7.7 | 5.2–7.6 | < 5.2 | 1275 | ≥ 7.2 | 4.9–7.1 | < 4.9 | 1112 |
| | MAPSE inferior | ≥ 9.7 | 7.6–9.6 | < 7.6 | 1271 | ≥ 9.2 | 6.8–9.1 | < 6.8 | 1130 |
| | MAPSE anterior | ≥ 8.8 | 6.8–8.7 | < 6.8 | 1273 | ≥ 7.8 | 5.5–7.7 | < 5.5 | 1130 |
| | TAPSE | ≥ 13.5 | 10.1–13.4 | < 10.1 | 1263 | ≥ 12.7 | 9.3–12.6 | < 9.3 | 1117 |
| **60–69** | MAPSE lateral | ≥ 9.9 | 8.2–9.8 | < 8.2 | 1185 | ≥ 9.2 | 7.2–9.1 | < 7.2 | 1019 |
| | MAPSE septal | ≥ 7.4 | 5.5–7.3 | < 5.5 | 1184 | ≥ 6.9 | 4.8–6.8 | < 4.8 | 1019 |
| | MAPSE inferior | ≥ 9.3 | 7.2–9.2 | < 7.2 | 1193 | ≥ 9.0 | 6.7–8.9 | < 6.7 | 1038 |
| | MAPSE anterior | ≥ 8.1 | 6.3–8.0 | < 6.3 | 1190 | ≥ 8.0 | 6.1–7.9 | < 6.1 | 1038 |
| | TAPSE | ≥ 13.2 | 10.2–13.1 | < 10.2 | 1174 | ≥ 12.5 | 9.0–12.4 | < 9.0 | 1011 |
| **70+** | MAPSE lateral | ≥ 9.3 | 7.8–9.2 | < 7.8 | 1167 | ≥ 9.2 | 7.4–9.1 | < 7.4 | 1264 |
| | MAPSE septal | ≥ 7.1 | 5.6–7.0 | < 5.6 | 1167 | ≥ 6.6 | 4.5–6.5 | < 4.5 | 1264 |
| | MAPSE inferior | ≥ 8.5 | 6.5–8.4 | < 6.5 | 1173 | ≥ 8.9 | 6.6–8.8 | < 6.6 | 1299 |
| | MAPSE anterior | ≥ 7.8 | 6.4–7.7 | < 6.4 | 1170 | ≥ 8.0 | 6.4–7.9 | < 6.4 | 1300 |
| | TAPSE | ≥ 13.3 | 10.5–13.2 | < 10.5 | 1163 | ≥ 12.3 | 8.7–12.2 | < 8.7 | 1241 |

**Footnote**: All annular plane systolic excursion measurements were derived using cvi42 software (Circle Cardiovascular Imaging Inc.) with standardized feature-tracking analysis. Mitral annular plane systolic excursion (MAPSE; lateral and septal) and tricuspid annular plane systolic excursion (TAPSE) are presented in millimetres (mm). Reference ranges are defined as the interval between the 2.5th and 97.5th centiles derived from age- and sex-specific generalized additive models for location, scale and shape (GAMLSS) using the Box–Cox Cole–Green distribution. Centile-based categories beyond the reference range are derived from LMS-based thresholds (P0.5 and P2.5) and describe the position of a measurement within the reference distribution rather than disease severity or diagnostic abnormality. For clarity, only the lower reference threshold (2.5th percentile) and corresponding lower centile cut-offs are presented in this table. Reference values are derived from a White adult population.

**Abbreviations**: MAPSE = mitral annular plane systolic excursion; TAPSE = tricuspid annular plane systolic excursion.

### Supplementary Table 37 Ventricular long-axis and atrioventricular junction strain metrics reference ranges and centile-based categories, by age group using cvi42 (Circle Cardiovascular Imaging Inc), *in a White adult population*

| | | Female | | | | Male | | | |
|---|---|---|---|---|---|---|---|---|---|
| **Age group** | Metrics (%) | Reference threshold | Mild | At least moderate | n | Reference threshold | Mild | At least moderate | n |
| **18–29** | LV long-axis strain 4CV | ≥ 10.9 | 7.8–10.8 | < 7.8 | 106 | ≥ 9.8 | 7.4–9.7 | < 7.4 | 111 |
| | LV long-axis strain avg | ≥ 13.1 | 10.7–13.0 | < 10.7 | 80 | ≥ 11.9 | 10.1–11.8 | < 10.1 | 99 |
| | LV AV-junction strain 4CV | ≥ 10.1 | 6.9–10.0 | < 6.9 | 106 | ≥ 9.3 | 7.0–9.2 | < 7.0 | 111 |
| | LV AV-junction strain avg | ≥ 12.3 | 9.8–12.2 | < 9.8 | 81 | ≥ 11.3 | 9.7–11.2 | < 9.7 | 99 |
| **30–39** | LV long-axis strain 4CV | ≥ 11.4 | 8.8–11.3 | < 8.8 | 138 | ≥ 10.0 | 7.8–9.9 | < 7.8 | 144 |
| | LV long-axis strain avg | ≥ 13.3 | 11.1–13.2 | < 11.1 | 124 | ≥ 11.9 | 10.1–11.8 | < 10.1 | 132 |
| | LV AV-junction strain 4CV | ≥ 10.7 | 8.0–10.6 | < 8.0 | 138 | ≥ 9.5 | 7.4–9.4 | < 7.4 | 144 |
| | LV AV-junction strain avg | ≥ 12.6 | 10.4–12.5 | < 10.4 | 125 | ≥ 11.4 | 9.7–11.3 | < 9.7 | 133 |
| **40–49** | LV long-axis strain 4CV | ≥ 11.8 | 9.5–11.7 | < 9.5 | 556 | ≥ 10.3 | 8.1–10.2 | < 8.1 | 478 |
| | LV long-axis strain avg | ≥ 13.4 | 11.4–13.3 | < 11.4 | 546 | ≥ 11.9 | 10.1–11.8 | < 10.1 | 459 |
| | LV AV-junction strain 4CV | ≥ 11.1 | 8.8–11.0 | < 8.8 | 557 | ≥ 9.8 | 7.7–9.7 | < 7.7 | 478 |
| | LV AV-junction strain avg | ≥ 12.8 | 10.9–12.7 | < 10.9 | 547 | ≥ 11.4 | 9.7–11.3 | < 9.7 | 460 |
| **50–59** | LV long-axis strain 4CV | ≥ 12.0 | 10.0–11.9 | < 10.0 | 1271 | ≥ 10.5 | 8.4–10.4 | < 8.4 | 1107 |
| | LV long-axis strain avg | ≥ 13.4 | 11.7–13.3 | < 11.7 | 1240 | ≥ 12.0 | 10.2–11.9 | < 10.2 | 1083 |
| | LV AV-junction strain 4CV | ≥ 11.4 | 9.5–11.3 | < 9.5 | 1270 | ≥ 10.0 | 8.0–9.9 | < 8.0 | 1107 |
| | LV AV-junction strain avg | ≥ 12.9 | 11.2–12.8 | < 11.2 | 1240 | ≥ 11.5 | 9.7–11.4 | < 9.7 | 1082 |
| **60–69** | LV long-axis strain 4CV | ≥ 12.2 | 10.4–12.1 | < 10.4 | 1182 | ≥ 10.7 | 8.8–10.6 | < 8.8 | 1017 |
| | LV long-axis strain avg | ≥ 13.4 | 11.8–13.3 | < 11.8 | 1154 | ≥ 12.0 | 10.2–11.9 | < 10.2 | 985 |
| | LV AV-junction strain 4CV | ≥ 11.6 | 9.9–11.5 | < 9.9 | 1182 | ≥ 10.3 | 8.4–10.2 | < 8.4 | 1017 |
| | LV AV-junction strain avg | ≥ 12.9 | 11.4–12.8 | < 11.4 | 1154 | ≥ 11.5 | 9.8–11.4 | < 9.8 | 986 |
| **70+** | LV long-axis strain 4CV | ≥ 12.3 | 10.7–12.2 | < 10.7 | 1165 | ≥ 11.1 | 9.2–11.0 | < 9.2 | 1263 |
| | LV long-axis strain avg | ≥ 13.4 | 11.9–13.3 | < 11.9 | 1128 | ≥ 12.1 | 10.3–12.0 | < 10.3 | 1215 |
| | LV AV-junction strain 4CV | ≥ 11.8 | 10.3–11.7 | < 10.3 | 1165 | ≥ 10.6 | 8.8–10.5 | < 8.8 | 1263 |
| | LV AV-junction strain avg | ≥ 12.9 | 11.5–12.8 | < 11.5 | 1127 | ≥ 11.6 | 9.9–11.5 | < 9.9 | 1215 |

**Footnote**: All ventricular long-axis and atrioventricular junction strain measurements were derived using cvi42 software (Circle Cardiovascular Imaging Inc.) with standardized feature-tracking analysis. Left ventricular (LV) long-axis strain and LV AV-junction strain were assessed in the 4-chamber view (4CV) and as averaged values across available views (avg), and are presented as absolute values (%). Reference ranges are defined as the interval between the 2.5th and 97.5th centiles derived from age- and sex-specific generalized additive models for location, scale and shape (GAMLSS) using the Box–Cox Cole–Green distribution. Centile-based categories beyond the reference range are derived from LMS-based thresholds (P0.5 and P2.5) and describe the position of a measurement within the reference distribution rather than disease severity or diagnostic abnormality. For clarity, only the lower reference threshold (2.5th percentile) and corresponding lower centile cut-offs are presented in this table. Reference values are derived from a White adult population.
**Abbreviations**: LV = left ventricle; AV = atrioventricular; 4CV = four-chamber view.

### Supplementary Table 38 Atrial long-axis and atrioventricular junction strain metrics reference ranges and centile-based categories, by age group using cvi42 (Circle Cardiovascular Imaging Inc), *in a White adult population*

| | | Female | | | | Male | | | |
|---|---|---|---|---|---|---|---|---|---|
| **Age group** | Metrics (%) | Reference threshold | Mild | At least moderate | n | Reference threshold | Mild | At least moderate | n |
| **18–29** | LA long-axis strain 4CV | ≥ 13.4 | 8.1–13.3 | < 8.1 | 86 | ≥ 9.9 | 6.1–9.8 | < 6.1 | 82 |
| | LA long-axis strain avg | ≥ 20.8 | 13.9–20.7 | < 13.9 | 64 | ≥ 18.2 | 13.6–18.1 | < 13.6 | 63 |
| | LA AV-junction strain 4CV | ≥ 13.8 | 9.3–13.7 | < 9.3 | 87 | ≥ 10.8 | 5.9–10.7 | < 5.9 | 81 |
| | LA AV-junction strain avg | ≥ 19.2 | 13.2–19.1 | < 13.2 | 64 | ≥ 16.1 | 12.1–16.0 | < 12.1 | 63 |
| | RA long-axis strain | ≥ 20.7 | 11.3–20.6 | < 11.3 | 93 | ≥ 18.4 | 11.1–18.3 | < 11.1 | 94 |
| | RA AV-junction strain | ≥ 21.1 | 15.1–21.0 | < 15.1 | 92 | ≥ 16.0 | 10.2–15.9 | < 10.2 | 94 |
| **30–39** | LA long-axis strain 4CV | ≥ 13.0 | 8.5–12.9 | < 8.5 | 122 | ≥ 9.4 | 5.8–9.3 | < 5.8 | 109 |
| | LA long-axis strain avg | ≥ 19.7 | 13.8–19.6 | < 13.8 | 103 | ≥ 17.5 | 12.8–17.4 | < 12.8 | 79 |
| | LA AV-junction strain 4CV | ≥ 13.2 | 9.2–13.1 | < 9.2 | 124 | ≥ 10.7 | 6.5–10.6 | < 6.5 | 110 |
| | LA AV-junction strain avg | ≥ 18.1 | 12.9–18.0 | < 12.9 | 103 | ≥ 15.6 | 11.5–15.5 | < 11.5 | 79 |
| | RA long-axis strain | ≥ 20.2 | 12.3–20.1 | < 12.3 | 124 | ≥ 17.9 | 11.1–17.8 | < 11.1 | 136 |
| | RA AV-junction strain | ≥ 20.1 | 14.4–20.0 | < 14.4 | 123 | ≥ 15.3 | 10.0–15.2 | < 10.0 | 135 |
| **40–49** | LA long-axis strain 4CV | ≥ 12.4 | 8.5–12.3 | < 8.5 | 516 | ≥ 9.0 | 5.5–8.9 | < 5.5 | 437 |
| | LA long-axis strain avg | ≥ 18.7 | 13.6–18.6 | < 13.6 | 483 | ≥ 16.9 | 12.3–16.8 | < 12.3 | 409 |
| | LA AV-junction strain 4CV | ≥ 12.4 | 8.8–12.3 | < 8.8 | 515 | ≥ 10.3 | 6.6–10.2 | < 6.6 | 438 |
| | LA AV-junction strain avg | ≥ 17.2 | 12.6–17.1 | < 12.6 | 484 | ≥ 15.2 | 11.0–15.1 | < 11.0 | 409 |
| | RA long-axis strain | ≥ 19.5 | 12.9–19.4 | < 12.9 | 548 | ≥ 16.2 | 10.2–16.1 | < 10.2 | 476 |
| | RA AV-junction strain | ≥ 19.0 | 13.8–18.9 | < 13.8 | 547 | ≥ 14.7 | 9.7–14.6 | < 9.7 | 476 |
| **50–59** | LA long-axis strain 4CV | ≥ 11.5 | 8.0–11.4 | < 8.0 | 1230 | ≥ 8.6 | 5.2–8.5 | < 5.2 | 1074 |
| | LA long-axis strain avg | ≥ 17.8 | 13.4–17.7 | < 13.4 | 1156 | ≥ 16.1 | 11.6–16.0 | < 11.6 | 1012 |
| | LA AV-junction strain 4CV | ≥ 11.2 | 8.0–11.1 | < 8.0 | 1231 | ≥ 9.3 | 6.0–9.2 | < 6.0 | 1076 |
| | LA AV-junction strain avg | ≥ 16.3 | 12.3–16.2 | < 12.3 | 1159 | ≥ 14.5 | 10.4–14.4 | < 10.4 | 1011 |
| | RA long-axis strain | ≥ 18.5 | 12.9–18.4 | < 12.9 | 1253 | ≥ 14.8 | 9.6–14.7 | < 9.6 | 1110 |
| | RA AV-junction strain | ≥ 17.7 | 12.8–17.6 | < 12.8 | 1251 | ≥ 14.2 | 9.5–14.1 | < 9.5 | 1110 |
| **60–69** | LA long-axis strain 4CV | ≥ 10.3 | 7.3–10.2 | < 7.3 | 1126 | ≥ 8.1 | 5.0–8.0 | < 5.0 | 974 |
| | LA long-axis strain avg | ≥ 16.8 | 13.0–16.7 | < 13.0 | 1078 | ≥ 14.9 | 10.7–14.8 | < 10.7 | 917 |
| | LA AV-junction strain 4CV | ≥ 9.8 | 6.9–9.7 | < 6.9 | 1129 | ≥ 8.0 | 5.1–7.9 | < 5.1 | 979 |
| | LA AV-junction strain avg | ≥ 15.4 | 11.9–15.3 | < 11.9 | 1079 | ≥ 13.4 | 9.5–13.3 | < 9.5 | 918 |
| | RA long-axis strain | ≥ 17.8 | 12.9–17.7 | < 12.9 | 1152 | ≥ 14.8 | 9.7–14.7 | < 9.7 | 1009 |
| | RA AV-junction strain | ≥ 16.4 | 11.8–16.3 | < 11.8 | 1151 | ≥ 13.6 | 9.2–13.5 | < 9.2 | 1009 |
| **70+** | LA long-axis strain 4CV | ≥ 8.9 | 6.3–8.8 | < 6.3 | 1122 | ≥ 7.6 | 4.7–7.5 | < 4.7 | 1211 |
| | LA long-axis strain avg | ≥ 15.6 | 12.4–15.5 | < 12.4 | 1061 | ≥ 13.4 | 9.6–13.3 | < 9.6 | 1157 |
| | LA AV-junction strain 4CV | ≥ 8.2 | 5.7–8.1 | < 5.7 | 1126 | ≥ 6.5 | 4.0–6.4 | < 4.0 | 1220 |
| | LA AV-junction strain avg | ≥ 14.2 | 11.3–14.1 | < 11.3 | 1062 | ≥ 11.9 | 8.4–11.8 | < 8.4 | 1151 |
| | RA long-axis strain | ≥ 17.5 | 13.1–17.4 | < 13.1 | 1141 | ≥ 14.2 | 9.6–14.1 | < 9.6 | 1231 |
| | RA AV-junction strain | ≥ 15.3 | 10.8–15.2 | < 10.8 | 1145 | ≥ 12.9 | 8.9–12.8 | < 8.9 | 1231 |

**Footnote**: All atrial long-axis and atrioventricular junction strain measurements were derived using cvi42 software (Circle Cardiovascular Imaging Inc.) with standardized feature-tracking analysis. Left atrial (LA) long-axis strain and LA AV-junction strain were assessed in the 4-chamber view (4CV) and as averaged values across available views (avg), while right atrial (RA) long-axis and AV-junction strain were derived from standard long-axis views; all values are presented as absolute percentages (%). Reference ranges are defined as the interval between the 2.5th and 97.5th centiles derived from age- and sex-specific generalized additive models for location, scale and shape (GAMLSS) using the Box–Cox Cole–Green distribution. Centile-based categories beyond the reference range are derived from LMS-based thresholds (P0.5 and P2.5) and describe the position of a measurement within the reference distribution rather than disease severity or diagnostic abnormality. For clarity, only the lower reference threshold (2.5th percentile) and corresponding lower centile cut-offs are presented in this table. Reference values are derived from a White adult population.
**Abbreviations**: LA = left atrium; RA = right atrium; AV = atrioventricular; 4CV = four-chamber view.

**Supplementary Table 39 Annular plane systolic excursion metrics reference ranges and centile-based categories, by age group using cvi42 (Circle Cardiovascular Imaging Inc),** ***in a Black adult population***

| | | Female | | | | Male | | | |
|---|---|---|---|---|---|---|---|---|---|
| **Age group** | Metrics (mm) | Reference threshold | Mild | At least moderate | n | Reference threshold | Mild | At least moderate | n |
| **40–49** | MAPSE lateral | ≥ 11.5 | 10.2–11.4 | < 10.2 | 10 | | | | |
| | MAPSE septal | ≥ 9.8 | 7.5–9.7 | < 7.5 | 10 | | | | |
| | MAPSE inferior | ≥ 11.9 | 11.0–11.8 | < 11.0 | 10 | | | | |
| | MAPSE anterior | ≥ 9.6 | 7.9–9.5 | < 7.9 | 10 | | | | |
| | TAPSE | ≥ 15.7 | 13.1–15.6 | < 13.1 | 10 | | | | |
| **50–59** | MAPSE lateral | ≥ 10.9 | 9.5–10.8 | < 9.5 | 94 | ≥ 9.9 | 8.3–9.8 | < 8.3 | 70 |
| | MAPSE septal | ≥ 8.9 | 6.9–8.8 | < 6.9 | 94 | ≥ 9.3 | 8.2–9.2 | < 8.2 | 70 |
| | MAPSE inferior | ≥ 11.6 | 10.5–11.5 | < 10.5 | 93 | ≥ 11.6 | 10.5–11.5 | < 10.5 | 67 |
| | MAPSE anterior | ≥ 9.4 | 7.8–9.3 | < 7.8 | 93 | ≥ 10.0 | 8.8–9.9 | < 8.8 | 67 |
| | TAPSE | ≥ 14.9 | 12.2–14.8 | < 12.2 | 93 | ≥ 15.5 | 13.5–15.4 | < 13.5 | 72 |
| **60–69** | MAPSE lateral | ≥ 9.9 | 8.5–9.8 | < 8.5 | 63 | ≥ 9.9 | 8.4–9.8 | < 8.4 | 44 |
| | MAPSE septal | ≥ 7.8 | 6.1–7.7 | < 6.1 | 63 | ≥ 7.7 | 6.3–7.6 | < 6.3 | 44 |
| | MAPSE inferior | ≥ 11.1 | 9.7–11.0 | < 9.7 | 63 | ≥ 11.1 | 9.8–11.0 | < 9.8 | 45 |
| | MAPSE anterior | ≥ 9.1 | 7.7–9.0 | < 7.7 | 63 | ≥ 9.9 | 8.6–9.8 | < 8.6 | 45 |
| | TAPSE | ≥ 13.8 | 11.0–13.7 | < 11.0 | 60 | ≥ 15.7 | 14.3–15.6 | < 14.3 | 45 |
| **70+** | MAPSE lateral | ≥ 8.9 | 7.5–8.8 | < 7.5 | 17 | ≥ 9.8 | 8.4–9.7 | < 8.4 | 12 |
| | MAPSE septal | ≥ 6.7 | 5.4–6.6 | < 5.4 | 17 | ≥ 6.0 | 4.1–5.9 | < 4.1 | 12 |
| | MAPSE inferior | ≥ 10.5 | 8.6–10.4 | < 8.6 | 17 | ≥ 10.6 | 9.0–10.5 | < 9.0 | 14 |
| | MAPSE anterior | ≥ 8.8 | 7.5–8.7 | < 7.5 | 17 | ≥ 9.8 | 8.5–9.7 | < 8.5 | 14 |
| | TAPSE | ≥ 12.8 | 10.1–12.7 | < 10.1 | 16 | ≥ 15.8 | 14.6–15.7 | < 14.6 | 10 |

**Footnote**: All annular plane systolic excursion measurements were derived using cvi42 software (Circle Cardiovascular Imaging Inc.) with standardized feature-tracking analysis. Mitral annular plane systolic excursion (MAPSE; lateral and septal) and tricuspid annular plane systolic excursion (TAPSE) are presented in millimetres (mm). Reference ranges are defined as the interval between the 2.5th and 97.5th centiles derived from age- and sex-specific generalized additive models for location, scale and shape (GAMLSS) using the Box–Cox Cole–Green distribution. Centile-based categories beyond the reference range are derived from LMS-based thresholds (P0.5 and P2.5) and describe the position of a measurement within the reference distribution rather than disease severity or diagnostic abnormality. For clarity, only the lower reference threshold (2.5th percentile) and corresponding lower centile cut-offs are presented in this table. Reference values are derived from a Black adult population.

**Abbreviations**: MAPSE = mitral annular plane systolic excursion; TAPSE = tricuspid annular plane systolic excursion.

### Supplementary Table 40 Ventricular long-axis and atrioventricular junction strain metrics reference ranges and centile-based categories, by age group using cvi42 (Circle Cardiovascular Imaging Inc), *in a Black adult population*

| | | Female | | | | Male | | | |
|---|---|---|---|---|---|---|---|---|---|
| **Age group** | Metrics (%) | Reference threshold | Mild | At least moderate | n | Reference threshold | Mild | At least moderate | n |
| **40–49** | LV long-axis strain 4CV | ≥ 12.1 | 10.3–12.0 | < 10.3 | 10 | | | | |
| | LV long-axis strain avg | ≥ 14.2 | 13.2–14.1 | < 13.2 | 10 | | | | |
| | LV AV-junction strain 4CV | ≥ 12.0 | 10.5–11.9 | < 10.5 | 10 | | | | |
| | LV AV-junction strain avg | ≥ 13.8 | 12.9–13.7 | < 12.9 | 10 | | | | |
| **50–59** | LV long-axis strain 4CV | ≥ 11.9 | 10.1–11.8 | < 10.1 | 94 | ≥ 10.3 | 8.6–10.2 | < 8.6 | 70 |
| | LV long-axis strain avg | ≥ 13.4 | 12.2–13.3 | < 12.2 | 92 | ≥ 12.3 | 11.0–12.2 | < 11.0 | 63 |
| | LV AV-junction strain 4CV | ≥ 11.6 | 10.2–11.5 | < 10.2 | 94 | ≥ 9.9 | 8.3–9.8 | < 8.3 | 70 |
| | LV AV-junction strain avg | ≥ 13.0 | 11.9–12.9 | < 11.9 | 92 | ≥ 11.7 | 10.5–11.6 | < 10.5 | 63 |
| **60–69** | LV long-axis strain 4CV | ≥ 11.4 | 9.8–11.3 | < 9.8 | 63 | ≥ 10.4 | 8.4–10.3 | < 8.4 | 44 |
| | LV long-axis strain avg | ≥ 11.9 | 10.1–11.8 | < 10.1 | 59 | ≥ 12.0 | 10.5–11.9 | < 10.5 | 43 |
| | LV AV-junction strain 4CV | ≥ 11.1 | 9.7–11.0 | < 9.7 | 63 | ≥ 9.9 | 8.0–9.8 | < 8.0 | 44 |
| | LV AV-junction strain avg | ≥ 11.5 | 9.8–11.4 | < 9.8 | 59 | ≥ 11.5 | 10.1–11.4 | < 10.1 | 43 |
| **70+** | LV long-axis strain 4CV | ≥ 11.0 | 9.5–10.9 | < 9.5 | 17 | ≥ 10.6 | 8.1–10.5 | < 8.1 | 12 |
| | LV long-axis strain avg | ≥ 9.7 | 6.3–9.6 | < 6.3 | 16 | ≥ 11.7 | 9.9–11.6 | < 9.9 | 12 |
| | LV AV-junction strain 4CV | ≥ 10.6 | 9.3–10.5 | < 9.3 | 17 | ≥ 9.9 | 7.5–9.8 | < 7.5 | 12 |
| | LV AV-junction strain avg | ≥ 9.1 | 5.9–9.0 | < 5.9 | 16 | ≥ 11.3 | 9.5–11.2 | < 9.5 | 12 |

**Footnote**: All ventricular long-axis and atrioventricular junction strain measurements were derived using cvi42 software (Circle Cardiovascular Imaging Inc.) with standardized feature-tracking analysis. Left ventricular (LV) long-axis strain and LV AV-junction strain were assessed in the 4-chamber view (4CV) and as averaged values across available views (avg), and are presented as absolute values (%). Reference ranges are defined as the interval between the 2.5th and 97.5th centiles derived from age- and sex-specific generalized additive models for location, scale and shape (GAMLSS) using the Box–Cox Cole–Green distribution. Centile-based categories beyond the reference range are derived from LMS-based thresholds (P0.5 and P2.5) and describe the position of a measurement within the reference distribution rather than disease severity or diagnostic abnormality. For clarity, only the lower reference threshold (2.5th percentile) and corresponding lower centile cut-offs are presented in this table. Reference values are derived from a Black adult population.
**Abbreviations**: LV = left ventricle; AV = atrioventricular; 4CV = four-chamber view.

**Supplementary Table 41 Atrial long-axis and atrioventricular junction strain metrics reference ranges and centile-based categories, by age group using cvi42 (Circle Cardiovascular Imaging Inc),** ***in a Black adult population***

| | | Female | | | | Male | | | |
|---|---|---|---|---|---|---|---|---|---|
| **Age group** | Metrics (%) | Reference threshold | Mild | At least moderate | n | Reference threshold | Mild | At least moderate | n |
| **40–49** | LA long-axis strain 4CV | | | | | | | | |
| | LA long-axis strain avg | | | | | | | | |
| | LA AV-junction strain 4CV | | | | | | | | |
| | LA AV-junction strain avg | | | | | | | | |
| | RA long-axis strain | ≥ 21.6 | 16.9–21.5 | < 16.9 | 10 | | | | |
| | RA AV-junction strain | ≥ 20.2 | 16.3–20.1 | < 16.3 | 10 | | | | |
| **50–59** | LA long-axis strain 4CV | ≥ 12.5 | 9.5–12.4 | < 9.5 | 95 | ≥ 11.9 | 9.5–11.8 | < 9.5 | 69 |
| | LA long-axis strain avg | ≥ 23.2 | 20.7–23.1 | < 20.7 | 92 | ≥ 19.3 | 16.3–19.2 | < 16.3 | 63 |
| | LA AV-junction strain 4CV | ≥ 12.9 | 10.2–12.8 | < 10.2 | 95 | ≥ 12.2 | 10.1–12.1 | < 10.1 | 70 |
| | LA AV-junction strain avg | ≥ 21.2 | 19.0–21.1 | < 19.0 | 92 | ≥ 17.7 | 15.1–17.6 | < 15.1 | 63 |
| | RA long-axis strain | ≥ 19.1 | 14.4–19.0 | < 14.4 | 94 | ≥ 18.6 | 14.7–18.5 | < 14.7 | 74 |
| | RA AV-junction strain | ≥ 17.9 | 13.9–17.8 | < 13.9 | 94 | ≥ 16.5 | 12.6–16.4 | < 12.6 | 74 |
| **60–69** | LA long-axis strain 4CV | ≥ 11.3 | 8.4–11.2 | < 8.4 | 64 | ≥ 10.0 | 7.5–9.9 | < 7.5 | 47 |
| | LA long-axis strain avg | ≥ 19.6 | 16.9–19.5 | < 16.9 | 59 | ≥ 18.3 | 14.6–18.2 | < 14.6 | 45 |
| | LA AV-junction strain 4CV | ≥ 11.5 | 9.0–11.4 | < 9.0 | 64 | ≥ 9.9 | 7.5–9.8 | < 7.5 | 47 |
| | LA AV-junction strain avg | ≥ 18.0 | 15.6–17.9 | < 15.6 | 59 | ≥ 16.7 | 13.3–16.6 | < 13.3 | 45 |
| | RA long-axis strain | ≥ 16.8 | 12.5–16.7 | < 12.5 | 65 | ≥ 17.6 | 14.1–17.5 | < 14.1 | 47 |
| | RA AV-junction strain | ≥ 16.6 | 12.9–16.5 | < 12.9 | 65 | ≥ 15.7 | 12.6–15.6 | < 12.6 | 47 |
| **70+** | LA long-axis strain 4CV | ≥ 10.1 | 7.4–10.0 | < 7.4 | 18 | ≥ 8.2 | 5.7–8.1 | < 5.7 | 12 |
| | LA long-axis strain avg | ≥ 15.8 | 12.9–15.7 | < 12.9 | 17 | ≥ 18.7 | 13.5–18.6 | < 13.5 | 12 |
| | LA AV-junction strain 4CV | ≥ 10.2 | 7.8–10.1 | < 7.8 | 18 | ≥ 7.6 | 5.0–7.5 | < 5.0 | 12 |
| | LA AV-junction strain avg | ≥ 14.5 | 11.8–14.4 | < 11.8 | 17 | ≥ 16.9 | 11.8–16.8 | < 11.8 | 12 |
| | RA long-axis strain | ≥ 16.8 | 13.2–16.7 | < 13.2 | 18 | ≥ 16.9 | 13.6–16.8 | < 13.6 | 11 |
| | RA AV-junction strain | ≥ 19.0 | 16.3–18.9 | < 16.3 | 18 | ≥ 15.2 | 12.5–15.1 | < 12.5 | 11 |

**Footnote**: All atrial long-axis and atrioventricular junction strain measurements were derived using cvi42 software (Circle Cardiovascular Imaging Inc.) with standardized feature-tracking analysis. Left atrial (LA) long-axis strain and LA AV-junction strain were assessed in the 4-chamber view (4CV) and as averaged values across available views (avg), while right atrial (RA) long-axis and AV-junction strain were derived from standard long-axis views; all values are presented as absolute percentages (%). Reference ranges are defined as the interval between the 2.5th and 97.5th centiles derived from age- and sex-specific generalized additive models for location, scale and shape (GAMLSS) using the Box–Cox Cole–Green distribution. Centile-based categories beyond the reference range are derived from LMS-based thresholds (P0.5 and P2.5) and describe the position of a measurement within the reference distribution rather than disease severity or diagnostic abnormality. For clarity, only the lower reference threshold (2.5th percentile) and corresponding lower centile cut-offs are presented in this table. Reference values are derived from a Black adult population.
**Abbreviations**: LA = left atrium; RA = right atrium; AV = atrioventricular; 4CV = four-chamber view.

**Supplementary Table 42 Annular plane systolic excursion metrics reference ranges and centile-based categories, by age group using cvi42 (Circle Cardiovascular Imaging Inc), *in a South Asian adult population***

| | | Female | | | | Male | | | |
|---|---|---|---|---|---|---|---|---|---|
| **Age group** | Metrics (mm) | Reference threshold | Mild | At least moderate | n | Reference threshold | Mild | At least moderate | n |
| **40–49** | MAPSE lateral | | | | | ≥ 11.8 | 9.7–11.7 | < 9.7 | 19 |
| | MAPSE septal | | | | | ≥ 9.5 | 7.9–9.4 | < 7.9 | 19 |
| | MAPSE inferior | | | | | ≥ 12.3 | 10.0–12.2 | < 10.0 | 20 |
| | MAPSE anterior | | | | | ≥ 10.2 | 8.8–10.1 | < 8.8 | 20 |
| | TAPSE | | | | | ≥ 14.1 | 11.2–14.0 | < 11.2 | 19 |
| **50–59** | MAPSE lateral | ≥ 10.8 | 9.3–10.7 | < 9.3 | 81 | ≥ 10.7 | 8.8–10.6 | < 8.8 | 132 |
| | MAPSE septal | ≥ 8.4 | 6.5–8.3 | < 6.5 | 81 | ≥ 8.8 | 7.1–8.7 | < 7.1 | 133 |
| | MAPSE inferior | ≥ 11.0 | 9.8–10.9 | < 9.8 | 83 | ≥ 11.5 | 9.7–11.4 | < 9.7 | 134 |
| | MAPSE anterior | ≥ 9.9 | 8.7–9.8 | < 8.7 | 83 | ≥ 9.7 | 8.2–9.6 | < 8.2 | 134 |
| | TAPSE | ≥ 14.6 | 12.5–14.5 | < 12.5 | 80 | ≥ 13.8 | 10.9–13.7 | < 10.9 | 134 |
| **60–69** | MAPSE lateral | ≥ 10.7 | 9.5–10.6 | < 9.5 | 58 | ≥ 9.9 | 8.1–9.8 | < 8.1 | 114 |
| | MAPSE septal | ≥ 8.4 | 7.0–8.3 | < 7.0 | 58 | ≥ 7.9 | 6.0–7.8 | < 6.0 | 114 |
| | MAPSE inferior | ≥ 10.4 | 9.1–10.3 | < 9.1 | 60 | ≥ 10.7 | 9.3–10.6 | < 9.3 | 117 |
| | MAPSE anterior | ≥ 9.1 | 7.8–9.0 | < 7.8 | 60 | ≥ 9.0 | 7.4–8.9 | < 7.4 | 117 |
| | TAPSE | ≥ 13.6 | 11.3–13.5 | < 11.3 | 59 | ≥ 13.5 | 10.6–13.4 | < 10.6 | 112 |
| **70+** | MAPSE lateral | ≥ 10.7 | 9.6–10.6 | < 9.6 | 30 | ≥ 9.7 | 8.1–9.6 | < 8.1 | 37 |
| | MAPSE septal | ≥ 8.2 | 7.2–8.1 | < 7.2 | 30 | ≥ 6.9 | 4.6–6.8 | < 4.6 | 37 |
| | MAPSE inferior | ≥ 9.7 | 8.4–9.6 | < 8.4 | 30 | ≥ 10.1 | 9.1–10.0 | < 9.1 | 40 |
| | MAPSE anterior | ≥ 8.1 | 6.8–8.0 | < 6.8 | 30 | ≥ 8.2 | 6.6–8.1 | < 6.6 | 40 |
| | TAPSE | ≥ 12.5 | 9.9–12.4 | < 9.9 | 28 | ≥ 13.2 | 10.4–13.1 | < 10.4 | 36 |

**Footnote**: All annular plane systolic excursion measurements were derived using cvi42 software (Circle Cardiovascular Imaging Inc.) with standardized feature-tracking analysis. Mitral annular plane systolic excursion (MAPSE; lateral and septal) and tricuspid annular plane systolic excursion (TAPSE) are presented in millimetres (mm). Reference ranges are defined as the interval between the 2.5th and 97.5th centiles derived from age- and sex-specific generalized additive models for location, scale and shape (GAMLSS) using the Box–Cox Cole–Green distribution. Centile-based categories beyond the reference range are derived from LMS-based thresholds (P0.5 and P2.5) and describe the position of a measurement within the reference distribution rather than disease severity or diagnostic abnormality. For clarity, only the lower reference threshold (2.5th percentile) and corresponding lower centile cut-offs are presented in this table. Reference values are derived from a South Asian adult population.
**Abbreviations**: MAPSE = mitral annular plane systolic excursion; TAPSE = tricuspid annular plane systolic excursion.

### Supplementary Table 43 Ventricular long-axis and atrioventricular junction strain metrics reference ranges and centile-based categories, by age group using cvi42 (Circle Cardiovascular Imaging Inc), *in a South Asian adult population*

| | | Female | | | | Male | | | |
|---|---|---|---|---|---|---|---|---|---|
| **Age group** | Metrics (%) | Reference threshold | Mild | At least moderate | n | Reference threshold | Mild | At least moderate | n |
| **40–49** | LV long-axis strain 4CV | | | | | ≥ 10.0 | 7.0–9.9 | < 7.0 | 19 |
| | LV long-axis strain avg | | | | | ≥ 14.2 | 13.1–14.1 | < 13.1 | 19 |
| | LV AV-junction strain 4CV | | | | | ≥ 9.7 | 6.8–9.6 | < 6.8 | 19 |
| | LV AV-junction strain avg | | | | | ≥ 13.6 | 12.6–13.5 | < 12.6 | 19 |
| **50–59** | LV long-axis strain 4CV | ≥ 12.8 | 11.3–12.7 | < 11.3 | 80 | ≥ 10.8 | 8.6–10.7 | < 8.6 | 132 |
| | LV long-axis strain avg | ≥ 14.4 | 13.2–14.3 | < 13.2 | 81 | ≥ 13.5 | 12.3–13.4 | < 12.3 | 132 |
| | LV AV-junction strain 4CV | ≥ 12.2 | 10.7–12.1 | < 10.7 | 80 | ≥ 10.5 | 8.4–10.4 | < 8.4 | 132 |
| | LV AV-junction strain avg | ≥ 13.8 | 12.6–13.7 | < 12.6 | 81 | ≥ 13.1 | 12.0–13.0 | < 12.0 | 132 |
| **60–69** | LV long-axis strain 4CV | ≥ 13.2 | 11.9–13.1 | < 11.9 | 58 | ≥ 11.6 | 9.9–11.5 | < 9.9 | 114 |
| | LV long-axis strain avg | ≥ 14.4 | 13.3–14.3 | < 13.3 | 56 | ≥ 13.1 | 11.8–13.0 | < 11.8 | 112 |
| | LV AV-junction strain 4CV | ≥ 12.5 | 11.2–12.4 | < 11.2 | 58 | ≥ 11.3 | 9.8–11.2 | < 9.8 | 114 |
| | LV AV-junction strain avg | ≥ 13.9 | 12.8–13.8 | < 12.8 | 56 | ≥ 12.8 | 11.6–12.7 | < 11.6 | 112 |
| **70+** | LV long-axis strain 4CV | ≥ 13.4 | 12.4–13.3 | < 12.4 | 30 | ≥ 12.5 | 11.2–12.4 | < 11.2 | 37 |
| | LV long-axis strain avg | ≥ 15.1 | 14.2–15.0 | < 14.2 | 30 | ≥ 13.3 | 11.7–13.2 | < 11.7 | 35 |
| | LV AV-junction strain 4CV | ≥ 12.7 | 11.6–12.6 | < 11.6 | 30 | ≥ 12.5 | 11.3–12.4 | < 11.3 | 37 |
| | LV AV-junction strain avg | ≥ 14.7 | 13.8–14.6 | < 13.8 | 30 | ≥ 13.0 | 11.6–12.9 | < 11.6 | 35 |

**Footnote**: All ventricular long-axis and atrioventricular junction strain measurements were derived using cvi42 software (Circle Cardiovascular Imaging Inc.) with standardized feature-tracking analysis. Left ventricular (LV) long-axis strain and LV AV-junction strain were assessed in the 4-chamber view (4CV) and as averaged values across available views (avg), and are presented as absolute values (%). Reference ranges are defined as the interval between the 2.5th and 97.5th centiles derived from age- and sex-specific generalized additive models for location, scale and shape (GAMLSS) using the Box–Cox Cole–Green distribution. Centile-based categories beyond the reference range are derived from LMS-based thresholds (P0.5 and P2.5) and describe the position of a measurement within the reference distribution rather than disease severity or diagnostic abnormality. For clarity, only the lower reference threshold (2.5th percentile) and corresponding lower centile cut-offs are presented in this table. Reference values are derived from a South Asian adult population.
**Abbreviations**: LV = left ventricle; AV = atrioventricular; 4CV = four-chamber view.

### Supplementary Table 44 Atrial long-axis and atrioventricular junction strain metrics reference ranges and centile-based categories, by age group using cvi42 (Circle Cardiovascular Imaging Inc), *in a South Asian adult population*

| | | Female | | | | Male | | | |
|---|---|---|---|---|---|---|---|---|---|
| **Age group** | Metrics (%) | Reference threshold | Mild | At least moderate | n | Reference threshold | Mild | At least moderate | n |
| **40–49** | LA long-axis strain 4CV | | | | | ≥ 12.0 | 7.7–11.9 | < 7.7 | 19 |
| | LA long-axis strain avg | | | | | ≥ 23.8 | 20.1–23.7 | < 20.1 | 19 |
| | LA AV-junction strain 4CV | | | | | ≥ 11.9 | 7.7–11.8 | < 7.7 | 19 |
| | LA AV-junction strain avg | | | | | ≥ 20.8 | 17.2–20.7 | < 17.2 | 19 |
| | RA long-axis strain | | | | | ≥ 19.9 | 14.7–19.8 | < 14.7 | 20 |
| | RA AV-junction strain | | | | | ≥ 19.1 | 15.1–19.0 | < 15.1 | 20 |
| **50–59** | LA long-axis strain 4CV | ≥ 12.9 | 10.3–12.8 | < 10.3 | 77 | ≥ 11.0 | 7.4–10.9 | < 7.4 | 132 |
| | LA long-axis strain avg | ≥ 21.7 | 18.3–21.6 | < 18.3 | 77 | ≥ 21.6 | 18.3–21.5 | < 18.3 | 127 |
| | LA AV-junction strain 4CV | ≥ 12.3 | 9.8–12.2 | < 9.8 | 77 | ≥ 10.6 | 7.0–10.5 | < 7.0 | 132 |
| | LA AV-junction strain avg | ≥ 19.3 | 16.3–19.2 | < 16.3 | 77 | ≥ 19.2 | 16.1–19.1 | < 16.1 | 126 |
| | RA long-axis strain | ≥ 18.6 | 13.5–18.5 | < 13.5 | 81 | ≥ 18.7 | 13.3–18.6 | < 13.3 | 135 |
| | RA AV-junction strain | ≥ 16.9 | 11.2–16.8 | < 11.2 | 81 | ≥ 17.8 | 13.3–17.7 | < 13.3 | 135 |
| **60–69** | LA long-axis strain 4CV | ≥ 11.0 | 8.1–10.9 | < 8.1 | 57 | ≥ 9.7 | 6.9–9.6 | < 6.9 | 113 |
| | LA long-axis strain avg | ≥ 19.2 | 16.1–19.1 | < 16.1 | 58 | ≥ 19.0 | 16.0–18.9 | < 16.0 | 111 |
| | LA AV-junction strain 4CV | ≥ 10.6 | 7.8–10.5 | < 7.8 | 57 | ≥ 9.1 | 6.2–9.0 | < 6.2 | 113 |
| | LA AV-junction strain avg | ≥ 17.3 | 14.4–17.2 | < 14.4 | 58 | ≥ 17.3 | 14.6–17.2 | < 14.6 | 111 |
| | RA long-axis strain | ≥ 18.9 | 14.1–18.8 | < 14.1 | 58 | ≥ 17.2 | 11.4–17.1 | < 11.4 | 113 |
| | RA AV-junction strain | ≥ 17.3 | 12.5–17.2 | < 12.5 | 58 | ≥ 16.0 | 10.5–15.9 | < 10.5 | 113 |
| **70+** | LA long-axis strain 4CV | ≥ 8.7 | 5.5–8.6 | < 5.5 | 30 | ≥ 8.4 | 6.2–8.3 | < 6.2 | 40 |
| | LA long-axis strain avg | ≥ 16.8 | 13.9–16.7 | < 13.9 | 27 | ≥ 16.2 | 13.6–16.1 | < 13.6 | 38 |
| | LA AV-junction strain 4CV | ≥ 8.7 | 5.7–8.6 | < 5.7 | 30 | ≥ 7.7 | 5.5–7.6 | < 5.5 | 40 |
| | LA AV-junction strain avg | ≥ 15.2 | 12.4–15.1 | < 12.4 | 27 | ≥ 15.1 | 12.9–15.0 | < 12.9 | 38 |
| | RA long-axis strain | ≥ 19.1 | 14.7–19.0 | < 14.7 | 29 | ≥ 15.4 | 9.2–15.3 | < 9.2 | 38 |
| | RA AV-junction strain | ≥ 17.3 | 13.3–17.2 | < 13.3 | 29 | ≥ 13.8 | 7.5–13.7 | < 7.5 | 38 |

**Footnote**: All atrial long-axis and atrioventricular junction strain measurements were derived using cvi42 software (Circle Cardiovascular Imaging Inc.) with standardized feature-tracking analysis. Left atrial (LA) long-axis strain and LA AV-junction strain were assessed in the 4-chamber view (4CV) and as averaged values across available views (avg), while right atrial (RA) long-axis and AV-junction strain were derived from standard long-axis views; all values are presented as absolute percentages (%). Reference ranges are defined as the interval between the 2.5th and 97.5th centiles derived from age- and sex-specific generalized additive models for location, scale and shape (GAMLSS) using the Box–Cox Cole–Green distribution. Centile-based categories beyond the reference range are derived from LMS-based thresholds (P0.5 and P2.5) and describe the position of a measurement within the reference distribution rather than disease severity or diagnostic abnormality. For clarity, only the lower reference threshold (2.5th percentile) and corresponding lower centile cut-offs are presented in this table. Reference values are derived from a South Asian adult population.

**Abbreviations**: LA = left atrium; RA = right atrium; AV = atrioventricular; 4CV = four-chamber view.

**Supplementary Table 45 Annular plane systolic excursion metrics reference ranges and centile-based categories, by age group using cvi42 (Circle Cardiovascular Imaging Inc),** ***in a Chinese adult population***

| | | Female | | | | Male | | | |
|---|---|---|---|---|---|---|---|---|---|
| **Age group** | Metrics (mm) | Reference threshold | Mild | At least moderate | n | Reference threshold | Mild | At least moderate | n |
| **18–29** | MAPSE lateral | ≥ 12.8 | 10.5–12.7 | < 10.5 | 14 | ≥ 12.0 | 10.0–11.9 | < 10.0 | 17 |
| | MAPSE septal | ≥ 8.4 | 6.2–8.3 | < 6.2 | 14 | ≥ 8.8 | 7.1–8.7 | < 7.1 | 17 |
| | MAPSE inferior | ≥ 10.1 | 8.6–10.0 | < 8.6 | 14 | ≥ 9.1 | 5.9–9.0 | < 5.9 | 17 |
| | MAPSE anterior | ≥ 11.5 | 10.3–11.4 | < 10.3 | 14 | ≥ 9.4 | 7.7–9.3 | < 7.7 | 17 |
| | TAPSE | ≥ 17.1 | 15.2–17.0 | < 15.2 | 14 | ≥ 12.3 | 9.1–12.2 | < 9.1 | 17 |
| **30–39** | MAPSE lateral | ≥ 12.4 | 10.5–12.3 | < 10.5 | 18 | ≥ 11.7 | 10.0–11.6 | < 10.0 | 16 |
| | MAPSE septal | ≥ 8.0 | 5.9–7.9 | < 5.9 | 18 | ≥ 8.6 | 7.0–8.5 | < 7.0 | 16 |
| | MAPSE inferior | ≥ 9.8 | 8.3–9.7 | < 8.3 | 18 | ≥ 9.0 | 6.0–8.9 | < 6.0 | 17 |
| | MAPSE anterior | ≥ 10.6 | 9.3–10.5 | < 9.3 | 18 | ≥ 8.9 | 7.2–8.8 | < 7.2 | 17 |
| | TAPSE | ≥ 16.9 | 15.1–16.8 | < 15.1 | 18 | ≥ 12.5 | 9.2–12.4 | < 9.2 | 16 |
| **40–49** | MAPSE lateral | ≥ 11.7 | 10.2–11.6 | < 10.2 | 24 | ≥ 11.3 | 9.9–11.2 | < 9.9 | 19 |
| | MAPSE septal | ≥ 7.6 | 5.5–7.5 | < 5.5 | 24 | ≥ 8.3 | 6.9–8.2 | < 6.9 | 19 |
| | MAPSE inferior | ≥ 9.6 | 7.9–9.5 | < 7.9 | 24 | ≥ 8.9 | 6.4–8.8 | < 6.4 | 20 |
| | MAPSE anterior | ≥ 9.7 | 8.3–9.6 | < 8.3 | 24 | ≥ 8.5 | 6.8–8.4 | < 6.8 | 20 |
| | TAPSE | ≥ 16.5 | 14.8–16.4 | < 14.8 | 24 | ≥ 12.5 | 9.3–12.4 | < 9.3 | 19 |
| **50–59** | MAPSE lateral | ≥ 10.9 | 9.5–10.8 | < 9.5 | 66 | ≥ 11.0 | 9.8–10.9 | < 9.8 | 46 |
| | MAPSE septal | ≥ 7.3 | 5.3–7.2 | < 5.3 | 66 | ≥ 8.1 | 6.8–8.0 | < 6.8 | 46 |
| | MAPSE inferior | ≥ 9.3 | 7.5–9.2 | < 7.5 | 65 | ≥ 8.9 | 6.8–8.8 | < 6.8 | 48 |
| | MAPSE anterior | ≥ 8.7 | 7.2–8.6 | < 7.2 | 66 | ≥ 8.0 | 6.4–7.9 | < 6.4 | 48 |
| | TAPSE | ≥ 15.7 | 14.1–15.6 | < 14.1 | 66 | ≥ 12.6 | 9.4–12.5 | < 9.4 | 47 |
| **60–69** | MAPSE lateral | ≥ 9.8 | 8.4–9.7 | < 8.4 | 52 | ≥ 10.7 | 9.6–10.6 | < 9.6 | 35 |
| | MAPSE septal | ≥ 6.9 | 5.0–6.8 | < 5.0 | 52 | ≥ 7.9 | 6.7–7.8 | < 6.7 | 35 |
| | MAPSE inferior | ≥ 9.0 | 7.0–8.9 | < 7.0 | 51 | ≥ 8.8 | 7.1–8.7 | < 7.1 | 36 |
| | MAPSE anterior | ≥ 7.7 | 6.2–7.6 | < 6.2 | 51 | ≥ 7.6 | 6.1–7.5 | < 6.1 | 36 |
| | TAPSE | ≥ 14.6 | 13.0–14.5 | < 13.0 | 52 | ≥ 12.8 | 9.5–12.7 | < 9.5 | 34 |
| **70+** | MAPSE lateral | ≥ 8.8 | 7.3–8.7 | < 7.3 | 12 | | | | |
| | MAPSE septal | ≥ 6.6 | 4.8–6.5 | < 4.8 | 12 | | | | |
| | MAPSE inferior | ≥ 8.7 | 6.6–8.6 | < 6.6 | 12 | | | | |
| | MAPSE anterior | ≥ 6.9 | 5.3–6.8 | < 5.3 | 11 | | | | |
| | TAPSE | ≥ 13.6 | 12.0–13.5 | < 12.0 | 12 | | | | |

**Footnote**: All annular plane systolic excursion measurements were derived using cvi42 software (Circle Cardiovascular Imaging Inc.) with standardized feature-tracking analysis. Mitral annular plane systolic excursion (MAPSE; lateral and septal) and tricuspid annular plane systolic excursion (TAPSE) are presented in millimetres (mm). Reference ranges are defined as the interval between the 2.5th and 97.5th centiles derived from age- and sex-specific generalized additive models for location, scale and shape (GAMLSS) using the Box–Cox Cole–Green distribution. Centile-based categories beyond the reference range are derived from LMS-based thresholds (P0.5 and P2.5) and describe the position of a measurement within the reference distribution rather than disease severity or diagnostic abnormality. For clarity, only the lower reference threshold (2.5th percentile) and corresponding lower centile cut-offs are presented in this table. Reference values are derived from a Chinese adult population.
**Abbreviations**: MAPSE = mitral annular plane systolic excursion; TAPSE = tricuspid annular plane systolic excursion.

### Supplementary Table 46 Ventricular long-axis and atrioventricular junction strain metrics reference ranges and centile-based categories, by age group using cvi42 (Circle Cardiovascular Imaging Inc), *in a Chinese adult population*

| | | Female | | | | Male | | | |
|---|---|---|---|---|---|---|---|---|---|
| **Age group** | Metrics (%) | Reference threshold | Mild | At least moderate | n | Reference threshold | Mild | At least moderate | n |
| **18–29** | LV long-axis strain 4CV | ≥ 15.7 | 14.6–15.6 | < 14.6 | 14 | ≥ 13.5 | 12.3–13.4 | < 12.3 | 17 |
| | LV long-axis strain avg | ≥ 16.3 | 15.3–16.2 | < 15.3 | 13 | ≥ 13.5 | 12.0–13.4 | < 12.0 | 17 |
| | LV AV-junction strain 4CV | ≥ 15.2 | 14.1–15.1 | < 14.1 | 14 | ≥ 13.1 | 12.0–13.0 | < 12.0 | 17 |
| | LV AV-junction strain avg | ≥ 15.8 | 14.8–15.7 | < 14.8 | 13 | ≥ 13.2 | 12.0–13.1 | < 12.0 | 17 |
| **30–39** | LV long-axis strain 4CV | ≥ 15.1 | 13.9–15.0 | < 13.9 | 18 | ≥ 13.1 | 11.8–13.0 | < 11.8 | 16 |
| | LV long-axis strain avg | ≥ 15.7 | 14.6–15.6 | < 14.6 | 18 | ≥ 13.4 | 12.1–13.3 | < 12.1 | 16 |
| | LV AV-junction strain 4CV | ≥ 14.6 | 13.4–14.5 | < 13.4 | 18 | ≥ 12.7 | 11.6–12.6 | < 11.6 | 16 |
| | LV AV-junction strain avg | ≥ 15.3 | 14.1–15.2 | < 14.1 | 18 | ≥ 13.1 | 11.9–13.0 | < 11.9 | 16 |
| **40–49** | LV long-axis strain 4CV | ≥ 14.4 | 13.0–14.3 | < 13.0 | 24 | ≥ 12.7 | 11.3–12.6 | < 11.3 | 19 |
| | LV long-axis strain avg | ≥ 15.0 | 13.8–14.9 | < 13.8 | 24 | ≥ 13.4 | 12.2–13.3 | < 12.2 | 19 |
| | LV AV-junction strain 4CV | ≥ 13.9 | 12.5–13.8 | < 12.5 | 24 | ≥ 12.3 | 11.1–12.2 | < 11.1 | 19 |
| | LV AV-junction strain avg | ≥ 14.6 | 13.3–14.5 | < 13.3 | 24 | ≥ 13.0 | 11.9–12.9 | < 11.9 | 19 |
| **50–59** | LV long-axis strain 4CV | ≥ 13.6 | 12.0–13.5 | < 12.0 | 65 | ≥ 12.2 | 10.8–12.1 | < 10.8 | 46 |
| | LV long-axis strain avg | ≥ 14.2 | 12.8–14.1 | < 12.8 | 66 | ≥ 13.3 | 12.2–13.2 | < 12.2 | 46 |
| | LV AV-junction strain 4CV | ≥ 13.1 | 11.5–13.0 | < 11.5 | 65 | ≥ 11.9 | 10.7–11.8 | < 10.7 | 46 |
| | LV AV-junction strain avg | ≥ 13.9 | 12.5–13.8 | < 12.5 | 66 | ≥ 12.9 | 11.8–12.8 | < 11.8 | 46 |
| **60–69** | LV long-axis strain 4CV | ≥ 12.6 | 10.7–12.5 | < 10.7 | 52 | ≥ 11.8 | 10.3–11.7 | < 10.3 | 35 |
| | LV long-axis strain avg | ≥ 13.4 | 11.7–13.3 | < 11.7 | 51 | ≥ 13.3 | 12.3–13.2 | < 12.3 | 34 |
| | LV AV-junction strain 4CV | ≥ 12.1 | 10.2–12.0 | < 10.2 | 52 | ≥ 11.5 | 10.2–11.4 | < 10.2 | 35 |
| | LV AV-junction strain avg | ≥ 13.1 | 11.7–13.0 | < 11.7 | 51 | ≥ 12.8 | 11.8–12.7 | < 11.8 | 34 |
| **70+** | LV long-axis strain 4CV | ≥ 11.7 | 9.3–11.6 | < 9.3 | 12 | | | | |
| | LV long-axis strain avg | ≥ 12.5 | 10.7–12.4 | < 10.7 | 12 | | | | |
| | LV AV-junction strain 4CV | ≥ 11.2 | 8.9–11.1 | < 8.9 | 12 | | | | |
| | LV AV-junction strain avg | ≥ 12.5 | 10.9–12.4 | < 10.9 | 12 | | | | |

**Footnote**: All ventricular long-axis and atrioventricular junction strain measurements were derived using cvi42 software (Circle Cardiovascular Imaging Inc.) with standardized feature-tracking analysis. Left ventricular (LV) long-axis strain and LV AV-junction strain were assessed in the 4-chamber view (4CV) and as averaged values across available views (avg), and are presented as absolute values (%). Reference ranges are defined as the interval between the 2.5th and 97.5th centiles derived from age- and sex-specific generalized additive models for location, scale and shape (GAMLSS) using the Box–Cox Cole–Green distribution. Centile-based categories beyond the reference range are derived from LMS-based thresholds (P0.5 and P2.5) and describe the position of a measurement within the reference distribution rather than disease severity or diagnostic abnormality. For clarity, only the lower reference threshold (2.5th percentile) and corresponding lower centile cut-offs are presented in this table. Reference values are derived from a Chinese adult population.
**Abbreviations**: LV = left ventricle; AV = atrioventricular; 4CV = four-chamber view.

### Supplementary Table 47 Atrial long-axis and atrioventricular junction strain metrics reference ranges and centile-based categories, by age group using cvi42 (Circle Cardiovascular Imaging Inc), *in a Chinese adult population*

| | | Female | | | | Male | | | |
|---|---|---|---|---|---|---|---|---|---|
| **Age group** | Metrics (%) | Reference threshold | Mild | At least moderate | n | Reference threshold | Mild | At least moderate | n |
| **18–29** | LA long-axis strain 4CV | ≥ 13.2 | 6.4–13.1 | < 6.4 | 14 | ≥ 8.7 | 5.2–8.6 | < 5.2 | 14 |
| | LA long-axis strain avg | ≥ 25.3 | 21.2–25.2 | < 21.2 | 13 | ≥ 20.5 | 17.2–20.4 | < 17.2 | 14 |
| | LA AV-junction strain 4CV | ≥ 12.8 | 6.5–12.7 | < 6.5 | 14 | ≥ 7.4 | 3.7–7.3 | < 3.7 | 14 |
| | LA AV-junction strain avg | ≥ 21.4 | 17.6–21.3 | < 17.6 | 13 | ≥ 17.3 | 14.3–17.2 | < 14.3 | 14 |
| | RA long-axis strain | ≥ 29.3 | 22.7–29.2 | < 22.7 | 13 | ≥ 20.7 | 13.8–20.6 | < 13.8 | 16 |
| | RA AV-junction strain | ≥ 26.3 | 20.1–26.2 | < 20.1 | 13 | ≥ 20.0 | 14.1–19.9 | < 14.1 | 16 |
| **30–39** | LA long-axis strain 4CV | ≥ 11.1 | 4.9–11.0 | < 4.9 | 18 | ≥ 8.5 | 5.1–8.4 | < 5.1 | 14 |
| | LA long-axis strain avg | ≥ 23.0 | 18.8–22.9 | < 18.8 | 18 | ≥ 19.0 | 15.6–18.9 | < 15.6 | 14 |
| | LA AV-junction strain 4CV | ≥ 10.6 | 4.8–10.5 | < 4.8 | 18 | ≥ 7.3 | 3.7–7.2 | < 3.7 | 14 |
| | LA AV-junction strain avg | ≥ 19.6 | 15.7–19.5 | < 15.7 | 18 | ≥ 16.2 | 13.2–16.1 | < 13.2 | 14 |
| | RA long-axis strain | ≥ 30.3 | 24.7–30.2 | < 24.7 | 17 | ≥ 18.0 | 12.0–17.9 | < 12.0 | 16 |
| | RA AV-junction strain | ≥ 27.7 | 22.2–27.6 | < 22.2 | 17 | ≥ 17.1 | 12.1–17.0 | < 12.1 | 16 |
| **40–49** | LA long-axis strain 4CV | ≥ 9.3 | 3.9–9.2 | < 3.9 | 23 | ≥ 8.4 | 5.0–8.3 | < 5.0 | 17 |
| | LA long-axis strain avg | ≥ 20.0 | 15.7–19.9 | < 15.7 | 23 | ≥ 17.3 | 13.9–17.2 | < 13.9 | 18 |
| | LA AV-junction strain 4CV | ≥ 8.7 | 3.5–8.6 | < 3.5 | 23 | ≥ 7.2 | 3.6–7.1 | < 3.6 | 17 |
| | LA AV-junction strain avg | ≥ 17.2 | 13.2–17.1 | < 13.2 | 23 | ≥ 15.1 | 12.2–15.0 | < 12.2 | 18 |
| | RA long-axis strain | ≥ 29.3 | 24.4–29.2 | < 24.4 | 24 | ≥ 17.5 | 11.7–17.4 | < 11.7 | 19 |
| | RA AV-junction strain | ≥ 27.2 | 22.3–27.1 | < 22.3 | 24 | ≥ 16.8 | 11.9–16.7 | < 11.9 | 19 |
| **50–59** | LA long-axis strain 4CV | ≥ 8.4 | 4.2–8.3 | < 4.2 | 64 | ≥ 8.2 | 4.9–8.1 | < 4.9 | 43 |
| | LA long-axis strain avg | ≥ 16.6 | 12.3–16.5 | < 12.3 | 61 | ≥ 15.7 | 12.2–15.6 | < 12.2 | 40 |
| | LA AV-junction strain 4CV | ≥ 8.0 | 3.8–7.9 | < 3.8 | 64 | ≥ 7.0 | 3.5–6.9 | < 3.5 | 43 |
| | LA AV-junction strain avg | ≥ 14.6 | 10.5–14.5 | < 10.5 | 61 | ≥ 14.0 | 11.2–13.9 | < 11.2 | 40 |
| | RA long-axis strain | ≥ 25.5 | 21.0–25.4 | < 21.0 | 66 | ≥ 17.2 | 11.5–17.1 | < 11.5 | 48 |
| | RA AV-junction strain | ≥ 23.7 | 19.2–23.6 | < 19.2 | 66 | ≥ 16.5 | 11.7–16.4 | < 11.7 | 48 |
| **60–69** | LA long-axis strain 4CV | ≥ 7.9 | 5.0–7.8 | < 5.0 | 49 | ≥ 8.1 | 4.8–8.0 | < 4.8 | 30 |
| | LA long-axis strain avg | ≥ 13.0 | 8.8–12.9 | < 8.8 | 48 | ≥ 14.0 | 10.5–13.9 | < 10.5 | 28 |
| | LA AV-junction strain 4CV | ≥ 7.8 | 5.1–7.7 | < 5.1 | 49 | ≥ 6.9 | 3.5–6.8 | < 3.5 | 31 |
| | LA AV-junction strain avg | ≥ 11.8 | 7.7–11.7 | < 7.7 | 48 | ≥ 13.0 | 10.3–12.9 | < 10.3 | 29 |
| | RA long-axis strain | ≥ 19.5 | 15.1–19.4 | < 15.1 | 51 | ≥ 15.6 | 10.4–15.5 | < 10.4 | 35 |
| | RA AV-junction strain | ≥ 17.9 | 13.5–17.8 | < 13.5 | 50 | ≥ 15.4 | 10.8–15.3 | < 10.8 | 35 |
| **70+** | LA long-axis strain 4CV | ≥ 7.5 | 5.3–7.4 | < 5.3 | 12 | | | | |
| | LA long-axis strain avg | ≥ 10.0 | 6.0–9.9 | < 6.0 | 12 | | | | |
| | LA AV-junction strain 4CV | ≥ 7.5 | 5.5–7.4 | < 5.5 | 12 | | | | |
| | LA AV-junction strain avg | ≥ 9.4 | 5.4–9.3 | < 5.4 | 12 | | | | |
| | RA long-axis strain | ≥ 13.8 | 9.6–13.7 | < 9.6 | 12 | | | | |
| | RA AV-junction strain | ≥ 12.2 | 7.9–12.1 | < 7.9 | 12 | | | | |

**Footnote**: All atrial long-axis and atrioventricular junction strain measurements were derived using cvi42 software (Circle Cardiovascular Imaging Inc.) with standardized feature-tracking analysis. Left atrial (LA) long-axis strain and LA AV-junction strain were assessed in the 4-chamber view (4CV) and as averaged values across available views (avg), while right atrial (RA) long-axis and AV-junction strain were derived from standard long-axis views; all values are presented as absolute percentages (%). Reference ranges are defined as the interval between the 2.5th and 97.5th centiles derived from age- and sex-specific generalized additive models for location, scale and shape (GAMLSS) using the Box–Cox Cole–Green distribution. Centile-based categories beyond the reference range are derived from LMS-based thresholds (P0.5 and P2.5) and describe the position of a measurement within the reference distribution rather than disease severity or diagnostic abnormality. For clarity, only the lower reference threshold (2.5th percentile) and corresponding lower centile cut-offs are presented in this table. Reference values are derived from a Chinese adult population.
**Abbreviations**: LA = left atrium; RA = right atrium; AV = atrioventricular; 4CV = four-chamber view.

### Supplementary Table 48 Annular plane systolic excursion metrics reference ranges and centile-based categories, by age group using cvi42 (Circle Cardiovascular Imaging Inc), *in a Mixed adult population*

| | | Female | | | | Male | | | |
|---|---|---|---|---|---|---|---|---|---|
| **Age group** | Metrics (mm) | Reference threshold | Mild | At least moderate | n | Reference threshold | Mild | At least moderate | n |
| **50–59** | MAPSE lateral | ≥ 11.0 | 9.5–10.9 | < 9.5 | 136 | ≥ 9.0 | 6.2–8.9 | < 6.2 | 64 |
| | MAPSE septal | ≥ 9.2 | 7.8–9.1 | < 7.8 | 136 | ≥ 9.4 | 8.0–9.3 | < 8.0 | 64 |
| | MAPSE inferior | ≥ 11.7 | 10.4–11.6 | < 10.4 | 132 | ≥ 11.2 | 9.3–11.1 | < 9.3 | 64 |
| | MAPSE anterior | ≥ 10.1 | 9.0–10.0 | < 9.0 | 133 | ≥ 9.4 | 7.6–9.3 | < 7.6 | 65 |
| | TAPSE | ≥ 15.4 | 12.5–15.3 | < 12.5 | 134 | ≥ 13.5 | 10.5–13.4 | < 10.5 | 65 |
| **60–69** | MAPSE lateral | ≥ 10.2 | 8.7–10.1 | < 8.7 | 108 | ≥ 8.8 | 6.0–8.7 | < 6.0 | 56 |
| | MAPSE septal | ≥ 8.3 | 7.0–8.2 | < 7.0 | 107 | ≥ 8.6 | 7.4–8.5 | < 7.4 | 56 |
| | MAPSE inferior | ≥ 9.8 | 8.0–9.7 | < 8.0 | 110 | ≥ 10.6 | 8.7–10.5 | < 8.7 | 59 |
| | MAPSE anterior | ≥ 9.3 | 8.2–9.2 | < 8.2 | 109 | ≥ 8.3 | 6.7–8.2 | < 6.7 | 58 |
| | TAPSE | ≥ 14.1 | 11.5–14.0 | < 11.5 | 107 | ≥ 13.6 | 10.6–13.5 | < 10.6 | 57 |
| **70+** | MAPSE lateral | ≥ 9.3 | 7.7–9.2 | < 7.7 | 47 | ≥ 8.7 | 5.9–8.6 | < 5.9 | 32 |
| | MAPSE septal | ≥ 7.2 | 6.1–7.1 | < 6.1 | 47 | ≥ 8.4 | 7.2–8.3 | < 7.2 | 32 |
| | MAPSE inferior | ≥ 7.0 | 4.0–6.9 | < 4.0 | 47 | ≥ 9.9 | 8.2–9.8 | < 8.2 | 35 |
| | MAPSE anterior | ≥ 8.3 | 7.4–8.2 | < 7.4 | 47 | ≥ 7.7 | 6.2–7.6 | < 6.2 | 35 |
| | TAPSE | ≥ 11.2 | 7.8–11.1 | < 7.8 | 47 | ≥ 14.0 | 10.9–13.9 | < 10.9 | 32 |

**Footnote**: All annular plane systolic excursion measurements were derived using cvi42 software (Circle Cardiovascular Imaging Inc.) with standardized feature-tracking analysis. Mitral annular plane systolic excursion (MAPSE; lateral and septal) and tricuspid annular plane systolic excursion (TAPSE) are presented in millimetres (mm). Reference ranges are defined as the interval between the 2.5th and 97.5th centiles derived from age- and sex-specific generalized additive models for location, scale and shape (GAMLSS) using the Box–Cox Cole–Green distribution. Centile-based categories beyond the reference range are derived from LMS-based thresholds (P0.5 and P2.5) and describe the position of a measurement within the reference distribution rather than disease severity or diagnostic abnormality. For clarity, only the lower reference threshold (2.5th percentile) and corresponding lower centile cut-offs are presented in this table. Reference values are derived from a Mixed adult population.
**Abbreviations**: MAPSE = mitral annular plane systolic excursion; TAPSE = tricuspid annular plane systolic excursion.

**Supplementary Table 49 Ventricular long-axis and atrioventricular junction strain metrics reference ranges and centile-based categories, by age group using cvi42 (Circle Cardiovascular Imaging Inc),** ***in a Mixed adult population***

| | | Female | | | | Male | | | |
|---|---|---|---|---|---|---|---|---|---|
| **Age group** | Metrics (%) | Reference threshold | Mild | At least moderate | n | Reference threshold | Mild | At least moderate | n |
| **50–59** | LV long-axis strain 4CV | ≥ 13.2 | 11.7–13.1 | < 11.7 | 136 | ≥ 10.4 | 8.4–10.3 | < 8.4 | 64 |
| | LV long-axis strain avg | ≥ 14.4 | 13.0–14.3 | < 13.0 | 132 | ≥ 12.3 | 10.8–12.2 | < 10.8 | 63 |
| | LV AV-junction strain 4CV | ≥ 12.9 | 11.6–12.8 | < 11.6 | 136 | ≥ 10.0 | 8.1–9.9 | < 8.1 | 64 |
| | LV AV-junction strain avg | ≥ 14.0 | 12.7–13.9 | < 12.7 | 132 | ≥ 11.7 | 10.3–11.6 | < 10.3 | 63 |
| **60–69** | LV long-axis strain 4CV | ≥ 12.9 | 11.5–12.8 | < 11.5 | 107 | ≥ 10.6 | 8.6–10.5 | < 8.6 | 56 |
| | LV long-axis strain avg | ≥ 13.9 | 12.5–13.8 | < 12.5 | 107 | ≥ 12.1 | 10.6–12.0 | < 10.6 | 54 |
| | LV AV-junction strain 4CV | ≥ 12.5 | 11.2–12.4 | < 11.2 | 107 | ≥ 10.2 | 8.3–10.1 | < 8.3 | 56 |
| | LV AV-junction strain avg | ≥ 13.3 | 11.9–13.2 | < 11.9 | 107 | ≥ 11.6 | 10.2–11.5 | < 10.2 | 54 |
| **70+** | LV long-axis strain 4CV | ≥ 12.6 | 11.3–12.5 | < 11.3 | 46 | ≥ 10.7 | 8.7–10.6 | < 8.7 | 32 |
| | LV long-axis strain avg | ≥ 13.2 | 11.8–13.1 | < 11.8 | 45 | ≥ 12.5 | 11.1–12.4 | < 11.1 | 32 |
| | LV AV-junction strain 4CV | ≥ 12.0 | 10.7–11.9 | < 10.7 | 46 | ≥ 10.3 | 8.4–10.2 | < 8.4 | 32 |
| | LV AV-junction strain avg | ≥ 12.5 | 11.0–12.4 | < 11.0 | 45 | ≥ 12.1 | 10.6–12.0 | < 10.6 | 32 |

**Footnote**: All ventricular long-axis and atrioventricular junction strain measurements were derived using cvi42 software (Circle Cardiovascular Imaging Inc.) with standardized feature-tracking analysis. Left ventricular (LV) long-axis strain and LV AV-junction strain were assessed in the 4-chamber view (4CV) and as averaged values across available views (avg), and are presented as absolute values (%). Reference ranges are defined as the interval between the 2.5th and 97.5th centiles derived from age- and sex-specific generalized additive models for location, scale and shape (GAMLSS) using the Box–Cox Cole–Green distribution. Centile-based categories beyond the reference range are derived from LMS-based thresholds (P0.5 and P2.5) and describe the position of a measurement within the reference distribution rather than disease severity or diagnostic abnormality. For clarity, only the lower reference threshold (2.5th percentile) and corresponding lower centile cut-offs are presented in this table. Reference values are derived from a Mixed adult population.
**Abbreviations**: LV = left ventricle; AV = atrioventricular; 4CV = four-chamber view.

### Supplementary Table 50 Atrial long-axis and atrioventricular junction strain metrics reference ranges and centile-based categories, by age group using cvi42 (Circle Cardiovascular Imaging Inc), *in a Mixed adult population*

| | | Female | | | | Male | | | |
|---|---|---|---|---|---|---|---|---|---|
| **Age group** | Metrics (%) | Reference threshold | Mild | At least moderate | n | Reference threshold | Mild | At least moderate | n |
| **50–59** | LA long-axis strain 4CV | ≥ 12.1 | 9.2–12.0 | < 9.2 | 131 | ≥ 6.8 | 3.0–6.7 | < 3.0 | 64 |
| | LA long-axis strain avg | ≥ 22.1 | 18.4–22.0 | < 18.4 | 119 | ≥ 19.5 | 15.6–19.4 | < 15.6 | 62 |
| | LA AV-junction strain 4CV | ≥ 11.7 | 8.9–11.6 | < 8.9 | 131 | ≥ 8.0 | 4.8–7.9 | < 4.8 | 65 |
| | LA AV-junction strain avg | ≥ 19.8 | 16.6–19.7 | < 16.6 | 121 | ≥ 16.6 | 12.9–16.5 | < 12.9 | 62 |
| | RA long-axis strain | ≥ 21.7 | 17.1–21.6 | < 17.1 | 132 | ≥ 20.4 | 16.8–20.3 | < 16.8 | 66 |
| | RA AV-junction strain | ≥ 20.7 | 16.7–20.6 | < 16.7 | 132 | ≥ 15.5 | 10.9–15.4 | < 10.9 | 66 |
| **60–69** | LA long-axis strain 4CV | ≥ 11.3 | 8.6–11.2 | < 8.6 | 106 | ≥ 7.0 | 3.1–6.9 | < 3.1 | 56 |
| | LA long-axis strain avg | ≥ 19.8 | 16.5–19.7 | < 16.5 | 103 | ≥ 16.7 | 13.4–16.6 | < 13.4 | 53 |
| | LA AV-junction strain 4CV | ≥ 10.9 | 8.3–10.8 | < 8.3 | 105 | ≥ 8.1 | 4.9–8.0 | < 4.9 | 57 |
| | LA AV-junction strain avg | ≥ 17.9 | 14.9–17.8 | < 14.9 | 103 | ≥ 14.8 | 11.5–14.7 | < 11.5 | 53 |
| | RA long-axis strain | ≥ 20.0 | 15.7–19.9 | < 15.7 | 107 | ≥ 18.9 | 15.6–18.8 | < 15.6 | 55 |
| | RA AV-junction strain | ≥ 19.0 | 15.4–18.9 | < 15.4 | 107 | ≥ 13.9 | 9.8–13.8 | < 9.8 | 55 |
| **70+** | LA long-axis strain 4CV | ≥ 10.4 | 7.9–10.3 | < 7.9 | 46 | ≥ 8.0 | 3.6–7.9 | < 3.6 | 29 |
| | LA long-axis strain avg | ≥ 17.2 | 14.4–17.1 | < 14.4 | 45 | ≥ 19.2 | 15.4–19.1 | < 15.4 | 30 |
| | LA AV-junction strain 4CV | ≥ 10.0 | 7.7–9.9 | < 7.7 | 46 | ≥ 9.5 | 5.7–9.4 | < 5.7 | 30 |
| | LA AV-junction strain avg | ≥ 15.7 | 13.1–15.6 | < 13.1 | 45 | ≥ 17.0 | 13.2–16.9 | < 13.2 | 30 |
| | RA long-axis strain | ≥ 18.2 | 14.3–18.1 | < 14.3 | 47 | ≥ 20.8 | 17.1–20.7 | < 17.1 | 29 |
| | RA AV-junction strain | ≥ 17.2 | 13.9–17.1 | < 13.9 | 47 | ≥ 16.1 | 11.4–16.0 | < 11.4 | 29 |

**Footnote**: All atrial long-axis and atrioventricular junction strain measurements were derived using cvi42 software (Circle Cardiovascular Imaging Inc.) with standardized feature-tracking analysis. Left atrial (LA) long-axis strain and LA AV-junction strain were assessed in the 4-chamber view (4CV) and as averaged values across available views (avg), while right atrial (RA) long-axis and AV-junction strain were derived from standard long-axis views; all values are presented as absolute percentages (%). Reference ranges are defined as the interval between the 2.5th and 97.5th centiles derived from age- and sex-specific generalized additive models for location, scale and shape (GAMLSS) using the Box–Cox Cole–Green distribution. Centile-based categories beyond the reference range are derived from LMS-based thresholds (P0.5 and P2.5) and describe the position of a measurement within the reference distribution rather than disease severity or diagnostic abnormality. For clarity, only the lower reference threshold (2.5th percentile) and corresponding lower centile cut-offs are presented in this table. Reference values are derived from a Mixed adult population.
**Abbreviations**: LA = left atrium; RA = right atrium; AV = atrioventricular; 4CV = four-chamber view.
